# Identifying cohorts at elevated risk of cancers using generative modeling of patient health states

**DOI:** 10.64898/2026.09.09.26362676

**Authors:** Asif Khan, Duncan T. Forster, Moshir Harsh, Chunlei Zheng, Erica T. Warner, Daniel Ritter, Allison Chang, Qi Wei, Tanya K. Sorensen, Debora S. Marks, Lecia V. Sequist, Jennifer Hadlock, Nathanael R. Fillmore, Chris Sander

## Abstract

While large language models are powerful generators of new text, forecasting disease progression from longitudinal health histories remains a challenging problem. We introduce GenEHR, an autoregressive generative model trained on electronic health records (EHRs) from millions of patients in diverse health systems. The model learns from real-world multimodal patient histories, including diagnoses, medications, lab values, and procedures while explicitly representing irregular intervals between visits to forecast future clinical events. We combine the general-purpose patient representation learned during foundational training with parameter-efficient supervised adaptation for the task of risk stratification for 17 cancer types. In five large EHR cohorts, supervised adaptation substantially improved prediction performance of a first cancer diagnosis within a five year horizon window. Our retrospective results support the prospective evaluation of GenEHR-CancerRisk as a clinical decision-support tool for prioritizing high-risk patients for screening and prevention of aggressive cancer types, such as pancreatic and ovarian cancer.

## Introduction

### Global challenge of early cancer detection

Cancer is a major global health challenge, and the stage at which it is detected is one of the strongest predictors of outcomes, healthspan, and survival. Once malignancy progresses to an advanced stage, therapeutic options are limited and outcomes are worse compared to treatment at an early, localized stage^1^. Screening can shift diagnosis earlier, but population-wide testing, even when recommended (e.g., by USPSTF), is constrained by cost, availability of tests, heterogeneity of true risk among the screen-eligible population, and the burden of false positives or poor uptake^2^. An immediate task is therefore to identify patients whose future risk is high enough to justify further investigation or closer surveillance to preempt disease progression. This requires models that use routinely collected, widely available longitudinal data, update patient’s risk as their health progresses over time and provide the information as clinical decision support.

### EHR histories contain distributed signals of future cancer risk

Structured electronic health records (EHRs) offer a rich, detailed view of a patient’s health. EHRs include a wide range of multimodal information such as diagnosis codes, medications, procedures, laboratory measurements, demographic factors, geocoding, and lifestyle factors collected during standard clinical care. Early signals relevant to cancer may be weak individually and distributed over years, and their information value depends on sequence and context. For example, iron-deficiency anemia can be a signal for colorectal cancer, while jaundice, evolving abdominal symptoms, and changes in glycaemic control may occur before pancreatic cancer^3–7^. These clinical events occur alongside epidemiological risk factors such as age, smoking, and obesity^8,9^, whose predictive value may depend on their timing and clinical context. Models trained on longitudinal EHRs can test whether these features add information beyond demographic or established clinical risk factors.

### EHR models for cancer risk prediction and patient health state representation

Existing supervised approaches to pancreatic, lung, gastrointestinal, and pan-cancer risk prediction have shown that models trained on longitudinal EHR data can be used to risk stratify patients months to years before the diagnosis^10–14^. However, when labeled data are scarce, supervised model performance can be sensitive to differences between cohorts and healthcare systems. Several aggressive cancers are rare–for example, pancreatic cancer has an annual incidence of approximately 14 per 100,000 people in the U.S.(all ages, SEER^15^). Unsupervised representation learning on broader EHR data can instead learn general features of patient health that can then be adapted to specific diseases such as cancer, especially rare ones.

Earlier methods such as Med2Vec^16^, Deep Patient^17^, multilevel medical embedding (MiME^18^) learn representations of clinical events, visits, and patient histories. More recently, transformer based architectures such as BEHRT^19^, MedBERT^20^, CEHR-BERT^21^, TransformEHR^22^, have shown the benefit of pre-training for downstream tasks such as mortality, readmission, and risk prediction^19^. Recent approaches such as CLMBR^23^ predict sets of future codes, whereas Foresight^24^, ETHOS^25^, and CoMET^26^ autoregressively generate future event sequences. CEHR-GPT^27^ similarly uses autoregressive modeling to generate synthetic longitudinal EHRs. MOTOR^28^ learns patient representation with a multi-task time-to-event objective but does not generate event sequences. SurvivEHR^29^ models competing time-to-event distributions, while Delphi^30^ models competing event rates; both can forecast trajectories. More recently, Apollo^31^ combined longitudinal EHR with clinical text and images using masked auto-encoder training to learn patient representations, which were then applied to clinical forecasting tasks.

### Inter-visit time intervals are essential for capturing the clinical signal

Inter-visit time intervals carry information about a patient’s health state. For example, a repeat blood test within weeks can signal an active health issue, whereas the same test after a year may reflect routine follow-up. Interleaving time tokens with clinical visits is a common approach, but existing methods generally encode time at a fixed resolution. Increasing temporal resolution expands the time vocabulary as well as context length. For example, representing gaps of up to ten years at day-level resolution^27^ would require 3650 distinct tokens. Many exact gaps are sparsely observed, making it difficult to learn each token’s representation and its relationship to other intervals. ETHOS^25^ and CoMET^26^ instead use a fixed vocabulary of interval tokens, but cannot flexibly capture fine-grained gaps. Such limitations can become problematic when precursor events cluster tightly before diagnosis or are separated by prolonged asymptomatic intervals. We introduce GenEHR (Fig 1) that decomposes each inter-visit time gap into a small set of digits at progressively longer time scales using RAdix Time Encoding (RATE). This allows the model to predict what happens next and when it happens at a day level resolution using a small set of time tokens.

**Figure 1:**
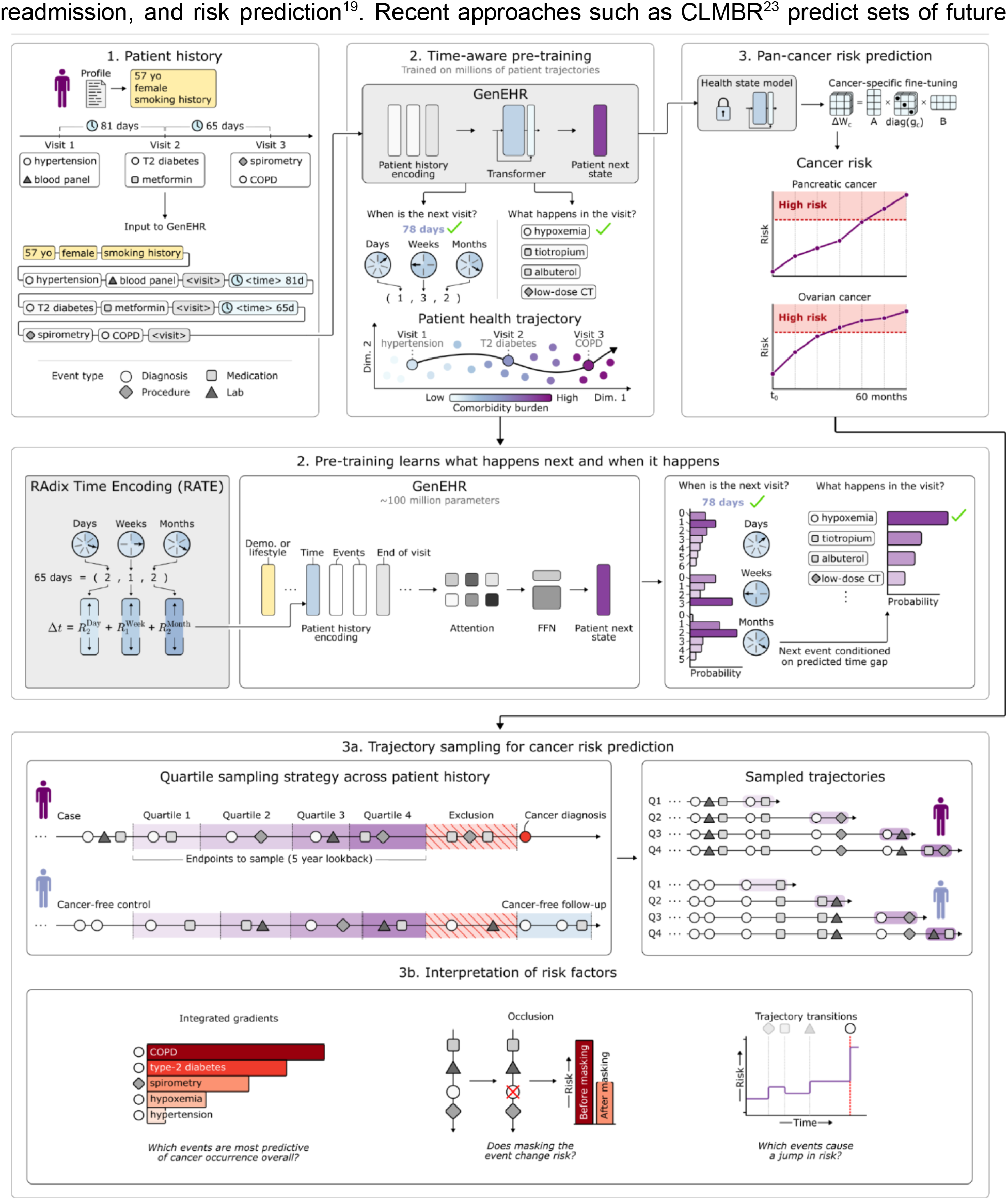
GenEHR: a foundation model of patient health state adapted for pan-cancer risk prediction. 1. Patient histories are tokenized as a combination of static (demographic and lifestyle) and dynamic (time-stamped events including diagnoses, medications, procedures, and laboratory results) variables. 2. GenEHR learns which clinical events occur next and after the end of visit, the five digits of the next inter-visit gap. 3a. Pre-trained GenEHR is combined with a piecewise exponential risk prediction head and cancer-gated low-rank adapters for pan-cancer risk stratification. For training and evaluation, the five year window preceding cancer diagnosis is divided into four equal size time bins and one trajectory endpoint is sampled from each bin, so as to assess risk at different points in a patient’s history. Trajectories start at an earlier time and training and predictions are made at the trajectory endpoint. We exclude events recorded in a three month window before cancer diagnosis to reduce information leakage from diagnostic workup. For no-cancer controls we require five years of observed follow-up after the trajectory endpoint. 3b. Features contributing to predicted cancer risk. We use IG to measure the importance of each event within a patient trajectory (left). We select top medication and diagnosis events based on IG and and by masking them measure the change in cancer log-odds across the cohort (middle). For high risk patients, we sequentially score all sub trajectories to identify events that consistently raise or lower the cancer log odds (right).

### Fine-tuning for cancer risk using registry-data and outcome-specific time-to-event modeling

While generative models learn broad patterns of disease progression and healthcare practices, adapting them to a specific clinical task usually requires updating model parameters. Full fine-tuning is computationally expensive and can cause forgetting, where the foundation model drifts from its pre-trained state^32^. We therefore use parameter-efficient fine-tuning (PEFT) with gated low-rank adaptation (LoRA), which freezes the pretrained model and learns a small set of adapter parameters in selected transformer layers^33^. We decompose each adapter into low-rank components shared across cancer types and a cancer-specific gate sets the strength of each component. Rare cancers can therefore benefit from the information in a shared direction instead of learning from their own few outcomes. We add multiple heads for pan-cancer risk prediction on top of GenEHR that learns to predict risk of all cancers simultaneously. Selected cancers are reported in Methods (Table 1). As diagnosis codes in the EHR are frequently recorded for the purpose of billing and reimbursement of tests and treatments rather than to actually confirm the presence of a diagnosis, and more than half of cancer diagnoses recorded are not confirmed by the registry (Fig S1c, S2c), we use the ‘gold standard’ cancer registry data for outcomes.

**Table 1:**
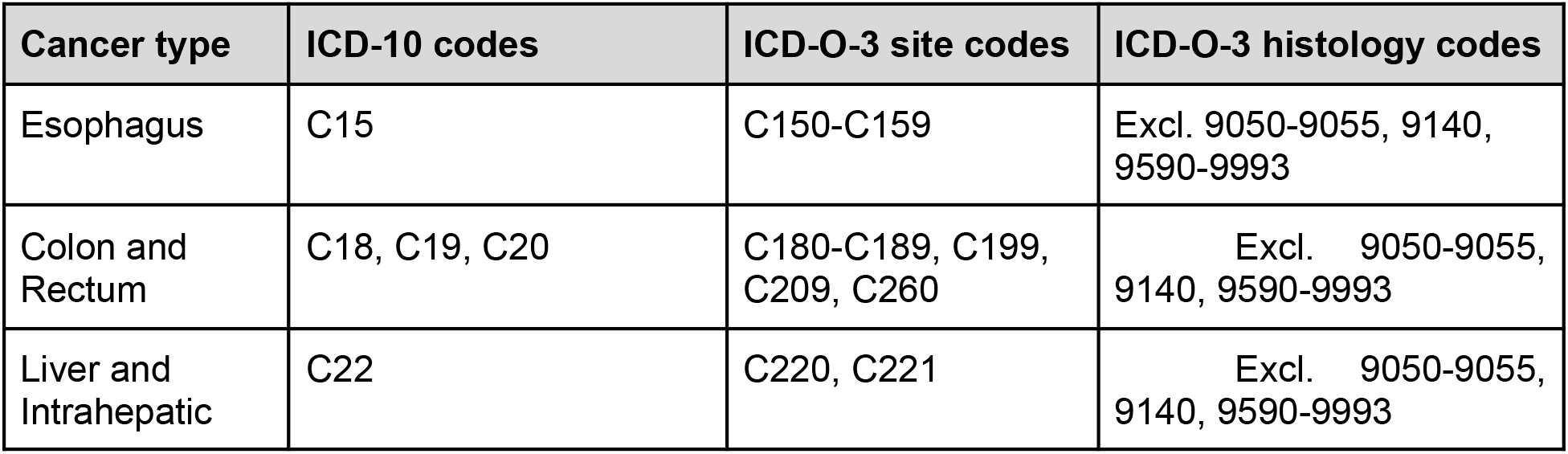
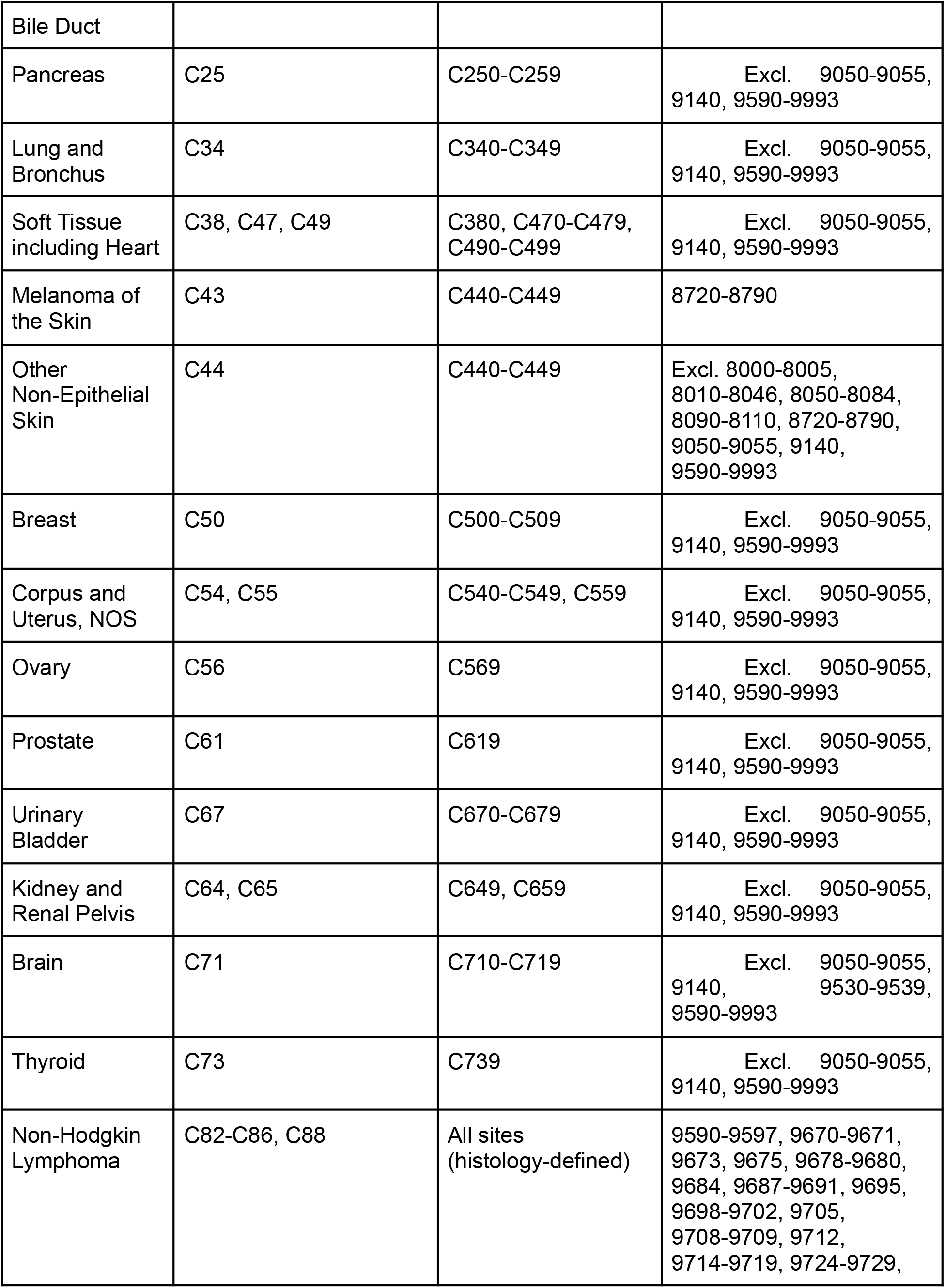
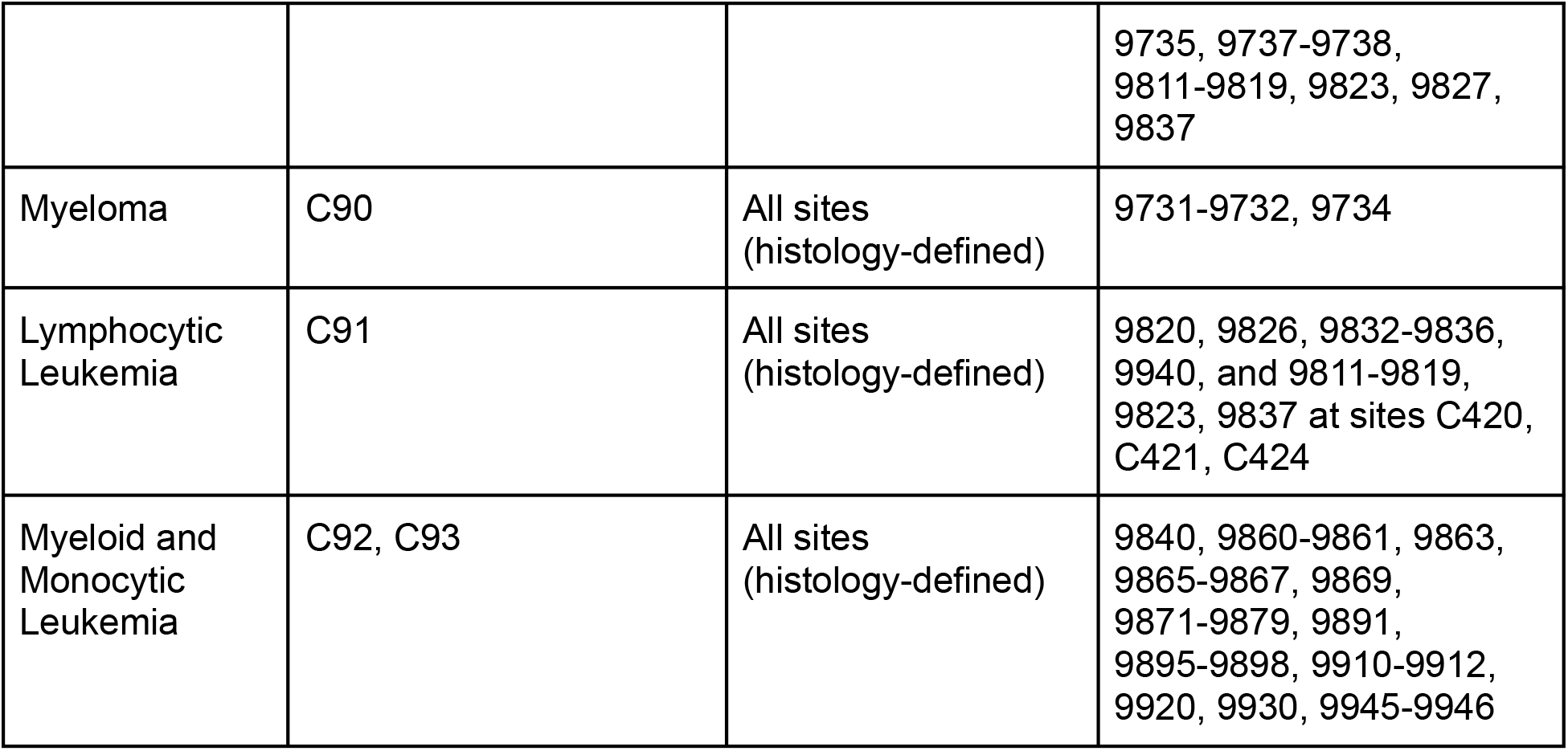
Cancer definition using SEER site code and pan-cancer risk prediction.

### A unified framework for pan-cancer risk prediction

We present GenEHR, a unified framework for generative modeling of patient trajectories and pan-cancer risk prediction for up to 17 cancer types (Table 1) using large-scale EHR data. We trained and evaluated cohort-specific GenEHR models using de-identified patient histories from Providence Health & Services (PROV), Mass General Brigham (MGB)^34^, U.S. Veterans-Affairs (US-VA)^35^, UK Biobank (UKBB)^36^, and NIH All of Us Research Program^37^. Across these cohorts, we demonstrated broad applicability of GenEHR on a range of tasks such as conditional prediction, trajectory generation, time-gap fidelity, learned health-state representations, and contextual event relationships reflecting model belief about data. GenEHR was then adapted for pan-cancer risk prediction across pre-specified horizons using supervised time-to-event modeling. Beyond overall discrimination performance (AUROC), GenEHR clinical utility is demonstrated at a clinical operating point using positive predictive value (PPV, defined as the proportion of selected high-risk patients followed by a cancer diagnosis within the prediction horizon, standardized incidence ratio (SIR, defined as the ratio of observed cancer cases to those expected from age- and sex-specific population incidence over the same prediction horizon), and “suggested number to screen” (SNS, defined as the reciprocal of PPV). We interpreted key features contributing to model risk using contextual pointwise mutual information (cPMI), integrated gradients (IG)^38^, and occlusion analysis. In summary, GenEHR is a time-gap aware foundation model of EHR specialized for targeted pan-cancer risk stratification, a step towards prospective clinical decision-making.

## Results

### Large-scale multi-institutional EHR cohorts

We used longitudinal patient histories from five large-scale EHR data sources, including three U.S. health systems Providence Health & Services (PROV), Mass General Brigham (MGB), U.S. Veterans Affairs (US-VA)), and two national research cohorts, namely the U.K. Biobank (UKBB) and the NIH All of Us Research Program (AoU) (Fig 2). Access to patient data in secure protected computing environments was regulated by IRB protocols at MGB, US-VA, and PROV. Data harmonization, tokenization of clinical events, cohort-specific inclusion criteria, observation periods, and detailed data statistics are in the Methods.

**Figure 2:**
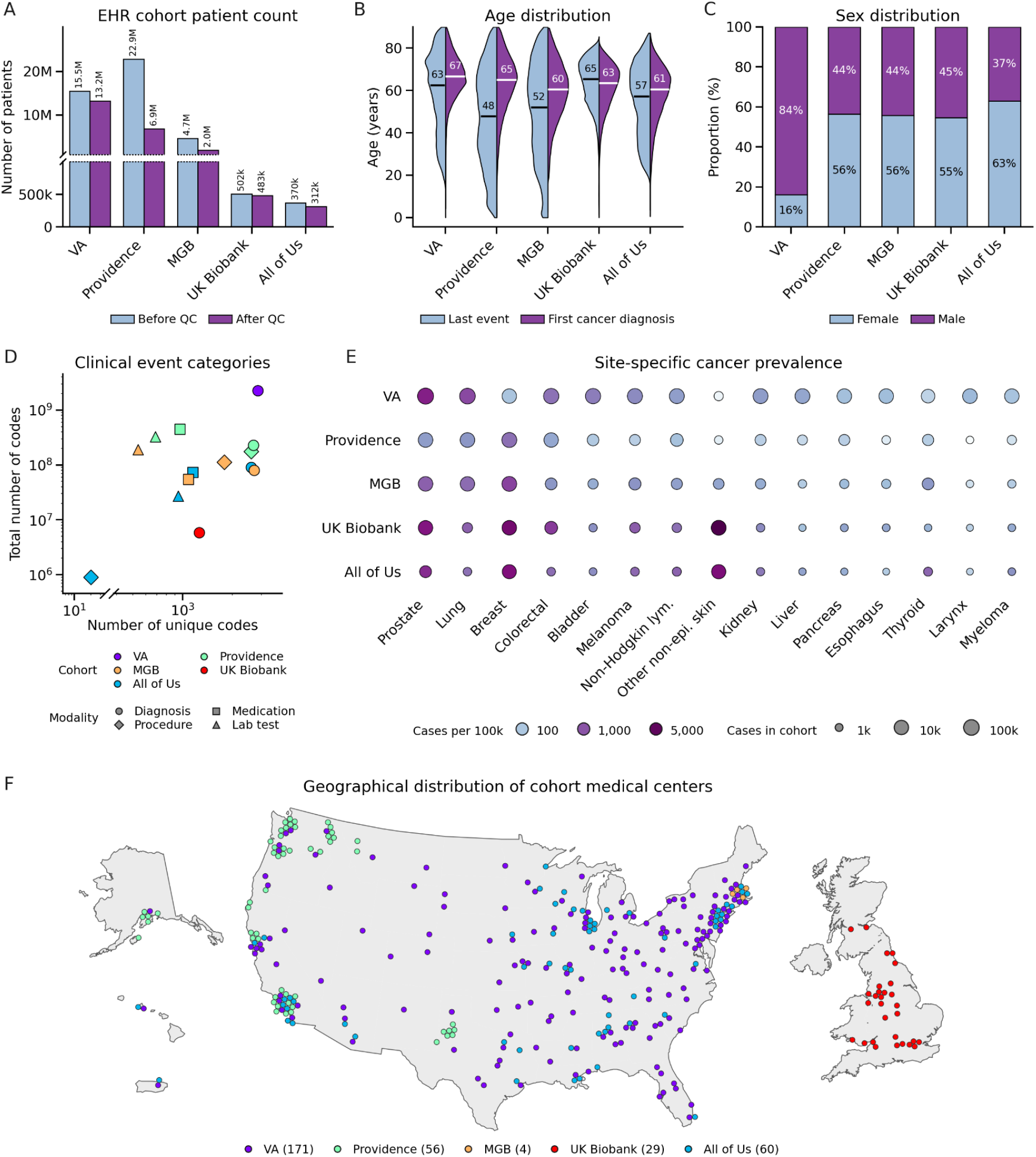
Datasets: large-scale multi-institutional EHR cohorts. (A) Millions of patients: cohort size after site specific quality control filter (Supplement 1). (B-C) Age and sex distribution. Age distribution at the end of the patient record (left) and at first cancer diagnosis (right) indicates broad coverage. Age distributions were truncated above 89 to prevent patient re-identification. (D) Rich data: total number of unique data types and events across modalities (diagnoses, medications, procedures, and laboratory tests). (E) Diverse cancer prevalence by cancer type and cohort. (F) Broad geographic distribution of institutions in healthcare systems. Details in Supplementary Fig S1–S5.

#### Cancer outcomes

MGB, US-VA, PROV, and UKBB used linked cancer registries to avoid known inaccuracies resulting from billing practices^39^. Registry records serve as the gold standard as they are confirmed by registrar teams based on pathology reports and other clinical records. For AoU, which does not have linked cancer-registry data, we require at least two occurrences of cancer-specific ICD codes to define cancer cases.

### Generative foundation model and multi-institutional training (GenEHR; Figure 1)

We represent each patient as a unified sequence of static (time-independent) and longitudinal (time sequence) tokens. The static prefix consists of demographic and lifestyle factors represented as discrete tokens. All events on the same date are grouped into a visit block, and we use a visit token to mark the end of a visit and a < *rate*_*gap* > token to represent the time from that visit to the next visit. All token embeddings except padding and static prefix are learned as a part of the training process.

### RATE learns intervisit intervals at day level resolution

The time gap between visits in EHR data is irregular and influences the model representation and interpretation of patient health. For example, a repeat blood test within a week of an abnormal result may indicate active illness, whereas a test one year apart can be a part of a routine annual checkup. We therefore encode each inter-visit interval, using a mixed-radix digit encoding that is designed to capture the essence of different time scales. The first digit represents 1-7 days, the second counts 1-4 seven day periods, and subsequent digits represent progressively longer intervals. Using radices (7, 4, 6, 5, 5) gives place values of (1, 7, 28, 168, 840) days. For example, a gap of 75 days is encoded as (5, 2, 2, 0, 0) because 75 = 5 × 1 + 2 × 7 + 2 × 28 + 0 × 168 + 0 × 840. Thus RATE preserves day-level resolution without requiring no-event^30^ or a separate token for every possible interval^25^. Similar to events, all digits embeddings are learned. The composition of digit embedding is used at the respective < *rate*_*gap* > position. For full details of tokenization refer to Methods.

### Model training, inference, and evaluation process

We train the model independently within each cohort using a combination of an autoregressive next event objective and a time gap objective. RATE heads predict the time gap digits from the hidden state of the preceding visit token. During generation, we sample event tokens autoregressively from the event head. After a visit token is sampled, its hidden state is given to the RATE heads to sample the time digits. Their embeddings are then composed and inserted at the < *rate*_*gap* > position, allowing events to be generated conditional on both the clinical history and the sampled elapsed time. Full details on tokenization and model formulation are in Supplement.

We evaluated predictive and generative performance together with the clinical structure of the learned representations. Conditional prediction asks whether held-out next events are ranked highly from an observed patient context at the end of a partial trajectory. Generation starts with a randomly sampled static-token (starting points), samples subsequent event tokens and RATE time gaps, and then comparison is done by comparing prevalence of events and time-gaps in generated and real data. We further examined whether the learned event and patient representation were organized by clinical category and disease burden. We also constructed a clinical belief graph using a cPMI matrix. Using context-dependent shifts in probability distributions for next event in subsequent visits, cPMI identifies recurrent, highly probable associations among clinical events within a cohort of held-out patients. Detailed evaluation scores are in the Methods with Table 1 providing the role and interpretation of each score. PROV and US-VA each used a fixed held-out retrospective cohort of one million patients, MGB used 500,000, All of Us used 50,000 and UKBB used 75,000 patients. The remaining cohort is further divided into training and validation sets. Final evaluation metrics were computed only on the retrospective held-out cohort.

We also carried out a scaling analysis by training multiple model architectures with increasing parameter counts and evaluating them on a common held-out dataset. Larger model size leads to a lower event perplexity, high next event accuracy and cancer-risk AUROC (Fig S14 MGB). Because of the computational cost, this analysis was limited to the MGB cohort.

### GenEHR foundation model learns contextual patient representations useful for clinical forecasting and hypothesis generation

We evaluated GenEHR on a range of tasks such as next visit forecasting, transitions in health state during disease progression, and downstream pan-cancer risk prediction.

### Conditional prediction, calibration, and trajectory generation

We evaluated next-visit retrieval at every eligible visit endpoint in the held-out patient trajectories, with results pooled across endpoints. Across cohorts, retrieval-AUC for next event in next visit, defined as the area under recall curve as a function of the fraction of the event vocabulary searched, ranged from 0.90 to 0.99 (Fig 3B). For the first occurrence of broader clinical categories such as the ICD chapter, one-vs-rest macro-AUROC across categories ranged from 0.98 to 0.99 (Fig 3B). Next, an autoregressive simulation tested whether the model captured the overall prevalence of events and time-gaps in observed data. Event prevalence in generated trajectories reproduced the observed prevalence in real-data with log-Pearson correlation of 0.93 to 0.99 and Spearman correlation of 0.90 to 0.98 (Fig 3). Among the hundred most prevalent observed codes pairwise co-occurrence was the fraction of trajectories in which both codes appeared at least once, regardless of whether they occurred in the same visit. Its log-Pearson-correlation was 0.92-0.97, showing that generated trajectories preserved broad code-pair associations in addition to individual frequencies. Generated inter-visit time gaps also correlated strong correlation with observed time gaps, with Spearman correlation of 0.89 to 0.98.

**Figure 3:**
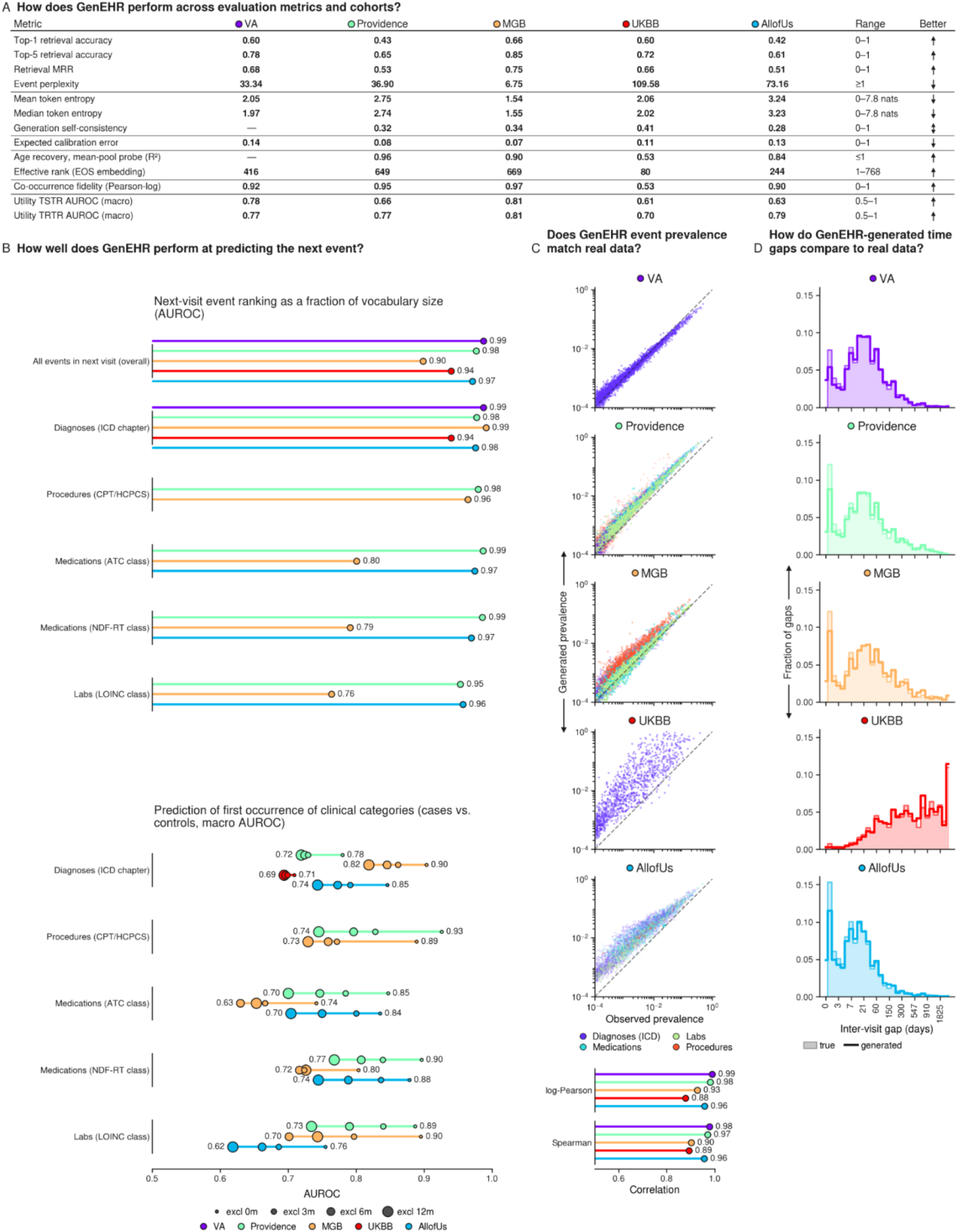
GenEHR foundation model learns contextual patient representations useful for clinical forecasting. (A) Model performance of general forecasting on held-out patient histories. For any patient trajectory, GenEHR predicts the next clinical visit after a time gap and recovers events in the visit. Details of evaluation metrics (MRR, TSTR, TRTR) in Supplement Methods. (B) Top: next visit event recovery assesses the model ranking of a correct event as a fraction of the event vocabulary. Bottom: For prediction of next-event clinical categories, we aggregate event probabilities within each clinical category and evaluate performance using one-vs-rest AUROC scoring. Exclusion window (‘excl 0-12m’): only evaluate trajectories without events in a certain time interval before the first occurrence of the clinical category. (C) Assessment of autoregressively generated trajectories: comparison of the prevalence of events in generated trajectories with the prevalence of events in the observed data; and log-Pearson and Spearman correlations of generated vs observed. (D) Distributions of observed and model-generated inter-visit time gaps. Together, (C-D) indicate how well events generated by the GenEHR model capture population prevalence of both clinical events and the temporal structure of patient histories.

Mean entropy of the next-event distribution quantified predictive uncertainty and ranged from 1.54 to 2.77 nats across cohorts. Lower entropy means that the probable next event is narrowed to fewer choices, whereas higher entropy means several events are plausible and the model is less certain which will occur. We used expected calibration error (ECE) to assess the reliability of next-event probabilities which ranged from 0.07 to 0.14 across cohorts. To assess how far in advance the model could predict ICD-chapter onset, we excluded events during 3-, 6-, and 12-month windows preceding chapter onset (bottom; Fig 3B). Performance suggests that the model learns from information present earlier in the patient trajectory rather than relying solely on events immediately preceding cancer diagnosis. We also carried out an ablation of time encoding by comparing RATE with ETHOS (a fixed set of time-gap tokens based on prior knowledge), relative-time encoding (a vocabulary with one token for each day up to ten years), no-time encoding (time tokens removed), and no-time-and-visit (both time and visit tokens removed). GenEHR with RATE performs best for predicting the next event in the next visit with the largest improvement in forecasting at longer time gaps (Fig S16b).

Across cohorts, GenEHR learned event and time-gap structure of patient histories with good accuracy and calibration, making it suitable for a broad range of downstream clinical tasks.

### GenEHR learns patient health state progression

We next asked whether the learned representations encode clinically meaningful differences between patients. We extracted patient-level representations from the penultimate layer of the model (using <eos> token) trained on the MGB cohort and visualized them in two dimensions using UMAP^40^. UMAP projection showed smooth gradients in Charlson comorbidity index (CCI)^41^, with higher disease burden histories mapped to nearby regions (bottom left; Fig 4). Because the representation using <eos> token summarizes overall health but not its evolution, we separately extracted the representation at each visit state. All visits across patients were projected into two dimensions and were connected using averaged displacement vectors within local regions of the UMAP (bottom right; Fig 4). The resulting visit path moved toward regions with higher average CCI and chronic-disease burden. This map shows a descriptive representation of longitudinal health trajectory over time. Results on other cohorts (MGB Fig S7; PROV S17, S19D; VA S25, S27D; UKBB S33, S35D; AoU S41, S43D).

**Figure 4:**
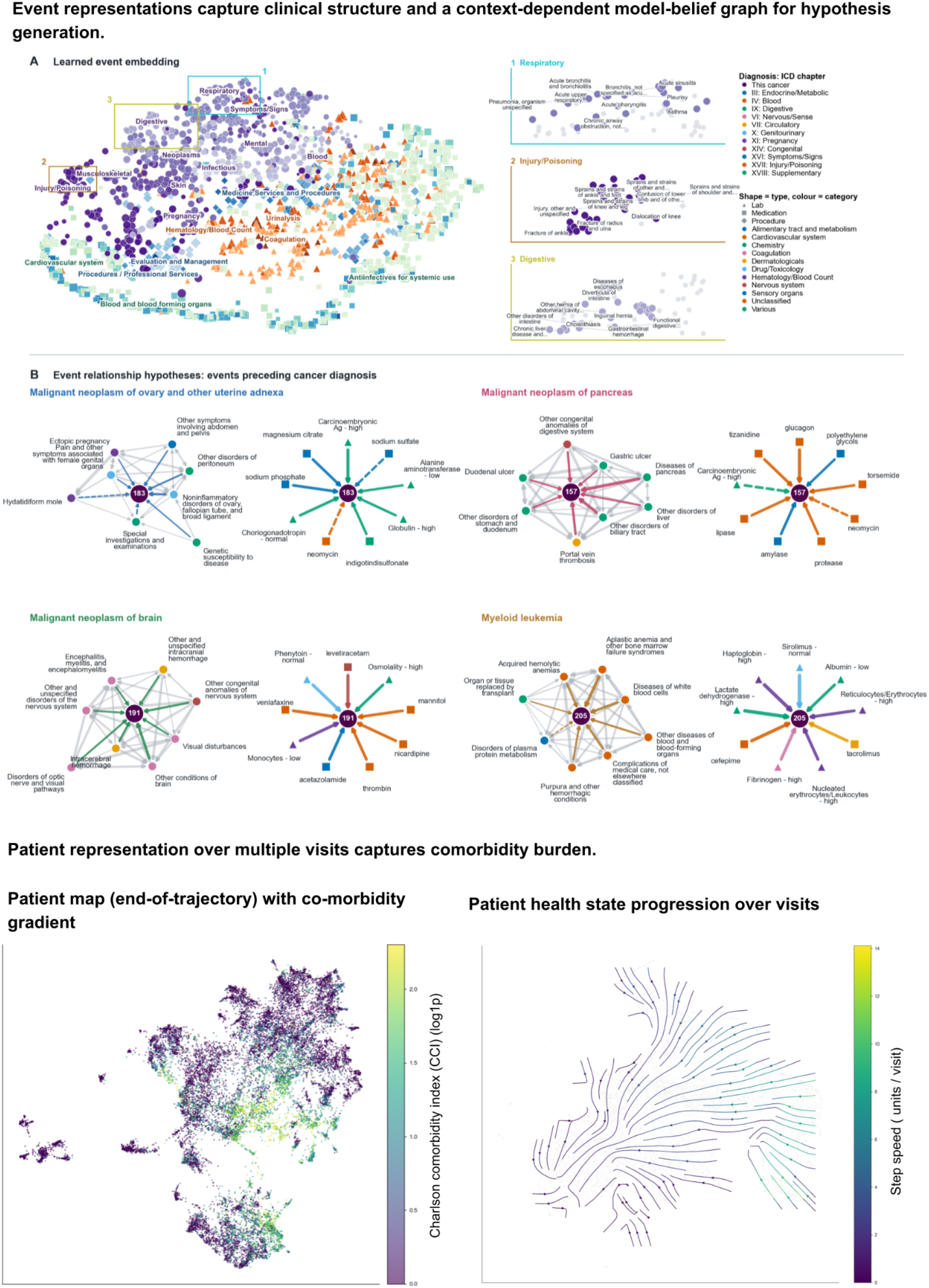
For the MGB dataset, other cohorts in Supplement. **(Top)** Left: **(A)** UMAP projection of learned event embeddings using cosine distance between embedding vectors. Right: closeup of three regions numbered 1 to 3 shows GenEHR learns hierarchy of clinical events without explicit prior knowledge and without using text. **(B)** Event relationship preceding cancer diagnosis inferred using contextual pointwise mutual information (cPMI) between pairs of events for selected cancers (ICD codes 183, 157, 191, 205). For held-out patients whose preceding visit contains an event e_1_, cPMI compares the average model probability assigned to e_2_ normalized against the model averaged probability of e_2_ across all positions and patients. In the cPMI belief graph nodes are clinical events and edges are informational links. Left: Diagnosis neighborhood of cancer with edges among preceding events, Right: Cross-modality hub - medications, procedures, and labs preceding cancer diagnosis. Dashed edges: events with weaker mutual information scores that are plausibly less specific to that cancer. (**Bottom)** UMAP neighborhoods of patients using their end-of-trajectory <eos> representation reflect gradient of comorbidity burden (left), and transitions along trajectories are informative about how the learned state of patients evolves over time using their <visit> representation over multiple visits. Flow (arrows) indicates how patient health evolves over visits (right) averaged over groups of held-out patients.

### Learned clinical event structure supports events-to-cancer hypothesis generation

We first evaluated whether the event representations learned by GenEHR captured any information structure. We projected the learned event embeddings using UMAP (Fig 4A) and graphically coloured the events by ICD chapters, medication ATC class, lab assay group, and CPT procedure section. Although no ICD hierarchy information was used during training, events clustered by clinical categories with refined substructure, e.g., three magnified regions show separation of acute from chronic respiratory disease, sprains from fracture, and upper gastrointestinal symptoms from hernia and hepatobiliary disease.

Next, we used GenEHR generation to construct a model-derived belief graph of event relations from held-out patient trajectories preceding cancer diagnosis. We excluded events on or after cancer diagnosis and within three months before it, to reduce prodromal signals. At each visit boundary, we used the clinical context through the current visit including an event A and the time gap to compute the probability distribution over the next event B. We then averaged the probability distribution over next event B across all positions and patients rather than looking at the most likely prediction. This conditional average was compared with the overall probability assigned to B across all contexts. A positive cPMI(A→B) therefore means that the model assigns a higher probability to B after A than expected from how often B generally occurs.

We mapped cPMI to a belief graph keeping sufficiently strong associations (Methods). Some edges in the graph reflect clinical prior knowledge (MGB Fig 4B, Fig S9a-c; PROV S19, S20a; VA S27, S28a; UKBB S35, S36a-b; AoU S43, S44a-b). Disease of the pancreas and disorders of biliary tract are top associations to pancreatic cancer followed by disorders of liver and gallbladder. These associations are linked to pancreatic inflammation, blockage of bile flow and may lead to the diagnosis of a previously undetected pancreatic tumor^42,43^.

The brain cancer neighborhood captures both broad brain disease and treatment context. Visual and optic-nerve disorders and haemorrhage cluster with anti-seizure medicines (levetiracetam and phenytoin), and treatment as well as monitoring for brain swelling and increased intracranial pressure (mannitol and serum osmolality)^44,45^. By contrast, the myeloid-leukaemia neighborhood reflects abnormal blood-cell production and its diagnostic workup. Aplastic anaemia and other bone-marrow failure disorders are top preceding signals alongside white-cell abnormalities, purpura, elevated lactate dehydrogenase, and nucleated erythrocyte and reticulocyte measurements. This pattern is consistent with bone marrow dysfunction and increased cellular turnover. Aplastic anaemia may represent an alternative diagnosis or a condition that later progressed to myelodysplastic syndrome or acute myeloid leukaemia^46^.

Since edges in cPMI represent learned model belief about the cohort they do not imply causality (Fig 4B). This model-belief graph can be constructed for any retrospective cohort, e.g., in different health systems, to analyze event relationships within patient context. The inferred edges are hypotheses learned from temporal patterns in clinical records, which may reflect disease progression, diagnostic work-up or healthcare use and not, or not necessarily, causal relationships. It is reassuring that the edges, derived in a data-driven mode, appear to agree with prior knowledge in many cases. However, the edges not explained by clinical practice may generate hypotheses for further epidemiological or experimental investigation.

### Parameter efficient fine-tuning improves pan-cancer risk prediction(GenEHR-CancerRisk)

Generative pre-training learns how diagnosis, medications, lab tests, procedures, and time intervals change the probability of future cancer events, including cancer-associated events. However, the next-event objective does not directly estimate the time to a first cancer diagnosis. We therefore adapt the pretrained GenEHR model for pan-cancer risk prediction with a time-to-event objective that uses each patient’s observed history and the period over which a new diagnosis could be observed. Selected cancer names and their definition are included in the Methods (Table 1).

### Trajectory sampling prevents data leakage and allows learning from earlier clinical histories

We sampled multiple endpoints per patient to learn from earlier clinical histories. Within a five year window ending at the last eligible endpoint before cancer diagnosis, we divided time ordered patient trajectory into four approximately equal sized segments and sampled one cutoff from each segment. To reduce the influence of immediate diagnostic work-up, we excluded events recorded during the three-month before the first cancer diagnosis. For cancer-free controls a reference date is assigned by randomly sampling from observed calendar dates of cancer diagnoses dates within each control’s eligible observation window. Prediction endpoints for controls were then sampled from the preceding history with requirement of at least five years of subsequent follow-up. Each retained trajectory contained at least three clinical visits.(Fig 2b; Supplement).

### Fine-tuning using gated low rank adapters trained with time-to-event loss

We introduce a pan-cancer risk head based on a piecewise-exponential model to estimate cancer risk in different horizons ending at 6, 12, 24, 36, and 60 months. For each cancer and horizon, the head combined the GenEHR cancer-token logit with a horizon-conditioned patient state. To obtain this state, we appended a visit token and the RATE encoding of the horizon endpoint to the observed trajectory and passed the augmented sequence through the GenEHR. The head is parameterized to learn a horizon baseline shared across cancers together with a cancer-specific deviation. The shared component stabilizes risk estimates for cancers with fewer outcomes, while the deviation captures cancer-specific rates. The interval specific rates are then integrated over observed at-risk time in the piecewise-exponential likelihood (Fig 2c; Supplement).

We introduced LoRA^33^ to combine shared representations with cancer-specific gates. LoRA specializes the model for a specific task by training a small set of additional parameters while not updating parameters of the pre-trained model. Specifically, the transformer backbone of GenEHR is frozen (not updated) except the additional inserted gated LoRA modules and the pan-cancer risk head trainable parameters. Each module learns a set of low-rank directions shared across cancers and a signed cancer-specific gate for each direction. This approach allows rare cancers with fewer outcomes to benefit from shared representation changes supported by other cancers while centered cancer deviations keep cancer specific information. (Fig 2c; Supplement).

### GenEHR-CancerRisk improves discrimination performance across cancer types

We compared three approaches to pan-cancer risk scoring: (1) direct scoring using the frozen GenEHR cancer-token logits, (2) supervised training of only a pan-cancer risk head over the frozen backbone (‘head only’), and (3) supervised joint fine-tuning of the risk head and cancer-type-specific gated LoRA parameters over the frozen backbone. All comparisons were carried out on the same patient trajectories for 17 cancer types in MGB, 17 in PROV, 13 in VA, 8 in All of US, and 8 in the UKBB, at prediction horizons of 6, 12, 24, 36, and 60 months after the end of the trajectory. In MGB, macro-AUROCs across horizons (see Methods) ranged from 0.71 to 0.77 for direct scoring, 0.80 to 0.84 for head only, and 0.80 to 0.84 for gated LoRA. The respective ranges were 0.60 to 0.67, 0.72 to 0.77, and 0.73 to 0.78 for PROV; 0.67 to 0.71, 0.80 to 0.81, and 0.82 to 0.83 for VA; 0.58 to 0.85, 0.67 to 0.82, and 0.68 to 0.84 for All of US; 0.54 to 0.80, 0.60 to 0.92, and 0.62 to 0.92 for UKBB. All individual cancer performances are in Fig 5.

**Figure 5:**
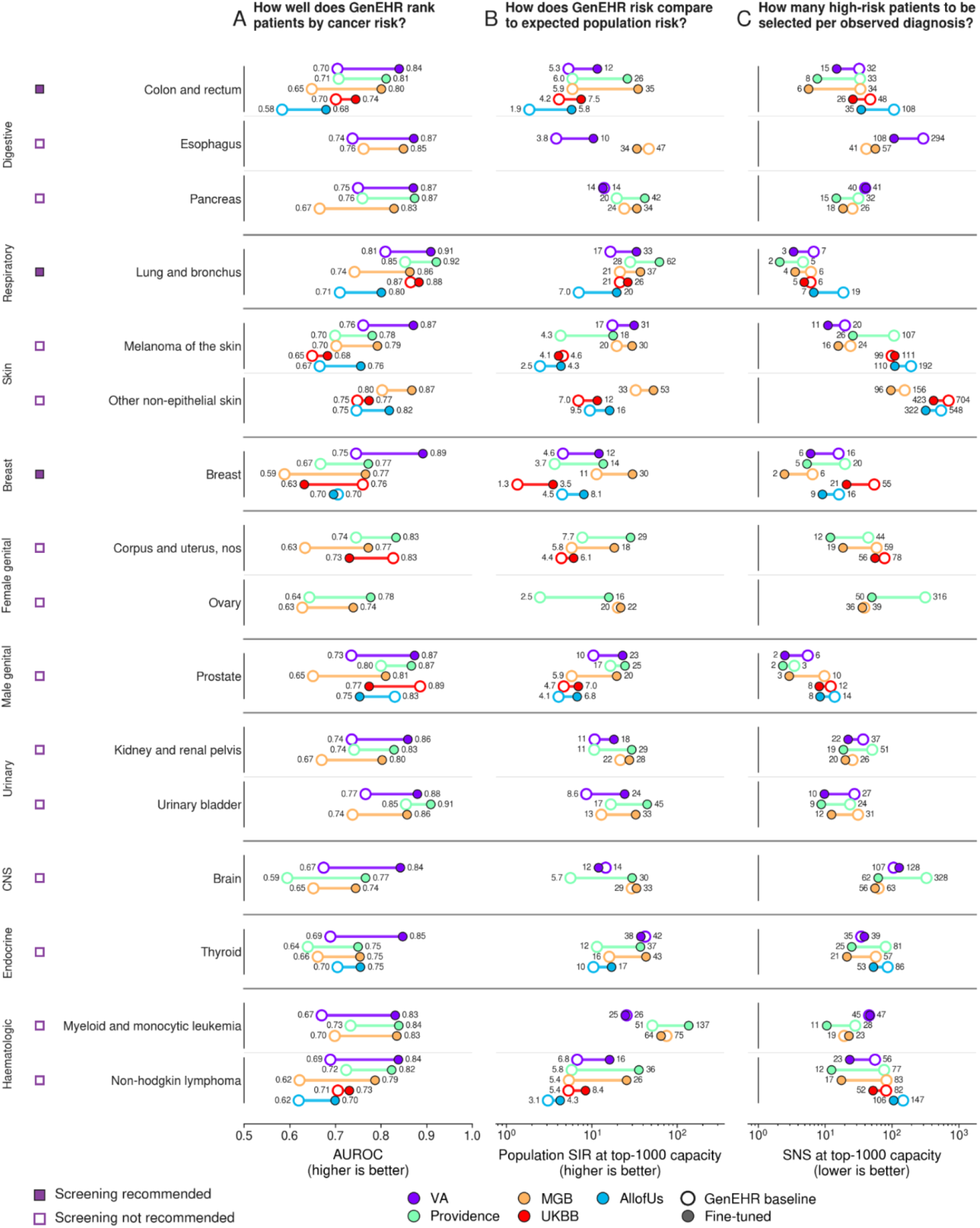
Performance of pan-cancer risk prediction (60 month prediction horizon). (A) Comparison of prediction scores from just the pretrained GenEHR with that from fine-tuned GenEHR for 19 cancer types in 5 health system datasets. Pretrained GenEHR learns cancer-associated signals useful for ranking cases and controls without any supervision from the cancer registry. Fine-tuned GenEHR uses a time-to-event objective, which improves AUROC. (B) SIR (standardized incidence ratio; higher is better) quantifies the increase in prediction performance for the 1000 highest scoring trajectories relative to the population-wide demographic baseline incidence for a given age&sex subpopulation (SEER data^1^). Before computing SIR we adjust for differences in cancer prevalence of respective cohorts and real data (C) SNS (number suggested to screen for one detection; lower is better) here quantifies the estimate as to how many patients are to be enrolled for high-risk screening to find one cancer case. The estimate is based on training on actually detected cancer cases and evaluation on withheld test sets. In new screening programs the number of detected cases will of course depend on the performance of the screening modalities, which may be higher or lower compared to that of clinical work-up in the training sets. Cancers shown are those with 2000 or more patients in at least two cohorts. Results on other prediction horizons (6,12,24, and 36 months) are in Supplement Fig S6a-d.

Direct scoring showed that GenEHR alone can discriminate reasonably well between patients with and without cancer, without cancer-specific supervised time-to-event training. The substantial improvement in risk scores after fine-tuning showed that explicit supervision on diagnosis timing adds information beyond unsupervised direct scoring. Gated LoRA fine-tuning achieved the best overall discrimination across cohorts and prediction horizons (60-months horizon, Fig 5). Results for other horizon windows are included in the supplement (Fig S6a-d). Comparisons with alternative baseline methods on the MGB cohort are in Fig S16. The pan-cancer risk model is well-calibrated (Fig S12, S23, S31, S39, S47). We also report the breakdown of performance by age-bracket and sex (Fig S10b, S21b, S29b, S37b, S45b) together with sensitivity analysis of age-matched and length matched cases and controls (Fig S10a, S21a, S29a, S37a, S45a). A fine-tuned model performs best in prediction performance estimated on withheld data across all comparisons. For cancer with low prevalence, the model relied more on shared representations, while cancer-specific gates contribute more for cancers with more data(Fig S15).

### Focus on highest-risk subgroups to facilitate targeted pre-cancer screening

AUROC assesses prediction accuracy integrated over all operating points but does not tell us how well a model nominates the highest risk cohort for a realistic screening program. This requires choosing an operating threshold that minimizes false positives and maximizes true positives, and also accounts for capacity of the healthcare system and burden of follow-up testing.

We therefore evaluated SIR and SNS at a clinically relevant operating threshold N, typically a small number compared to the size of the population for high-risk focus (Fig 5). A lower SNS implies that fewer patients would need to be evaluated through an AI-informed screening program in order to find one cancer. This number differs from the standard ‘number needed to screen’ (NNS)^47^ metric that assesses the strength of a given screening modality on its ability to reduce cancer mortality. The traditional NNS quantifies not only the sensitivity of a specific screening platform for early stage detection of a specific type of cancer in a specific screening cohort but is also meant to assess the ability to cure that cancer once detected. In contrast, our model estimates the risk enrichment for cancer diagnosis, but not the ultimate effect of therapeutic outcomes. Our suggested SNS estimates also do not take into account the performance of the clinical tests to be used in a newly configured screening program, as the datasets used for model training typically have only limited information about the clinical workup and no information about the number of patients seen with suspected cancer. One can use the suggested number to screen (or the model’s PPV at a given operational threshold) to compare different models, models applied to different cohorts and as a guide for the design of high-risk screening programs. SIR compares observed cancer cases in the high-risk cohort (as predicted by the model, evaluated on the withheld test set) and the expected number of cancer cases based on population wide incidence for a given age and sex. Higher SIR means better enrichment of high-risk cohorts compared to a baseline estimate based on population incidence.

At an operating threshold of N=1000 highest-risk patients, we report cancer-specific SIR, and SNS, across health system cohorts (Fig 5). At the 60-month prediction horizon, SIR, and SNS ranged from 18-224, 2-96 in MGB; 6-224, 2-136 in PROV; 3-91, 6-299 in VA; 3-27, 5-570 in UKBB; and 4-20, 6-322 in All of Us, respectively. For pancreatic, ovarian, and lung cancer, across cohorts SIR ranged from 34-48, 6-22, and 20-57, while SNS ranged from 13-18, 36-136, and 2-7 respectively. Because cancer incidence varies across cancer types and cohorts, we report SIR and PPV as a function of N for all cancer types across sites (Fig S11a-c; S22a-c; S30; S38; S45a-b). The SIR values are substantially higher than standard risk models, even those developed using curated data in specialized cohorts.

For nominated patients with correctly predicted cancer occurrence in the withheld test set, we further look up the time interval between the last clinical event in the observed patient history used for the prediction and the actual recorded cancer diagnosis date in the registry data. We report the distribution of these real-data intervals across cancer types and cohorts (Fig S6e-f). Many flagged cases might have been identified several months to years prior to actual diagnosis. At the 60-month horizon and an operating threshold of N=1000 across cancer types, median lead time ranged 5.5-17.6 months and max lead time 32.6-53.3 months in MGB; median 8.1-26.8 months and max 21.4-58.5 PROV; median 15.2-29.8 months and max 45.2-59.9 months in VA; median 18.9-24.2 months and max 53.2-59.9 months in UKBB; median 10.7-20.2 and 43.2-60 months in All of Us. For highly malignant cancer such as pancreatic cancer the median/max was 17/59.5 months in MGB, 9.3/29.8 in PROV, 15.9/45.2 months in VA. For ovarian cancer, 8.9/31.1 months in MGB and 26.8/47.1 in PROV. These results suggest the model may learn signals corresponding to future cancers which potentially were missed by standard care, suggesting the possibility of significantly widening the detection window, which may allow some patients to be diagnosed at an earlier stage of disease.

### Interpretation of features contributing to cancer risk

We carried out a three-step interpretability analysis (Methods) to identify clinical events contributing most to cancer-risk prediction. The analysis used the fine-tuned model trained with a three-month exclusion and evaluated the 12 and 60-month prediction horizons. First, we used IG to measure the contribution score of each event to cancer log-odds. Rather than using typical <pad> as the IG baseline, we used trajectories from multiple control patients. For each token in a given trajectory, we computed its attribution relative to each control baseline and averaged the scores. We then ranked events within each trajectory and recorded how often each event appeared in the top decile of attributions among true positive patients at the selected N=1000 operating threshold. A set of frequently contributing events shared across multiple cancers (Fig 6A;MGB) and cancer-specific events for pancreatic, ovarian, and lung cancer (Fig 6B;MGB). Second, we tested model sensitivity by masking top ranked medication and procedure tokens one at a time and recomputing log-odds. Finally, transition analysis assessed how often the predicted cancer log-odds increased or decreased when a medication or procedure code was recorded along the trajectory, quantified by (*n*_*up*_ − *n*_*down*_)/(*n*_*up*_ + *n*_*down*_).

**Figure 6:**
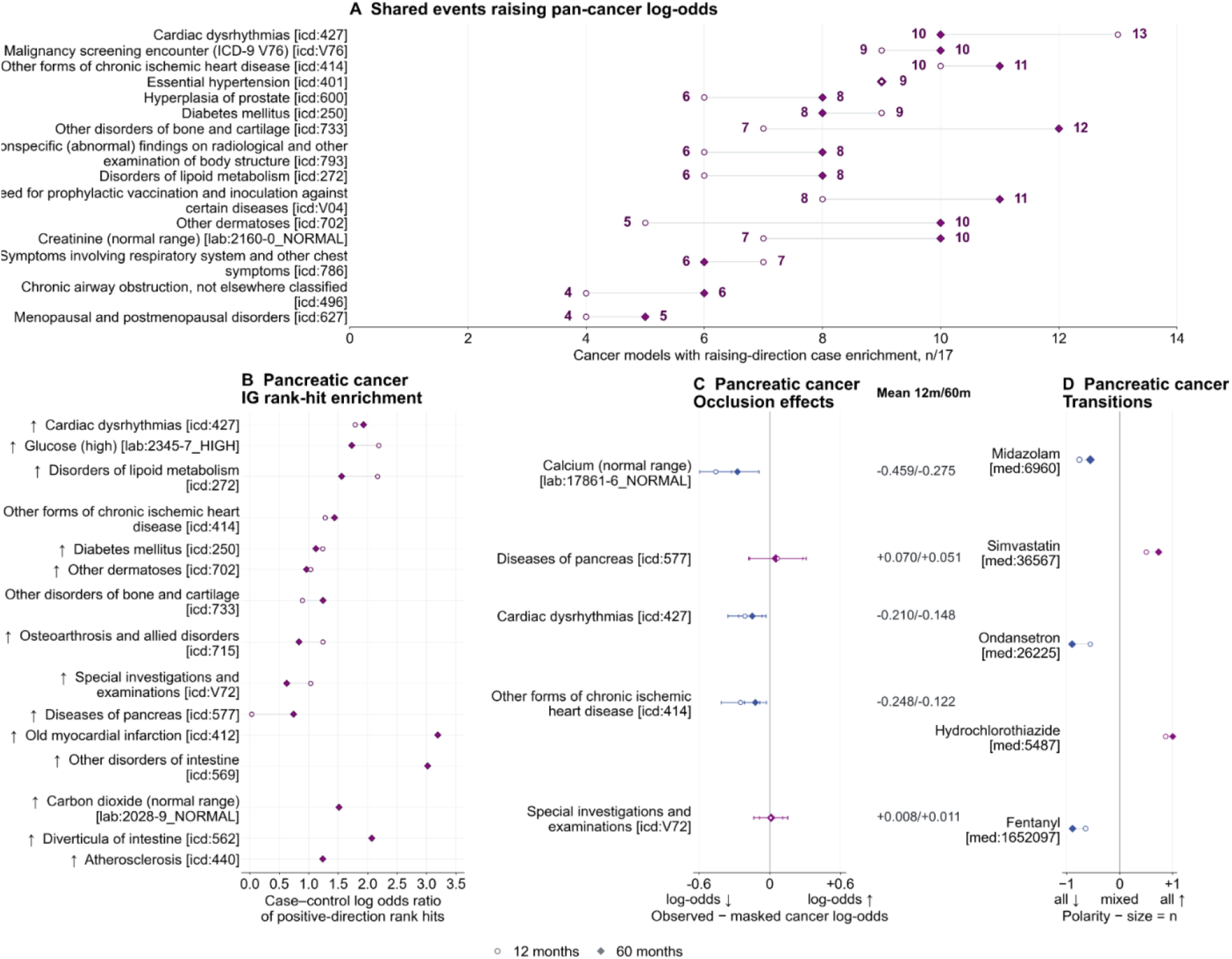
Interpretation of clinical events contributing to cancer-risk predictions in MGB. IG scores computed for the 12- and 60-month cancer-risk models using a three-month exclusion window. (A) Cancer-general features: events which are enriched among top-decile across chosen cancer types (x-axis is the number of cancer types in which an event was consistently ranked high). (B-D) Cancer-specific features for pancreatic cancer. (B) Case-control log2 odds ratio of events ranked at top among cancer patients based on IG score. (C) We mask each eligible (in top decile of trajectory length) occurrence of an event and compute observed minus masked change in cancer log-odds averaged across trajectories. (D) We carry out transition analysis by incrementally adding events to a patient’s trajectory and track transition in cancer log-odds (n_up_) or decrease (n_down_) and transition polarity is then given by (n_up_ − n_down_)/(n_up_ + n_down_). Results on other cancer types in Supplementary Fig S13a-S13f.

Across cancer types, cardiac dysrhythmias, ischaemic heart disease, hypertension, diabetes, lipid disorders, malignancy-screening encounters, abnormal images, and respiratory symptoms are some of the events that increase cancer log-odds (Fig 6A;MGB). For pancreatic cancer, top IG features include high glucose, intestinal disorders, diabetes, lipid disorders, and pancreatic disease contribute most to increasing log-odds and these patterns have been reported in patients with pancreatic cancer^6^. Masking abdominal or thoracic-computed tomography lowered the log-odds of pancreatic cancer at both 12 and 60 month horizons, which indicates imaging probably contributes to the risk score as a diagnostic workup for suspected cancer. Hydrochlorothiazide was associated with upward score transitions for pancreatic and ovarian cancer. Previous studies have associated thiazide use with ovarian cancer incidence, including a possible histotype-specific association^48,49^. In pancreatic cancer patients stopping antihypertensive treatment has been observed before cancer diagnosis^50^ (Fig 6B;MGB). Since EHR data only records when hydrochlorothiazide appears in the trajectory, we cannot determine whether this reflects the drug, or a change in treatment. Ovarian cancer IG features included menopausal disorders, diabetes, family history, and general comorbidity (Fig 6C;MGB). For lung cancer, respiratory symptoms, pneumonia, pleurisy, and other lung disease appear as top events, while chest computed tomography and radiography show larger log-odds with increased risk (Fig 6D;MGB). Detailed results for other cancer types are in Supplementary Fig S13a-S13f and for other cohorts PROV in Fig S24, VA in Fig S32, UKBB in Fig S40, and AoU in Fig S48. Interpreted isolated features should be interpreted with caution, as they actually function in the full context of health trajectories and should not be considered causative. Some of the reported features do agree with prior knowledge, while others provide a basis for hypothesis generation which could be further investigated through causal inference and subsequent clinical trials of preventative interventions.

## Discussion

### GenEHR – an inter-visit gap aware foundation model for pan-cancer risk prediction

We report a new pan-cancer EHR foundation model, with fine-tuning for disease risk prediction, with the clinical goals of improving cancer risk assessment and ‘catching cancer early’. The method combines the power of generative large models, analogous to large language models, with supervised adaptive learning for specific subtasks. Application of GenEHR to large-scale data sets in five different healthcare systems achieves reasonable predictive performance for the probability of a first cancer diagnosis within different horizons ending at 6, 12, 24, 36, and 60 months after the end of the trajectory used for the prediction. While the retrospective performance, based on evaluation in withheld test sets, is encouraging, real-world clinical implementation of screening programs assisted by the model will require prospective testing. We now discuss both strengths, limitations and challenges.

### Clinical utility of risk stratification models

An operating threshold trades false positive rate against true positive rate to minimize the potential harm of follow-up testing^51^. Choosing an optimal threshold is essential for nominating patients for AI-informed screening programs because screening tests and their downstream consequences have both financial and other resource implications. For effective resource utilization, we need to address key operational questions. How many individuals should be selected for AI-informed screening, and what proportion of those will eventually be diagnosed with cancer? We prioritize our evaluation of prediction performance using PPV, SIR and SNS. Here, SNS is an approximation to the number of nominated patients per subsequent cancer diagnosis and does not account for the performance of a screening test. SIR quantifies the risk enrichment within high risk stratum at a given operating threshold compared to a demographically adjusted baseline population incidence. These scores allow us to assess and compare the utility of various models for actionable intervention pathways by guiding patients toward targeted AI-informed screening programs, but admittedly do not capture the full spectrum of heterogeneity inherent in various available intervention pathways for individual cancer types.

### GenEHR improvements over existing autoregressive EHR foundation models

Recent generative EHR models differ in their handling of time intervals and the patient populations used for training. ETHOS^25^ is trained on MIMIC^52^ and evaluated as zero-shot, and CoMET^26^ adopts ETHOS tokenization but is trained at a much larger scale on the Epic Cosmos database. Both represent time intervals using a fixed set of time-interval tokens, so fine-grained temporal resolution is not attained. Delphi^30^ learns next event rates with a competing exponential model trained on first occurrence of ICD codes in the UKBB. Delphi handles time gaps using artificial no-event tokens and its own ablation evaluation shows that the performance depends on the rate at which these tokens are inserted. It also treats each level-3 ICD code as a separate competing outcome, without grouping codes that belong to the same disease. For example, colorectal cancer can be represented by any of the following codes C18, C19, C20, and C26; thus when the model predicts one of these codes, related codes may already be present in the patient’s history and bias risk prediction. MOTOR^28^ fits a piecewise exponential objective, but via a discriminative head rather than in a generative model of trajectories. GenEHR builds on these approaches by first learning time-interval aware patient health state representations through large-scale unsupervised pre-training. Given a patient’s history up to the present, GenEHR can learn how past clinical events interact with the queried time gap to influence their relative cancer risk across future prediction horizons, whereas, a standard Cox model in this setting preserves patients’ risk ranking across horizons. The supervised stage uses cancer registry confirmed labels for pan-cancer risk prediction, excluding patients whose EHR-recorded cancer diagnoses are not confirmed in the registry to reduce noise from EHR coding practices.

#### Screening programs for early cancer intervention

Compared with our earlier work on risk stratification for pancreatic cancer^10,11^, GenEHR affords an opportunity to design and test customized intervention programs for many tumor types. Cancer types differ in incidence, available screening tests, prevention protocols, and the balance between screening benefits and harms. U.S. Preventive Services Task Force (USPSTF) recommends population-wide screening only for breast^53^, colorectal^54^, cervical cancer^55^, and lung cancer^56^ typically conditioned on age (e.g., mammogram every two years for women above age 40; low-dose CT-scan for adults aged 50-80 years with smoking history). Key obstacles to USPSTF recommendations supporting screening for other cancers include the inability to accurately identify the population at sufficient risk and lack of efficient screening tools to adequately visualize or detect an early stage cancer before it metastasizes^57,58^. In addition, some existing screening criteria incompletely capture the at-risk population–for example, only about 50% of patients diagnosed with lung cancer would have qualified for screening prior to their diagnosis^59^. Use of GenEHR could help with the barrier of identifying populations at risk. AI-informed screening in a GenEHR-identified cohort could be analogous to enhanced screening methods currently recommended for those with inherited genetic variants (e.g. in BRCA1/2, Lynch syndrome)^60,61^, family history of cancer or toxic exposure. GenEHR could be applied for real-world population risk stratification using widely available longitudinal data as it reaches or exceeds prediction performance over most of the specialized models without the need for specialized data.

### Prospective evaluation of intervention programs for high-risk subpopulations

Implementation of GenEHR in the clinic would require prospective trials in which risk stratification is followed by a carefully designed intervention program (Fig 7). These trials would test screening and preventive interventions in those at highest risk. The population benefit of AI-informed risk-based screening depends on the rate at which screen-nominated patients develop cancer, and whether screening platforms used in conjunction with the model can clinically detect cancer early enough to alter the disease outcomes. Cost-benefit analyses considering screening capacity, performance, costs, potential screening related harms, and expected gains in healthy life-span or survival will be necessary^62^.

**Figure 7:**
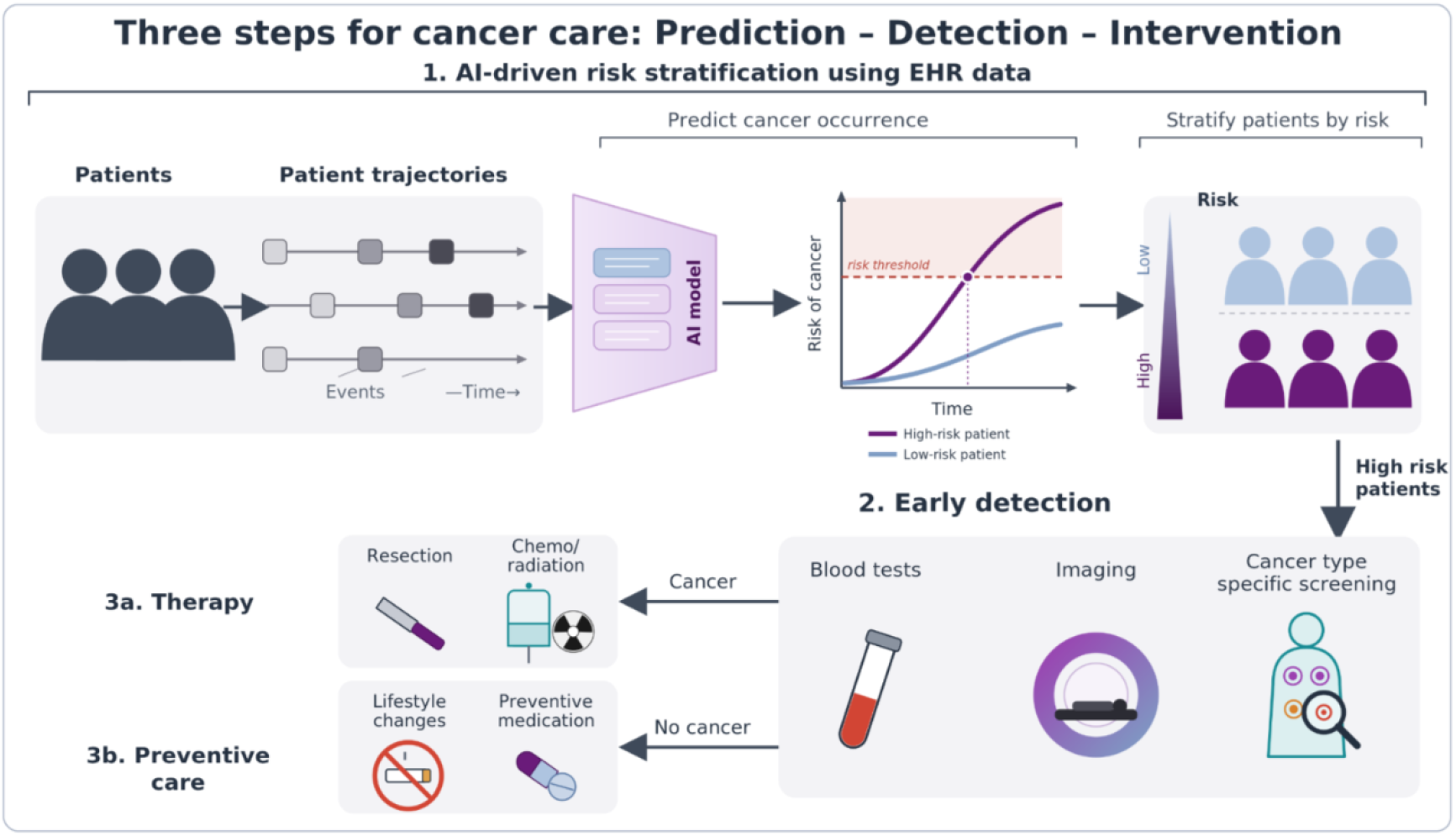
Three steps for getting AI risk stratification into clinical practice. This work addresses the first step in a three step process potentially leading to patient benefit. Early detection in step 2 may be particularly effective in high-risk groups, due to affordable intense workup. If no cancer is detected in screening, the prediction of high risk may still be considered for enrolment in prevention programs.

### Global reach and prospective testing

In our previous work and now here, we have trained and evaluated cancer risk models in cohorts from different countries^10^. The ambition is to train a global model across healthcare systems via local collaborative agreements that provide governed access to protected data and secure computing. We can then successively apply adaptive training on genEHR using location-specific data. We have seen interest in collaborative efforts in clinical settings in, e.g., the US, Canada, the UK, Sweden, Denmark, Turkey and Germany. As such collaborations develop, in the next steps we intend to implement prospective silent evaluation, in which patients will be scored using genEHR without influencing their clinical care^63^. This phase should assess reliability, temporal drift, calibration, and whether retrospective performance holds in the tracked patient population. Because no intervention occurs, this provides a projected SNS that can be used to judge feasibility of operation threshold. These efforts would benefit patients and lighten the economic burden of cancer care.

### Challenges and future outlook

Moving from retrospective risk prediction to clinical implementation requires models that are well calibrated and robust to data noise and dataset shift ^64,65^. Current AI methods can handle some level of noise, but systematic errors or missing diagnoses can bias the predictions^66^. Recording accurate and complete diagnosis remains a major challenge in electronic health record systems. While we have seen higher accuracy in the Danish clinical records, the accuracy of ICD codes in US healthcare systems remains challenging ^67,68^. We have partly addressed this limitation by using cancer registry confirmed outcomes as the labels for cancer risk stratification. Future opportunities include inclusion of free text notes, and more accurate computational phenotyping combining domain-agnostic machine learning, ontologies, or large language models. This would likely incur increased resource use, but it could improve predictive performance and explainability of per-patient or population wide risks for malignancy ^69,70^. An additional challenge within countries without a single payer system is that individuals often seek care within multiple different healthcare systems in sequence, and so integrating trajectories across these different settings can limit the model’s predictive power for those individuals.

Clinical adoption also requires AI-supported screening programs to be acceptable to patients and clinicians, evaluated for clinical benefit and integrated into existing workflows^71^. Predictions from the model should be supported by interpretable reports that clinicians can review when guiding patients to appropriate screening programs^72,73^. AI methods typically perform differently across cohorts when certain demographics groups are unequally represented in training or data acquisition differs, e.g. in specialized cohorts. At the same time, cancer incidence can vary across populations because of environmental, socioeconomic, genetics, and other factors^74^. Performance of AI methods, even if well-balanced in training, should therefore be evaluated across demographic, socioeconomic, and environmental subpopulations with the aim to capture true population specific differences in risk from disparities introduced by data collection or healthcare access ^74,75^.

Cancer risk stratification from EHR data can also benefit from richer longitudinal data collected using standardized measurement units. For example, an annually repeated blood test with detailed profiles of DNA, proteins and metabolites, in which one can analyze changes rather than just snapshots, could be particularly informative ^76,77^. Because molecular profiles of liquid biopsies vary substantially between healthy individuals, within-person changes over time may be more informative than a single measurement^78^. These year to year changes can capture an evolving health state and provide early evidence of changing disease state or risk.

In practice, clinical use of the genEHR-CancerRisk prediction tool is step one of three steps on the way to patient benefit: risk stratification, interception program, and early treatment (Fig 7)^79,80^. A full clinical program to “catch cancer early” is a challenge for a collaborative consortium involving AI experts, technology developers of medical tests, clinical professionals and patients.

## Data Availability

Codebase will be made open source, on github under academic license, restricted commercial use.
Model weights on request from each data-providing institution with the Sander lab at HMS.

## Author contributions

Concepts: A.K., C.S.

Data acquisition and refinement: A.K., E.T.W, (MGB); C.Z., N.F. (VA); D.T.F., Q.W., J.H. (PROV); M.H., A.K. (UKBB); M.H., A.K. (AoU).

Methodology, evaluations, software system: A.K.

Software components: A.K., D.T.F., M.H., C.Z., D.R.

Model runs and evaluation: A.K. (MGB), D.T.F. (PROV), M.H. (UKBB, AoU), C.Z. (US-VA)

Results discussion: A.K., D.T.F., M.H., C.Z, D.R., D.S.M., E.T.W., J.H., N.F., L.V.S., C.S.

Epidemiology: E.T.W., J.H.

Clinical collaborators: E.T.W., L..V.S., A.C., T.S., J.H.

Supervision: J.H., N.F., L.V.S., C.S.

Displayed items: D.T.F., M.H., C.Z., A.K.

Manuscript: A.K., C.S.

Manuscript edits: all authors

## Data and code availability

Open source, on github.

Academic license, restricted commercial use.

Model weights on request from each data-providing institution with the Sander lab at HMS.

## Potential competing interests

D.S.M: Advisor - GenBio.ai, Dyno Therapeutics, Tectonic Therapeutic, Catalio Ventures; Cofounder - Seismic Therapeutic.

Q.W., J.H.: Institutional research support for unrelated work from BMS, Gilead, Janssen, Delfina.

L.V.S.: Institutional grant funding from AstraZeneca, Delfi Diagnostics.

C.S.: SAB Cytoreason Ltd.

## Acknowledgements

We thank Gary Bader, Rob Grant, Brian Wolpin, Abdullah Kuziez, and members of Marks lab for discussions and guidance. We gratefully acknowledge All of Us participants for their contributions, without whom this research would not have been possible. We also thank the National Institutes of Health’s All of Us Research Program for making available the participant data and cohort examined in this study. This research has been conducted using the UK Biobank Resource under application number 53995.

## IRB protocols

Approved protocols at MGB, PROV and VA. For Providence Health and Services, procedures were reviewed and approved by the Institutional Review Board at PROV through expedited review (STUDY2019000389). Patient consent was waived because disclosure of protected health information for the study was determined to involve no more than a minimal risk to the privacy of individuals. For MGB, IRB approval was granted for access to the RPDR research database (Protocol 2024P000851). For VA, IRB approval was granted for access to the VA Corporate Data Warehouse (CDW). For All of US and UK-Biobank standard approval for research use after application.

## Funding for this work

ICI philanthropy (grant to Harvard Medical School (HMS)), US-CDMRP (grant #HT9425-23-1-0464 to HMS & the Boston VA Research Institute), Uniformed Services University/Henry Jackson Foundation #HU00012420063, subaward #67659, ISB (Institute for Systems Biology), and the VA Cooperative Studies Program. The views expressed are those of the authors and do not necessarily reflect the position or policy of the Department of Veterans Affairs or the U.S. government. This project is sponsored by the Uniformed Services University of the Health Sciences (USU); however, the information or content and conclusions do not necessarily represent the official position or policy of, nor should any official endorsement be inferred on the part of, USU, the Department of Defense, or the U.S. Government.

## Methodology

### Data access, cohort preparation, and harmonization

#### Data access and protected compute resources

We used longitudinal EHR databases from Providence Health and Services (PROV), Mass General Brigham (MGB), US Veterans Affairs (US-VA), UK Biobank (UKBB), and the NIH All of Us (AoU) research program. Each dataset was accessed under an approved IRB protocol and data use agreements. Patient level data, cohort analysis, model training and evaluations were all carried out within approved secure computing environments. We did not use any clinical notes or medical images data in this paper. Only structured diagnoses, medications, procedures, laboratory results, demographics, and lifestyle features (where applicable) were used. Cohort specific inclusion and exclusion criteria and counts are in the respective flowcharts (Fig. S1a-S5a).

### Cohort preparation

#### Providence Health and Services

PROV covers 51 hospitals and 1,014 clinics across 7 US states (Alaska, California, Montana, New Mexico, Oregon, Washington, and Texas). We used the EHR snapshot last updated on May 31, 2023, containing 22,863,545 patients. Codes occurring before January 1, 2012 were grouped by setting their dates to December 31, 2011. This constitutes a “bag-of-codes” setting so historical codes (before cohort start) can be modelled, but unreliable longitudinal information is discarded. Patients with death dates before January 1, 2012 were also removed. This filtering resulted in a cohort size of 6,884,118 patients. PROV had a particularly large drop in cohort size as a result of filtering. This is due to the requirement that valid patients have at least one systolic and diastolic blood pressure reading, indicating a meaningful encounter with the PROV health system.

### Mass General Brigham

We used longitudinal EHRs from four MGB sites: Brigham and Women’s Hospital (BWH), Brigham and Women’s Faulkner Hospital (BWFH), Massachusetts General Hospital (MGH), and Newton-Wellesley Hospital (NWH). Under our IRB protocol, the observation period extends from January 1, 2000 through January 1, 2024. The source cohort contained 4,673,744 patients, of whom 4,432,955 patients who have at least one diagnosis, medication, procedure and events. We removed events without a valid date and collapsed duplicate occurrences of the same event on the same day. Patients with invalid birth date or an age of 100 years or older were excluded. This resulted in a cohort of 1,999,232 patients.

### United States Veterans Affairs

We used nationwide EHR data from the VA Corporate Data Warehouse (CDW) covering 1999 through 2022. The initial cohort contained 15,510,644 patients. We removed patients with age greater than 100 years at the end of follow-up. After applying trajectory level filters the final cohort contained 13,187,301 patients.

### NIH All Of Us research program

We used the version 8 of the AoU Curated Data Repository (CDR) Registered Tier. The source cohort contained EHR data of 369,775 patients. We first required patients to have at least three distinct visits with diagnosis codes, leaving us with 312,495 patients, of which 312,181 patients are retained in the final cohort after the filtering steps. The filtering steps and corresponding numbers are laid out in Fig S5a.

### UK Biobank

We used linked UK Biobank health records from 502,409 participants available through April 1, 2022. The source population included diagnosis, lifestyle and demographic codes. UKBiobank provides birth month and year rather than the exact date of birth, therefore, patient age was computed using the available month and year. Dates designated as missing or invalid by UK Biobank were handled according to the data dictionary. If any event date is before the birth date of a patient, it is replaced by the patient’s birth date. If any date lies in the future, i.e. after the cohort cut-off date, then it is removed. Certain special (past) augmented diagnosis dates in the UK biobank (‘1901-01-01’,’1902-02-02’,’1903-03-03’) are replaced by the respective patient’s birthdate and diagnosis with two other special dates (‘1900-01-01’ and ‘2037-07-07’) are excluded since they indicate issues with the source data^36^. The final cohort contained 482,920 participants after filtering steps. The filtering steps and corresponding numbers are laid out in Fig S4a.

### Data harmonization

We implemented a common data harmonization and tokenization scheme across all cohorts. Representing each event using a standardized concept code, its event type, and its recorded date. Events occurring on the same date were assigned to the same visit and duplicate occurrences of the same concept within a visit were collapsed.

### Diagnoses

Diagnoses were represented using International Classification of Diseases Clinical Modification (ICD-CM), either version 9 or 10. ICD codes were truncated to the three character category level, reducing sparsity while preserving the major clinical events. This process resulted in a final set of 232,464,302 diagnosis events in PROV; 78,801,262 events across 2,700 categories in MGB; 2,255,262,266 events in VA across 2,853 categories; 5,879,445 events across 1,259 categories in UKBB; and 100,290,151 across 2561 categories in AoU.

### Medications

Medication records were represented using the RxNorm concept unique identifier (RxCUI). Because RxNorm assigns separate codes to different variants of the same drug (dosages or brand names) resulting in multiple codes for therapeutically similar drugs.

Therefore, we mapped each code to its respective ingredients. Drug to ingredient mapping for each RxNorm code was obtained by querying the RxNav web resource^81^ for ingredient (term type “IN”) and precise ingredient (term type “PIN”). For obsolete or remapped codes, we used RxNav to identify historical IN and PIN mappings where possible. We removed ingredient codes with a low number of orders to ensure each code had a sufficient number of occurrences for robust modelling and to avoid increasing the model vocabulary size unnecessarily. To do this, we ranked ingredient codes by their most to least frequent occurrence in the cohort, and took the top K codes such that at least 99.5% of all orders were covered. The final medication data included 1,885 ingredient RxCUIs and 1,208,554,728 ingredient-level events in PROV, 1,084 ingredients and 54,322,402 events in MGB, and 2,296 ingredients and 145,487,656 events in AoU. Medication records were not included in the UKBB and US-VA cohorts.

### Procedures

Procedures include medical services such as surgeries and radiology, which were represented using Current Procedural Terminology (CPT) codes or ICD10 Procedure Coding System (ICD10PCS). We filtered out any CPT codes which were infrequent by retaining a code set covering 99.5% of all procedures. This resulted in 2,589 CPT codes covering 178,115,584 procedure events in PROV; 1,786 (ICD10PCS or CPT) codes covering 111,518,760 events in MGB, and 251 procedure codes covering 1,015,214 events in AoU.

### Lab tests

Laboratory tests were represented using the Logical Observation Identifiers Names and Codes (LOINC). LOINC consists of a broad range of measurements, such as blood tests, physical exams and patient surveys. We filtered LOINC codes to only include quantitative laboratory concepts with a usable value, unit, and reference range. Lab values and thresholds were frequently entered as free text. For example, “negative” or “absent” could be used to indicate lab values of 0, “10-20” could indicate a range in which the measured value lies, and “<5” or “>30” could indicate specific value thresholds. We harmonized these entries by replacing free text values with numeric values where possible, took the mean of values with ranges, and removed any entries that could not be harmonized.

For PROV and AoU the majority of these test results were within the reference range. To prevent frequent normal measurements from dominating patient trajectories, we retained only values outside the recorded reference range. Each abnormal lab value was tokenized with the corresponding LOINC code along with a “low” or “high” indicator, specifying whether the value is less than the low threshold or greater than the high threshold, respectively. We then retained the smallest set of LOINC codes covering at least 99.5% of abnormal laboratory events. This resulted in 357 LOINC codes covering 345,660,408 events in PROV, 537 codes covering 190,198,452 events in MGB, and 404 codes covering 32,410,269 events in AoU.

### Lifestyle and demographics

In addition to longitudinal clinical events, we included available birthdate, sex, race, ethnicity, and lifestyle as cohort-specific static tokens. Ambiguous or missing categories were represented as “Unknown” tokens. PROV additionally included Area Deprivation Index (ADI) that groups 17 socioeconomic measures into a national index ranging from 1 (lowest deprivation) to 100 (highest deprivation) across U.S. census block groups^82^. Since PROV covers 7 U.S. states, we used the national rather than state-specific index to ensure comparability across patients in different states. We assigned ADI by matching each patient’s census block group identifiers to the ADI dataset and grouped values into 10 equal-width bins. Unmatched ADI values were set to “Unknown”.

Lifestyle information was available in PROV, AoU, and UKBB but not in VA and MGB due to institutional IRB restrictions. Lifestyle variables included body mass index (BMI), tobacco use and alcohol consumption. For each patient, we used the closest measurement or survey date to the cohort start for BMI measurements, tobacco use and alcohol consumption. BMI was tokenized into underweight (less than 18.5), healthy weight (18.5 to less than 25), overweight (25 to less than 30) and obese (30 or greater) categories according to National Institutes for Health BMI ranges^83^. Patients without a BMI measurement were tokenized as “Unknown”. Tobacco use was recorded as current smoker (“Yes”), former smoker (“Quit”), occasional tobacco use (“Passive”) and never smoked (“Never”). Alcohol consumption was recorded as current consumption (“Yes”), previous consumption (“Not currently”) and no alcohol consumption (“Never” and “No”). Tobacco use and alcohol consumption were likewise tokenized using these categories, as well as an “Unknown” token for patients without a survey entry (e.g. recorded as “Not asked” or a missing value).

### Time-ordered patient trajectory

For each patient, the available diagnoses, medications, procedures, and laboratory events were combined into a single time-ordered sequence. Events occurring on the same date formed a visit, and duplicate codes within a visit are removed. Consecutive visits were separated by a RATE token representing the elapsed time between them. To ensure sufficient longitudinal history, we restrict the cohort to patients with at least three distinct visits. We then rank patients by trajectory length and exclude trajectories longer than the 99.9th-percentile threshold that typically reflect data-quality artefacts. The resulting patient trajectory consists of available demographic and lifestyle tokens, followed by a sequence of visits and inter-visit time intervals.

### Cancer definition and linking to cancer registry

For pan-cancer risk scoring we used confirmed cancer outcomes from linked cancer registries. We constructed a separate fine-tuning cohort for PROV, MGB, US-VA, and UKBB, by matching cancer outcomes in EHR and registry. Registry uses ICD-O-3 site and histology code pairs to uniquely determine a cancer diagnosis, or histology code alone for hematological cancers without a site code. We mapped each record in the registry to a SEER cancer group using the SEER site recode cancer definitions^84^. This definition is used in epidemiological cancer incidence reporting which allows us to compare high risk cohort enrichment with population wide incidence. For each patient EHR trajectory, we also mapped ICD-9-CM and ICD-10-CM cancer diagnosis codes to the corresponding SEER cancer group label (Tab S1). We removed cancers of neuroendocrine pancreas with ICD-O-3 site code C254 from the registry to be consistent with previous PDAC risk prediction work^10,11^.

Patients can have multiple cancer codes in the EHR or multiple cancers recorded in the registry, and the dates of these events can differ. To determine an appropriate label for first cancer diagnosis, we first identified a matching pair of SEER cancer group labels between a patient’s EHR trajectory and registry records, and then used the cancer with the earliest registry date as the first cancer outcome. Patients with cancer recorded exclusively in the EHR, only in the registry, or mismatched diagnoses between EHR and registry were excluded from the cohort because the cancer status of these patients could not be confirmed. Discrepancies in cancer occurrence dates between the EHR and registry can occur because, for example, a suspected cancer diagnosis was entered into the EHR before confirmation in the registry, or conversely, that the diagnosis was not noted until after histology confirmed the cancer. To address these discrepancies, we set the cancer diagnosis date in the EHR trajectory to the earlier of the EHR and registry dates. We excluded any patients with a cancer diagnosis date occurring before the cohort start date. This process resulted in a final set of 165,951 cancer patients in PROV; 133, 960 patients in MGB; 955,307 patients in US-VA, and 103,037 in the UKBB.

To ensure sufficient samples for reliable evaluation, we limited our fine-tuning analysis to a set of cancers which have at- least 2,000 patients across the entire data-set. We used cohort subsets for site specific fine-tuning and evaluation:

#### MGB

Breast; prostate; colon and rectum; lung and bronchus; melanoma of skin; thyroid; corpus and uterus; non-Hodgkin lymphoma; kidney and renal pelvis; urinary bladder; brain; pancreas; ovary; esophagus; soft tissue including heart; other non-epithelial skin; and myeloid and monocytic leukemia.

#### PROV

Colon and rectum; pancreas; liver & intrahepatic bile duct; lung & bronchus; melanoma of the skin; breast; corpus & uterus; ovary; prostate; kidney & renal pelvis; urinary bladder; brain; thyroid; myeloma; lymphocytic leukemia; myeloid & monocytic leukemia, and non-Hodgkin lymphoma.

#### US-VA

Breast; prostate; colon and rectum; lung and bronchus; melanoma of skin; thyroid; non-Hodgkin lymphoma; kidney and renal pelvis; urinary bladder; brain; pancreas; esophagus; and myeloid & monocytic leukemia.

#### UKBB

Other non-epithelial skin; female breast; prostate; melanoma of skin; colon and rectum; lung and bronchus; corpus and uterus; non-Hodgkin lymphoma.

#### AoU

Other non-epithelial skin; female breast; prostate; melanoma of skin; colon and rectum; lung and bronchus; thyroid; non-Hodgkin lymphoma.

### Population cancer incidence

For US cohorts, we used SEER incidence rates to compute reference estimates and SIR scores^1^. For UKBB, cancer incidence rates were obtained from the NHS England National Disease Registration Service (NDRS) “Cancer registration statistics” dashboard^85^, which provides accredited official statistics on annual cancer registrations in England, including counts and crude and age-standardised rates for combinations of cancer site, geography, sex, age group and calendar year. We group the cancer incidence rates according to the sex, averaged over all the years available and the cancer type to get the overall sex, age and cancer specific incidence rates.

### Train, validation, and evaluation splits

For each cohort, we partitioned patients into two disjoint sets one for model development and another for held-out evaluation. We withheld 1M patients from PROV, 500K from MGB, 1M from US-VA, 75K from UKBB, and 50K from All Of Us. The remaining cohort was split into 80-10-10 at patient-level, where we use 80% for training, 10% for validation, and remaining 10% for calibration purposes. Respective cohort stats for both GenEHR and for fine-tuning are in Fig 2.

### GenEHR foundation model: Generative model of patient health histories

We introduce a generative foundation model trained to model the conditional distribution of the next health event and its corresponding time given patient history. The same learned distribution supports next visit prediction, disease risk scoring with respect to a pre-specified time horizon, as well as trajectory forecasting. We use an autoregressive transformer to model longitudinal EHRs as heterogeneous sequences containing static conditioning tokens, clinical event tokens, visit delimiters, and compositional time-gap tokens. We separate the observed clinical record from its tokenized representation because visit delimiters and gap placeholders are model control symbols rather than clinical events.

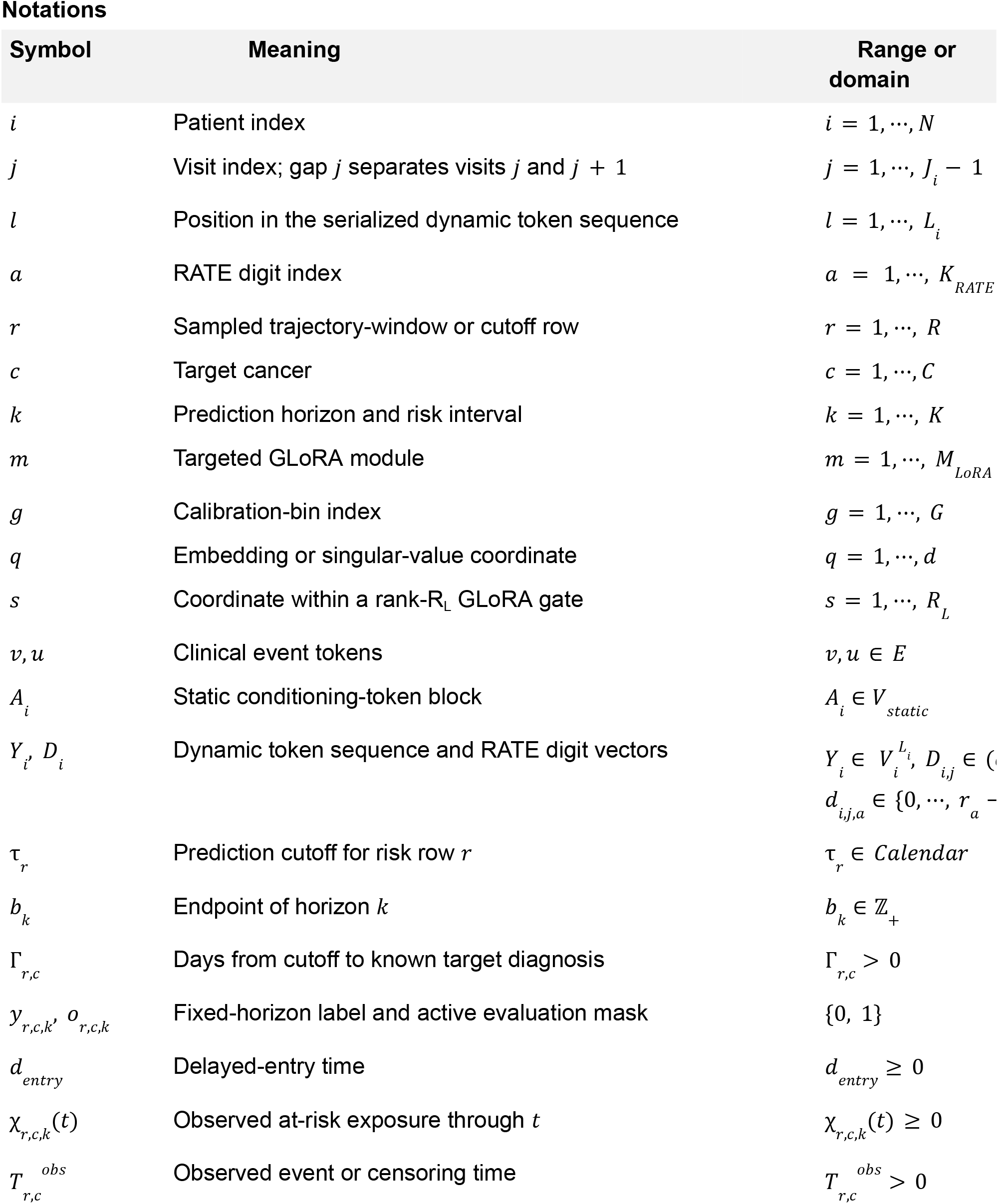
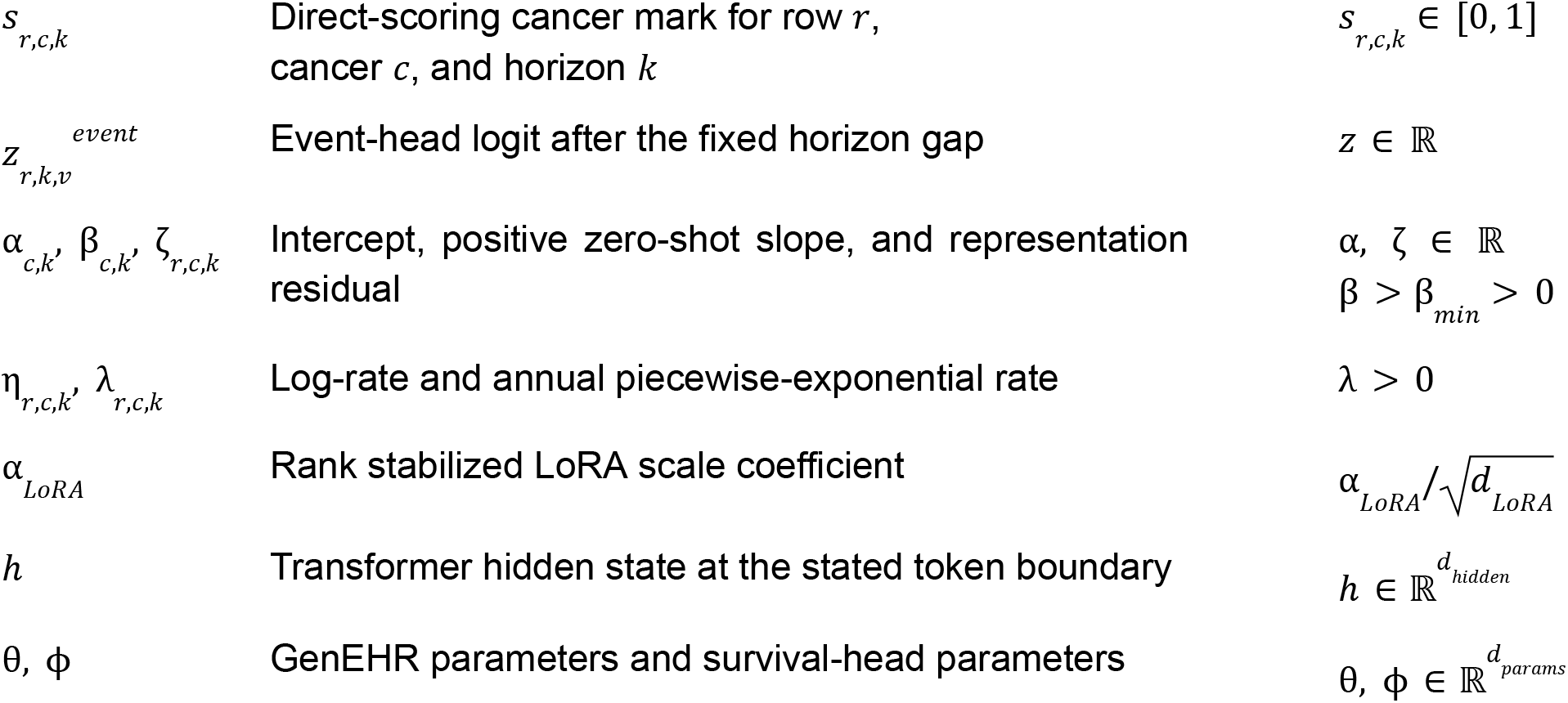

### Representation of patient health record

To model the evolving trajectory of a patient’s health, we represent their longitudinal history as a chronological stream of discrete tokens encompassing demographic descriptors, clinical visits, recorded medical events, and varying time gaps between consecutive visits. For patient *i*, let 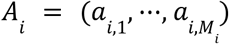 denote the ordered static input token block that includes sex, race, ethnicity, and initial age bucket. Let 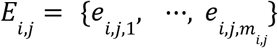 be the set of clinical events recorded at visit *j* with timestamp *t*_*i,j*_, where visit dates satisfy 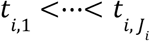. All events form a vocabulary *E*_*event*_= *E*_*diagu*_*∪ E*_*med*_ *∪ E*_*proc*_ *∪ E*_*diagu*_, where *E*_*med*_, *E*_*proc*_, *E*_*lab*_, and *E* are a unique set of diagnoses, medications, procedures, and lab measurements. The inter-visit gap after visit *j* is Δ_*i,j*_ = *t*_*i,j*+1_ – *t*_*i,j*_ . Let 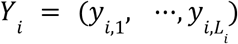 denote the tokenized sequence and *X*_*i*_ be the transformer input,

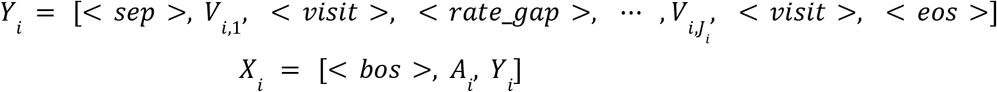

where < *bos* > is beginning-of-sequence token, < *sep* > is a separator token (between static and dynamic tokens), < *visit* > is an end of visit token, < *rate*_*gap* > is a token for an inter-visit time interval and its token embedding is replaced by the learned mixed-radix embedding of patient record. Δ_*i,j*_ . The final visit is followed by < *eos* > token that captures the end of the patient record.

### Conditional generative model with static tokens and time gaps

The static prefix is observed conditioning information rather than a random sequence generated by the model. Let *A*_*i*_ be the observed static prefix, *Y*_*i*_ be the dynamic clinical and structural tokens, for a time gap 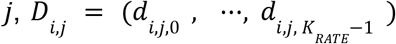be its *K*_*RATE*_ -digits, and *g*_*i,j*_ be the token position of the associated gap token. At token position *l, D*_*i*,<*l*_ contains only gaps whose placeholders precede that position *l*. Let 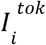 be the non-padding dynamic positions included in the loss function and 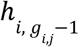 be the transformer hidden state immediately before gap *j* is drawn. Conditional on *A*_*i*_ the joint distribution is factorized as,

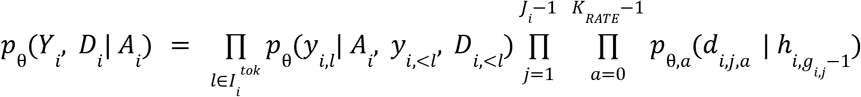

The first product scores clinical and structural tokens and the second scores the *K* digits of each gap from the shared pre-gap hidden state of the transformer. The digit heads are conditionally independent given the shared history (this is a modeling assumption, not that time scales are marginally independent).

### Radix time encoding (RATE) for irregular inter-visit time gaps

EHR data consists of irregular inter-visit gaps, two visits separated by a day or by a year have different implications in terms of severity of health state. We introduce RATE to represent time gaps between consecutive visits with a fixed number of categorical predictions while capturing exact day resolution over the supported interval. For radices 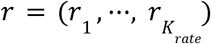, let the place values be 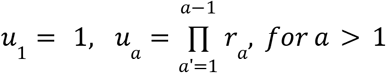. Then, an observed time gap Δ_*j*_ is a composition of a digit vector 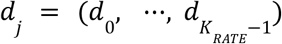 such that

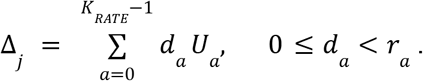

We used *r* = (7, 4, 6, 5, 5), giving *u* = (1, 7, 28, 168, 840) days and range *R* = 4200 days. Any time interval of 4, 200 days or longer were clamped to 4, 199 days. For digit vector *D*, the embedding inserted at a gap position < *rate*_*gap* > is constructed compositionally (look up a learned embedding for each digit *E*_*a*_ [*d*_*a*_], apply a learned digit scale, sum the digit vectors, and pass it through layer normalization (*LN* (.))^86^ and a two layer projection,

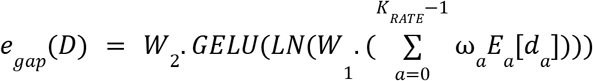

where *W*_1_, *W*_2_ are projection matrix, and *GELU*(.) is a non-linearity. A worked example further clarifies the placement of static, visit and gap tokens. Consider a patient with static tokens female, black, non-hispanic, and age bin 65-69. Let’s say on January 10 the patient had hypertension and a complete blood count, on February 15 the patient had iron-deficiency anemia and ferritin testing, and on May 1 the patient underwent a colonoscopy. The day gaps are 36 and 75 days, with *r* = (7, 4, 6, 5, 5), 36 days with digits (1, 1, 1, 0, 0) as 36 = 1 × 1 + 1 × 7 + 1 × 28; 75 days with digits 75 = (5 × 1 + 2 × 7 + 2 × 28). The full trajectory is (5, 2, 2, 0, 0) as [< *bos* >, *sex*_*female, race*_*black, ethnicity*_*non*_*hispanic, age*_65_69, < *sep* >, *icd*_401, *lab*_58410_*normal*, < *visit* >, < *rate*_*gap* > [1, 1, 1, 0, 0], *icd*_280, *lab*_22760_*low*,< *visit* >, < *rate*_*gap* > [5, 2, 2, 0, 0], *proc*_45378, < *visit* >, < *eos* >]

### Training objective

The training objective combines a standard autoregressive token log likelihood (in analogy to language modeling) with a mean digitwise cross-entropy for valid inter-visit gaps. The event token loss is the negative conditional log-likelihood over scored positions,

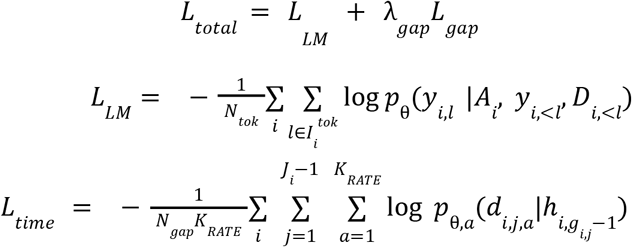

Here,*N*_*tok*_ is the total number of scored token positions over the batch,*N*_*gap*_ is the number of valid inter-visit gaps and λ_*gap*_= 1 is a hyperparameter.

### Pan-cancer risk prediction

#### Trajectory sampling to answer how far back in time the model can predict risk

A patient’s risk changes as new events are encountered over time. That is a patient may be at low risk at an earlier cutoff and high risk later. We therefore use a sampling procedure to draw several cutoff trajectories per patient rather than using a fixed representation of their entire clinical history. Let *r* be an index of a sampled cutoff trajectory, *c* a cancer type, and *k* an ordered horizon index, *i*(*r*) be the patient attached to trajectory *r*, and τ_*r*_ be the prediction cutoff.

Let *T*_*i*(*r*),*c*_ be the diagnosis date of first occurrence of cancer for patient with trajectory *r* and cancer *c*, and Γ_*r,c*_ be days from cutoff to the first target-cancer diagnosis, *U*_*r,c*_ be observable target-cancer free follow-up, *d*_*entry*_= 3 months be the delayed-entry boundary. For horizon *b*_*k*_, the cumulative label and observability mask are,

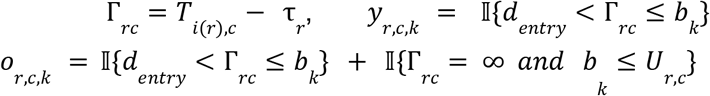

where *I*{.} is an indicator function, the indicator in *y*_*r,c,k*_ is true when diagnosis occurs after the cutoff and before the horizon. For a patient whose cancer occurs within the maximum prediction horizon, all cumulative bins are observed (bins before the event are 0 and the event bin and all later bins are 1). If a patient doesn’t have an observed target cancer, only horizons whose right edge is no later than the censor boundary enter the loss. We use a prespecified prediction interval endpoint of 3, 6, 12, 24, 36, and 60 months.

Only the first recorded cancer group is treated as positive. In case registry data is not available the first chronological occurrence within the health trajectory is used instead. If an event timestamp in the cancer registry occurs after the EHR timestamp, the entry is adjusted to align with the registry, ensuring that no other cancer codes can precede the confirmed diagnosis. Each sampled trajectory is considered valid if it contains a minimum of three events to ensure sufficient clinical context for prediction. Positive trajectory cutoffs had to be more than three months before diagnosis. For healthy control patients, we enforce a minimum follow-up after the cutoff, and for cross-task controls (a patient with another first cancer could be a control sample) cutoff was at least two years before their first cancer. These constraints aim to ensure that the model learns predictive risk factors rather than exploiting quasi-symptoms that may appear right before cancer diagnosis.

To allow the model to learn from a patient’s evolving health status, we sample multiple trajectories from the set with valid cut-off points. Since clinical events cluster more densely immediately preceding a cancer diagnosis, to avoid bias we use a quartile sampling strategy. Specifically, we partition temporal interval from the timestamp of the final event prior to diagnosis *t*_0_, back to *t*_0_ − 5 years prior into four disjoint quartiles and sample one endpoint from each quartile. This approach provides a longitudinal signal of how a patient who is initially classified as not at risk within specific horizons can progressively develop risk factors over time.

### Scoring cancer risk using pre-trained GenEHR

#### Fixed-gap point score

For risk trajectory *r* with cutoff τ_*r*_, let *C*_*c*,9_ and *C*_*c*,10_ be the target cancer token sets in ICD9 and ICD10, for horizon *b*_*k*_, we append its respective RATE encoding to obtain the logit *z*_*r,k,v*_ = *f*_θ_ (*X*_*i*_ (τ_*i*_), < *visit* >, *RATE*(*b*_*k*_)) and map it to event-normalized cancer risk score,

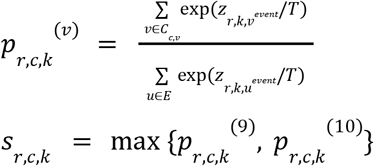

By taking the maximum score the model extracts cancer signals from the most strongly contributing coding systems (ICD9, ICD10).

### Parameter-efficient fine-tuning for improving cancer risk prediction

GenEHR learns cancer associated signals by forecasting the next clinical event, but it is not explicitly optimized for time-to-event prediction or to account for patients whose records end before cancer is observed. We therefore use PEFT to adapt the pretrained state with a piecewise-exponential likelihood, following each patient from the prediction cutoff until cancer diagnosis or the end of available follow-up.

We compare three approaches: (1) the direct scoring using frozen zero-shot decoder, (2) a prediction head-only adaptation for pan-cancer risk, and (3) joint optimization of head and pan-cancer low-rank adapters. Next, we discuss specific configurations for training and validation.

### Monotone cumulative-risk scoring using a time-to-event objective

For each trajectory *r*, cancer *c*, and interval *k*, the log-odds are computed from the frozen model’s output over event tokens,

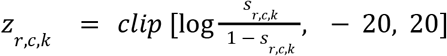

here *clip* is a boundary function to truncate extreme tails which prevents unstable log-rates and extreme gradients. A cancer-interval-specific log-rate is

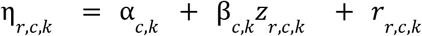

Here, α_*c,k*_ is a baseline log rate intercept and the coefficient β_*c,k*_ learns the contribution of the GenEHR score to the interval rate. Introducing a separate baseline and intercept for each cancer can make learning unstable for rare cancers, we therefore decompose these components as,

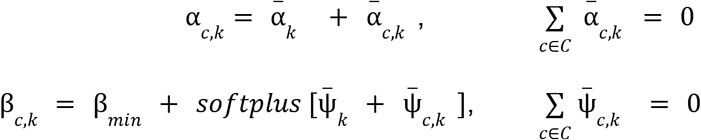

The zero-sum constraints decouple shared and cancer specific components. Without this, a constant could be added to the shared term and subtracted from every cancer deviation without changing any effective baseline or slope. The shared term 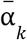 is a learnable baselinelog-rate for interval *k* and 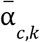 is the centered cancer-specific deviation. The shared parameter 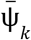 and deviation 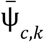 determine the cancer-specific slope adjustment on the fixed point score of the foundation model. The *softplus* ensures β_*c,k*_ > β_*min*_ > 0, so the fitted interval rate is non-decreasing in *z*_*r,c,k*_ when the representation residual is held fixed.

#### Low rank residual representation

We further introduce a low-rank projection matrix 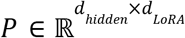 where *d*_*LoRA*_ << *d*_*hidden*_, normalizing each column of *P* by its norm 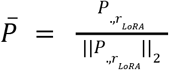. This low rank formulation *P* captures primary shared risk directions by using supervised signal across all cancers, requiring only *d*_*LoRA*_ residual directions per cancer type. Let 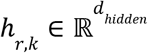 be the horizon conditioned trajectory representation, then the resulting cancer risk residual is formulated as,

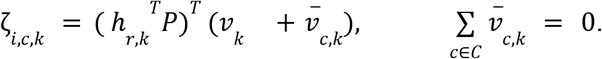

*v*_*k*_ controls the importance of feature across time-intervals 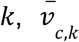 determines the weighting for cancer *c*. Sharing the parameter allows the model to capture risk factors with common temporal effects (short or long term), while centering decouples the global shared effect from the cancer-specific residual difference.

#### Piecewise-exponential likelihood for time-to-event

Let *d*_*entry*_ be the delayed entry in days, 0 = *b*_0_ < *b*_1_ <…< *b*_*K*_ be the rounded-day endpoints, *L*_*K*_ be the effective start of interval *k, W*_*k*_ be width of interval, and χ_*r,c,k*_ (*t*) be an indicator of whether cancer *c* has occurred in interval *k* through event or censoring time *t*,

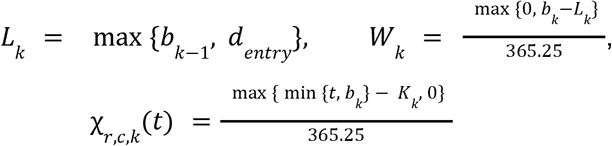

We use exact survival likelihood for the fine-tuning stage. The final log-rate is defined as,

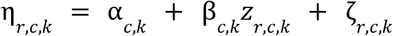

and respective cancer event intensity as λ_*r,c,k*_ = exp(η_*r,c,k*_). Let δ_*r,c,k*_ indicate the exact target event occurred in interval *k*, and *T* _*r,c*_^*obs*^ be the exact event or censoring time truncated at *b*_*K*_ ; then per trajectory cumulative hazard function is

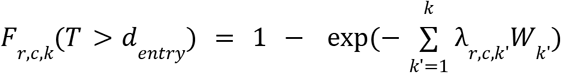

Since, the exposure is measured in years so λ_*r,c,k*_ is an annual rate. The likelihood uses exact event and censoring times which handles unequal follow-up and maintains a strong gradient for rare events. The full risk is given by,

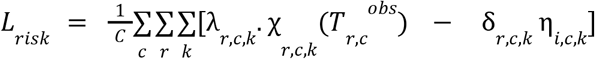

We further apply *L*2 regularization to the shared and cancer-specific parameters. The above formulation is an extension of a standard piecewise-exponential survival likelihood^87^, which has been used for structured EHR time-to-event modeling^28^. Our paper extends this formulation with a delayed entry, pooled calibration of frozen GenEHR and a shared low-rank representation residual.

### Pan-cancer fine-tuning only learning time-to-event head on GenEHR representation

In head-only mode, we restrict training to the cancer-risk predictor that includes cancer-specific embeddings, horizon-related biases, and linear terms within a single prediction head for each cancer type. Let θ_*b*_ denote the frozen GenEHR parameters and ϕ denote the trainable cancer risk head parameters. For patient *i*, a representation for prediction horizon τ_*k*_ is obtained by appending a visit token and the corresponding pre-trained temporal gap encoding

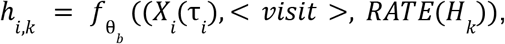

*h*_*i,k*_*∈ R* is treated as a fixed patient horizon representation. Since the backbone is frozen 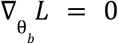 and the optimization is,

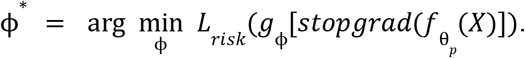

The sample trajectory cutoffs, prediction horizons, labels and censoring rules are the same as the one used in GenEHR evaluations.

### Pan-cancer gated low-rank adapter for improving pan-cancer risk prediction

Many cancers share risk factors that can be captured by a compact set of representation directions, with cancer-specific signed weights determining the contribution of each direction. For example, a cancer similarity graph derived from cPMI shows stronger relationships among cancers in related organ systems. Training a separate LoRA adapter for each cancer does not exploit the shared structure of pre-cancer trajectories and may provide poor estimates for cancers with relatively few cases. On the other hand a single adapter shared across every cancer patient cannot represent cancer-specific factors. We therefore use a variation of LoRA^88^ that adapts weights accounting, at least to some extent, for both shared and specific factors.

Unique to our paper we introduce zero-sum constraints to account for shared gate and cancer deviations in pan-cancer risk modeling. Let 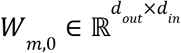 be the frozen weight matrix, 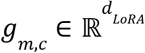 be a cancer-specific gate of LoRA module *m*; we then write the adapted weights as,

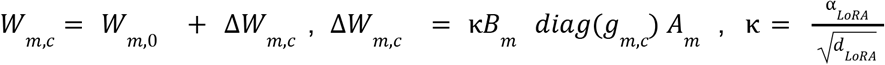

where α_*LoRA*_ is a hyperparameter that controls overall magnitude of the low rank update, *d*_*LoRA*_ is a rank of *LoRA* matrices, and 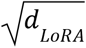 stabilizes the scale when the adapter rank changes. So κ directly controls the signed magnitude of feature component *j* for cancer *c*. The gate *g*_*m,c*_ for LoRA is decomposed into a shared component and a cancer specific deviation term,

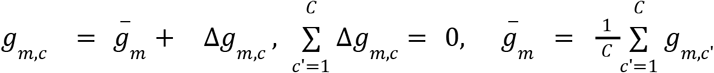

Introducing zero-sum constraint makes this decomposition unique. Specifically, 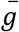 is the mean gate across cancers and Δ*g*_*m,c*_ is each cancer unique deviation from that mean. We further apply partial pooling to the cancer deviations because rare cancers otherwise receive weekly estimated cancer specific updates. This is implemented as a penalty over LoRA modules *M*_*LoRA*_,

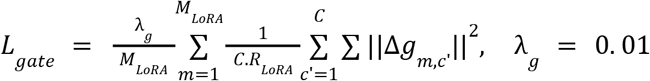

#### Control-trajectory subsampling for efficient finetuning

We implement a control subsampling approach to reduce the cost associated with the highly imbalanced fine tuning objective. Every trajectory with a finite-horizon event is kept and each control trajectory is selected independently with a probability *q*_*ctrl*_. Let π_*i,c*_ be the inclusion probability for a likelihood trajectory (*r, c*), and let 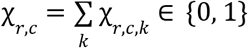 indicates whether a trajectory contains an observed event. Then,

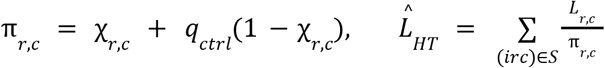

To match the expectation of the complete cohort during training, we apply Horvitz-Thompson (HT)^89^ weighting to scale down the mini-batches via a normalized weight 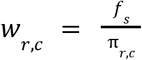, where 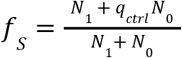, where *N*_1_ and *N*_0_ are event and control row counts.. This normalization factor approximately preserves the scale of the complete cohort mean objective under trajectory sampling.

### Evaluation of performance, reliability, and interpretability of GenEHR and GenEHR-CancerRisk

We first evaluate the generative capability of our EHR foundation model, measuring its calibration, reliability, representation quality, and generative fidelity in modeling clinical sequences. Next, we evaluate the task-specific performance of the fine-tuned model, including its predictive performance across multiple cancer types, its clinical utility in identifying high-risk cohorts, and its interpretability through attribution and counterfactual analyses. All score names, purpose, and interpretation are listed in Table 2.

**Table 2:** Breakdown of evaluation metrics and their interpretation in the context of this paper.

| Score | Purpose | Direction |
| --- | --- | --- |
| Event-only perplexity | Teacher-forced likelihood after renormalizing over clinical event tokens. | Down |
| Top-K event accuracy | Fraction of observed clinical events ranked within the first K event tokens. | Up |
| Mean reciprocal rank (MRR) | Average reciprocal rank of the observed event over the event vocabulary. | Up |
| Multiset precision, recall, F1 | Count-aware overlap between observed and generated continuation events after mapping. | Up |
| Next-visit rank-recall AUC | Vocabulary-rank recovery of the event set in the next observed visit. | Up |
| Generated next-visit precision, recall, F1 | Set recovery in a sampled next visit rather than under teacher forcing. | Up |
| RATE digit accuracy | Exact accuracy for each mixed-radix digit at a valid inter-visit gap. | Up |
| Event ECE | Bin-weighted gap between maximum event confidence and exact next-event accuracy. | Down |
| Reliability curve and Brier score | Calibration and squared probability error produced by the evaluator. | Toward diagonal; down |
| Mean token entropy | Average spread of the event-renormalized next-event distribution. | Qualitative |
| Log-prevalence Pearson correlation | Multiplicative agreement of observed and generated code frequencies across the long tail. | Up |
| Raw-prevalence Spearman correlation | Agreement in the untransformed rank ordering of code frequencies. | Up |
| Observed-code coverage | Fraction of observed event types with at least one occurrence in generation. | Up |
| Gap-prevalence agreement | Pearson on log gap-bin prevalence, Spearman on raw prevalence, and coverage. | Correlations and coverage up |
| Rare-event weighted RMSLE | Log-frequency error with greater attention to low-prevalence codes. | Down |
| TSTR and TRTR | Same classifier trained on synthetic or real data and tested on the same held-out real set. | Up; ratio toward 1 |
| Effective rank | Entropy-based dimensionality of trajectory-representation matrices. | Up if collapse is the concern |
| AUROC | Pairwise ranking of observable case and control rows. | Up |
| AUPRC | Precision across recall thresholds at the observed evaluation prevalence. | Up |
| Severity-Charlson Spearman | Monotone association between learned severity and comorbidity burden. | Up |
| Body-system kNN purity | Local clinical-label agreement relative to a frequency-matched random baseline. | Up |
| Patient retrieval overlap@K | Shared clinical concepts among nearest patient representations. | Up |
| Cancer-onset surprise AUROC | Discrimination of first cancer-coded visits from model surprise. | Up |
| ICD-chapter onset AUROC | First-onset discrimination using chapter-level event mass. | Up |
| Medication MoA AUROC | Next-event recovery after grouping medicines by mechanism of action. | Up |
| PPV at an operating threshold | Observed positive-window among the highest-ranked trajectories. | Up |
| Suggested number to screen (SNS) | Reciprocal of trajectory-level PPV; it quantifies the number of patients screened to find one positive case as per cancer-registry ground truth (not mortality-based NNS). | Down |
| Population-translated pos. pred. value (PPV) | PPV after replacing sample prevalence with a stated external incidence prior. | Up |
| Standardized incidence ratio (SIR) | Observed incidence relative to expected population incidence. | Up |
| Lead time | Cutoff-to-diagnosis interval among correctly predicted trajectories at operating threshold. | Qualitative; longer may be more actionable |
| Age and length matching | Sensitivity of discrimination to demographic and record-length imbalance. | Stability preferred |
| Stratified macro AUROC | AUROC breakdown across cancer, sex, and age cells. | Up |
| Directional IG importance | Population share of positive or negative attribution mass assigned to each code. | Up within direction |
| Top-decile attribution rate | Fraction of rows in which a code enters the selected directional attribution set. | Up |
| PMI enrichment and EB shrinkage | Attribution enrichment relative to prevalence with stabilization of sparse codes. | Up |
| Occlusion impact | Score change after masking one selected event while preserving the time representation. | Magnitude and sign |
| cPMI | Model-inferred directed association of a target event given the contexts ending in a source event. | Signed qualitative |
| Modality-adjusted cPMI | Association after removing changes in total target-modality mass. | Signed qualitative |
| Disparity-filter score | Share of a source event's positive outgoing association mass carried by one edge. | Low $\alpha$ retained |
| UMAP and trajectory plots | Qualitative visualization of event, visit, patient, and cancer geometry. | Qualitative |

### Scores used to evaluate generative fidelity

We initially assess the performance of the pretrained generative model to forecast future clinical events, to generate realistic synthetic trajectories, and to model time intervals between hospital visits.

#### Prompt completion

Here, we analyze whether high probability or rank is attributed to the true next token, next event category, and next clinical visit. Using our validation dataset, we divide patient trajectories into two segments: an observed initial prefix and a held-out continuation sequence. Prefix is given as an input to the model and the subsequent events are generated using an autoregressive process. We compute semantic multiset overlap between real and generated events across patients and evaluate the precision, recall, and F1 score. Furthermore, we evaluate population level alignment using Pearson and Spearman correlation coefficients between the actual and model predicted code prevalences across patient cohorts. The Pearson correlation is evaluated on a log scale to focus on consistency across the long-tailed frequency distribution which avoids score inflation due to highly frequent codes. Conversely, the Spearman correlation uses raw prevalence to assess whether the model accurately reproduces the rank order of frequent and infrequent codes.

### Conditional trajectory generation

Next event evaluation does not test whether the model can generate a long context of events. We sampled long trajectories to assess whether the model captures the event prevalence, code coverage, and time-gap distribution of real observed data. We uniformly at random sample static tokens (demographic information at the beginning of the trajectory) and used them as the initial prompt. Clinical events are sampled autoregressively from the event head while RATE digits are sampled from the corresponding radix-head distributions. Because event and time-gap tokens have separate heads, their sampling temperatures can be configured independently. We cap generation at a predefined maximum of valid clinical event tokens per trajectory and for both observed as well as predicted trajectories we remove their shared initial prompts prior to scoring to prevent prefix leakage. This step ensures comparisons are made on actual event codes. We report Pearson correlation between log-transformed observed *f*_*v*_ and generated 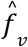 code prevalence to assess agreement across the long-tailed frequency distribution. We also report Spearman correlation to compare frequency rankings, and whether observed code reflects coverage of events in real data Separately, we evaluate temporal fidelity by comparing the histograms of observed and generated gap-days distributions, radix prevalence, and gap distributions across ICD chapters.

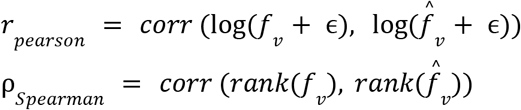

#### Synthetic-data utility

We measure the utility of synthetic data using TSTR (train synthetic and test real) and TRTR (train real and test real). Here, we train a simple classifier to predict whether a patient has cancer or not. We build a bag of event feature vectors (excluding cancer code) for cases and controls and train a logistic regression classifier on real trajectories as well as synthetic trajectories separately. We evaluate both classifier results on held out real data and report the prediction performance.

### Scores to evaluate generative fidelity

#### Perplexity (PPL)

Perplexity is a measure of how well the probability distribution of future events predicted by the model aligns with the actual patient trajectory. A lower perplexity means the model is less “surprised” by the patient’s future event. We compute PPL across event only tokens (to assess that the model is not simply learning to predict syntactic structure).

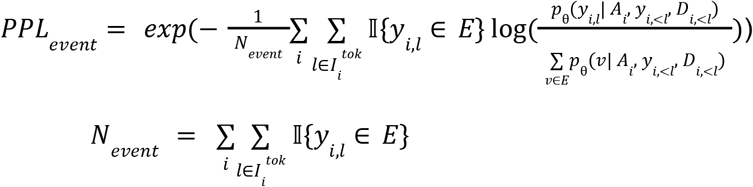

#### Event ranking

*Top-k accurac*y measures whether the correct future clinical event appears in the model’s top *k* most probable predictions. Let the rank of the observed event be 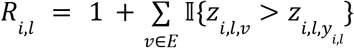 that counts event tokens with a strictly larger logit than the observed event (equal logits therefore get the best tied rank), where *z*_*i,l,v*_ is the logit for event *v*. Top-K event accuracy is

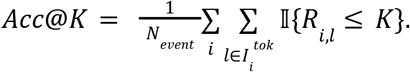

#### Mean reciprocal rank (MRR)

evaluates the position of the correct future event in the full ranked list of predictions,

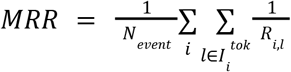

### Calibration, uncertainty, and representation quality

#### Calibration

A requirement for clinical utility is the reliability of the model’s probability estimates. If a model assigns a 0. 95 probability to an event then among comparable predictions with confidence near 0. 95 the event should occur approximately 95% of the time. We report expected calibration error (ECE) which summarizes the discrepancy between predicted event probability and empirical event frequencies. Given *N*_*event*_ evaluation predictions, we partition the interval [0, 1] into Ĝ confidence bins. Let ß ι denote the set of samples in bin ι, *acc*(ß_ι_) be the event frequency in that bin, and *conf*(ß_ι_) the mean predicted confidence. Then,

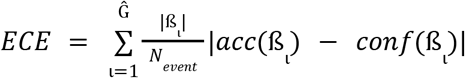

Lower ECE indicates better calibration.

#### Uncertainty quantification

We use mean token entropy (MTE) to summarize the spread of the next-token distribution across valid positions. MTE is defined as a Shannon entropy *H* of the predicted distribution 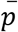 over token values at each time step.

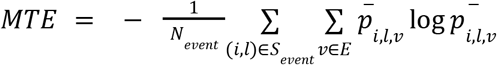

We compute MTE under teacher-forcing on held-out patient trajectories.It ranges from 0 to log |*E*|, high score means that probability is distributed across several event tokens and low means probability mass is concentrated on fewer alternatives. MTE can be high in cases where several future events may be clinically plausible.

#### Effective rank

We assess the diversity and non-degeneracy of the learned patient representation by computing the effective rank of the patient embedding matrix. For each patient in the test cohort, a fixed-dimensional representation is extracted from the hidden state at the final token (the < *eos* > embedding). By stacking these individual hidden states, we construct the patient representation matrix *E ∈* ℝ ^*N*×*d*^, let σ_*g*_ represent its singular values, with *d* *= min (*N, d*), the effective rank is defined as,

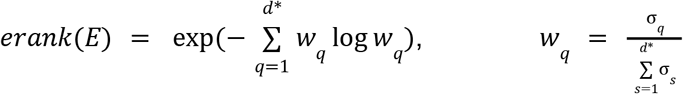

A lower value means representation collapse indicating representation fails to capture patient diversity. A high effective rank implies a rich structure in the representation space.

### Downstream task performance

#### Next-event category

At each scored event position (*r, l*) probabilities are summed over hierarchy defined groups such as ICD chapter, ATC class, medication mechanism, CPT/HCPCS section, and LOINC class. Category masses are renormalized within modality and evaluated using one-vs-rest AUROC and macro AUROC (separately in each group, then averaged overall, disregarding of group size) scores.

#### Retrieval of events in the next visit

At each true visit boundary *j*, we can rank everyevent*v ∈ E*using the history through visit *j*. We then assess the recall of clinical events at various rank thresholds and report rank-recall AUROC which summarizes whether true events in the next visit are ranked high.

#### ICD-chapter onset

For predicting the onset of ICD chapter, patient trajectories are censored at the first diagnosis in that chapter. We use gap-boundary scoring based on cancer logit matching the setup of GenEHR multi-cancer risk assessment, and report per-chapter AUROC and macro AUROC across chapters.

### Patient embedding analysis

#### Event-space geometry

We project event-token representations with UMAP and color the points by modality and configured clinical ontology labels, including ICD chapter, procedure section, laboratory class, ATC class, and medication mechanism. These plots are qualitative and are used to inspect whether clinically related events occupy nearby regions. We do not infer discrete clusters or clinical validity from UMAP alone.

#### Cancer-events geometry

To map the relationships between cancer type event codes in the representation space, we construct a k-nearest neighbor graph with k=4. Within this graph, edges are annotated as strong associations if the connected cancer codes are mutual nearest neighbors of each other.

#### Patient-embedding geometry

To evaluate patient embedding geometry, we analyze whether the embedding space successfully places patients with clinically similar trajectories in close proximity. For each patient, we average the final-layer contextual hidden states at clinical event positions and project the resulting representation using UMAP with cosine distance. We overlay CCI, dominant ICD chapter, dominant body system, record recency, number of distinct medications, and recent acuity. The dominant ICD chapter is the chapter containing the largest number of diagnosis events in the patient’s history. The dominant body system is the most frequent mapped system across clinical events, excluding the unmapped ‘Other’ category when a mapped system is present. These overlays assess whether clinically related gradients are retained in the representation without treating visual separation as a quantitative performance measure.

#### Visit space and clinical-state analysis

A patient can contribute several points. The visit index is the zero-based chronological order of retained visits after sequence truncation, not elapsed time. We construct a shared two-dimensional diffusion-UMAP layout and trace successive visits in chronological order. For the population flow field, we divide the layout into a fixed 24 by 24 grid and displacement vectors are averaged for transitions starting in each cell. Cells with fewer than six transitions are omitted.

### Cancer-risk prediction

#### Per-cancer, stratified macro, and age sensitivity discrimination performance

For each cancer and prediction horizon we report AUROC to measure pairwise ranking of observable cases and controls in the retrospective set, and AUPRC for precision across recall thresholds under the observed class prevalence. We then perform age-matched and stratified sensitivity analyses to quantify how much discrimination is associated with demographic differences. Controls are matched to cases within five-year age bins. Next, AUROC is computed within cancer-, sex-, and age-specific groups and reported across cancer types using a macro average. This analysis assesses the sensitivity of model performance to age and sex but it does not remove age or sex from the prediction model.

### High-risk cohort enrichment

To assess clinical utility with focus on effective intervention programs, especially for cancer types without current population-wide screening recommendations, we emphasize performance within the highest-risk cohort, defined as the top-*N* patients ranked by their predicted risk. **Positive predictive value (PPV@N)** for a high risk top-N cohort is defined as,

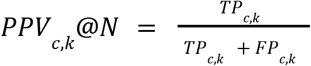

where *TP*_*c,k*_, *FP*_*c,k*_ are true positives and false positives for cancer *c* in interval *k* at threshold *N*(e.g., N=1000 at top risk).

**Suggested number needed to screen (SNS@N)** for a high risk cohort is a good estimate for the number of patients needed to screen to detect one cancer case, defined as

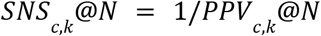

**Standardized incidence ratio (SIR)** is defined as the ratio of the number of observed cases to the number of expected cases within the top-N high risk cohort.

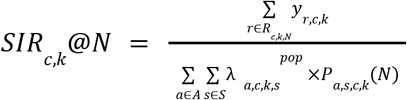

where λ_*a,s,k,s*_^*pop*^ is the real-world population-level incidence for cancer *c*, horizon *k*, age *a*, sex group *s*, and *P*_*a,s,c,k*_(*N*) is the number of selected patients for the same cancer, demographic,and prediction horizon.

### Lead-time and exclusion-window sensitivity

Lead time of cancer is an endpoint-to-diagnosis interval Δ*T*_*r*_= *T*_*i*(*r*),*c*_− τ_*r*_ between the timestamp of the last event of a patient’s trajectory used for risk scoring and the true cancer diagnosis. We compute the time to cancer Δ*T*_*r*_ for every trajectory *r* in the true-positive cohort (flagged as high-risk who got cancer). In other words, for those (in the test set) predicted correctly to have a cancer diagnosis within 36 months, when did the cancer actually occur? We aggregate these values 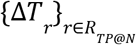to report a population level distribution of each cancer type, indicating the range of time intervals in which intervention is advised, in response to the clinical question “how far in advance can future cancer diagnosis be reliably predicted?”.

### Operating threshold sensitivity analysis (Metrics as a function of N)

The selection of the operating threshold *N* involves a tradeoff between resource constraints and sensitivity of detection. Testing larger subpopulations in order to find a larger fraction of actual cancer cases becomes increasingly expensive. Evaluation of the prediction methods on withheld data shows that the efficiency of cancer detection is highest for those at highest predicted risk. To characterize this operating landscape, we perform a sensitivity analysis by varying the high-risk cohort size *N* from 100 to a million trajectories. We visualize the performance by plotting the SIR, and PPV as a function of the operating threshold *N*.

### Interpretation of features contributing to cancer log-odds

We identify risk factors driving cancer log-odds using feature interpretability. Specifically, we use IG to calculate event-level scores for individual patients, which are further aggregated across the cohort to derive a global importance score. In addition, we use an occlusion trajectory analysis, which highlights events that appear to cause substantial health state transitions (details below). And, to analyze how the model learns associations among events, we compute the conditional mutual information of events in GenEHR. This helps us to evaluate whether these relationships capture known clinical knowledge or are fertile ground for novel hypotheses.

### Top cancer risk features using integrated gradients

We compute token-level attributions using the IG attribution to measure contribution of individual events in each patient trajectory. IG assigns a signed attribution to each event token along a path from a prespecified baseline (e.g, padding token) to the observed trajectory representation. Here, we are interested in tokens of a sequence that explain a trajectory’s cancer-risk score. A positive IG means tokens/features that increase the cancer log-odds whereas negative IG means features that decrease the cancer log-odds.

For trajectory *r*, let *X*_*r*_ be the embedding of observed trajectory, 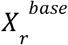 be the embedding of baseline, and *S* be the risk score, the IG attribution for the *l*-th clinical event is defined as path integral of the gradients from 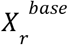 to *X*_*r*_,

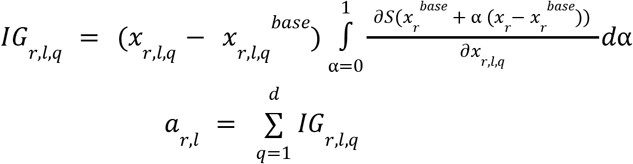

*a*_*r,l*_ is a cumulative score summed across all *d* dimensions of embedding matrix.

Aggregating IG across patients to obtain global feature importance is non-trivial because IG is sample-specific. For each trajectory, positive and negative attribution scores are separated and scaled by trajectory’s total absolute contribution. We then aggregate the contributions by clinical code which prevents a few long trajectories from dominating the global ranking.

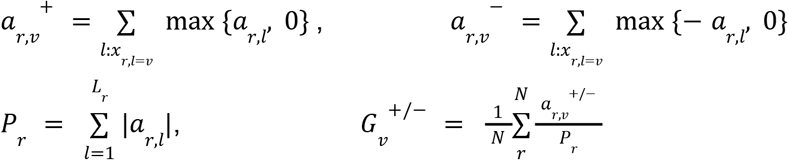

For code *v*, global score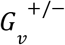normalizes for code prevalence (a frequent code must consistently appear among the highest-attribution tokens across a set of patients to achieve a high global importance score).

### Occlusion analysis on most important features as per IG

We combine IG with occlusion-based counterfactuals to find critical features. Given a patient trajectory *X*_*r*_ = [*x*_1_, Δ*t*_1_, *x*_2_, Δ*t*_2_, ···], we define the patient-specific risk Δ*Risk*_*r,l*_ as the marginal effect of event *x* _*l*_ on the predicted risk. This is derived from ablation by masking one by one top-k tokens as identified by IG.

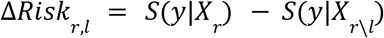

A positive Δ*Risk*_*r,l*_ implies *x*_*l*_ contributes to increased risk, while a negative value implies itlowers the risk. To further aggregate this feature importance across a cohort we compute the metrics:

#### Mean absolute impact (MAI)

This score measures the magnitude of a code’s general influence on the model’s decision boundary ignoring directionality,

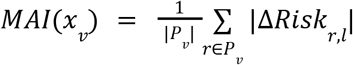

#### Risk directionality (RD)

This score whether on average a code points towards or away from cancer outcome,

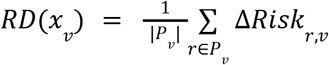

*RD*(*x*_*v*_) > 0 increases log-odds of cancer, *RD*(*x*_*v*_) < 0 decreases log-odds of cancer,*RD*(*x*_*v*_) ≈ 0 means the code is context-dependent (increasing log-odds in some patients whiledecreasing it in others).

### Event belief-graph inference using conditional pointwise mutual information

We constructed a GenEHR decoder-association graph over clinical event tokens. Let *C*(*e*_*context*_) denote the set of every held out gap position whose preceding visit contained *e*_*context*_, and let *N*(*e*_*context*_) be the total weight of those positions weighted by the inverse of its position count across all patients and visits. For each context *h*_*i,l*_for the members of *C*(*e*_*context*_) we used modality normalized probability of a next event *e*_*next*_. These probabilities were averaged over all eligible occurrences of *e*_*context*_ and compared with the model averaged over all weighted positions,

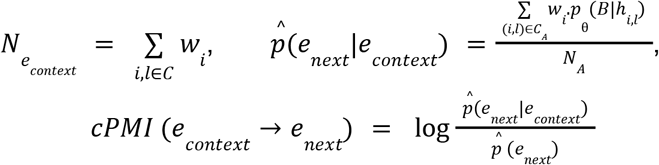

In cPMI matrix a positive value means that a context ending in *e*_*context*_ assigns higher probability to *e*_*next*_ than its model-implied baseline, zero means no change relative to baseline, and a negative value implies suppression. For strength of cross-modality associations, we applied an adjustment that renormalizes the probability mass across the target modality prior to comparing all pairwise source target associations.

We transformed the cPMI matrix to a directed graph by filtering out weak associations. Specifically, for every source with *N*_*econtext*_≥ 50, its top 15 scoring target.

### Events contributing to transition of cancer log-odds over time

A patient may move from a low-risk score at an earlier cutoff to a higher-risk score as new events are encountered over time. For event *v*, transition polarity compares the number of score increasing and decreasing instances associated with that event. To define thresholds for transition we define discrete risk strata (e.g., low, medium, high) based on population-calibrated thresholds γ _*low*_ and γ _*high*_,

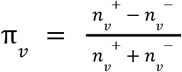

where 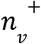, 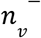are the counts of *v* resulting in risk increase and risk decrease. A code with high π_*v*_ > 0are events that frequently lead to increase of risk.

### Baseline models for pan-cancer risk prediction

We compared fine-tuning with several supervised baselines trained using the same data split as in fine-tuning. Demographic baseline is a logistic regression trained using three features (standardized age, sex, and an indicator if age was observed). We also constructed a5,479-dimensional count vector of clinical events from the training data. For a feature *j*, let *n*_*r,j*_be a count of event *j* in trajectory *r*, the transformed feature was *x*_*r,j*_ = log(1 + *n*_*r,j*_). We then_*r,j*_ used L2-regularized logistic regression, random forest and XGBoost^90^ models using these features. We separately extracted the GenEHR representation of trajectory (<eos>) and trained random forest^91^ and XGBoost on this representation.

A separate supervised model was fitted for each of seventeen cancers at horizons of 6, 12, 24, 36, and 60 months, using a three-month delayed entry period. The random forest used 64 trees, a maximum depth of 10, and a minimum leaf size of 20. XGBoost used a maximum depth of 4, a learning rate of 0.05, and up to 200 boosting rounds with early stopping based on validation loss. Cancer-specific AUCROC,and SIR and PPV at 1000 operating points are included in Fig S16.

### Model Implementation and hyperparameters

We used PyTorch^92^ and the Hugging Face transformers library^93^ for implementation optimized with AdamW^94^. Cohort specific learning rate and architecture details are in Tab S3.

#### Network architecture and training scheme

MGB, PROV, AoU, and UKBB used cohort-specific variants of a Llama decoder architecture (Tab S3), US-VA used GPT-Neo. The Llama-based models were instantiated as LlamaForCausalLM from the HuggingFace Transformers library (version>=4.45,<6.0). The decoder had two output components. An autoregressive head predicted clinical/structural tokens autoregressively, while a RATE head predicted the inter-visit interval. The RATE decoder consists of five parallel linear subheads with categorical output sizes [7, 4, 6, 5, 5], corresponding to place values [1, 7, 28, 168, 840] days. Conditioned on the shared hidden representation, each subhead predicted one time-gap digit independently. This representation covers integer intervals from 0 through 4,199 days using 27 output logits rather than a single 4,200-class output layer.

Compute-optimal scaling was adopted for defining different model-size tiers, dependent on the training dataset tokens (model size ∼= total_tokens / 20)^95^. Different tiers use increasing factors of roughly 1x / 2x / 4x of this budget. All models were trained to a Chinchilla optimum so that the model sees a fixed budget of target tokens during training dependent on the number of parameters. Target tokens were set to 24 times the number of (active) parameters. The maximum number of training steps was calculated as the total target tokens divided by the product of effective batch size and average sequence length. For convergence, we also monitored the val loss using a patience of 5, i.e. stop the training if no improvement in val loss for 5 consecutive epochs.

#### Fine-tuning

In the fine-tuning phase only the gated shared basis LoRA adapters and cancer risk heads were updated. We used rank-4 adapters into all linear projections in each of the 14 transformer layers (query, key, value, and output projections, gate, MLP projections) resulting in 98 adapted modules. For rank=4, we set α_*LORA*_ to 8. Each module has its shared basis A, B across all 17 cancers and 17 × 4 parameters of cancer-specific gates. We normalized the basis vector and a shrinkage penalty λ_*gate*_ was set to 0.01. LoRA parameters were optimized jointly with cancer heads, with 830, 360 overall trainable parameters.

