## Supplementary Materials for "Identifying cohorts at elevated risk of cancers using generative modeling of patient health states"

| **Hyper-parameter/ Cohort** | **MGH** | **PROV** | **All of Us** | **VA** | **UKB** |
|---|---|---|---|---|---|
| Model architecture | llama_large | llama_2b_small | llama_medium | gpt-neo | llama_large |
| Architecture details | hidden_size: 768<br>num_attention_heads: 12<br>num_hidden_layers: 14<br>intermediate_size: 2048<br>max_position_embeddings: 512<br>rms_norm_eps: 1.0e-06 | hidden_size: 768<br>num_attention_heads: 12<br>num_hidden_layers: 12<br>intermediate_size: 2048<br>max_position_embeddings: 512<br>rms_norm_eps: 1.0e-06 | hidden_size: 384<br>num_attention_heads: 12<br>num_hidden_layers: 8<br>intermediate_size: 1280<br>max_position_embeddings: 1024<br>rms_norm_eps: 1.0e-06 | hidden_size: 768<br>num_heads: 12<br>num_layers: 20<br>intermediate_size: 3072<br>max_position_embeddings: 512<br>Attention_types:<br>global:10<br>local:10 | hidden_size: 128<br>num_attention_heads: 4<br>num_hidden_layers: 4<br>intermediate_size: 384<br>max_position_embeddings: 256<br>rms_norm_eps: 1.0e-6 |
| Total parameters | ~108.3M | ~95.7M | ~19.44M | ~145M | ~1.18M |
| Vocab size | 6147 | 6944 | 4694 | 2885 | 1281 |
| Batch size | 32 | 32 | 126 | 64 | 16 |
| Data-types | [icd, proc, lab, med] | [icd, proc, lab, med] | [icd, proc, lab, med] | [icd] | [icd] |
| Lifestyle information | No | Yes (Alcohol, Smoking, BMI) | Yes (Alcohol, Smoking, BMI) | No | Yes (Alcohol, Smoking, BMI) |
| Demographics | Yes (Gender, Race, Ethnicity) | Yes (Gender, Race, Ethnicity) | Yes (Gender, Race and Ethnicity) | Yes (Gender, Race and Ethnicity) | Yes (Gender and Ethnicity) |
| Exposure information | No | Yes (Area Deprivation Index) | No | No | No |

| | | | | | |
|---|---|---|---|---|---|
| Age bucket edges | [30, 35, 40, 45, 50, 55, 60, 65, 70, 75, 80, 85] | [30, 35, 40, 45, 50, 55, 60, 65, 70, 75, 80, 85] | [30, 35, 40, 45, 50, 55, 60, 65, 70, 75, 80, 85] | [30, 35, 40, 45, 50, 55, 60, 65, 70, 75, 80, 85] | [30, 35, 40, 45, 50, 55, 60, 65, 70, 75, 80, 85] |
| Stratified buckets ratios | [short: 0.17, medium: 0.20, long: 0.63] | [short: 0.17, medium: 0.16, long: 0.67] | [short: 0.23, medium: 0.16, long: 0.61] | [short: 0.20, medium: 0.26, long: 0.54] | [short: 0.11, medium: 0.07, long: 0.82] |
| Min events | 3 | 3 | 2 | 5 | 2 |
| Max length | 512 | 512 | 1024 | 512 | 256 |
| Learning rate (LR) | 5e-5 | 5e-5 | 5e-5 | 1e-4 | 5e-5 |
| Weight decay | 0.01 | 0.01 | 0.01 | 0.1 | 0.01 |
| Warmup steps | 10% | 10% | 10% | 10% | 10% |
| Cosine annealing total steps | 137500 | 246500 | 21000 | 10000 | 25000 |
| Max steps | 137500 | 246500 | 21000 | -1 (3 epochs (steps)) | -1 (trained to val loss saturation) |
| GPU Used | A100, 40GB | H100, 94GB | A100, 40GB | H100, 100GB | A100, 40GB |

***Table S1. Cohort and model specific hyper-parameters.*** *List of the key hyper-parameters used for GenEHR model training for the specific cohort and the specific models.*

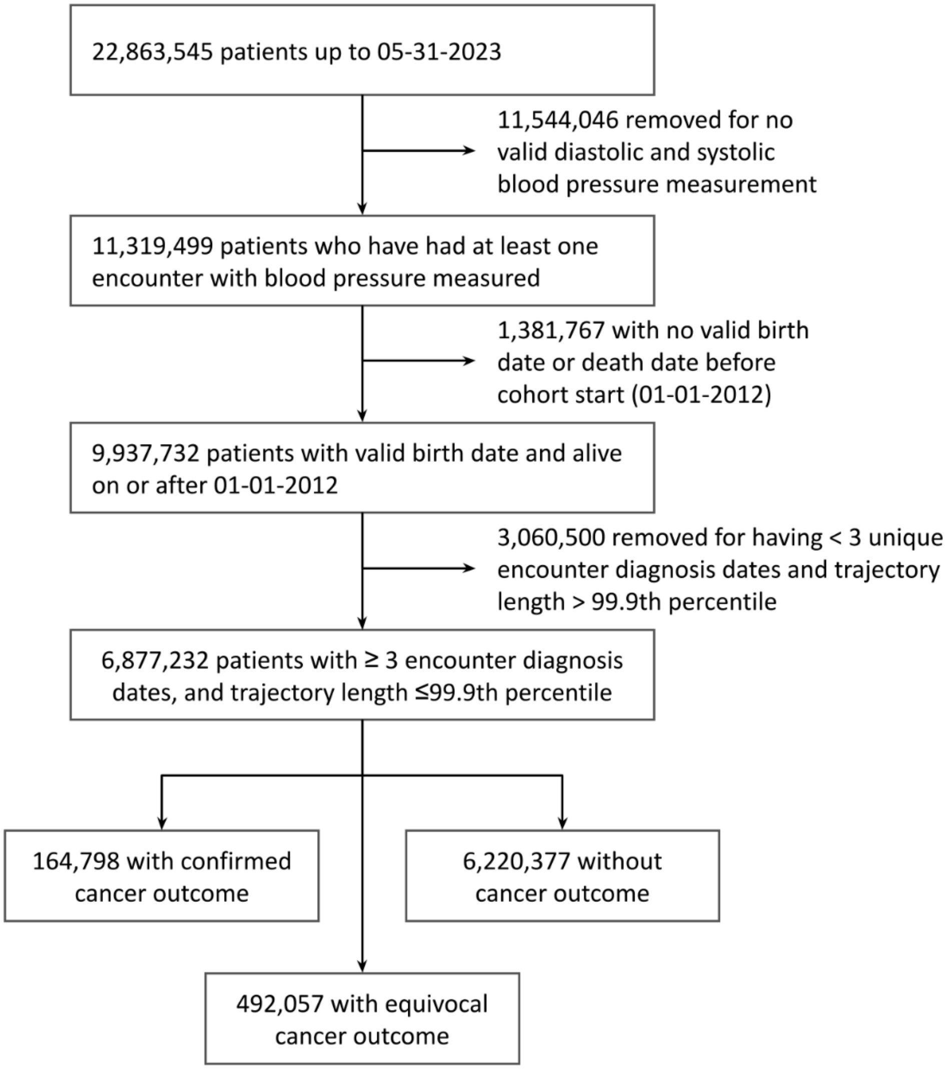


***Figure S1a: Data preparation and quality control filtering of the PROV cohort.*** *Patient level inclusion and exclusion steps which results in a cohort of 6,877,232 patients of whom 164,798 had a cancer outcome confirmed through the linked cancer registry. Filtering against cancer registry gives clean labels useful for supervised fine-tuning and reliable estimate of model performance.*

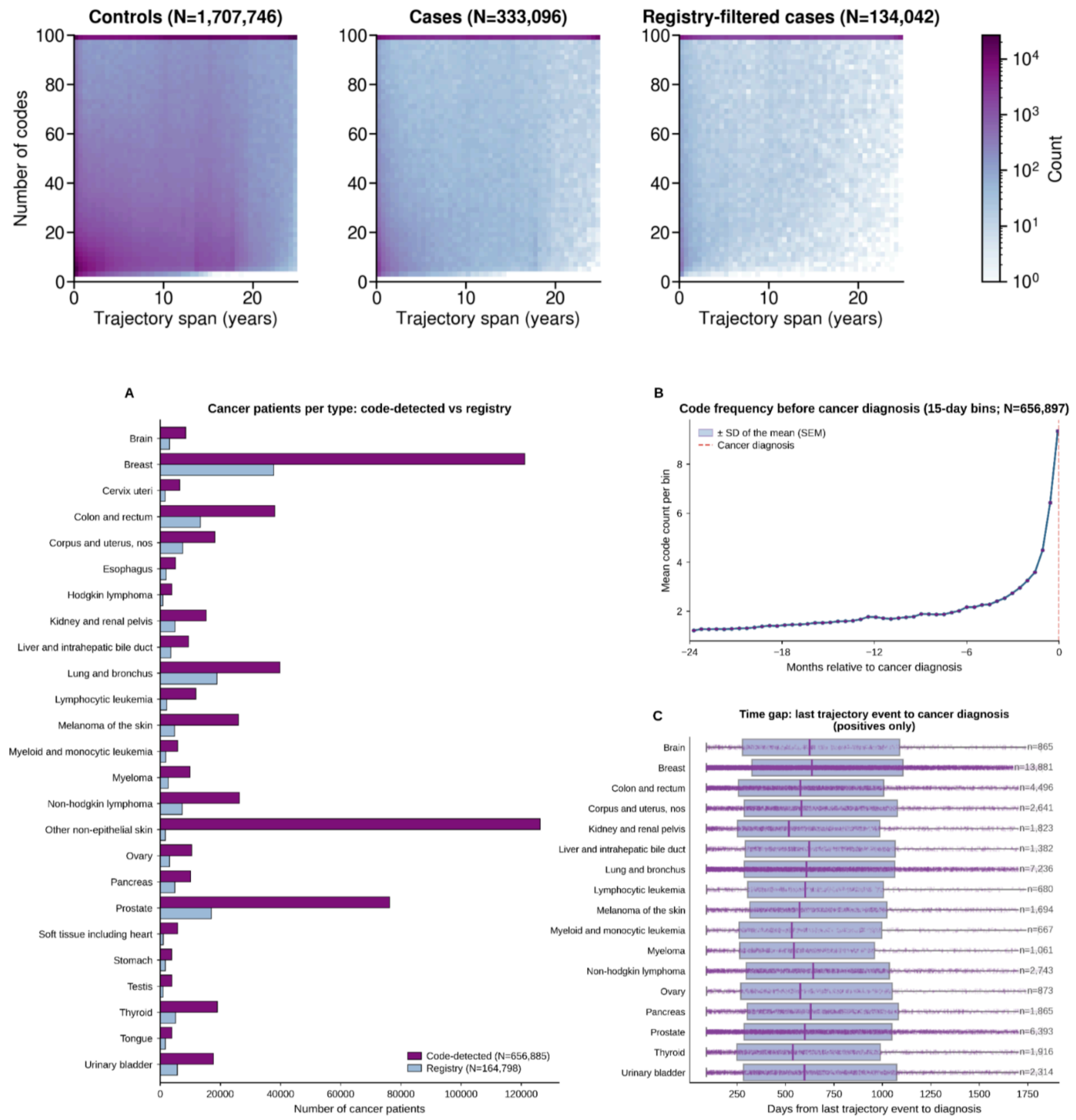


***Figure S1b: Data statistics summary of PROV cohort. (Top)*** *Event count and trajectory time span distribution shown for three groups: controls (no-cancer event), cases (cancer event in EHR), and overlap (cancer cases in EHR also present in registry). Cases and controls have a good coverage of event counts and trajectory durations, registry filtering resulted in a smaller cohort which highlights the challenge of relying on ICD codes for diagnostic accuracy.* ***(Bottom)*** *Cancer registry filtering and trajectory sampling for cancer risk scoring. (A) Cancer counts by site before and after matching EHR diagnosis with linked cancer registry outcomes. This comparison shows the unreliability of cancer outcomes from EHR codes alone. We excluded patients with cancer recorded either only in EHR or only in registry. (B) Cancer diagnosis is often preceded by diagnostic work-up codes that become more frequent closer to the diagnosis. (C) To answer how far ahead in time our model can predict risk, we sampled multiple*

*trajectories per patient. The preceding window was divided into four equal size segments with one endpoint sampled from each segment which prevents concentrating trajectories close to diagnosis. Plotted endpoint distribution across cancer types*.

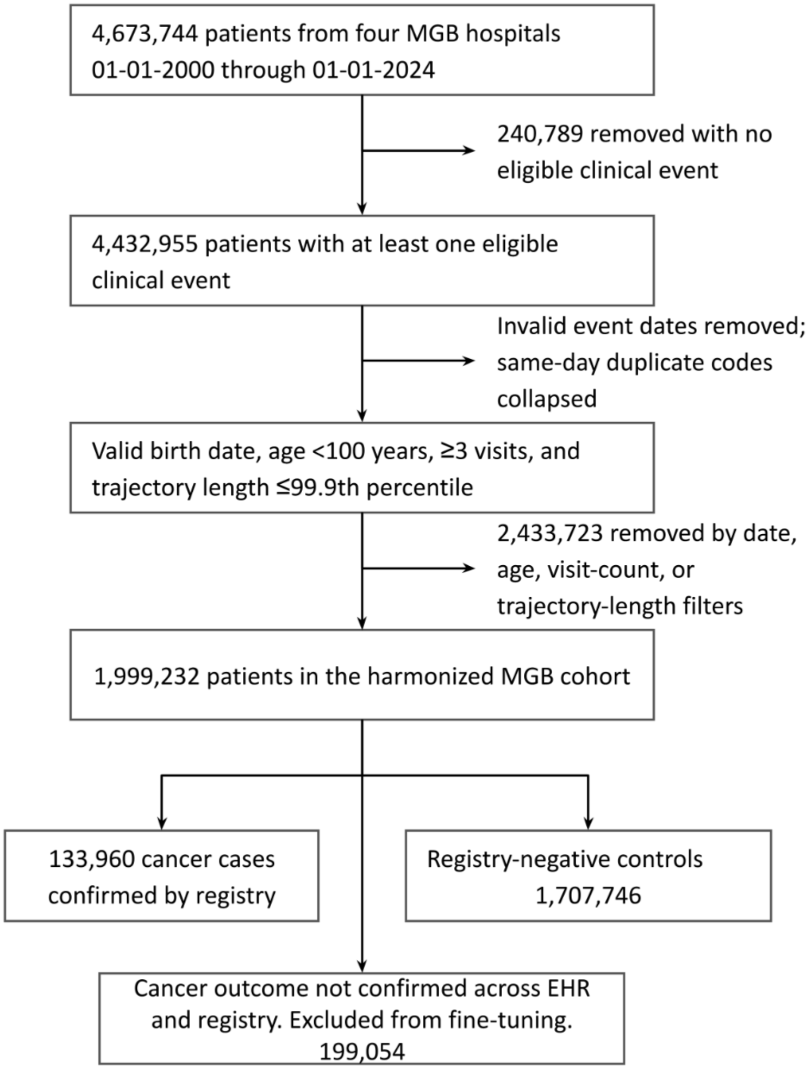


***Figure S2a: Data preparation and quality control filtering of the MGB cohort.*** *Patient level inclusion and exclusion steps which results in a cohort of 1,999,232 patients of whom 133,960 had a cancer outcome confirmed through the linked cancer registry. Filtering against cancer registry gives clean labels useful for supervised fine-tuning and reliable estimate of model performance.*

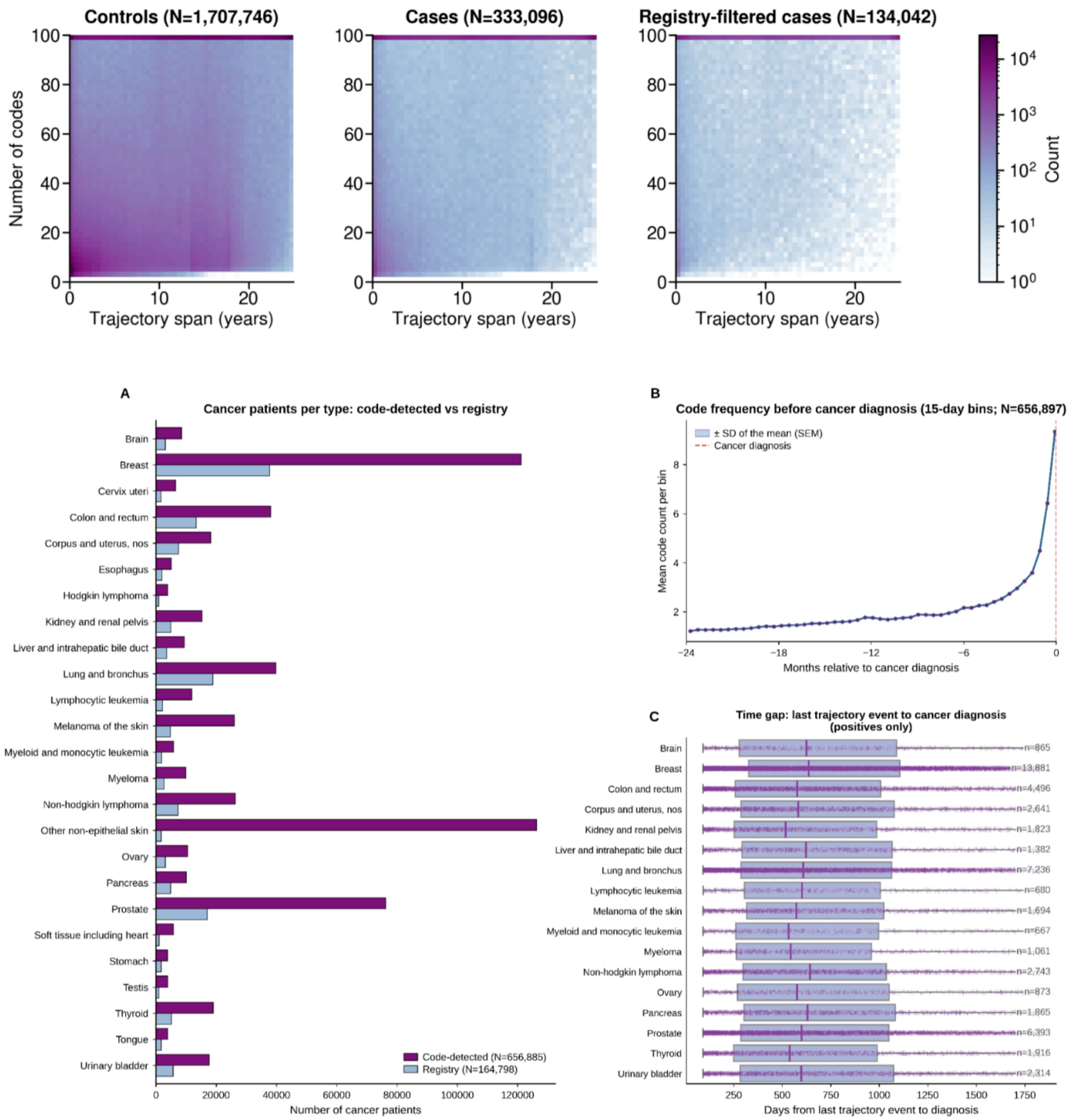


***Figure S2b: Data statistics summary of MGB cohort. (Top)** Event count and trajectory time span distribution shown for three groups: controls (no-cancer event), cases (cancer event in EHR), and overlap (cancer cases in EHR also present in registry). Cases and controls have a rich coverage in terms of event counts and trajectory durations, registry filtering resulted in a smaller cohort which highlights the challenge of relying on ICD codes for diagnostic accuracy. **(Bottom)** Cancer registry filtering and trajectory sampling for cancer risk scoring. (A) Cancer counts by site before and after matching EHR diagnosis with linked cancer registry outcomes. This comparison shows the unreliability of cancer outcomes from EHR codes alone. We excluded patients with cancer recorded either only in EHR or only in registry. (B) Cancer diagnosis is often preceded by diagnostic work-up codes that become more frequent closer to the diagnosis. (C) To answer how far ahead in time our model can predict risk, we sampled*

*multiple trajectories per patient. The preceding window was divided into four equal size segments with one endpoint sampled from each segment which prevents concentrating trajectories close to diagnosis. Plotted endpoint distribution across cancer types.*

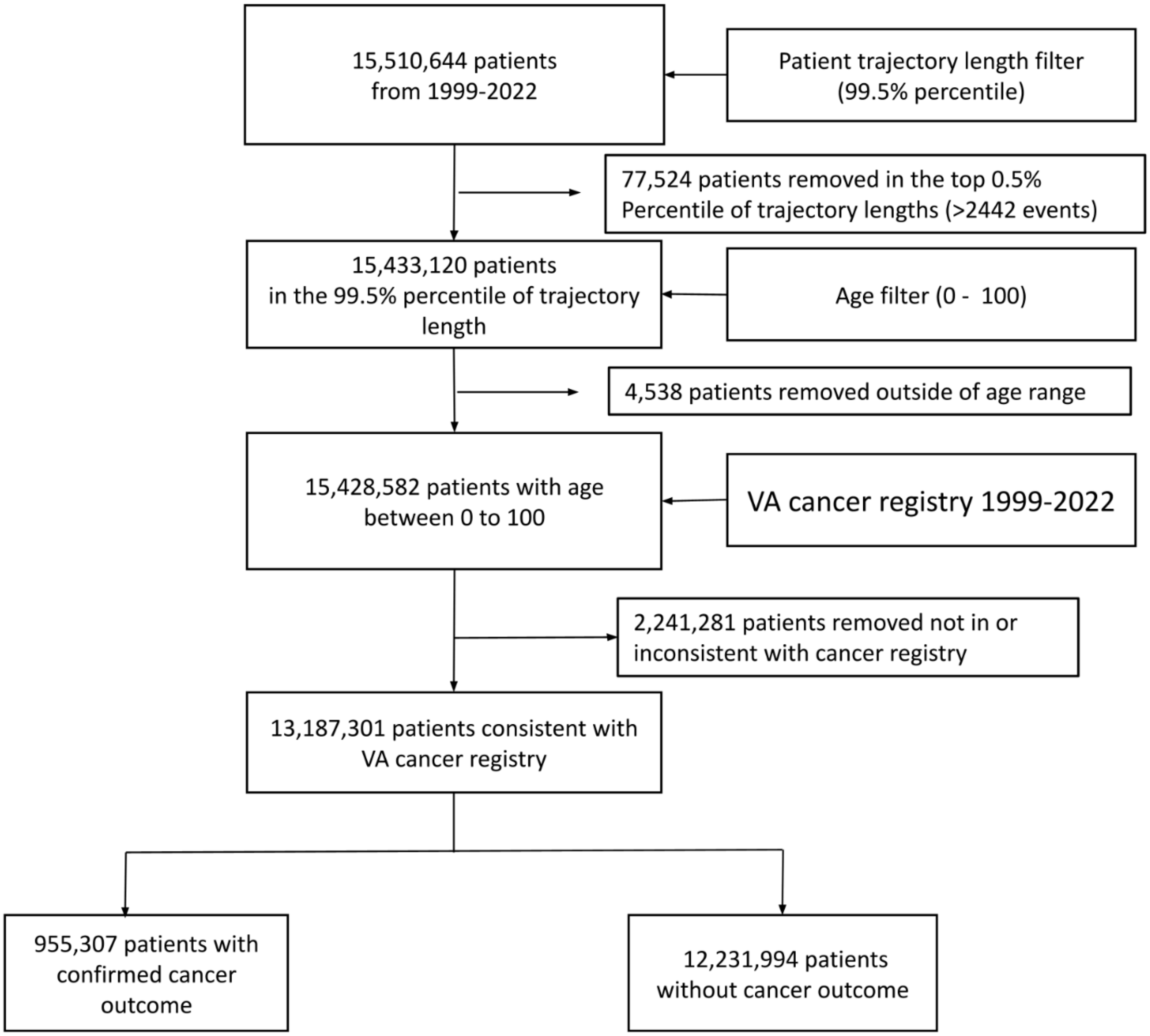


***Figure S3a: Data preparation and quality control filtering of the VA cohort.** Patient level inclusion and exclusion steps which results in a cohort of 13,187,301 patients of whom 955,307 had a cancer outcome confirmed through the linked cancer registry. Filtering against cancer registry gives clean labels useful for supervised fine-tuning and reliable estimate of model performance.*

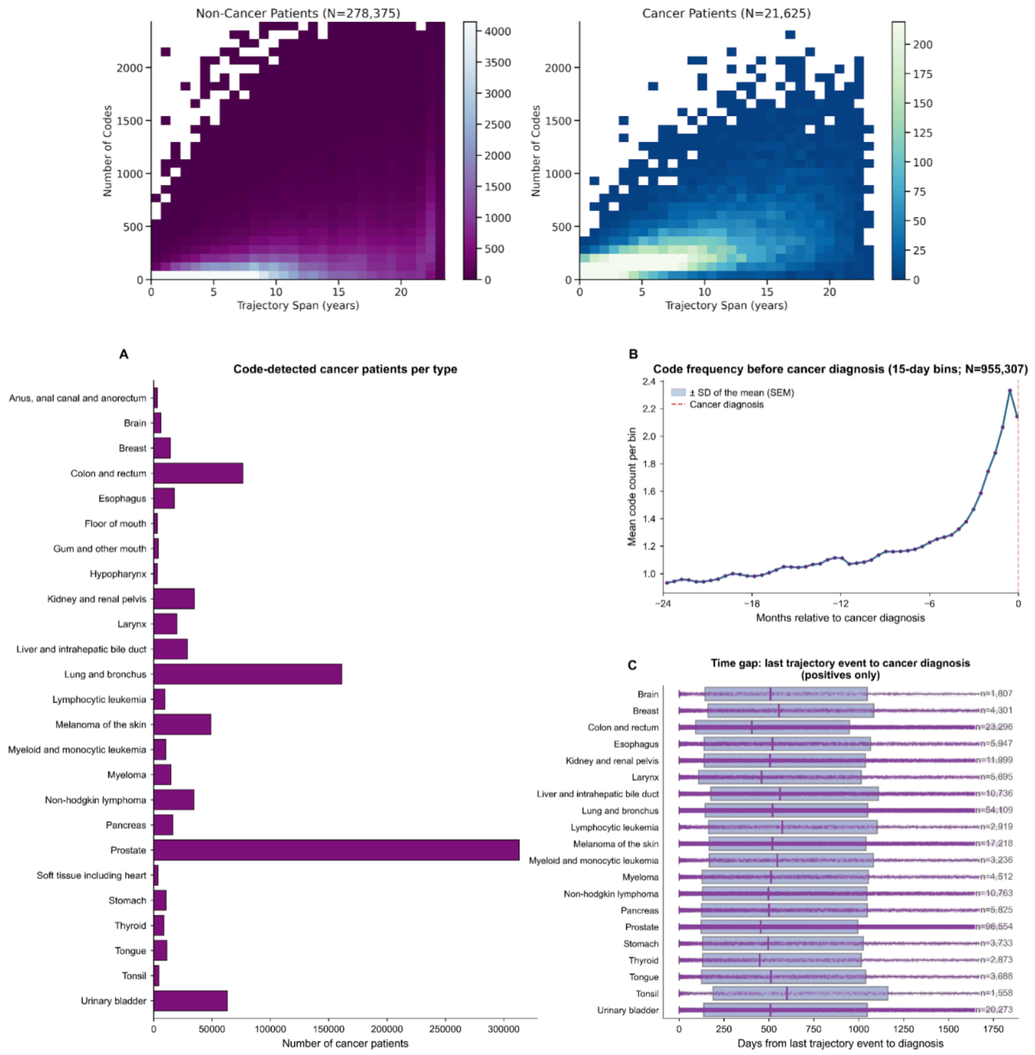


***Figure S3b: Data statistics summary of VA cohort. (Top)*** *Cases and controls have a rich coverage in terms of event counts and trajectory durations, registry filtering resulted in a smaller cohort which highlights the challenge of relying on ICD codes for diagnostic accuracy. Age distribution at the first event, last event, and time of cancer diagnosis together show a broad coverage across different age brackets, with higher age for cancer patients showing its a strong predictor of risk.* ***(Bottom)*** *Cancer registry filtering and trajectory sampling for cancer risk scoring. (A) Cancer counts by site before and after matching EHR diagnosis with linked cancer registry outcomes. This comparison shows the unreliability of cancer outcomes from EHR codes alone. We excluded patients with cancer recorded either only in EHR or only in registry. (B) Cancer diagnosis is often preceded by diagnostic work-up codes that become more frequent closer to the diagnosis. (C) To answer how far ahead in time our model can predict risk, we sampled multiple trajectories per patient. The preceding window was divided into four equal size*

*segments with one endpoint sampled from each segment which prevents concentrating trajectories close to diagnosis. We report resulting endpoint distribution across cancer types.*

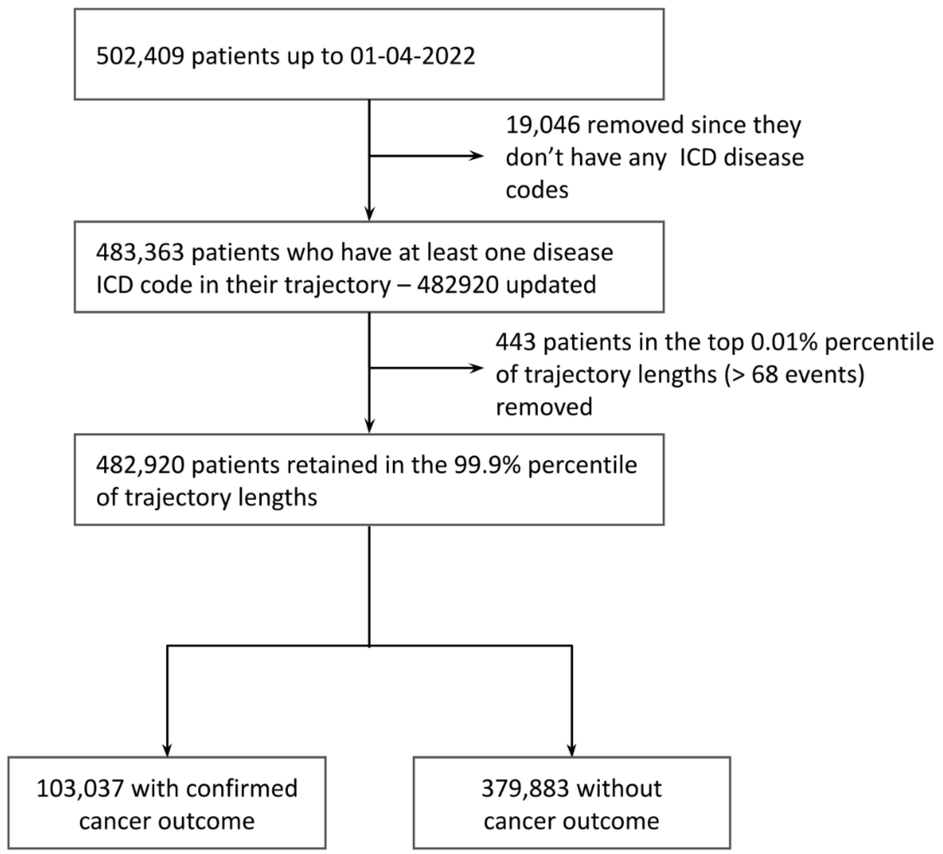


***Figure S4a: Data preparation and quality control filtering of the UKBB cohort.*** *Patient level inclusion and exclusion steps which results in a cohort of 502,409 patients of whom 103,037 had a cancer outcome confirmed through the linked cancer registry. Filtering against cancer registry gives clean labels useful for supervised fine-tuning and reliable estimate of model performance.*

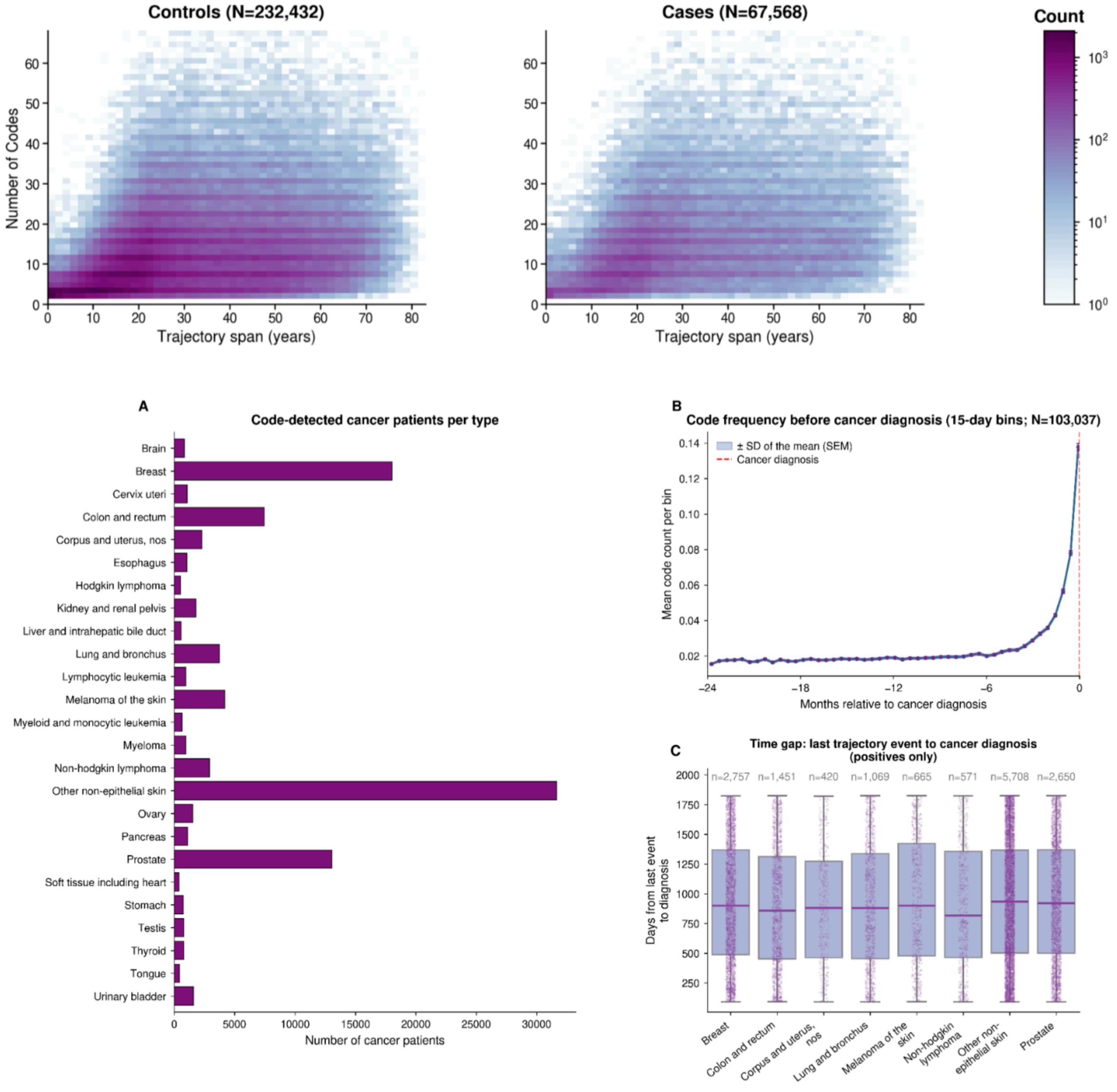


***Figure S4b: Data statistics summary of UKBB cohort. (Top)** Cases and controls have a rich coverage in terms of event counts and trajectory durations, registry filtering resulted in a smaller cohort which highlights the challenge of relying on ICD codes for diagnostic accuracy. Age distribution at the first event, last event, and time of cancer diagnosis together show a broad coverage across different age brackets, with higher age for cancer patients showing its a strong predictor of risk. **(Bottom)** Cancer registry cases and trajectory sampling for cancer risk scoring. (A) All cancer outcomes in UKBB first-occurrence EHR come from cancer registry. (B) Cancer diagnosis is often preceded by diagnostic work-up codes that become more frequent closer to the diagnosis. (C) To answer how far ahead in time our model can predict risk, we sampled multiple trajectories per patient. The preceding window was divided into four equal size*

*segments with one endpoint sampled from each segment which prevents concentrating trajectories close to diagnosis. We report resulting endpoint distribution across cancer types.*

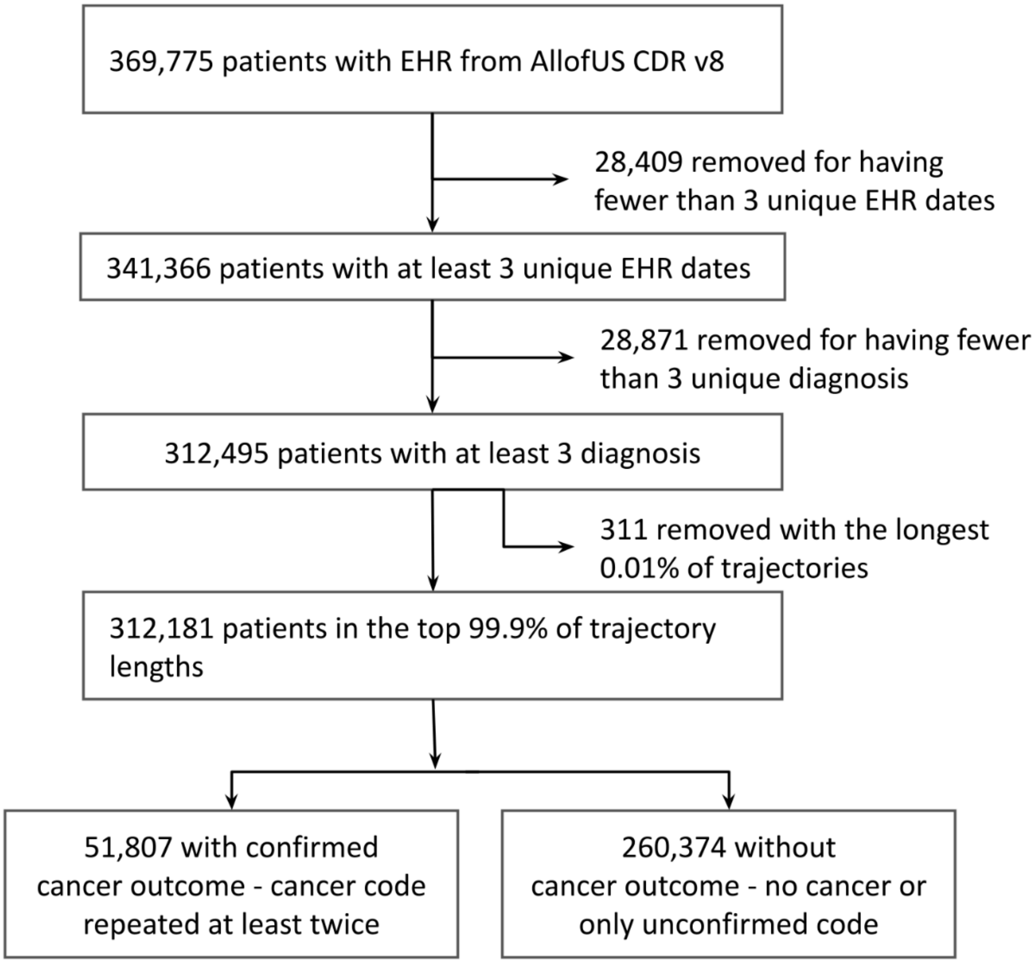


***Figure S5a: Data preparation and quality control filtering of the AoU cohort.*** *Patient level inclusion and exclusion steps which results in a cohort of 312,181 patients of whom 51,807 had a cancer outcome confirmed through the linked cancer registry. Filtering against cancer registry gives clean labels useful for supervised fine-tuning and reliable estimate of model performance.*

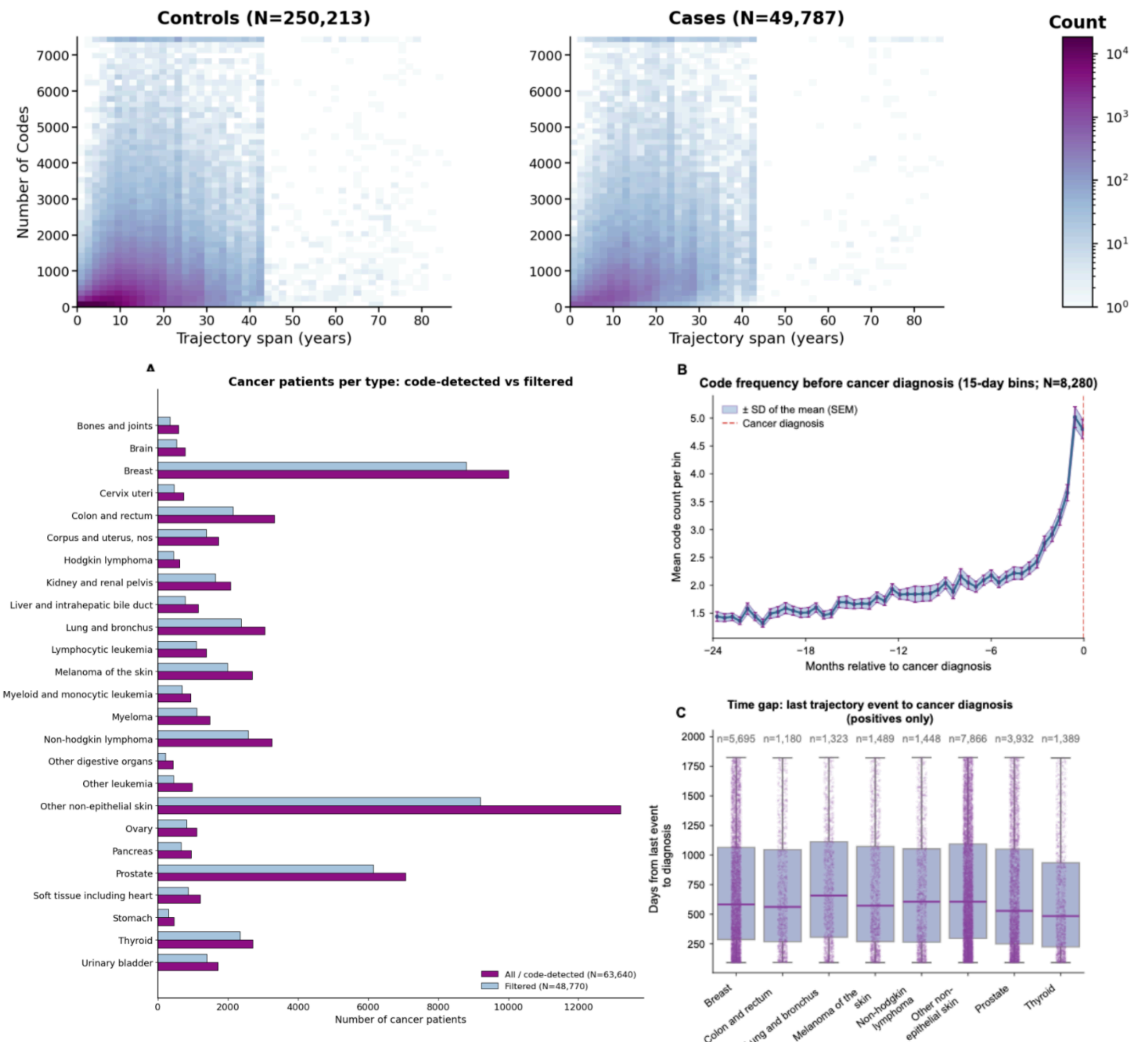


***Figure S5b: Data statistics summary of AoU cohort. (Top)*** *Cases and controls have a rich coverage in terms of event counts and trajectory durations, registry filtering resulted in a smaller cohort which highlights the challenge of relying on ICD codes for diagnostic accuracy. Age distribution at the first event, last event, and time of cancer diagnosis together show a broad coverage across different age brackets, with higher age for cancer patients showing its a strong predictor of risk.* ***(Bottom)*** *Cancer cases filtering and trajectory sampling for cancer risk scoring. (A) Cancer counts by site from the EHR diagnosis and after filtering by retaining only those patients which have the same code repeated in their EHR. Since All of Us does not have a cancer registry, we adopt this procedure to filter out the unreliable cancer outcomes from EHR alone. We excluded patients with cancer recorded either only in EHR or only in registry. (B) Cancer diagnosis is often preceded by diagnostic work-up codes that become more frequent closer to the diagnosis. (C) To answer how far ahead in time our model can predict risk, we sampled multiple trajectories per patient. The preceding window was divided into four equal size segments with one endpoint sampled from each segment which prevents concentrating trajectories close to diagnosis. We report resulting endpoint distribution across cancer types.*

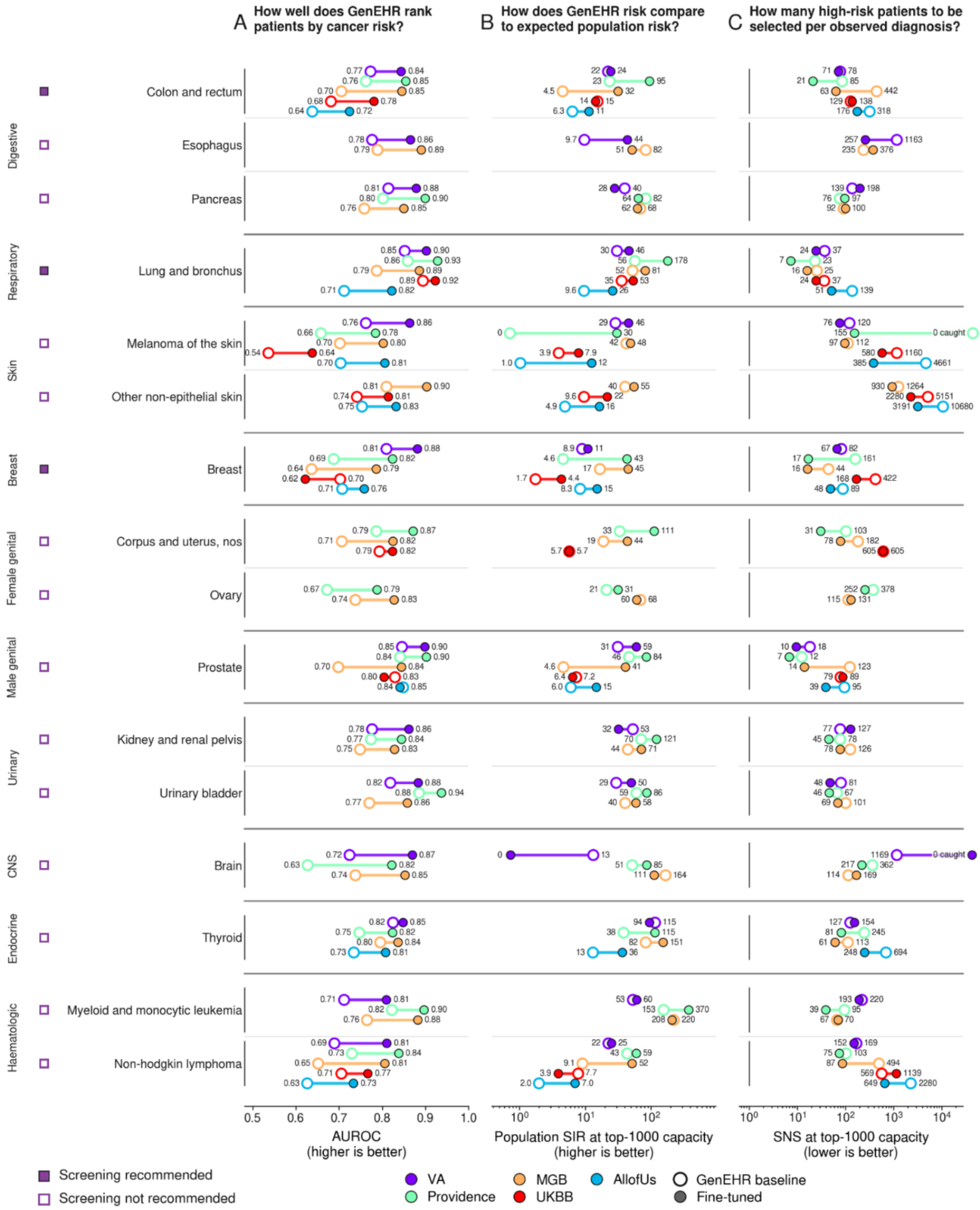


***Figure S6a: Breakdown of model performance for pan-cancer risk prediction (6 month horizon window).*** *Comparison of pretrained and fine-tuned GenEHR for cancer risk prediction.*

*Rows are cancer types. Fine-tuning GenEHR using a time-to-event objective improves AUROC (A). For (B-C), we select the 1000 highest scoring trajectories and evaluate their enrichment relative to the population-wide demographic baseline incidence from SEER for U.S. cohorts and NDRS for UKBB. SNS measures how many patients to be enrolled for advanced screening to find one cancer case, since we know future recorded outcomes here it serves a purpose of a retrospective workload. In practice SNS would depend on the sensitivity of the screening test. Panel (B-C) are on logarithmic scale. Open circles (GenEHR), filled circles (fine-tuned GenEHR). Colors distinguish cohorts: MGB (orange), PROV (green), VA (purple), AoU (cyan), and UKBB (red). Cancers shown are those with 2000 or more patients in at least two cohorts.*

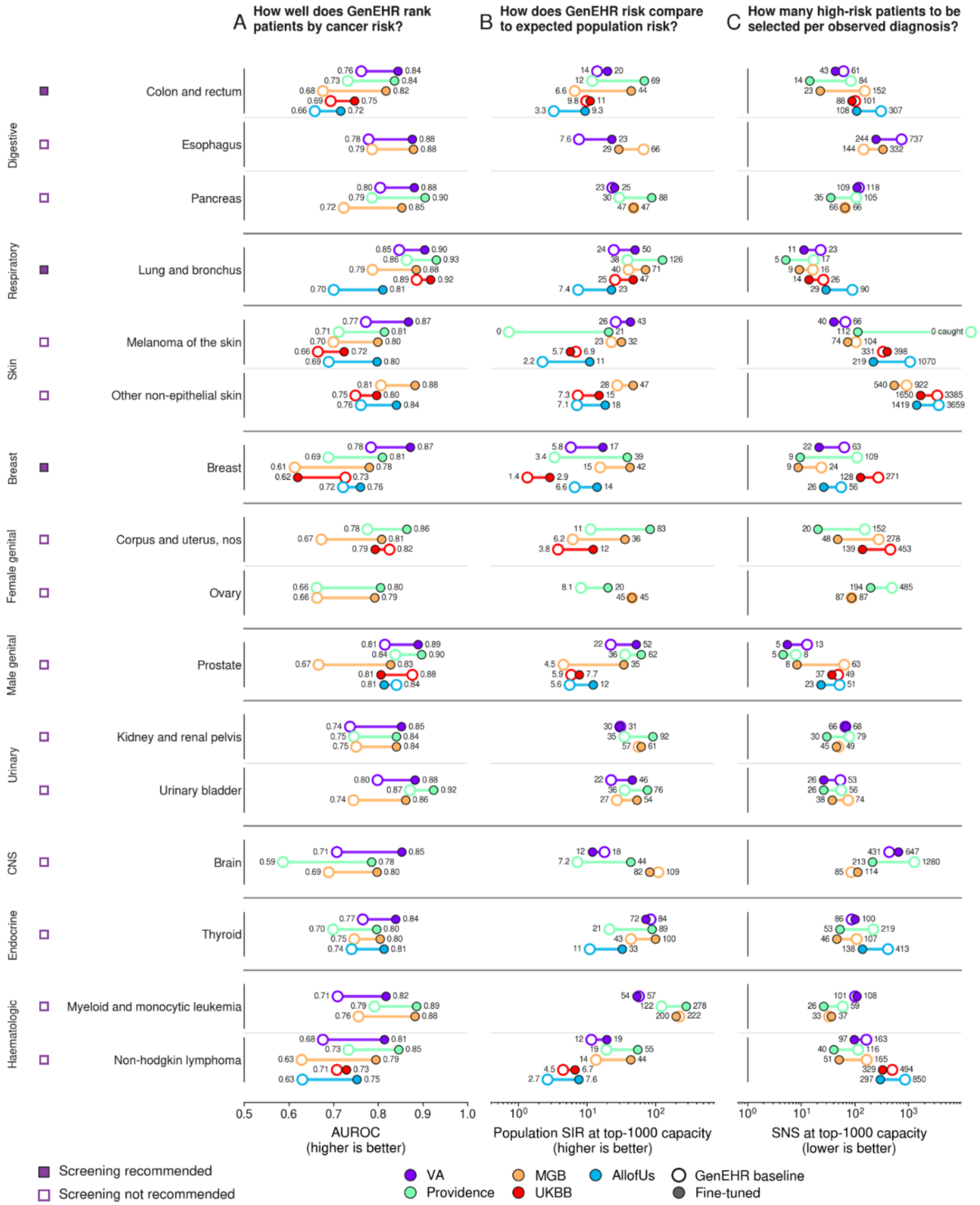


**Figure S6b:** ***Breakdown of model performance for pan-cancer risk prediction (12 month horizon window).*** *Comparison of pretrained and fine-tuned GenEHR for cancer risk prediction. Rows are cancer types. Fine-tuning GenEHR using a time-to-event objective improves AUROC (A). For (B-C), we select the 1000 highest scoring trajectories and evaluate their enrichment*

*relative to the population-wide demographic baseline incidence from SEER for U.S. cohorts and NDRS for UKBB. SNS measures how many patients to be enrolled for advanced screening to find one cancer case, since we know future recorded outcomes here it serves a purpose of a retrospective workload. In practice SNS would depend on the sensitivity of the screening test. Panel (B-C) are on logarithmic scale. Open circles (GenEHR), filled circles (fine-tuned GenEHR). Colors distinguish cohorts: MGB (orange), PROV (green), VA (purple), AoU (cyan), and UKBB (red). Cancers shown are those with 2000 or more patients in at least two cohorts.*

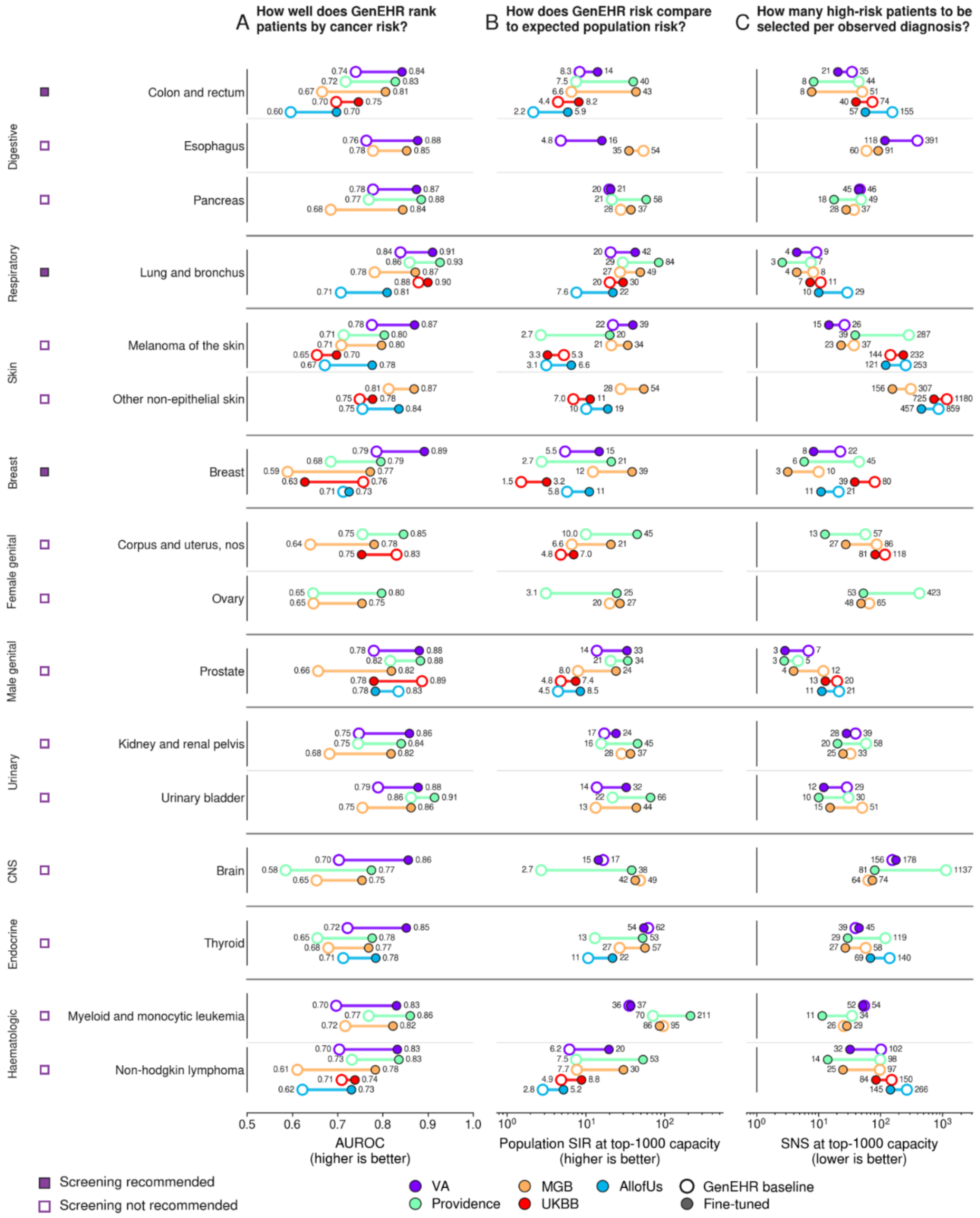


**Figure S6c:** ***Breakdown of model performance for pan-cancer risk prediction (24 month horizon window).*** *Comparison of pretrained and fine-tuned GenEHR for cancer risk prediction. Rows are cancer types. Fine-tuning GenEHR using a time-to-event objective improves AUROC (A). For (B-C), we select the 1000 highest scoring trajectories and evaluate their enrichment*

*relative to the population-wide demographic baseline incidence from SEER for U.S. cohorts and NDRS for UKBB. SNS measures how many patients to be enrolled for advanced screening to find one cancer case, since we know future recorded outcomes here it serves a purpose of a retrospective workload. In practice SNS would depend on the sensitivity of the screening test. Panel (B-C) are on logarithmic scale. Open circles (GenEHR), filled circles (fine-tuned GenEHR). Colors distinguish cohorts: MGB (orange), PROV (green), VA (purple), AoU (cyan), and UKBB (red). Cancers shown are those with 2000 or more patients in at least two cohorts.*

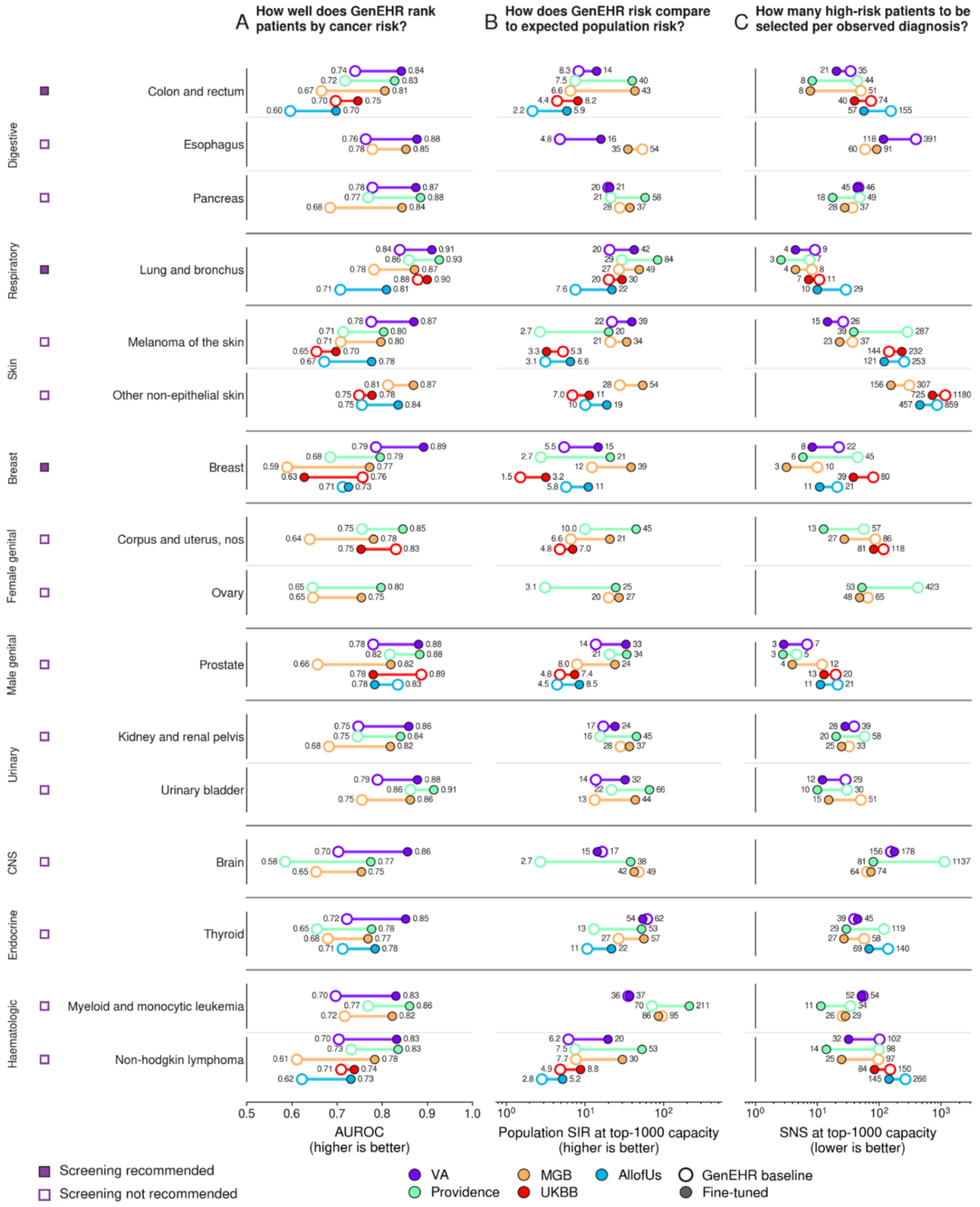


**Figure S6d: *Breakdown of model performance for pan-cancer risk prediction (36 month horizon window).*** *Comparison of pretrained and fine-tuned GenEHR scores for cancer risk prediction. Rows are cancer types. Pretrained GenEHR learns cancer-associated signals useful for ranking cases and controls without any supervision from the cancer registry. Fine-tuning further improves AUROC (A). For (B-C), we select the 1000 highest scoring trajectories and*

*evaluate their enrichment relative to the population-wide demographic baseline incidence from SEER for U.S. cohorts and NDRS for UKBB. SNS measures how many patients to be enrolled for advanced screening to find one cancer case, since we know future recorded outcomes here it serves a purpose of a retrospective workload. In practice SNS would depend on the sensitivity of the screening test. Open circles (GenEHR), filled circles (fine-tuned GenEHR). Colors distinguish cohorts: MGB (orange), PROV (green), VA (purple), AoU (cyan), and UKBB (red). Cancers shown are those with 2000 or more patients in at least two cohorts.*

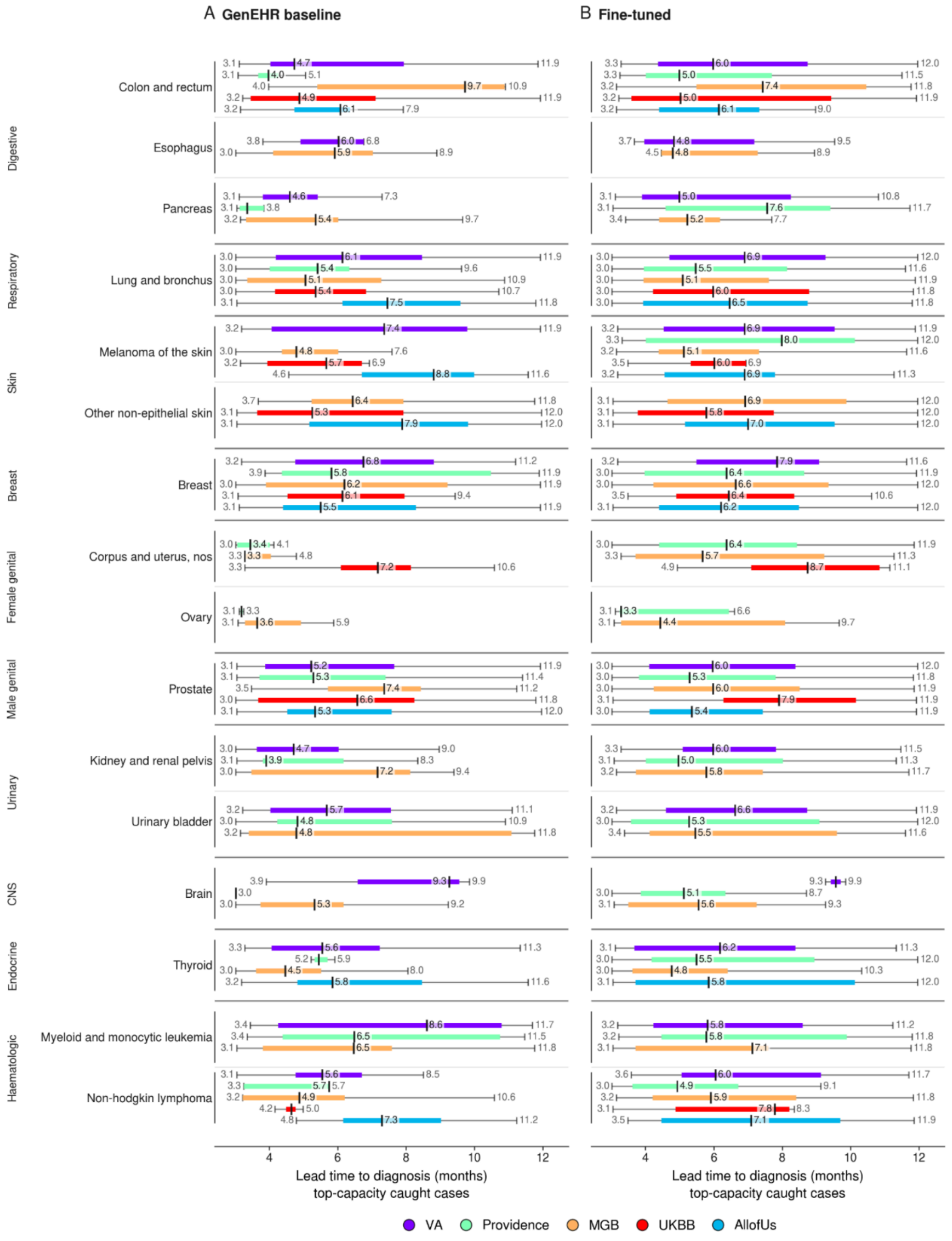


***Figure S6e: Lead-time among true-positive patients at operating threshold N=1000 (12 month horizon window).*** *For patients ranked within the top 1000 who developed cancer within the prediction horizon, we plot the interval between the endpoint of trajectory and the registry*

*recorded diagnosis date. Box plot whiskers indicate 1.5 x inter quartile range (IQR) below and above first and third quartiles, respectively. (A) Pretrained GenEHR and (B) Fine-tuned GenEHR. Horizontal axis is the number of months from the endpoint of trajectory to cancer diagnosis. Cancers shown are those with 2000 or more patients in at least two cohorts.*

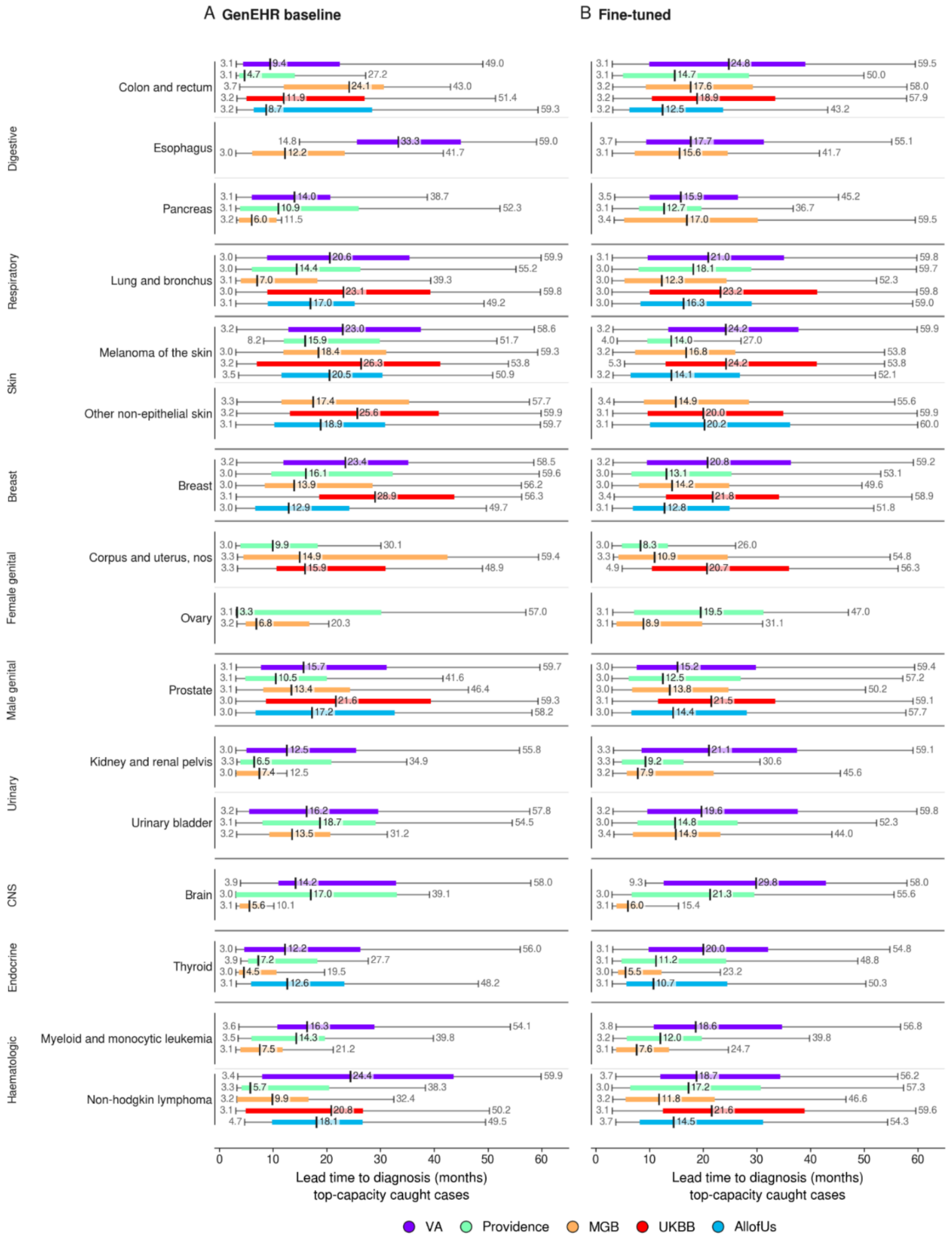


***Figure S6f: Lead-time among true-positive patients at operating threshold N=1000 (60 month horizon window).*** *For patients ranked within the top 1000 who developed cancer within*

the prediction horizon, we plot the interval between the endpoint of trajectory and the registry recorded diagnosis date. Box plot whiskers indicate 1.5 x inter quartile range (IQR) below and above first and third quartiles, respectively. (A) Pretrained GenEHR and (B) Fine-tuned GenEHR. Horizontal axis is the number of months from the endpoint of trajectory to  cancer diagnosis. Cancers shown are those with 2000 or more patients in at least two cohorts.*

**MGB Results.**

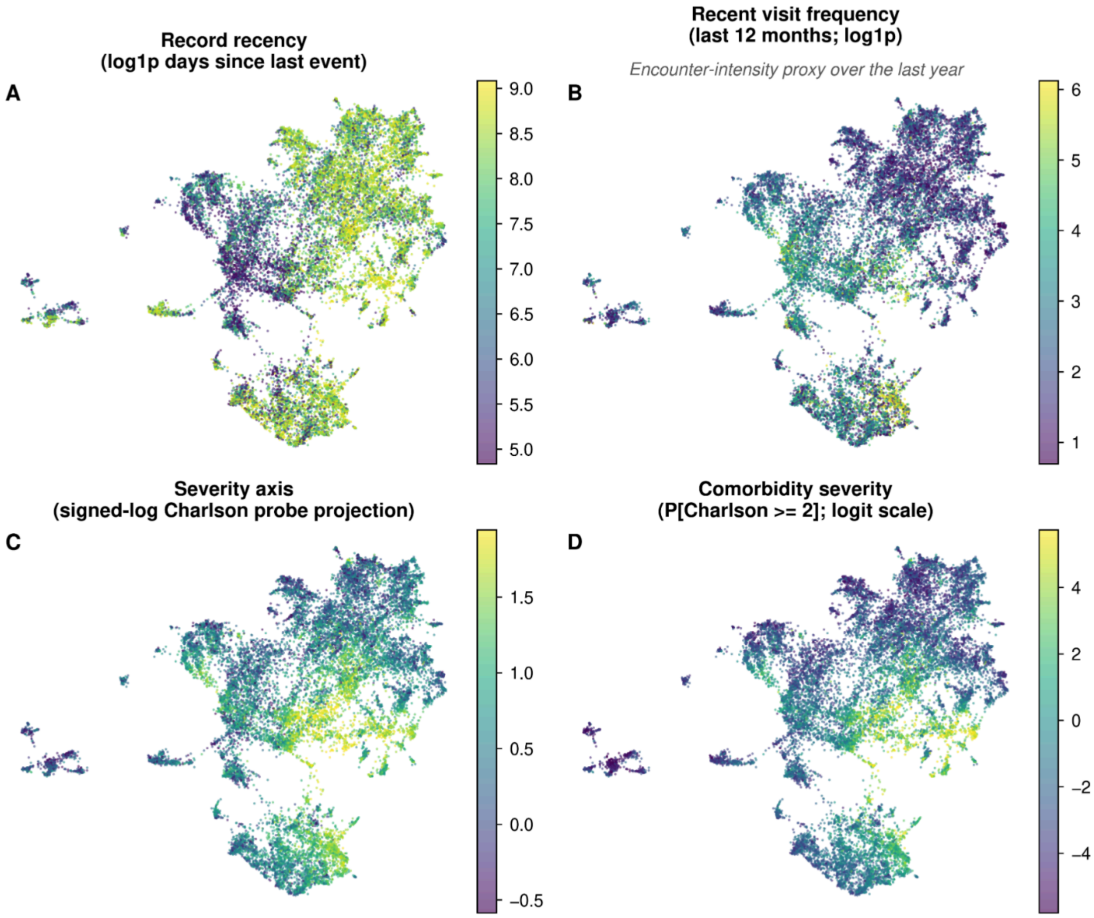


***Figure S7: Patient health state representation (MGB Cohort).*** *All four panels are two dimensional projections of patient trajectory representation obtained from the penultimate layer of GenEHR. Each has a different overlay: (A) time gap since last visit (B) number of visits in preceding year as a proxy for acuity. (C) a linear probe was fitted on feature representation for predicting CCI  overlay with probe prediction (D) only patients who have CCI above chosen threshold. Gradients in CCI imply that representations capture information about overall morbidity and healthcare use.*

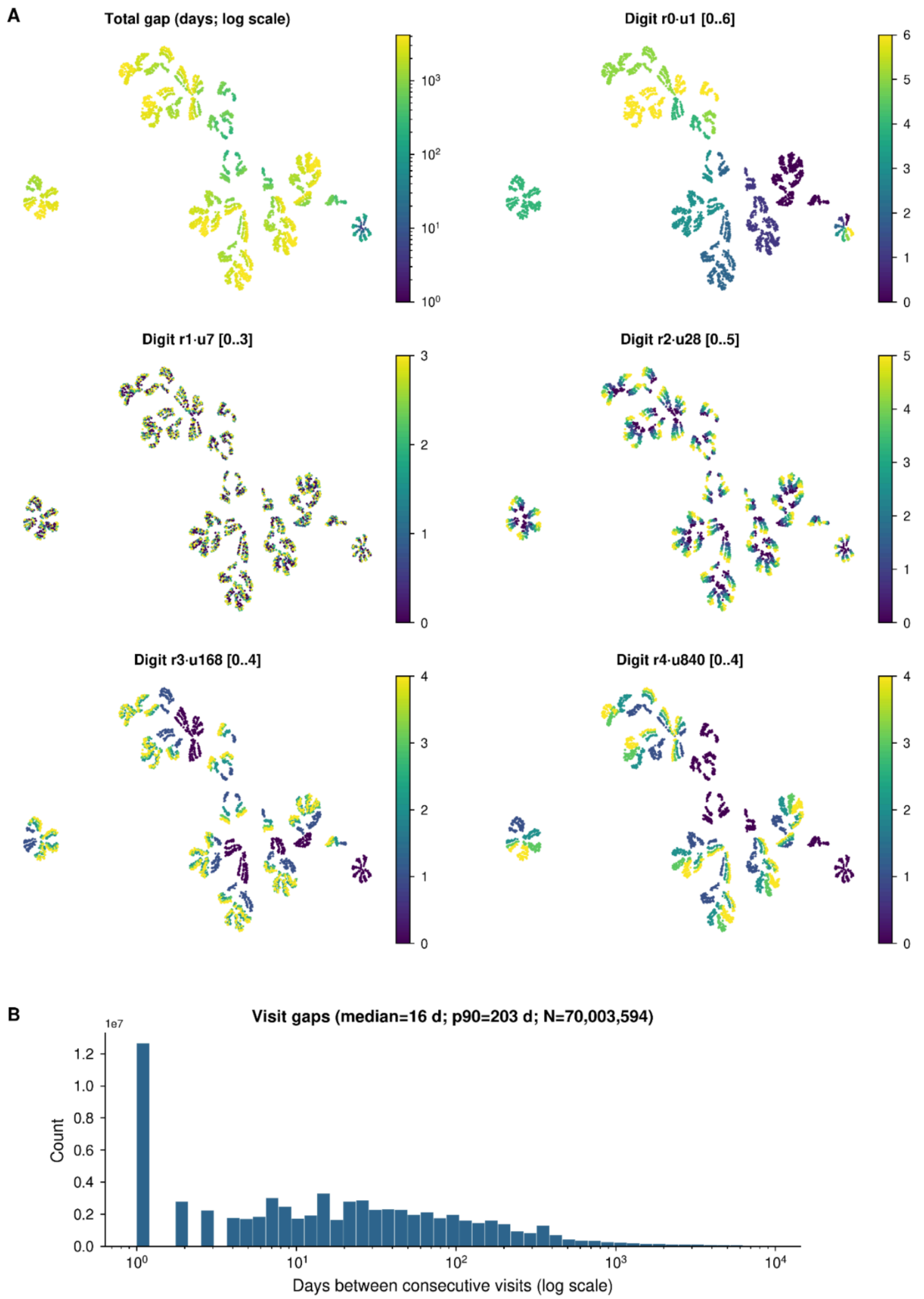


***Figure S8: Learned RATE representation and observed time gaps in MGB.*** *UMAP projection of learned representation of 4200 days, colored by the value of each radix digit together with the observed distribution of inter-visit gaps. (A) Each point represents one encoded gap from 1 to 4200 days where color is the gap length. The other five show the digit*

*with place values 1, 7, 28, 168, and 840 days with color indicating lower to higher values on each scale. (B) Inter-visit time interval distribution.*

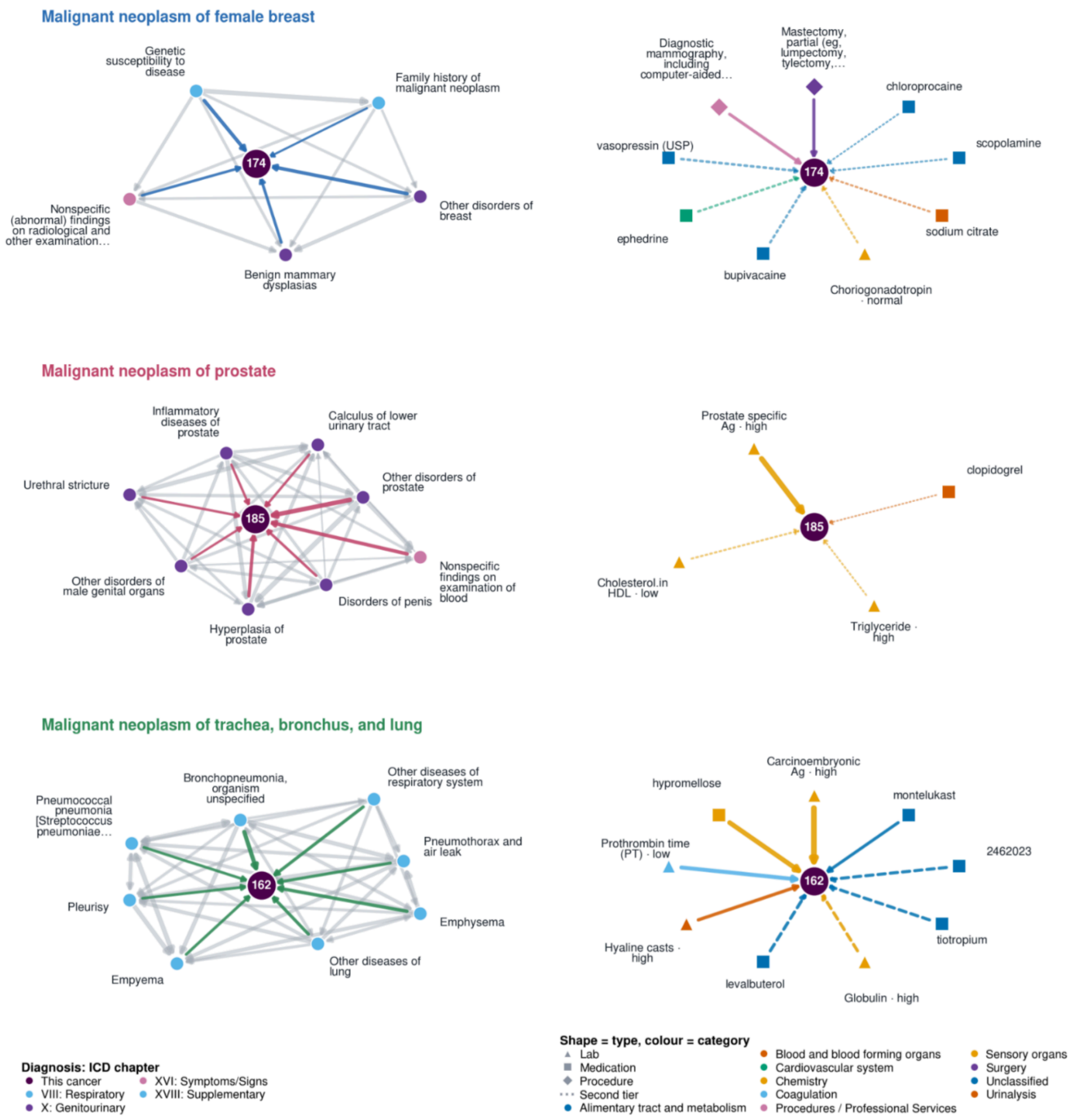


***Figure S9a: Breakdown of within and across modality belief graphs across cancer types.*** *Graphs are derived from the held-out data using cPMI of the decoder. Nodes are clinical events and directed edges show positive cPMI after filtering noise and weak associations (Supplement Methods). The neighborhoods are associations learned from data they do not capture causal relations. (Left) Diagnoses neighbors; (Right) neighborhood from other modalities. Edge width reflects retained association strength, and node size is the support as defined in Methods.*

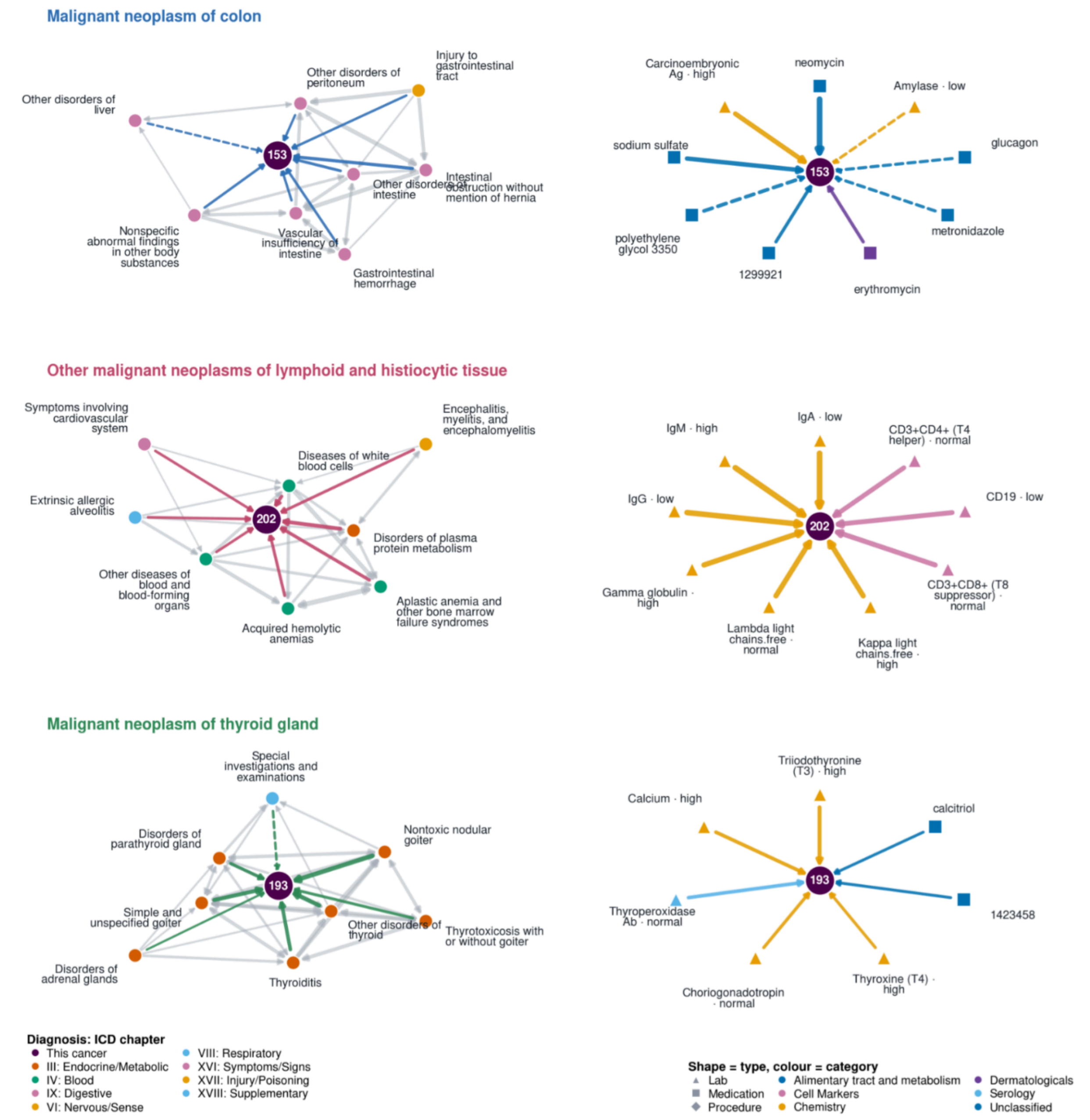


***Figure S9b: Breakdown of within and across modality belief graphs across cancer types.*** *Graphs are derived from the held-out data using cPMI of the decoder. Nodes are clinical events and directed edges show positive cPMI after filtering noise and weak associations (Supplement Methods). Edge width reflects retained association strength, and node size is the support as defined in Methods. (Left) Diagnoses neighbors; (Right) neighborhood from other modalities. The neighborhoods are associations learned from data they do not capture causal relations.*

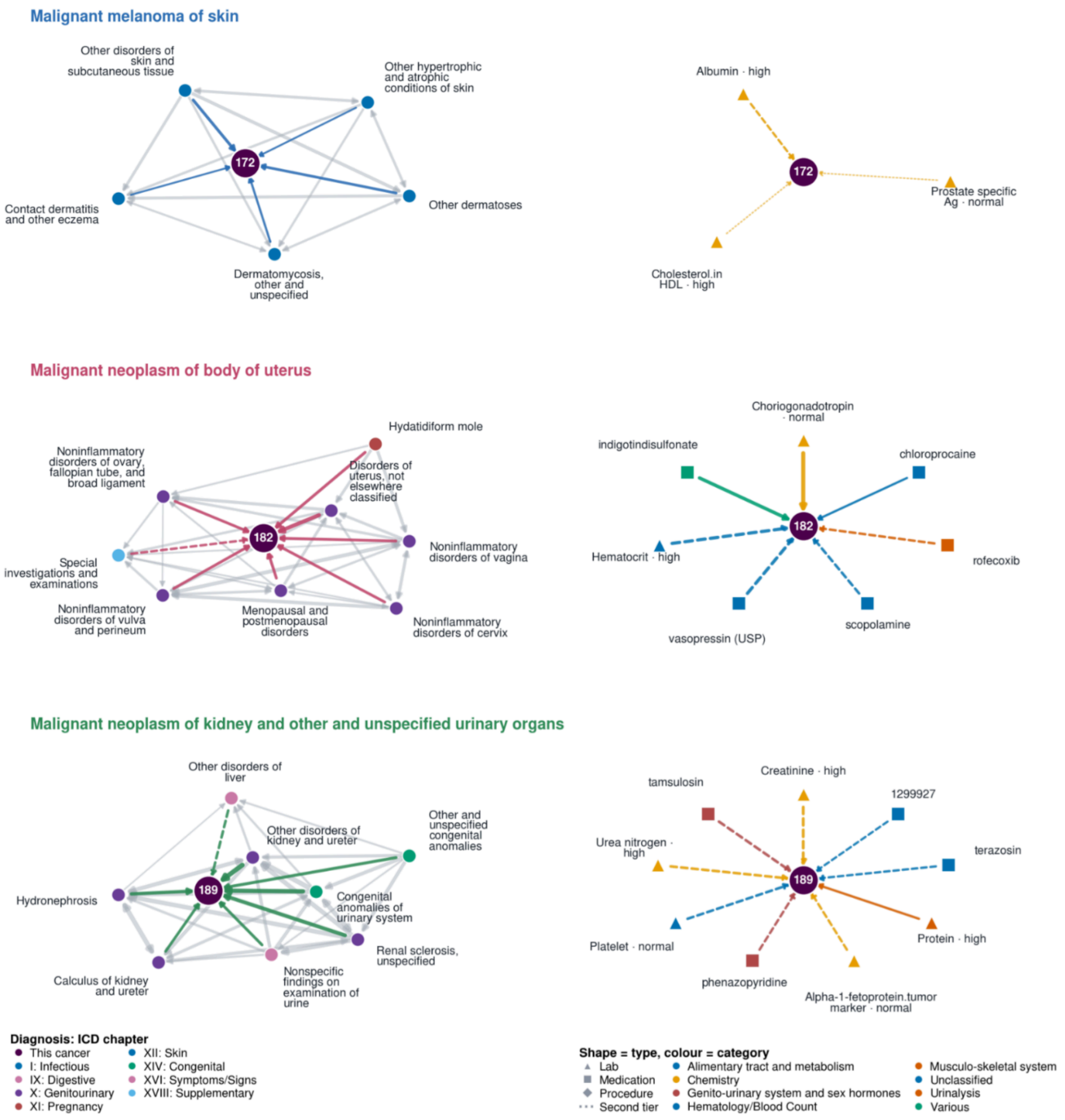


***Figure S9c: Breakdown of within and across modality belief graphs across cancer types.*** *Graphs are derived from the held-out data using cPMI of the decoder. Nodes are clinical events and directed edges show positive cPMI after filtering noise and weak associations (Supplement Methods). Edge width reflects retained association strength, and node size is the support as defined in Methods. (Left) Diagnoses neighbors; (Right) neighborhood from other modalities. The neighborhoods are associations learned from data they do not capture causal relations.*

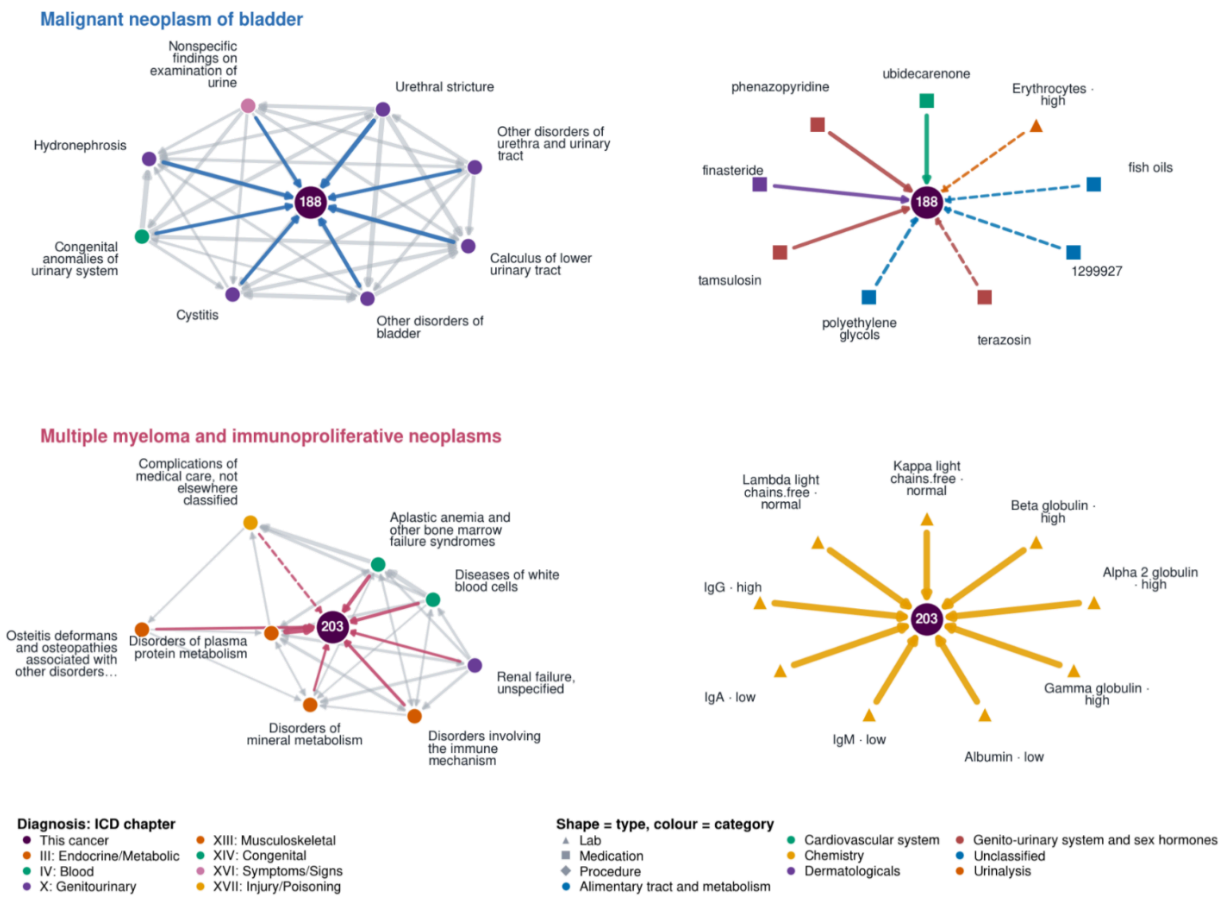


***Figure S9d: Breakdown of within and across modality belief graphs across cancer types.*** *Graphs are derived from the held-out data using cPMI of the decoder. Nodes are clinical events and directed edges show positive cPMI after filtering noise and weak associations (Supplement Methods). Edge width reflects retained association strength, and node size is the support as defined in Methods. (Left) Diagnoses neighbors; (Right) neighborhood from other modalities. The neighborhoods are associations learned from data they do not capture causal relations.*

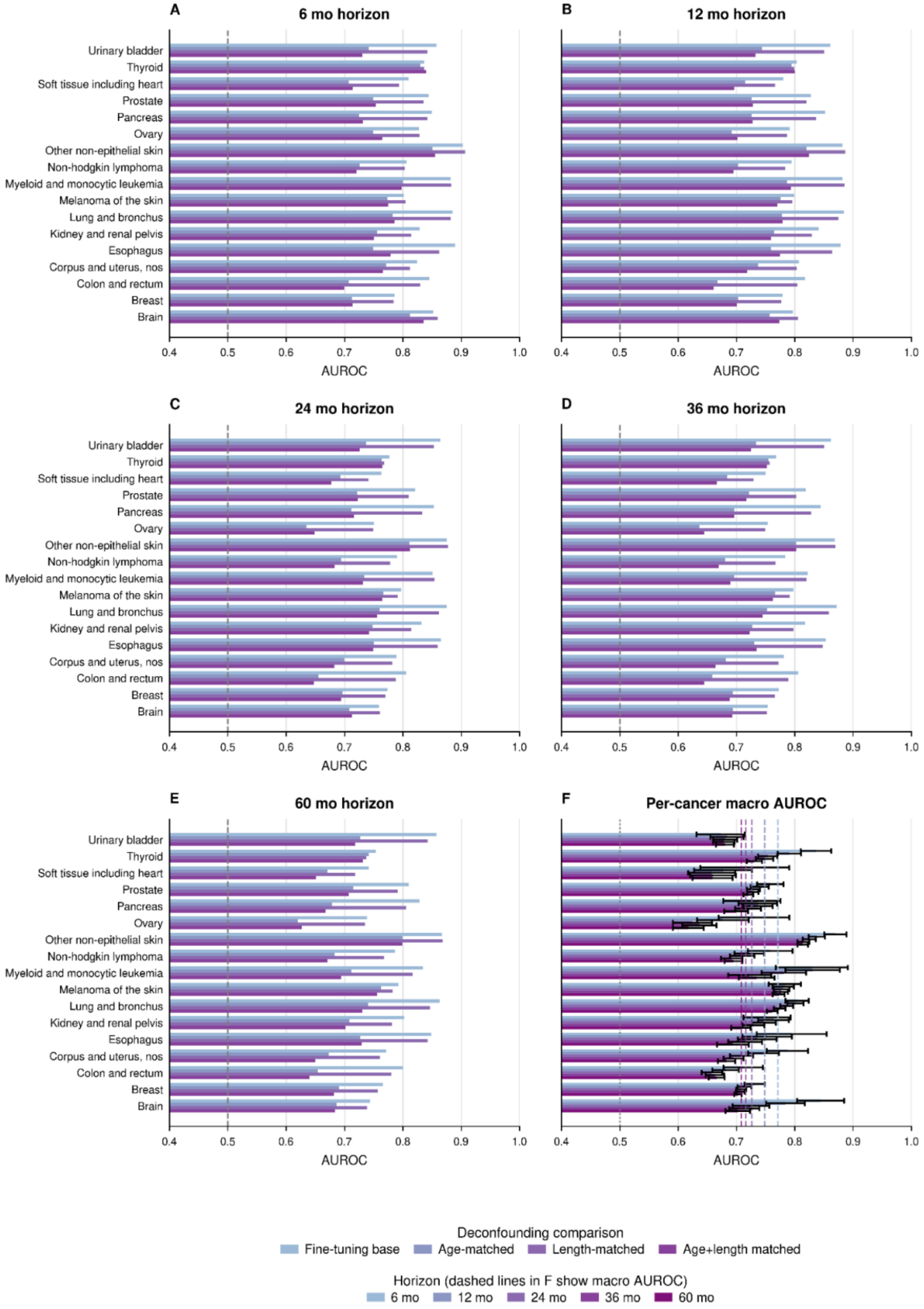


***Figure S10a: Sensitivity of MGB cancer discrimination to age and trajectory length on cancer risk.*** *Cases and controls were matched by age bracket at the endpoint of trajectory as well as with an equal number of events. Age matched results in a drop of performance, but overall performance shows the model just doesn't rely on age and learns from the two combined. Length matched shows the results are not sensitive to the number of codes in patient*

trajectory. (A-E) are results for 6, 12, 24, 36, and 60 month horizons, with each row comparing original evaluation with age matching, event count matching, and both combined.

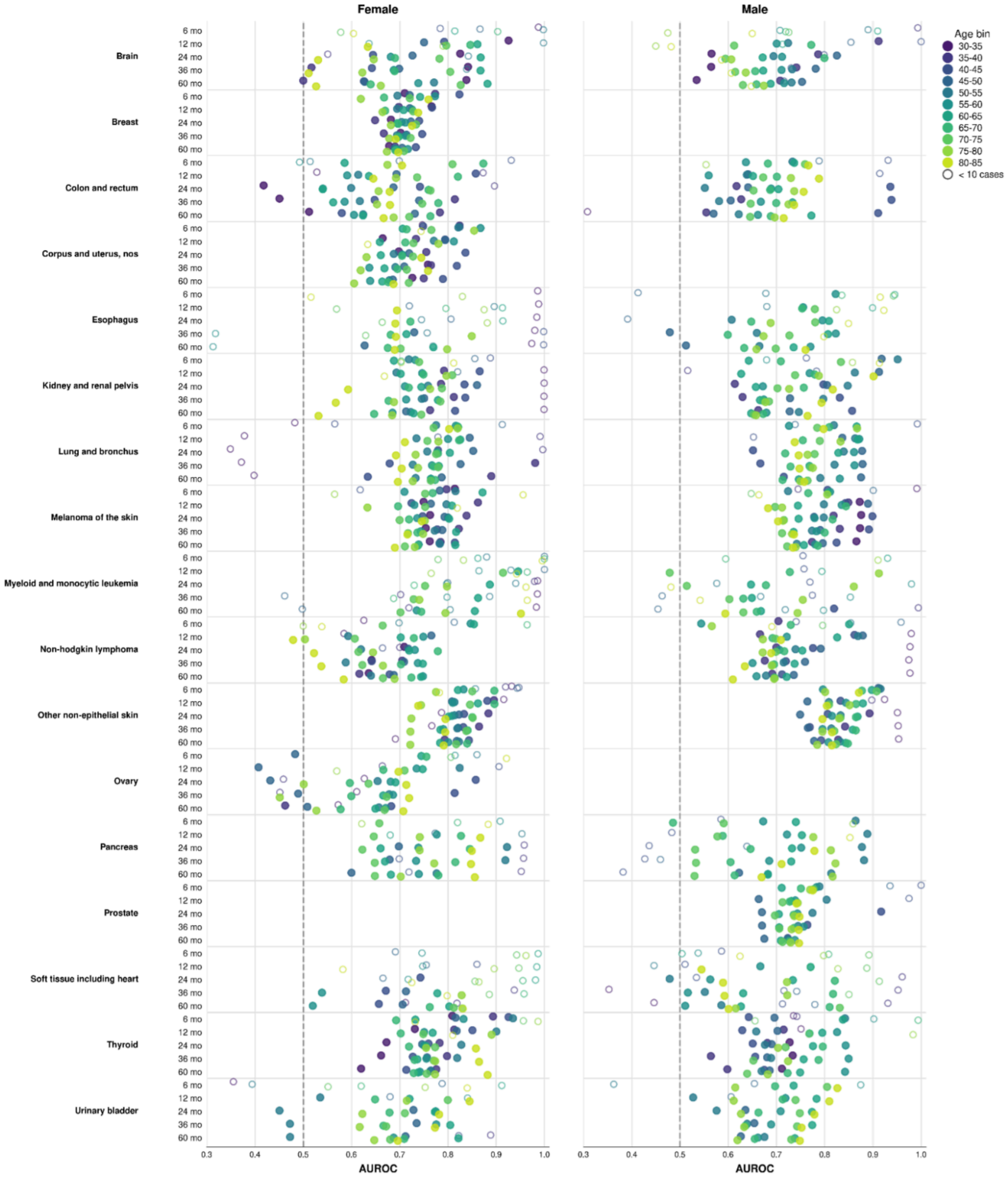


***Figure S10b: Breakdown of predictive performance by age-brackets and sex.*** *Performance by age and sex bracket for different groups assessing heterogeneity and data sparsity. (Left) Performance on female population for each cancer (Right) Performance on male population for each cancer. Both are reported for all six (6, 12, 24, 36, and 60 months) prediction horizons.*

*Color encodes five year age groups, and hollow circle means fewer than 10 positive outcomes in each bin.*

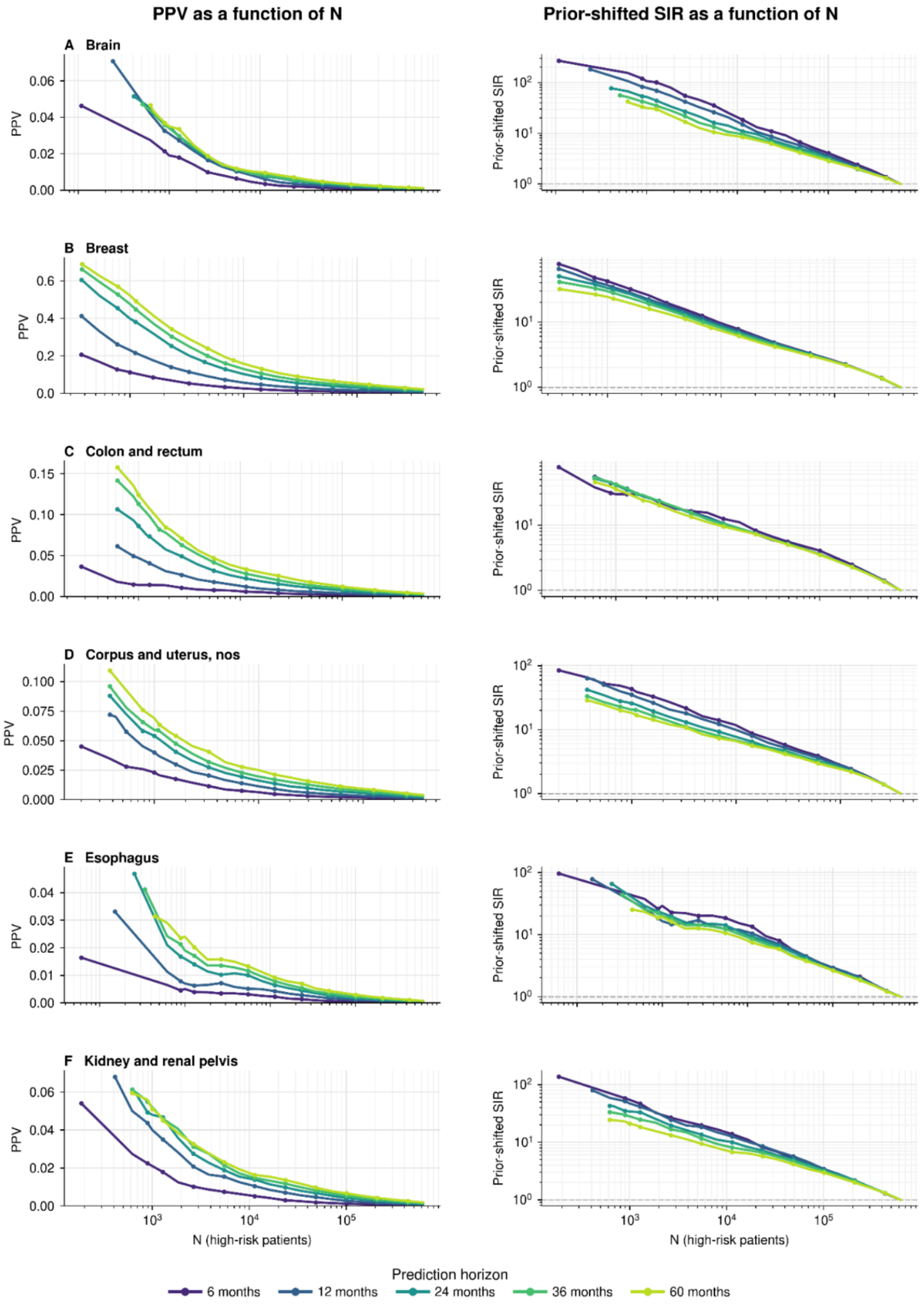


***Figure S11a: Clinical utility of GenEHR fine-tuned model for nominating patients for screening by choosing an operating threshold****. PPV (left) and SIR (right) plotted as a*

*function of operating threshold N shows a net benefit of detecting one cancer at the cost of increasing false positives. (A-F) brain, breast, colon/rectum, corpus/uterus, esophagus, and kidney/renal-pelvis cancer.*

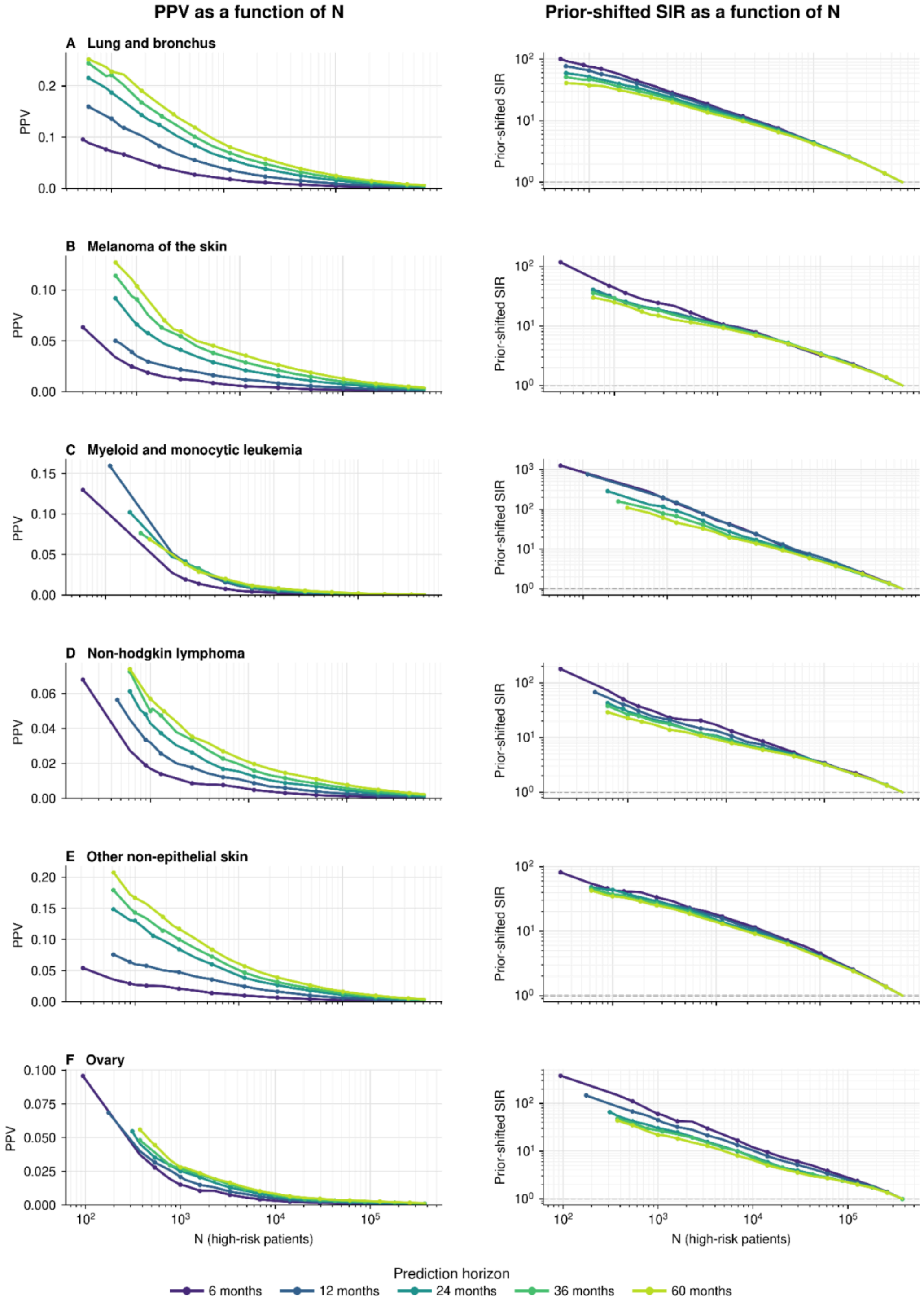


***Figure S11b: Clinical utility of GenEHR fine-tuned model for nominating patients for screening by choosing an operating threshold****. We plot PPV (left) and SIR (right) as a*

*function of operating threshold, this shows a net benefit of detecting one cancer at the cost of increasing false positives. (A-F) lung/bronchus, melanoma, myeloid/monocytic leukemia, non-Hodgkin lymphoma, other non-epithelial skin cancer, and ovary cancer.*

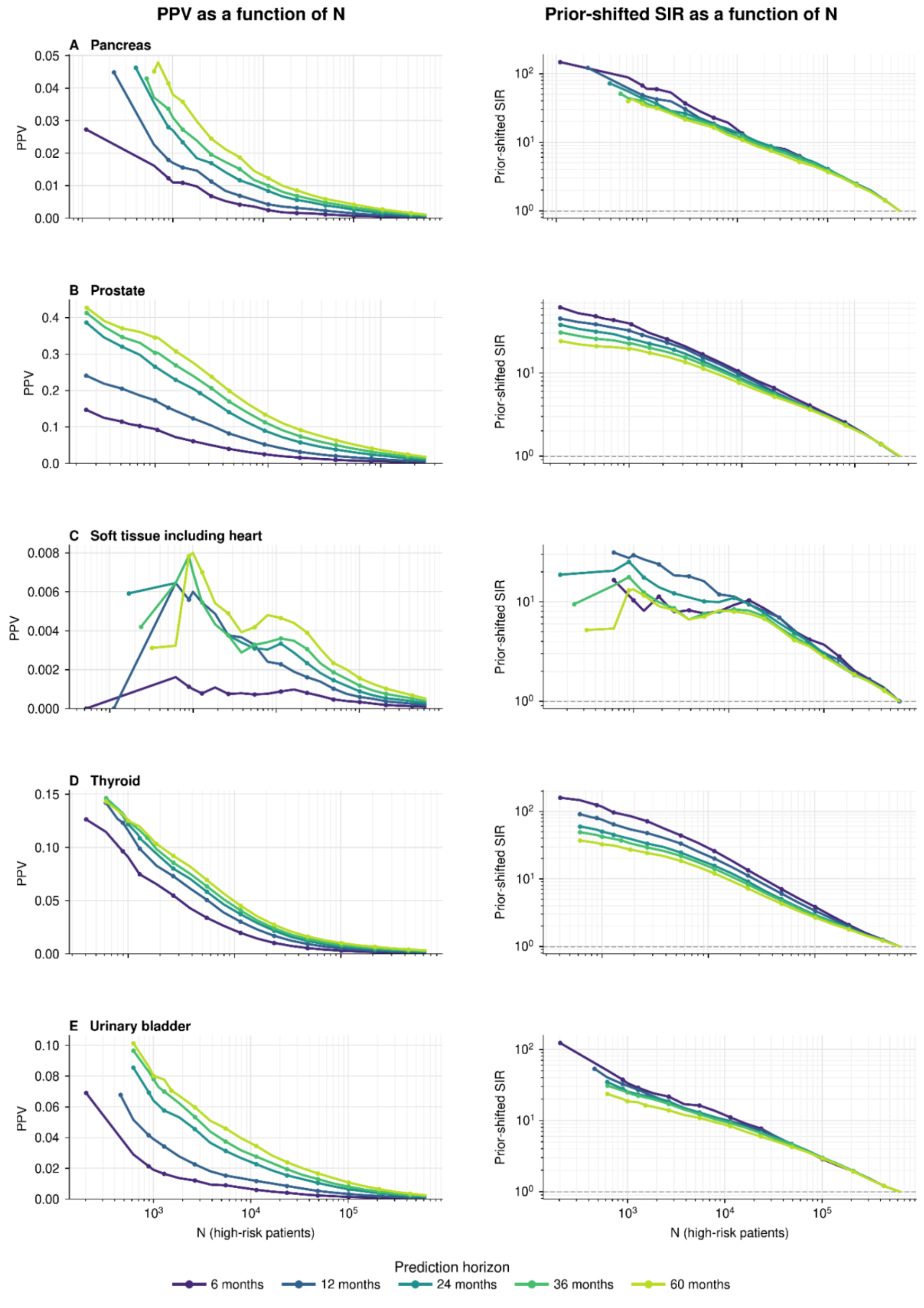

***Figure S11c: Clinical utility of GenEHR fine-tuned model for nominating patients for screening by choosing an operating threshold**. We plot PPV (left) and SIR (right) as a function of operating threshold, this shows a net benefit of detecting one cancer at the cost of increasing false positives. (A-F) pancreas, prostate, soft tissue/heart, thyroid, and urinary-bladder cancer.*

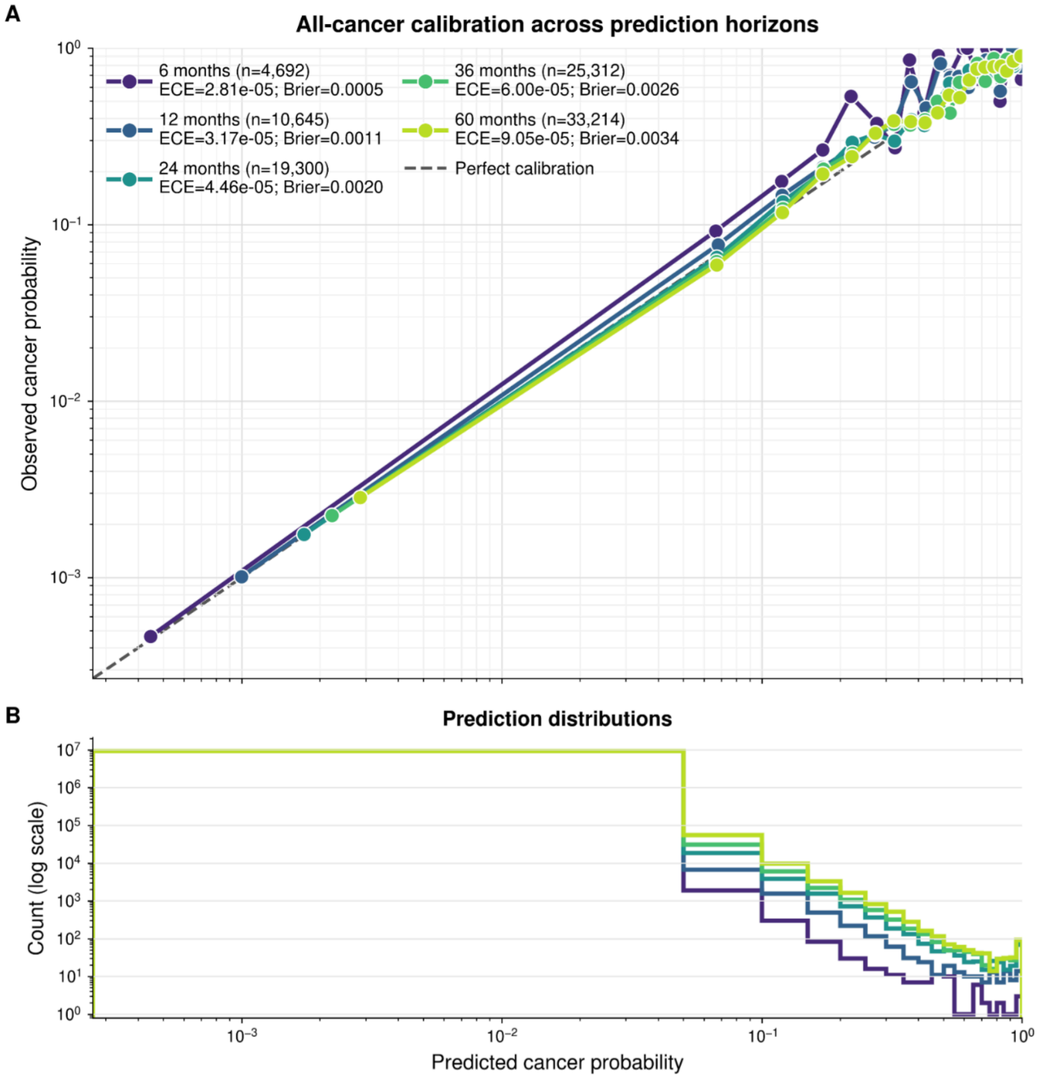


***Figure S12: Pan-cancer risk model is well calibrated across horizons and cancer types.** Observed cancer frequency against predicted probability across calibration bins for different horizons, where diagonal line indicates perfect calibration. Both ECE and Brier score describe fine-tuned cancer-risk probabilities (not next event calibration in Fig 3).*

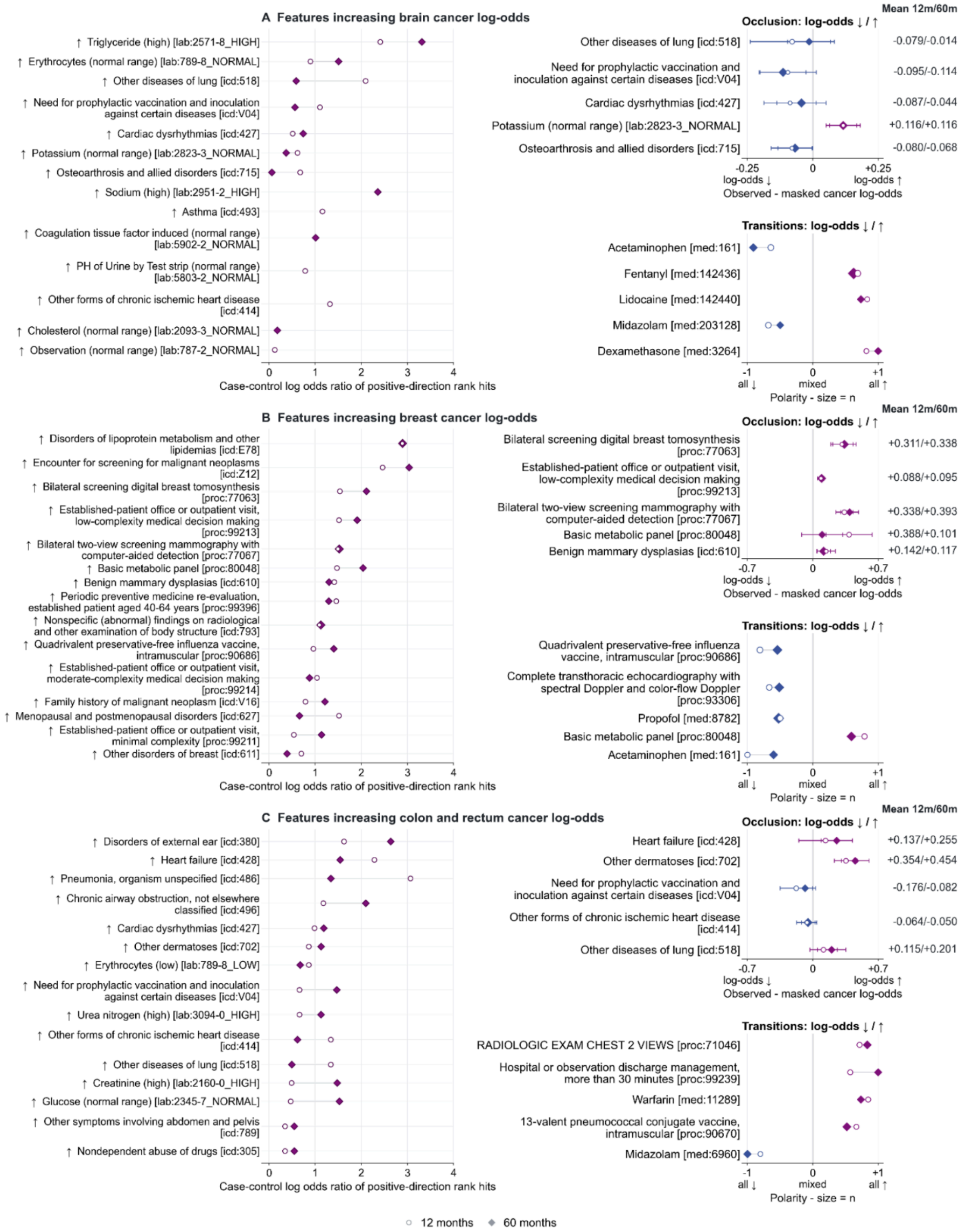


***Figure S13a: Interpretation of features for selected cancer types (brain; breast; colon and rectum) in MGB cohort.*** *(Left) Case-control enrichment among top-ranked positive IG scores, x-axes values are log odds ratio of feature rank hits. (Upper right) Occlusion shows mean*

*predicted log odds with the event observed minus masked, with whiskers of +/-1.96 occurrence level errors, positive means observed feature score is higher. (Lower right) Score-change directions around medication and procedure transitions (-1 all retained changes decrease the score, +1 all increase it).*

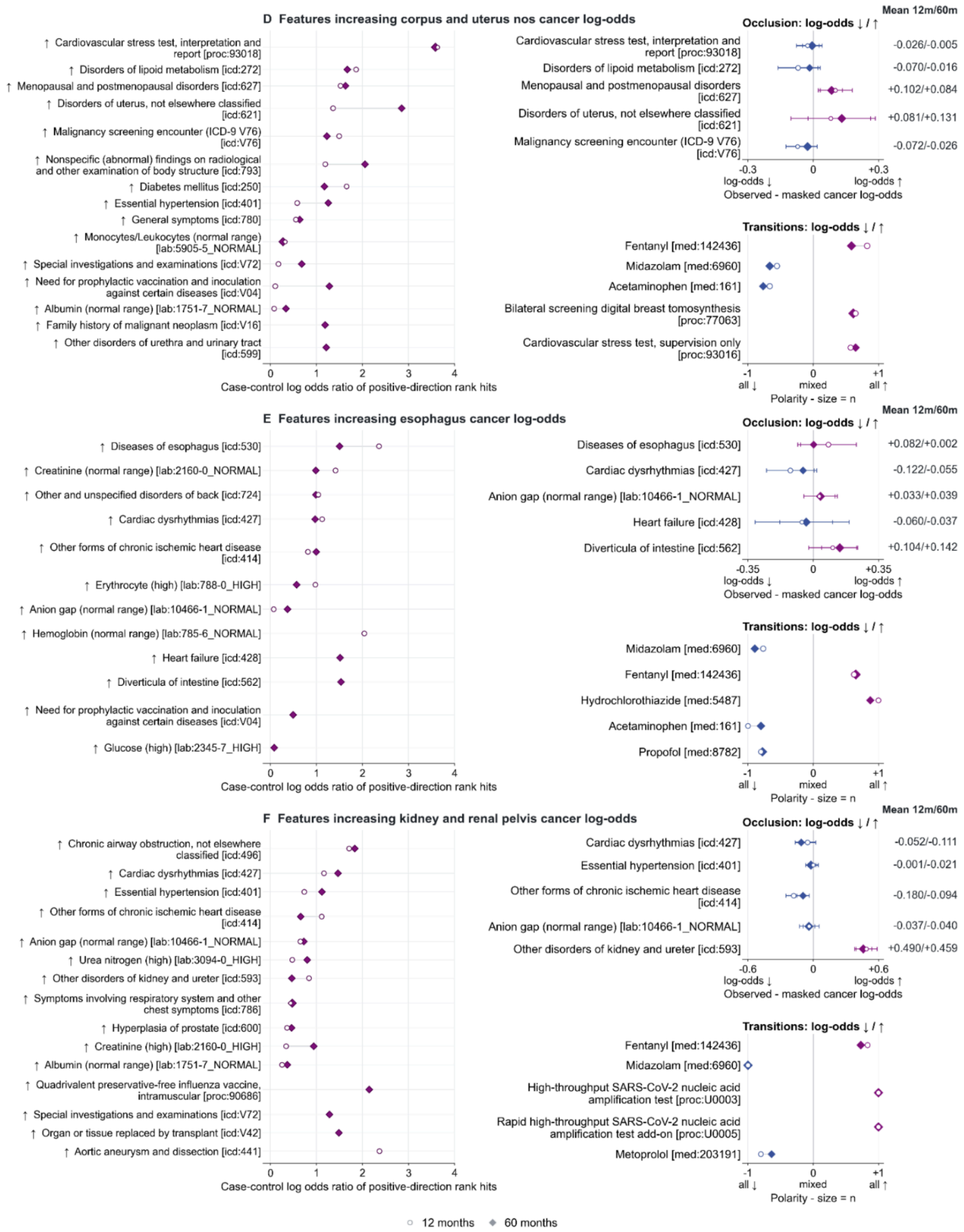


***Figure S13b: Interpretation of features for selected cancer types (corpus and uterus; esophagus; kidney and renal pelvis) in MGB cohort.*** *(Left) Case-control enrichment among top-ranked positive IG scores, x-axes values are log odds ratio of feature rank hits. (Upper right) Occlusion shows mean predicted log odds with the event observed minus masked, with*

*whiskers of +/-1.96 occurrence level errors, positive means observed feature score is higher. (Lower right) Score-change directions around medication and procedure transitions (-1 all retained changes decrease the score, +1 all increase it).*

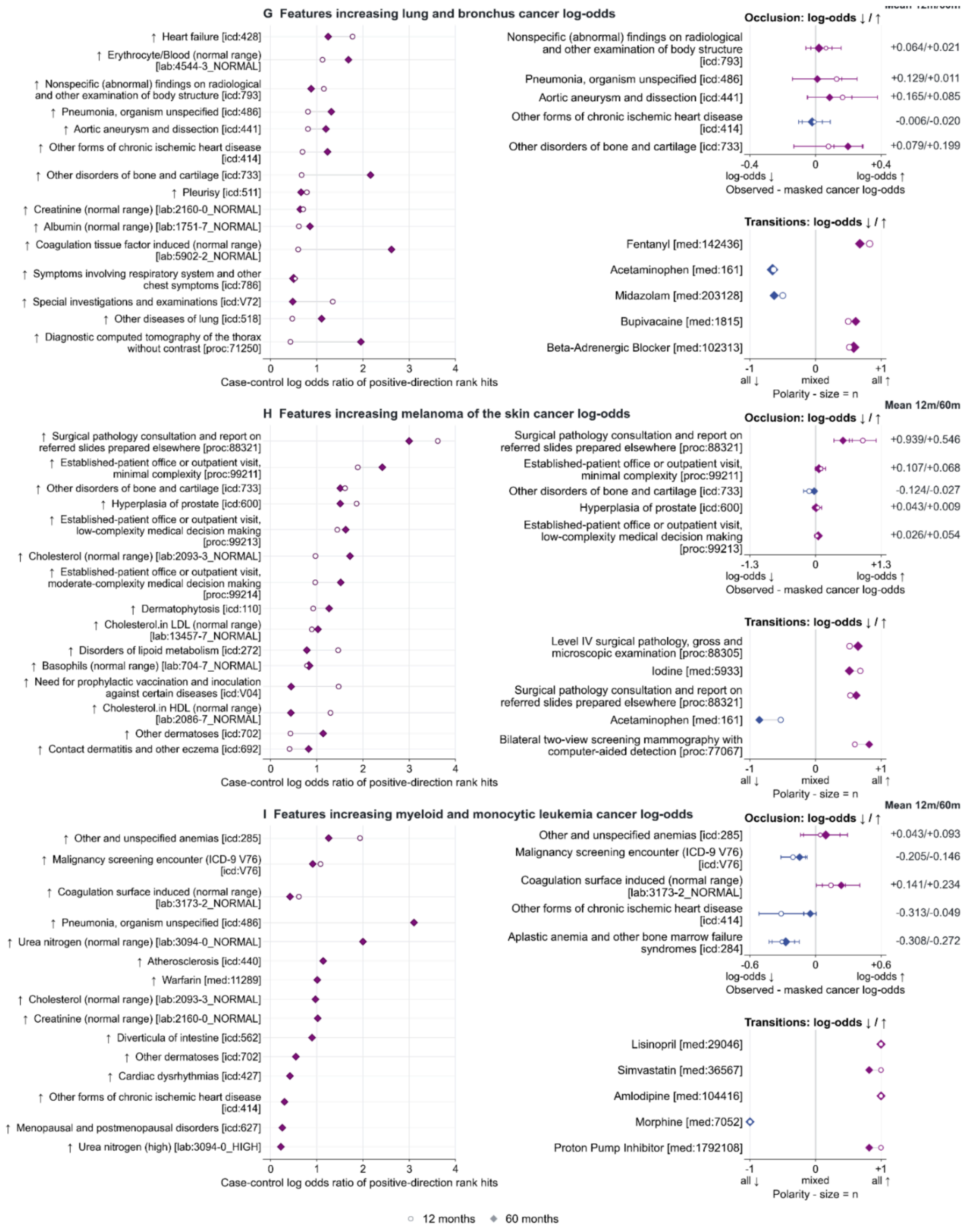


***Figure S13c: Interpretation of features for selected cancer types (lung and bronchus; melanoma of the skin;myeloid and monocytic leukemia) in MGB cohort.*** *(Left) Case-control enrichment among top-ranked positive IG scores, x-axes values are log odds ratio of feature rank hits. (Upper right) Occlusion shows mean predicted log odds with the event*

*observed minus masked, with whiskers of +/-1.96 occurrence level errors, positive means observed feature score is higher. (Lower right) Score-change directions around medication and procedure transitions (-1 all retained changes decrease the score, +1 all increase it).*

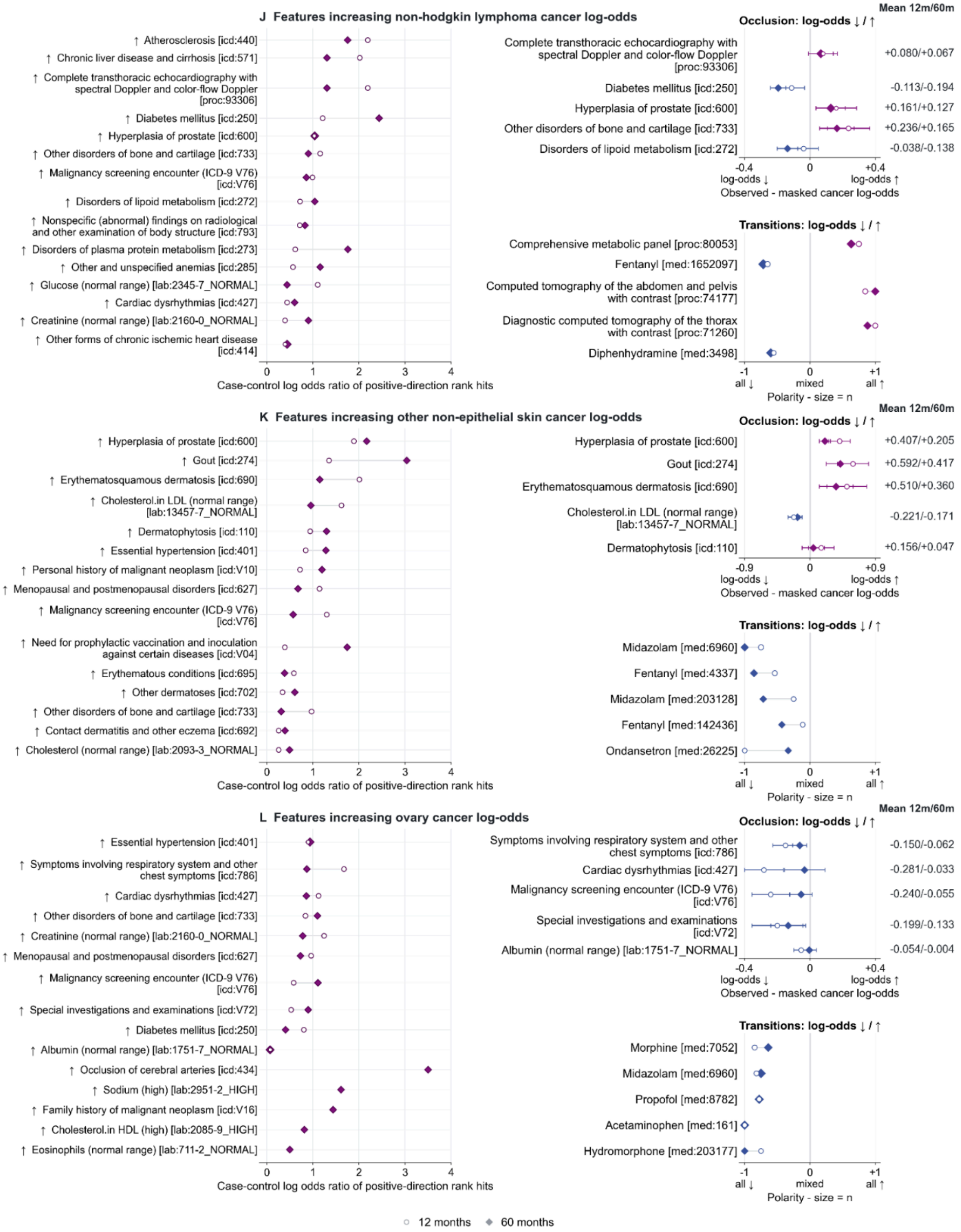

***Figure S13d: Interpretation of features for selected cancer types in MGB cohort (non-Hodgkin lymphoma, other non-epithelial skin cancer, and ovary cancer).*** *(Left) Case-control enrichment among top-ranked positive IG scores, x-axes values are log odds ratio of feature rank hits. (Upper right) Occlusion shows mean predicted log odds with the event observed minus masked, with whiskers of +/-1.96 occurrence level errors, positive means observed feature score is higher. (Lower right) Score-change directions around medication and procedure transitions (-1 all retained changes decrease the score, +1 all increase it).*

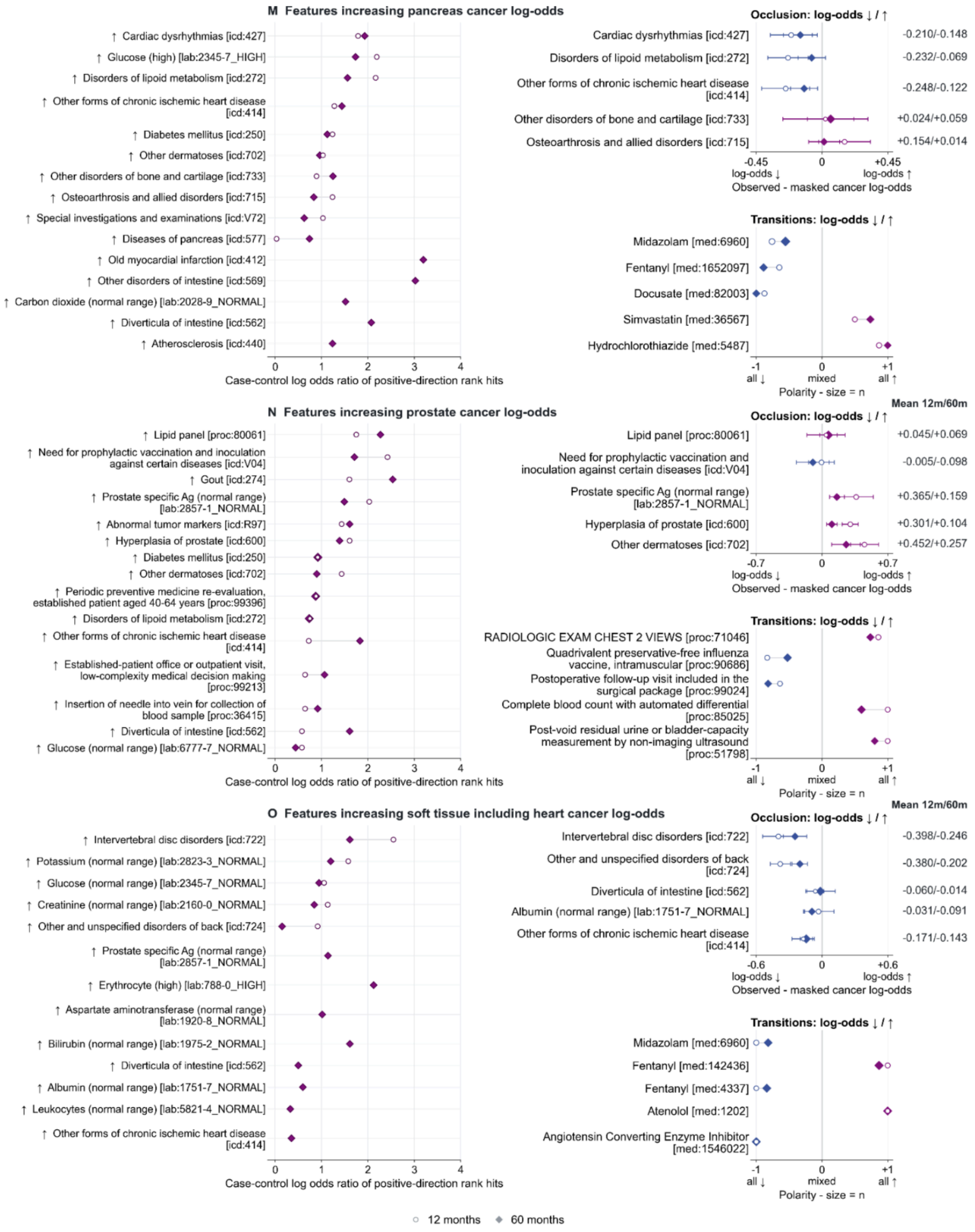


***Figure S13e: Interpretation of features for selected cancer types in MGB cohort (pancreas, prostate, and soft tissue/heart cancer).*** *(Left) Case-control enrichment among top-ranked positive IG scores, x-axes values are log odds ratio of feature rank hits. (Upper right) Occlusion shows mean predicted log odds with the event observed minus masked, with*

*whiskers of +/-1.96 occurrence level errors, positive means observed feature score is higher. (Lower right) Score-change directions around medication and procedure transitions (-1 all retained changes decrease the score, +1 all increase it).*

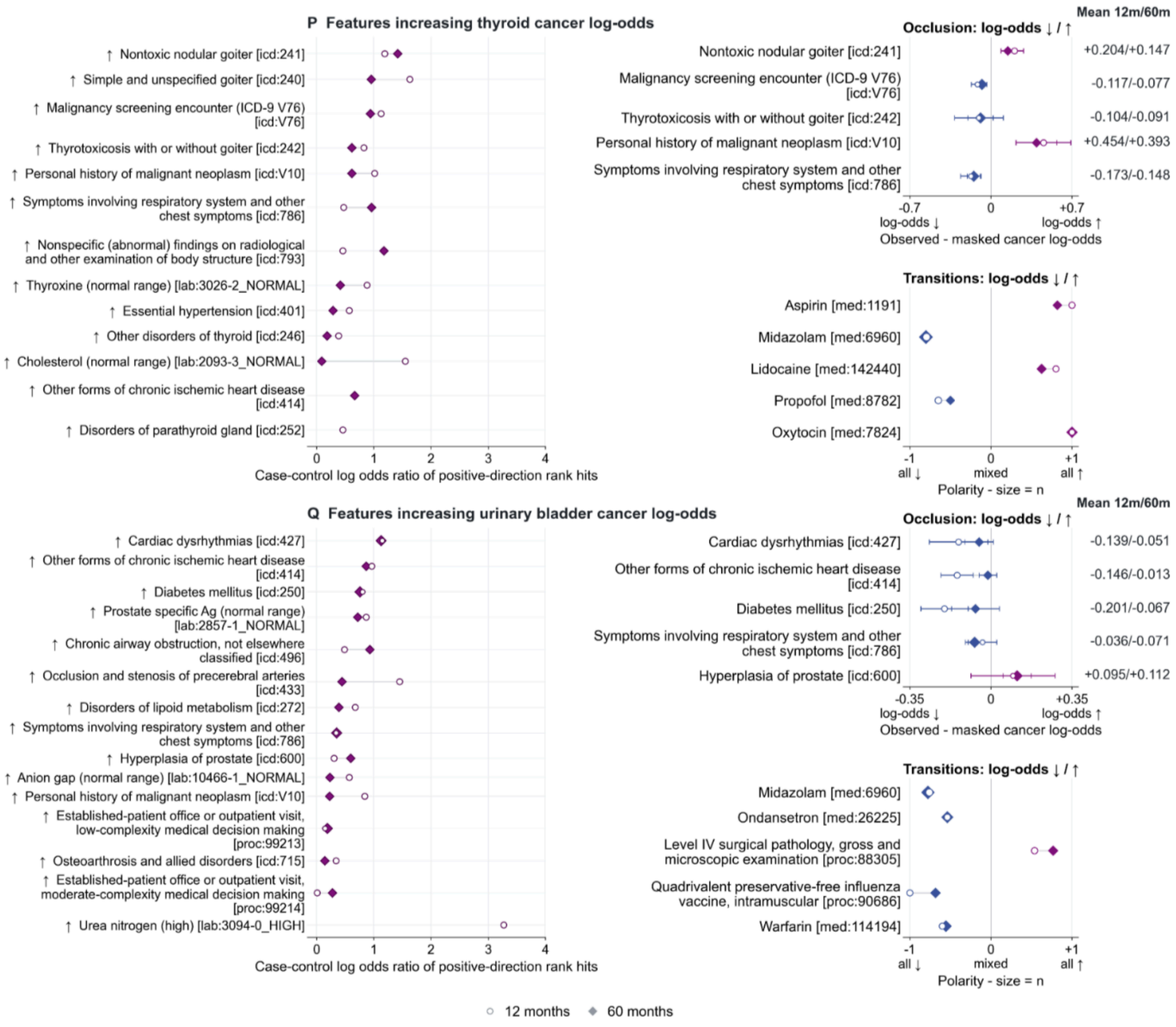


***Figure S13f: Interpretation of features for selected cancer types in MGB cohort (thyroid and urinary-bladder cancer).*** *(Left) Case-control enrichment among top-ranked positive IG scores, x-axes values are log odds ratio of feature rank hits. (Upper right) Occlusion shows mean predicted log odds with the event observed minus masked, with whiskers of +/-1.96 occurrence level errors, positive means observed feature score is higher. (Lower right) Score-change directions around medication and procedure transitions (-1 all retained changes decrease the score, +1 all increase it).*

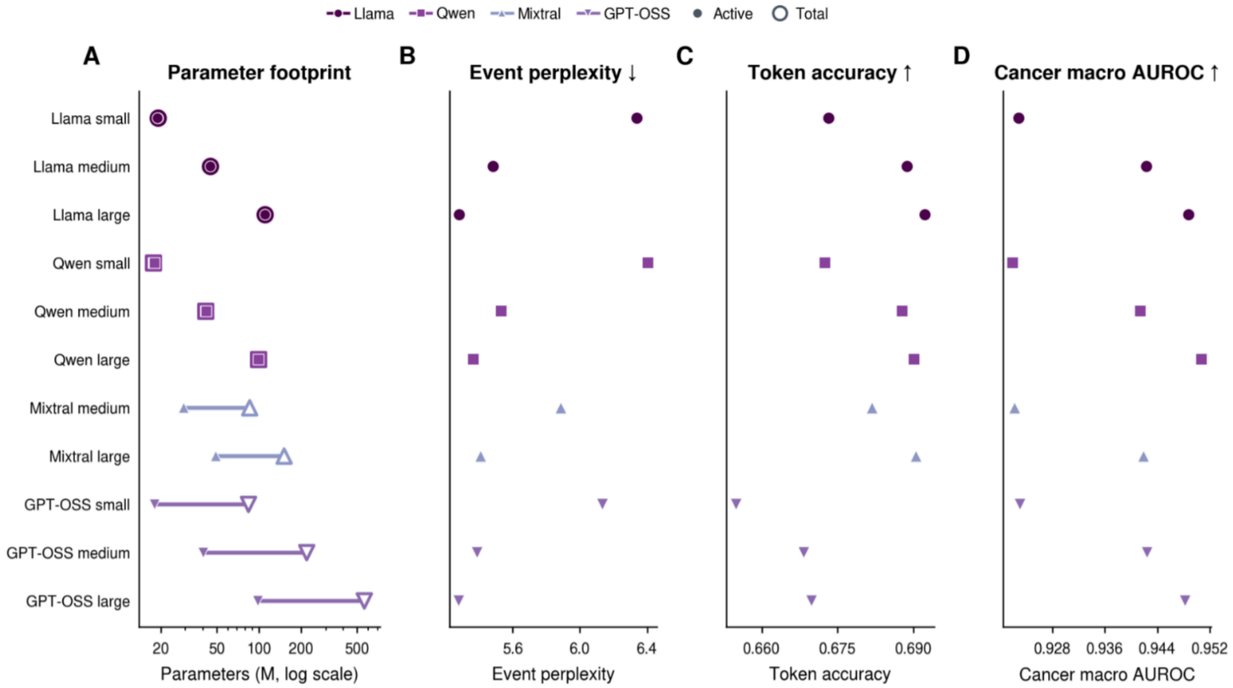


***Figure S14: Scaling model architecture improves downstream predictive performance.*** *Performance of different model sizes and architectures evaluated across tasks. Increasing model size improves performance consistent across both architectures. Llama large achieves the best performance and is used for all other experiments and analysis in this paper. Rows compare sizes of Llama, Qwen, GPT, and Mixtral, identified by circles, squares, and triangles. Due to high-computing we carry this analysis only on the MGB cohort.*

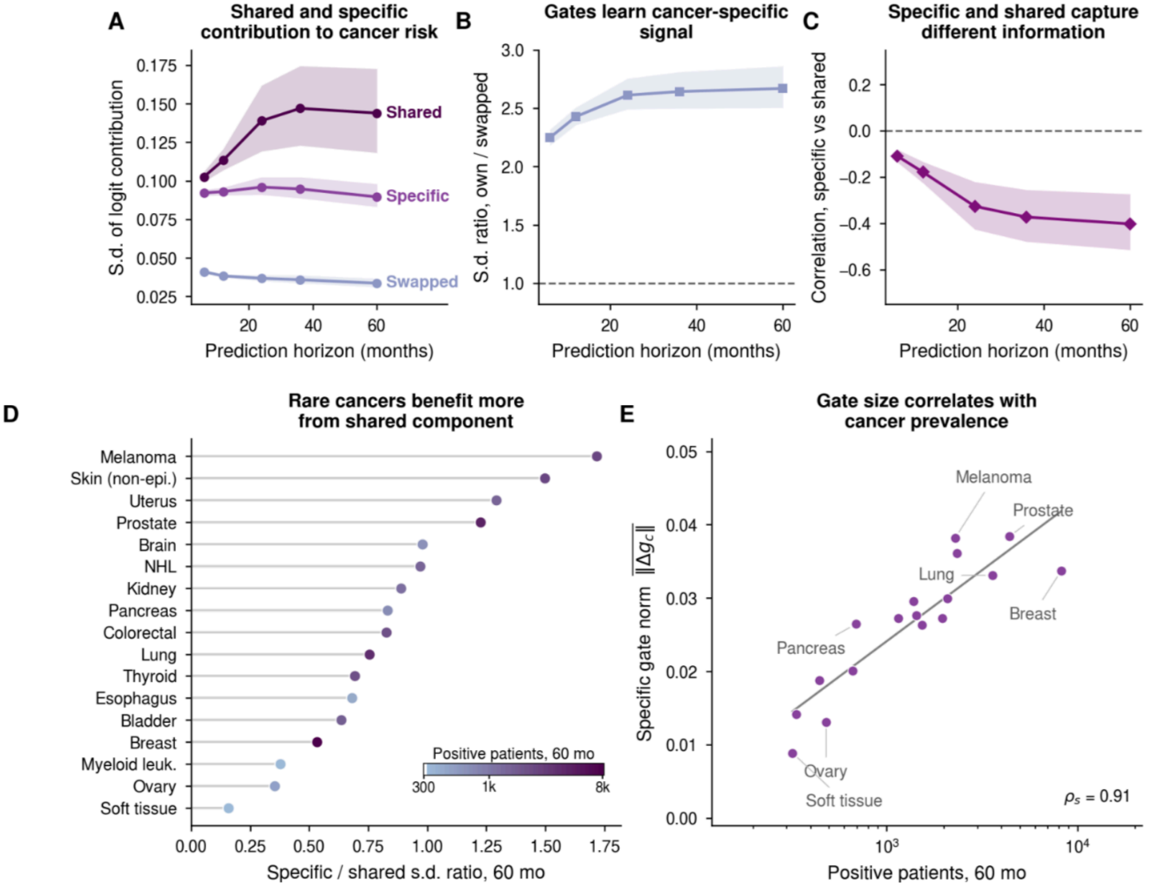


***Figure S15: Contribution of shared and specific components to pan-cancer risk.*** *GenEHR-CancerRisk combines a shared gate architecture with cancer specific deviations to learn stable risk estimates for rare cancers by leveraging from signal in shared baseline with other cancer types. (A) Both specific and shared contribute to risk; random shuffling of cancer gates results in a drop of performance indicating the gate mechanism learns cancer specific features. (B) Relative performance of correct vs swapped gates further validates the specialized role of learned gates. (C) Shared and specific components become more anti-correlated over longer horizons, indicating they capture distinct clinical signals. (D) Breakdown of contribution by cancer types, shared directions provide a larger relative contribution for rare cancers compared to high-prevalence types. (E) Gate-norm drift is correlated with cancer prevalence. This figure summarizes the benefit of decomposing risk as a combination of shared and specific components.*

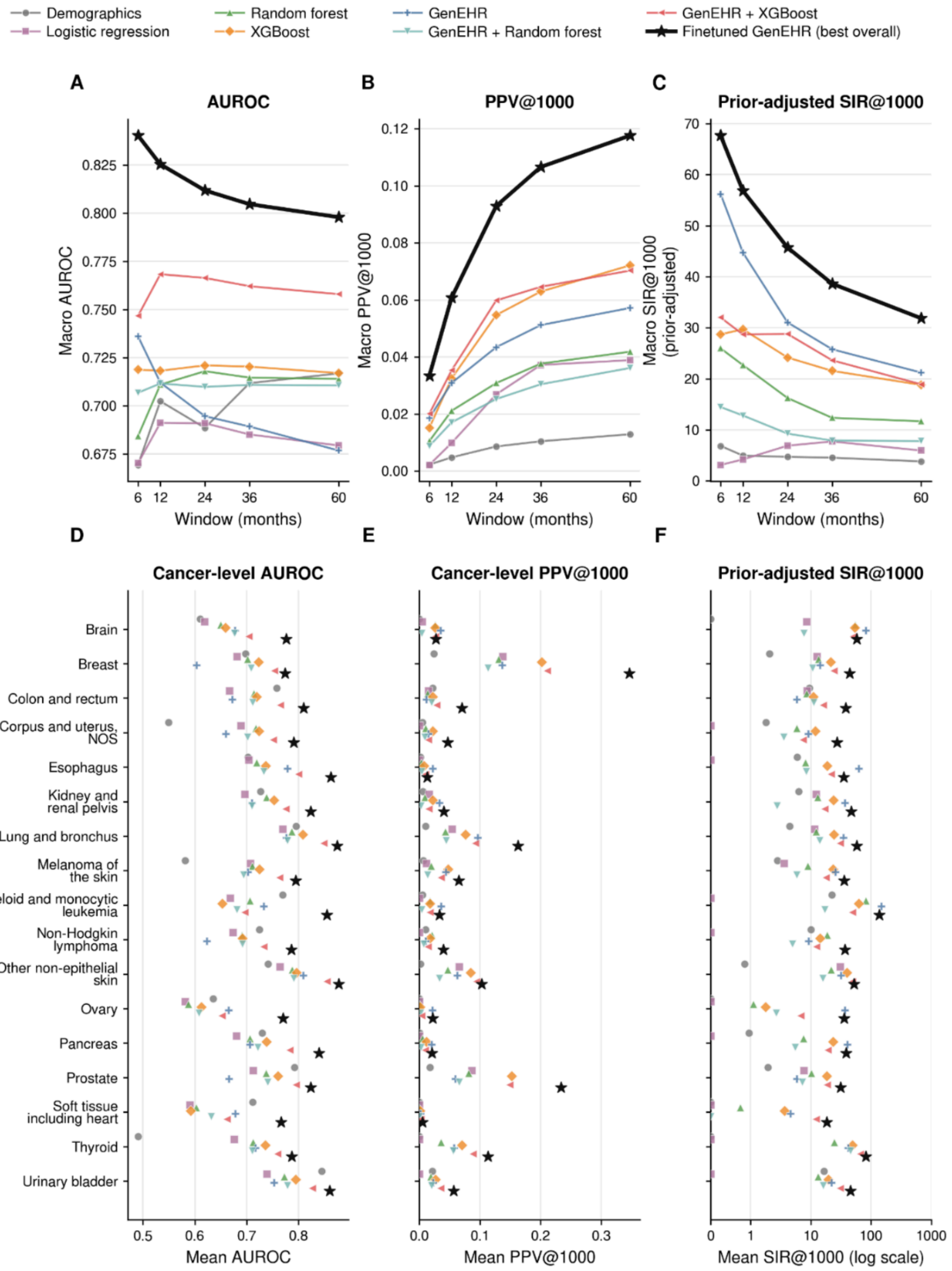


***Figure S16a: Pan-cancer baseline comparisons.*** *(A-C) Overall performance in terms of AUROC, PPV, and SIR across 17 cancers at six prediction horizons (6, 12, 24, 36, and 60 months). PPV and SIR are reported at operating threshold N=1000. Comparison is done with a*

*demographic-only baseline using a logistic model; bag-of-words features (5479 features) constructed using training data after trajectory sampling (5479 total features) with logistic regression, random forest and XGBoost. Fine-tuning consistently outperforms all baselines across metrics.*

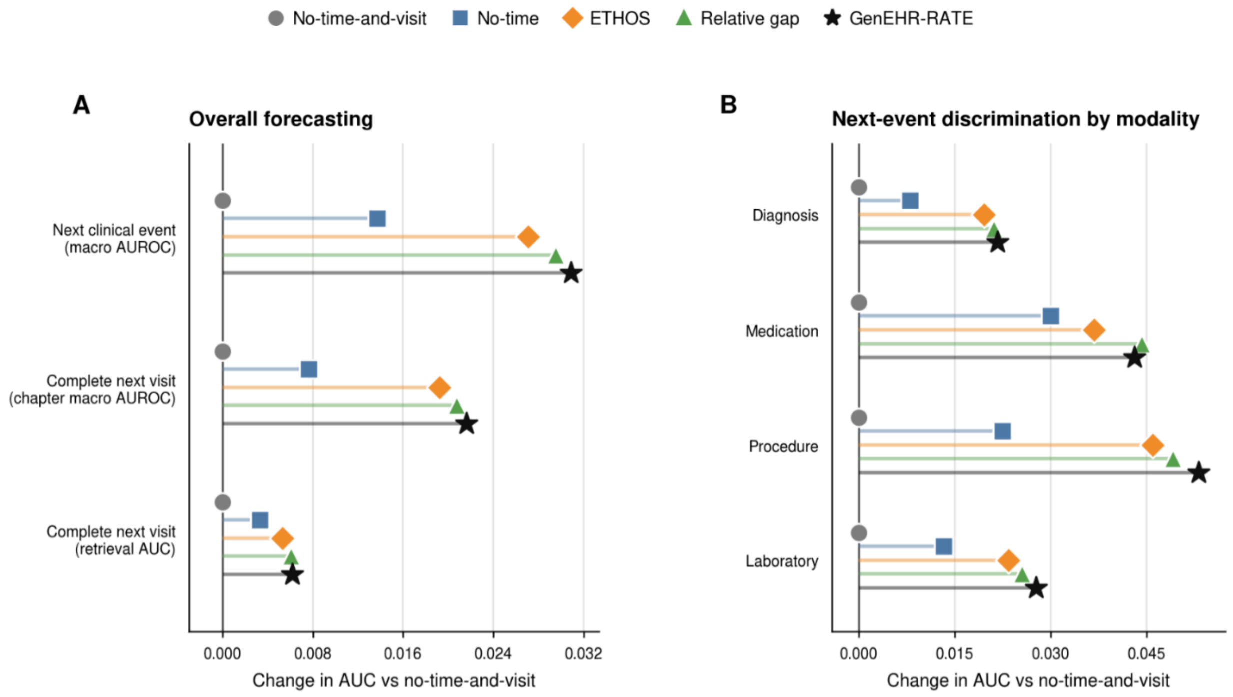


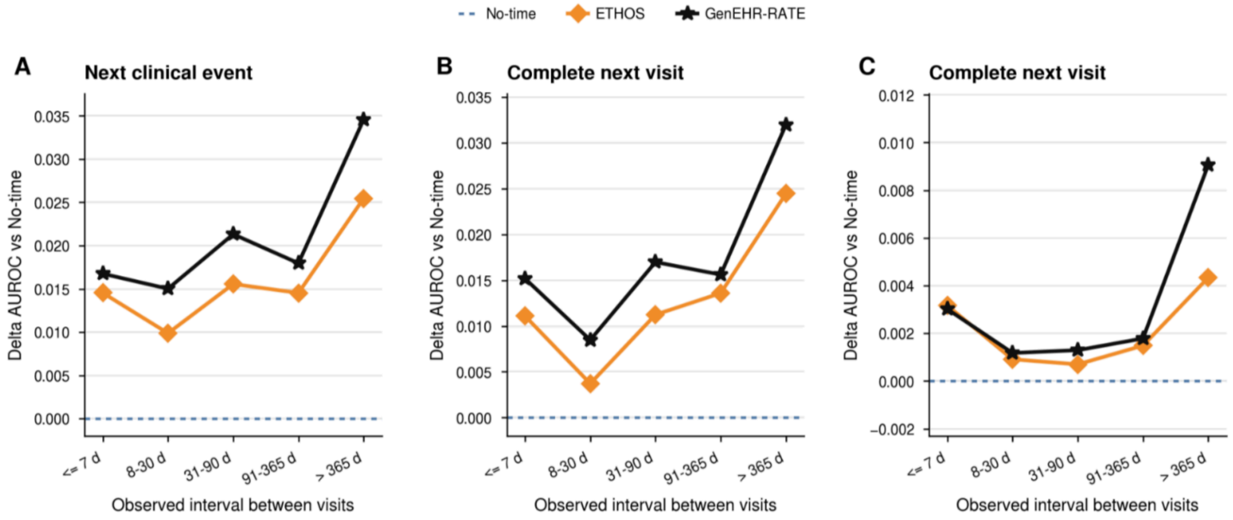


***Figure S16b: Ablation of time tokenization.*** *Comparison of different time tokenization approaches. No-time-and-visit (model trained on patient trajectories without any visit or time tokens in trajectory), No-time (model trained on patient trajectories without any time tokens), ETHOS (using a pre-specified set of gap tokens introduced introduced in ETHOS), relative gap (one token for each day with maximum ten years of gap), and RATE (our mixed radix encoding) Top: (A-B) Overall improvement in forecasting performance over no-time-and-visit in terms of*

*AUROC. Bottom (A-C) Forecasting performance at increasing time gaps. All comparisons are done on models trained with the same size, training budget, and hyperparameters.*

### Providence Health and Services Results.

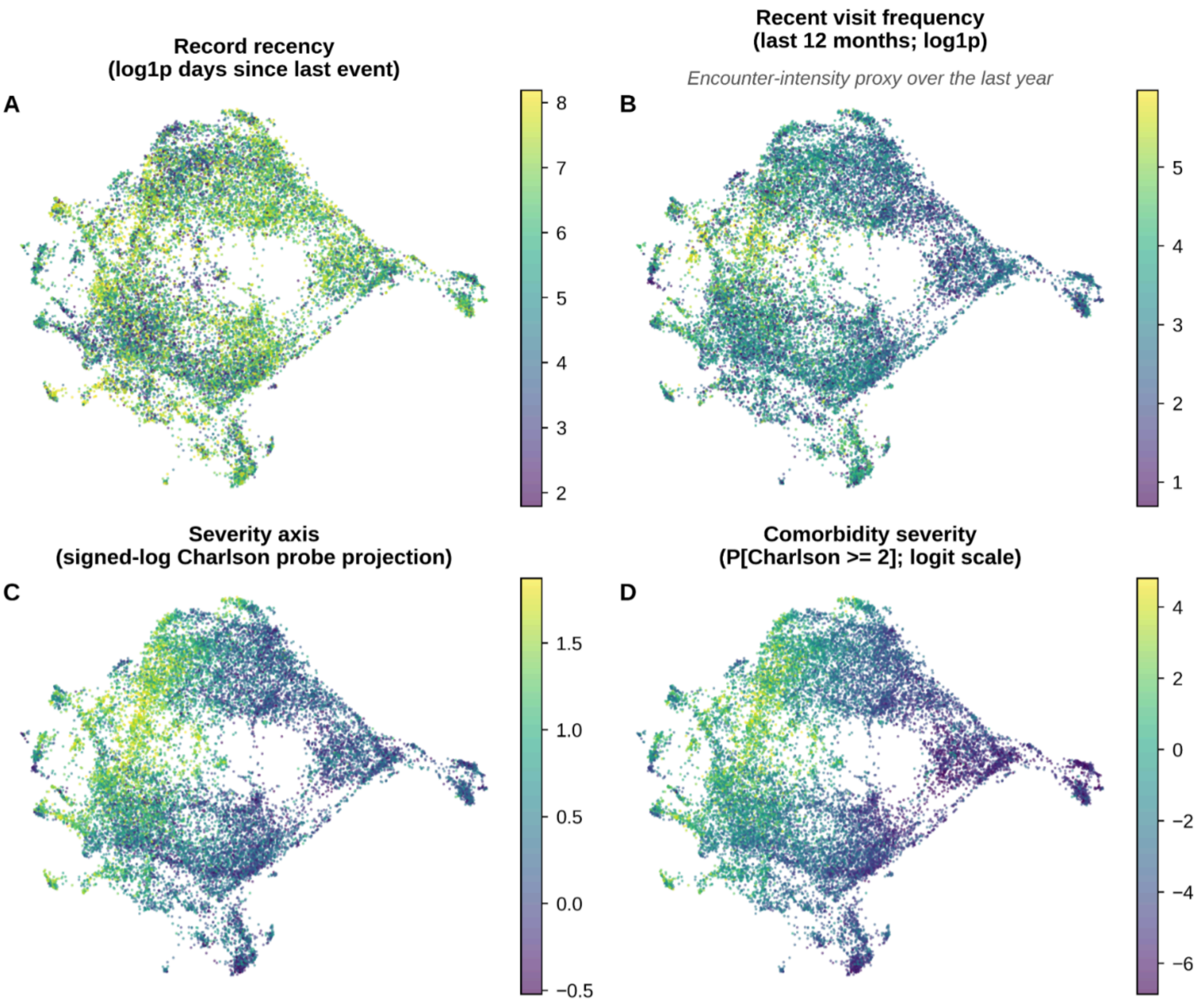


***Figure S17: Patient health state representation given by pooled encoding of all events in patient's trajectory (PROV cohort).*** *All four panels are two dimensional projections of patient trajectory representation obtained from the penultimate layer of GenEHR. Each has a different overlay: (A) time gap since last visit (B) number of visits in preceding year as a proxy for acuity. (C) a linear probe was fitted on feature representation for predicting CCI overlay with probe prediction (D) only patients who have CCI above chosen threshold. Gradients in CCI imply that representations capture information about overall morbidity and healthcare use.*

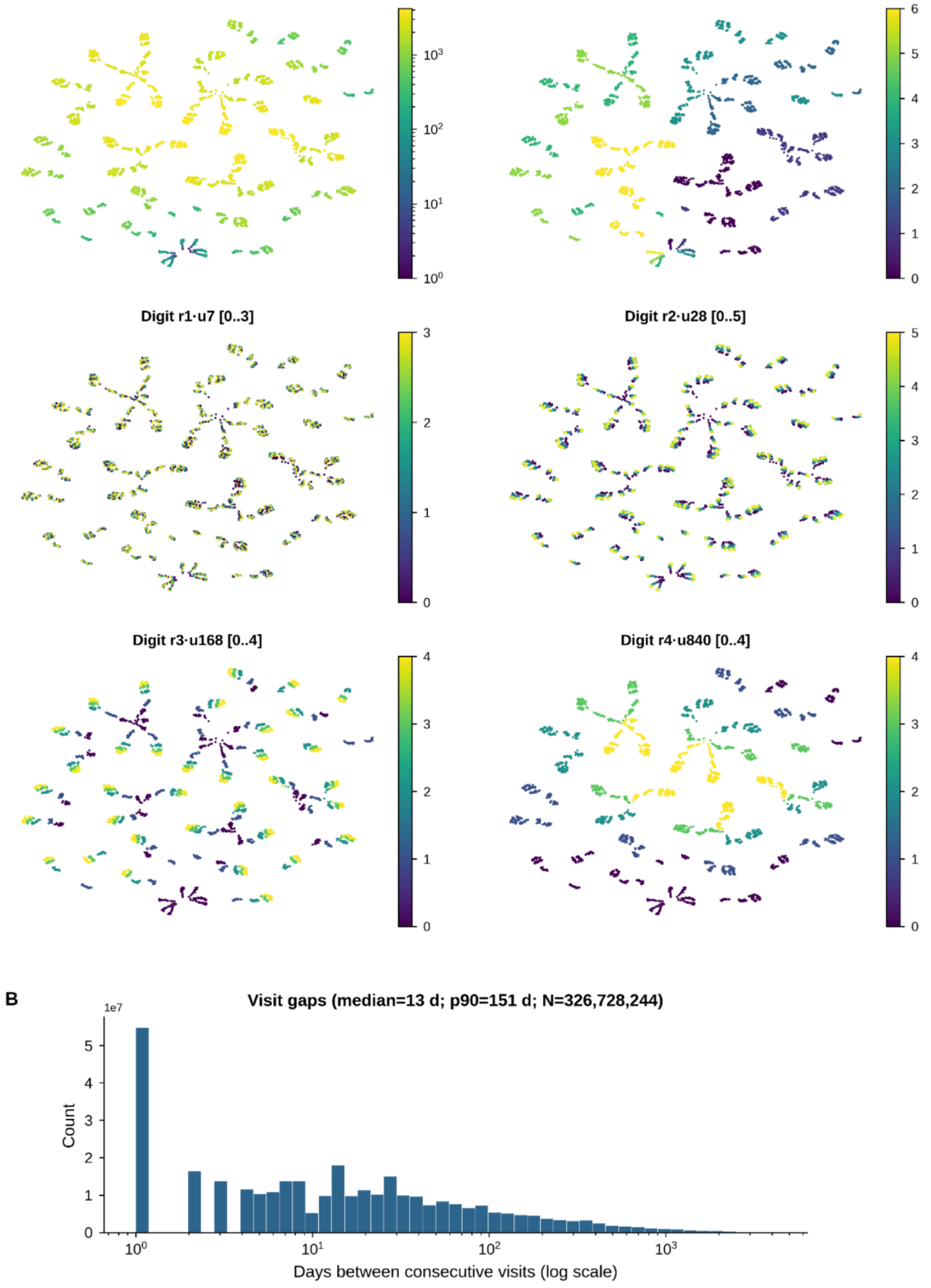


***Figure S18: Learned RATE representation and observed time gaps in PROV cohort.***
*UMAP projection of learned representation of 4200 days, colored by the value of each radix digit together with the observed distribution of inter-visit gaps. (A) Each point represents one encoded gap from 1 to 4200 days where color is the gap length. The other five show the digit with place values 1, 7, 28, 168, and 840 days with color indicating lower to higher values on each scale. (B) Inter-visit time interval distribution.*

**Event representations capture clinical structure and a context-dependent model-belief graph for hypothesis generation.**

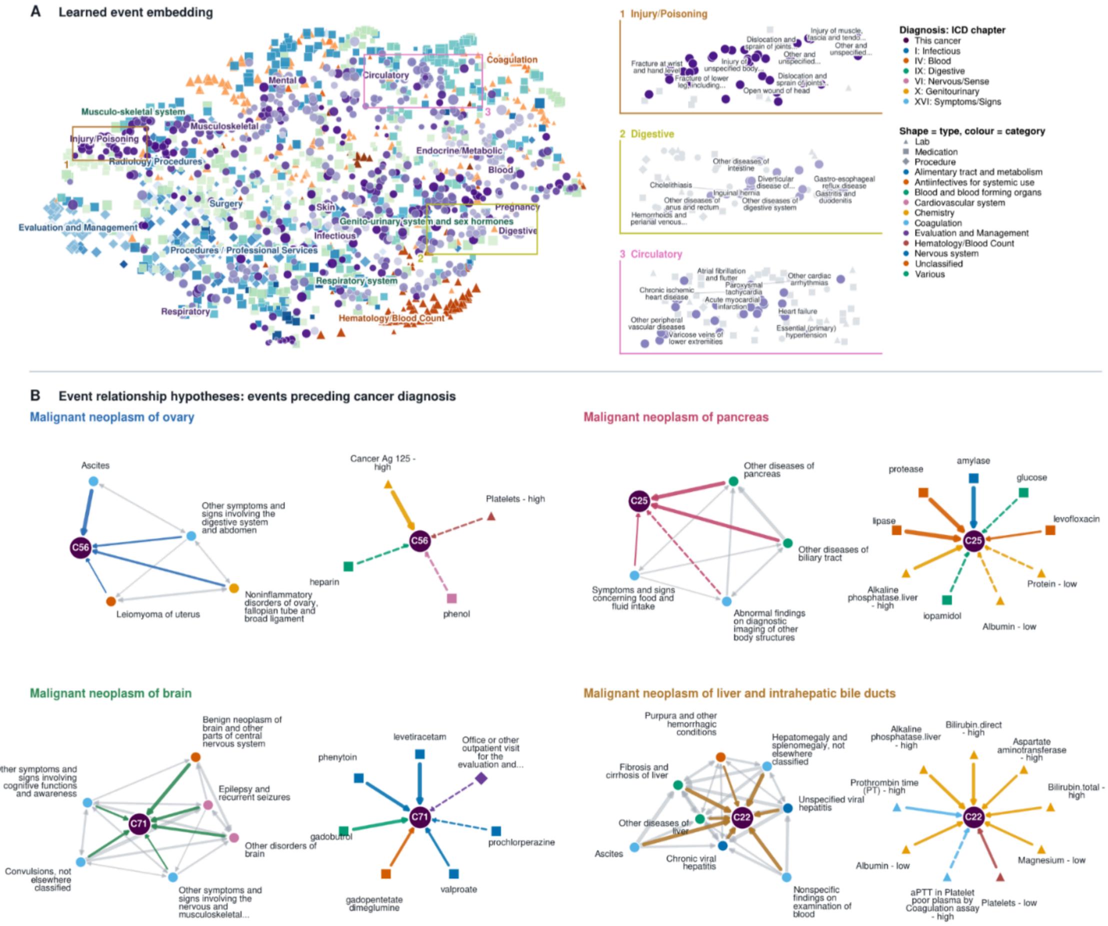


**Patient representation over multiple visits captures comorbidity burden.**

**Patient map (end-of-trajectory) with co-morbidity gradient**

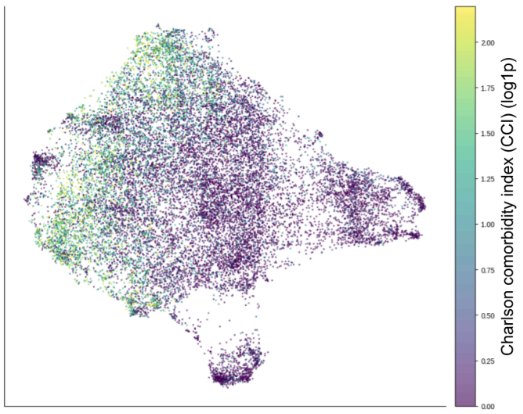


**Patient health state progression over visits**

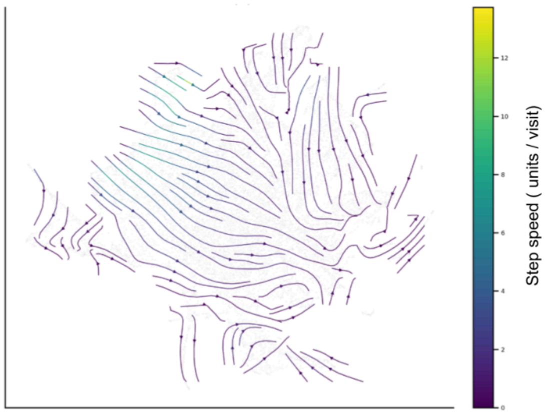


***Figure S19: For the PROV cohort.*** *Left:* ***(A)*** *UMAP projection of learned event embeddings using cosine distance between embedding vectors. Right: closeup of three regions numbered 1*

*to 3 shows GenEHR learns hierarchy of clinical events without explicit prior knowledge and without using text.* ***(B)*** *Event relationship preceding cancer diagnosis inferred using cPMI between pairs of events for selected cancers (ICD codes C56, C25, C71, C22). For held-out patients whose preceding visit contains an event* $e_1$*, cPMI compares the average model probability assigned to* $e_2$ *normalized against the model averaged probability of* $e_2$ *across all positions and patients. In the cPMI belief graph nodes are clinical events and edges are informational links. Left: Diagnosis neighborhood of cancer with edges among preceding events, Right: Cross-modality hub - medications, procedures, and labs preceding cancer diagnosis. Dashed edges: events with weaker mutual information scores that are plausibly less specific to that cancer. (****Bottom)*** *UMAP neighborhoods of patients using their end-of-trajectory <eos> representation reflect gradient of comorbidity burden (left), and transitions along trajectories are informative about how the learned state of patients evolves over time using their <visit> representation over multiple visits. Flow (arrows) indicates how patient health evolves over visits (right) averaged over groups of held-out patients.*

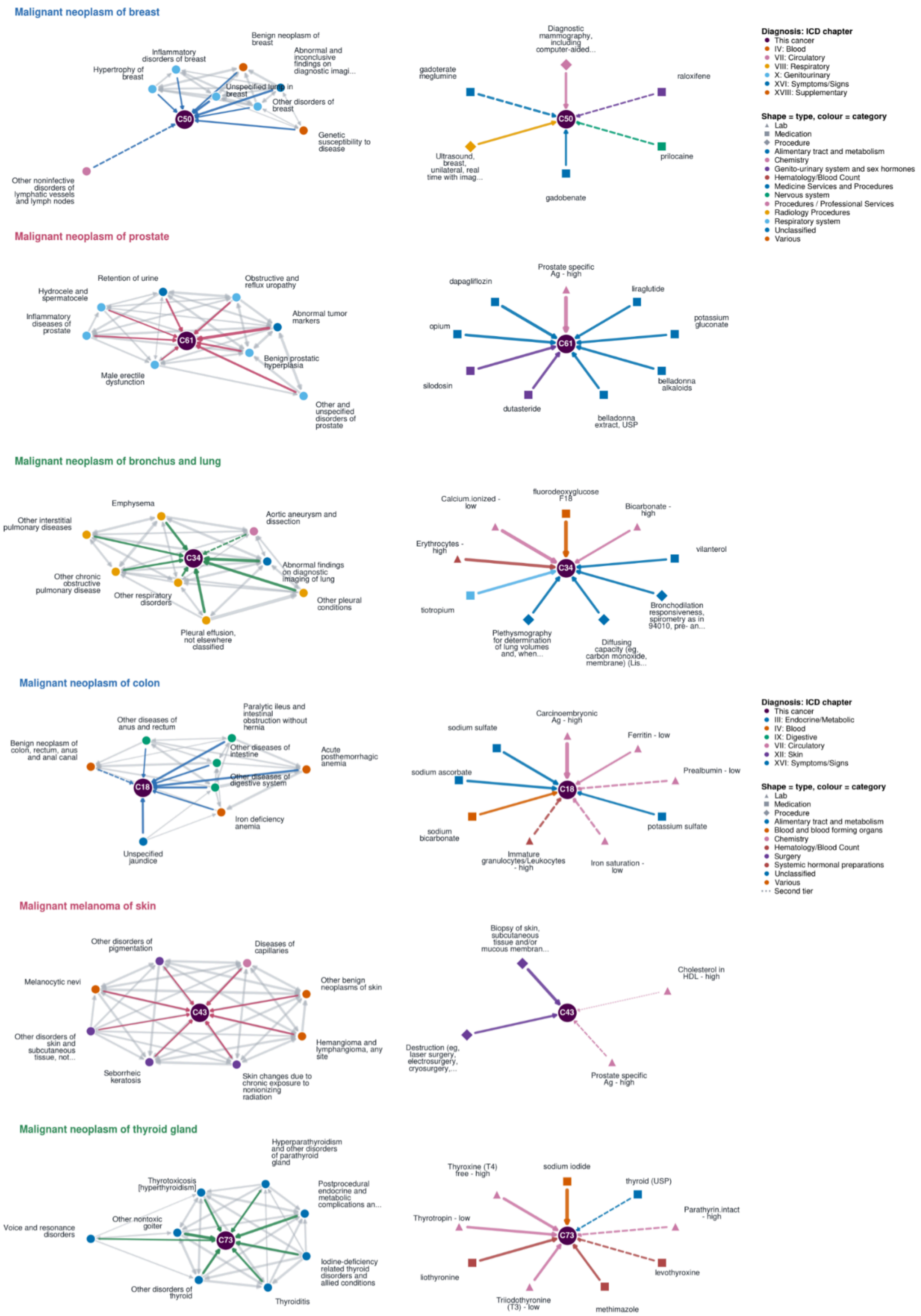


***Figure S20a: Breakdown of within and across modality belief graphs across cancer types (PROV Cohort).*** *Event relationship preceding cancer diagnosis inferred using cPMI between pairs of events for selected cancers (ICD codes C50, C61, C18, C53, C73). For held-out patients whose preceding visit contains an event* $e_1$*, cPMI compares the average model probability assigned to* $e_2$ *normalized against the model averaged probability of* $e_2$ *across all*

*positions and patients. In the cPMI belief graph nodes are clinical events and edges are informational links. Left: Diagnosis neighborhood of cancer with edges among preceding events, Right: Cross-modality hub - medications, procedures, and labs preceding cancer diagnosis. Dashed edges: events with weaker mutual information scores that are plausibly less specific to that cancer.*

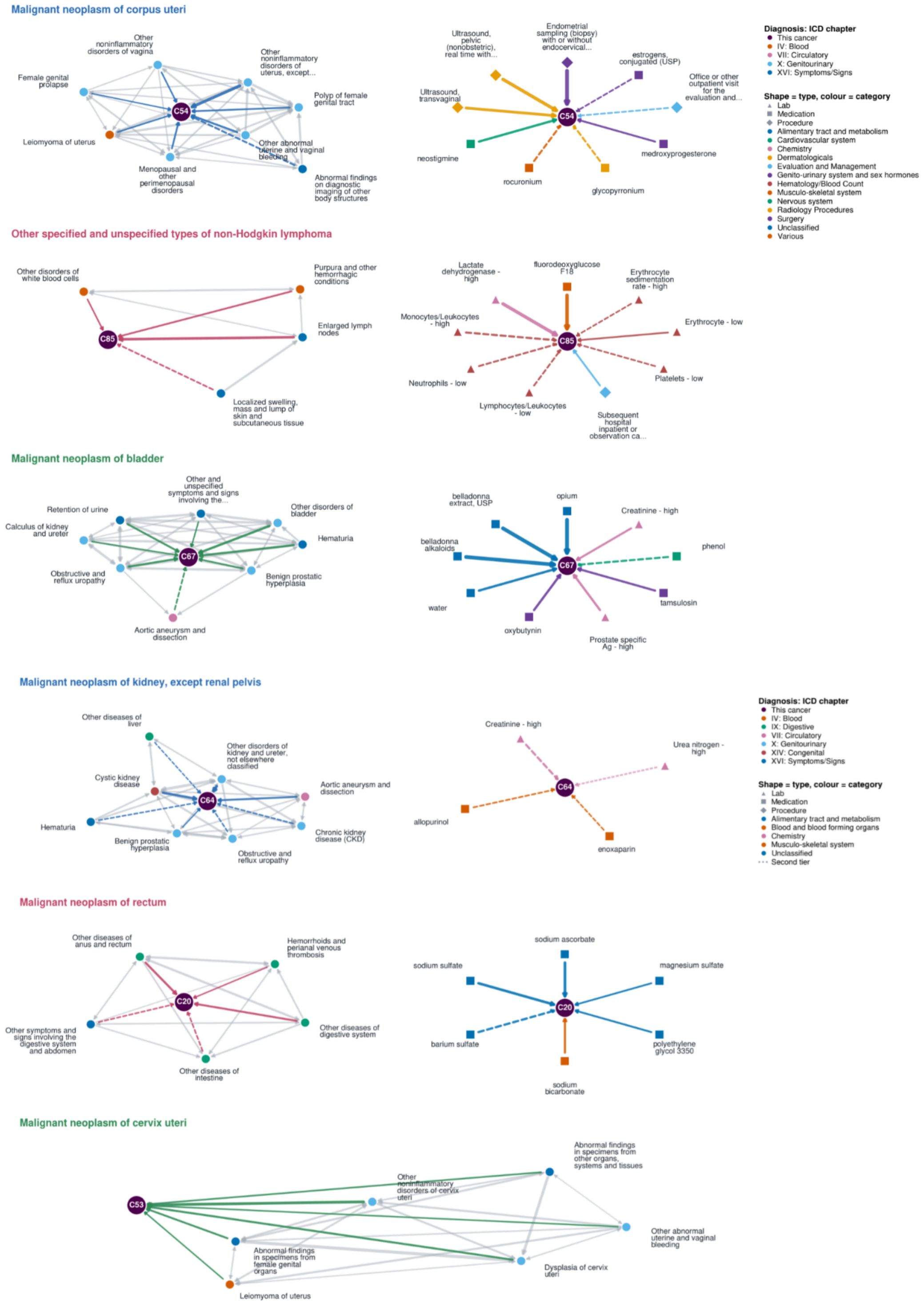

***Figure S20b: Breakdown of within and across modality belief graphs across cancer types (PROV cohort).*** *Event relationship preceding cancer diagnosis inferred using cPMI between pairs of events for selected cancers (ICD codes C54, C85, C67, C64, C20, C53). For held-out patients whose preceding visit contains an event* $e_1$*, cPMI compares the average model probability assigned to* $e_2$ *normalized against the model averaged probability of* $e_2$ *across all positions and patients. In the cPMI belief graph nodes are clinical events and edges are informational links. Left: Diagnosis neighborhood of cancer with edges among preceding events, Right: Cross-modality hub - medications, procedures, and labs preceding cancer diagnosis. Dashed edges: events with weaker mutual information scores that are plausibly less specific to that cancer.*

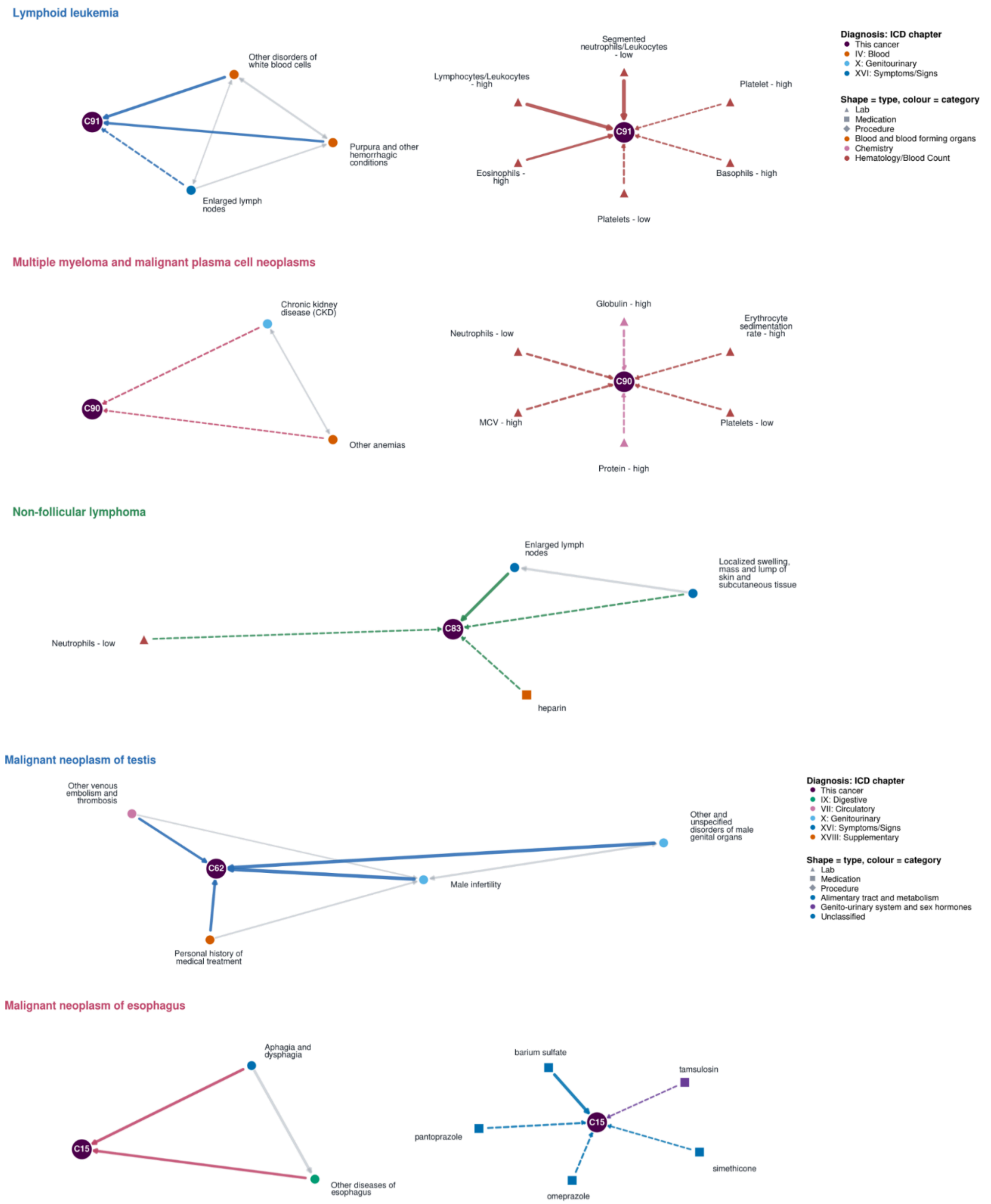


***Figure S20c: Breakdown of within and across modality belief graphs across cancer types (PROV cohort).*** *Event relationship preceding cancer diagnosis inferred using cPMI between pairs of events for selected cancers (ICD codes C91, C90, C83, C62, C15). For held-out patients whose preceding visit contains an event* $e_1$*, cPMI compares the average model probability assigned to* $e_2$ *normalized against the model averaged probability of* $e_2$ *across all*

*positions and patients. In the cPMI belief graph nodes are clinical events and edges are informational links. Left: Diagnosis neighborhood of cancer with edges among preceding events, Right: Cross-modality hub - medications, procedures, and labs preceding cancer diagnosis. Dashed edges: events with weaker mutual information scores that are plausibly less specific to that cancer.*

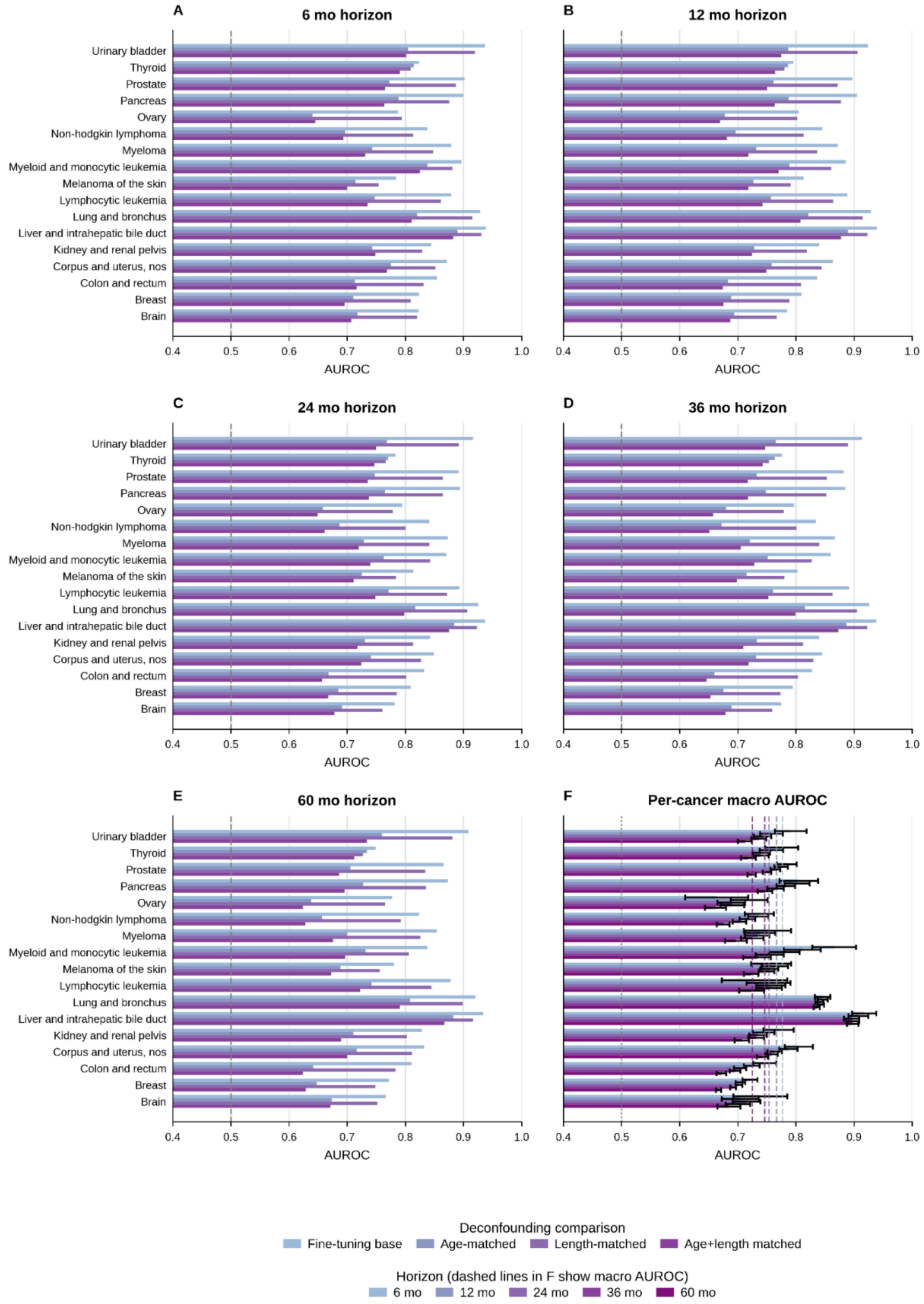


***Figure S21a: Sensitivity of PROV cancer discrimination to age and trajectory length on cancer risk.*** *Cases and controls were matched by age bracket at the endpoint of trajectory as*

*well as with an equal number of events. Age matched results in a drop of performance, but overall performance shows the model just doesn't rely on age and learns from the two combined. Length matched shows the results are not sensitive to the number of codes in patient trajectory. (A-E) are results for 6, 12, 24, 36, and 60 month horizons, with each row comparing original evaluation with age matching, event count matching, and both combined.*

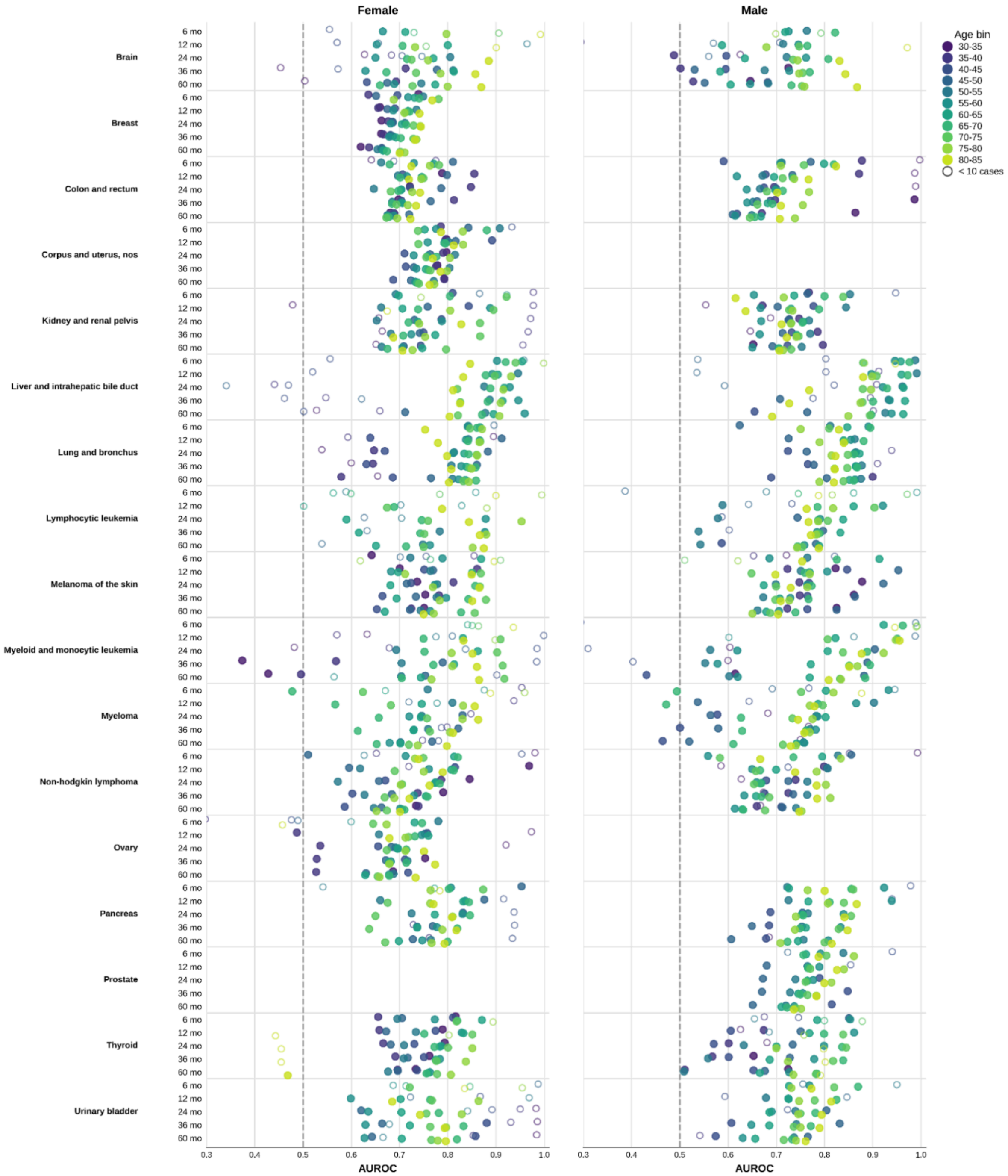


***Figure S21b: (PROV Cohort) Breakdown of predictive performance by age-brackets and sex.*** *Performance by age and sex bracket for different groups assessing heterogeneity and data sparsity. (Left) Performance on female population for each cancer (Right) Performance on male*

*population for each cancer. Both are reported for all six (6, 12, 24, 36, and 60 months) prediction horizons. Color encodes five year age groups, and hollow circle means fewer than 10 positive outcomes in each bin.*

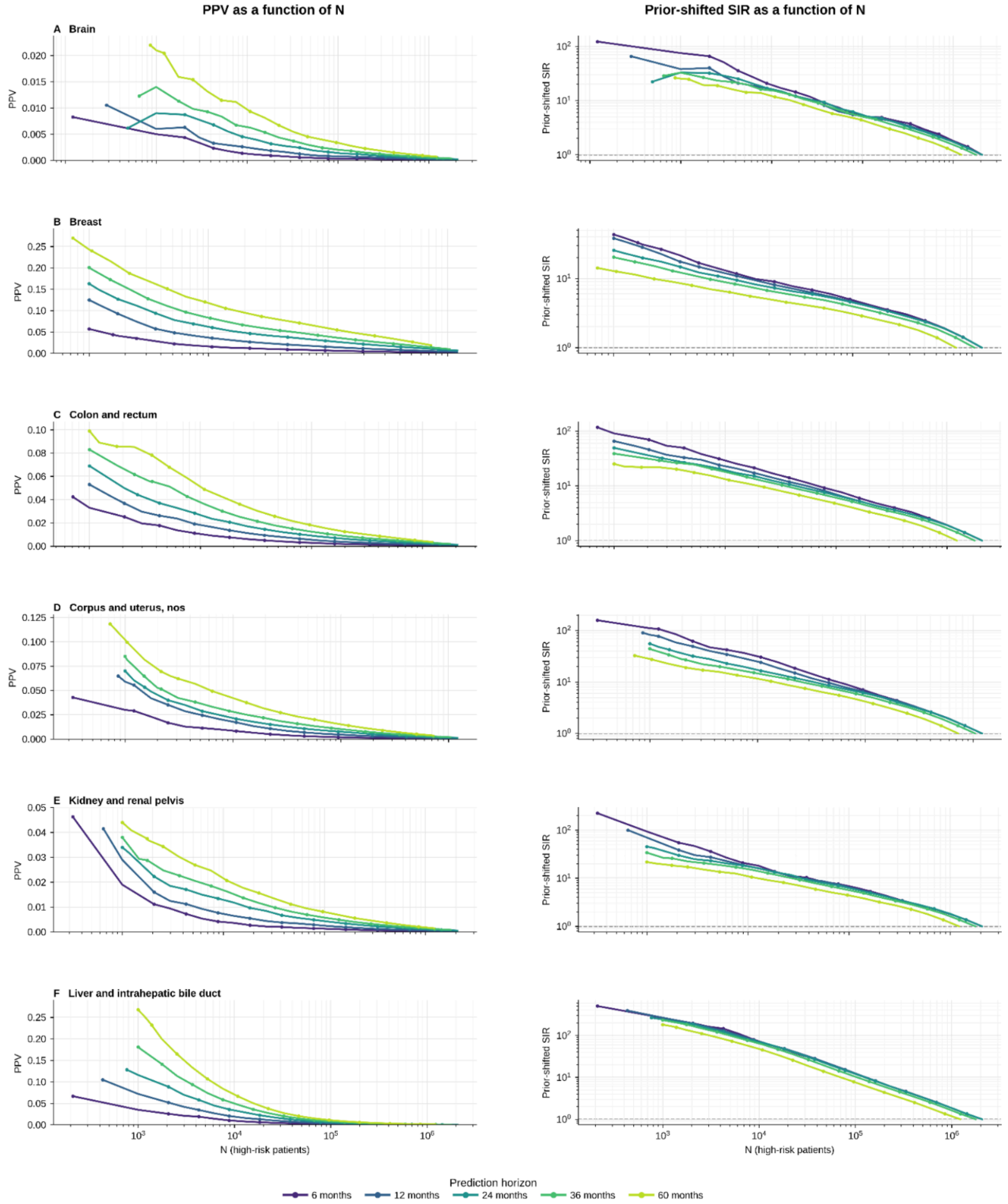

***Figure S22a: (PROV Cohort) Clinical utility of GenEHR fine-tuned model for nominating patients for screening by choosing an operating threshold****. PPV (left) and SIR (right) plotted as a function of operating threshold N shows a net benefit of detecting one cancer at the cost of increasing false positives. (A-F) brain, breast, colon/rectum, corpus/uterus, esophagus, and kidney/renal-pelvis cancer.*

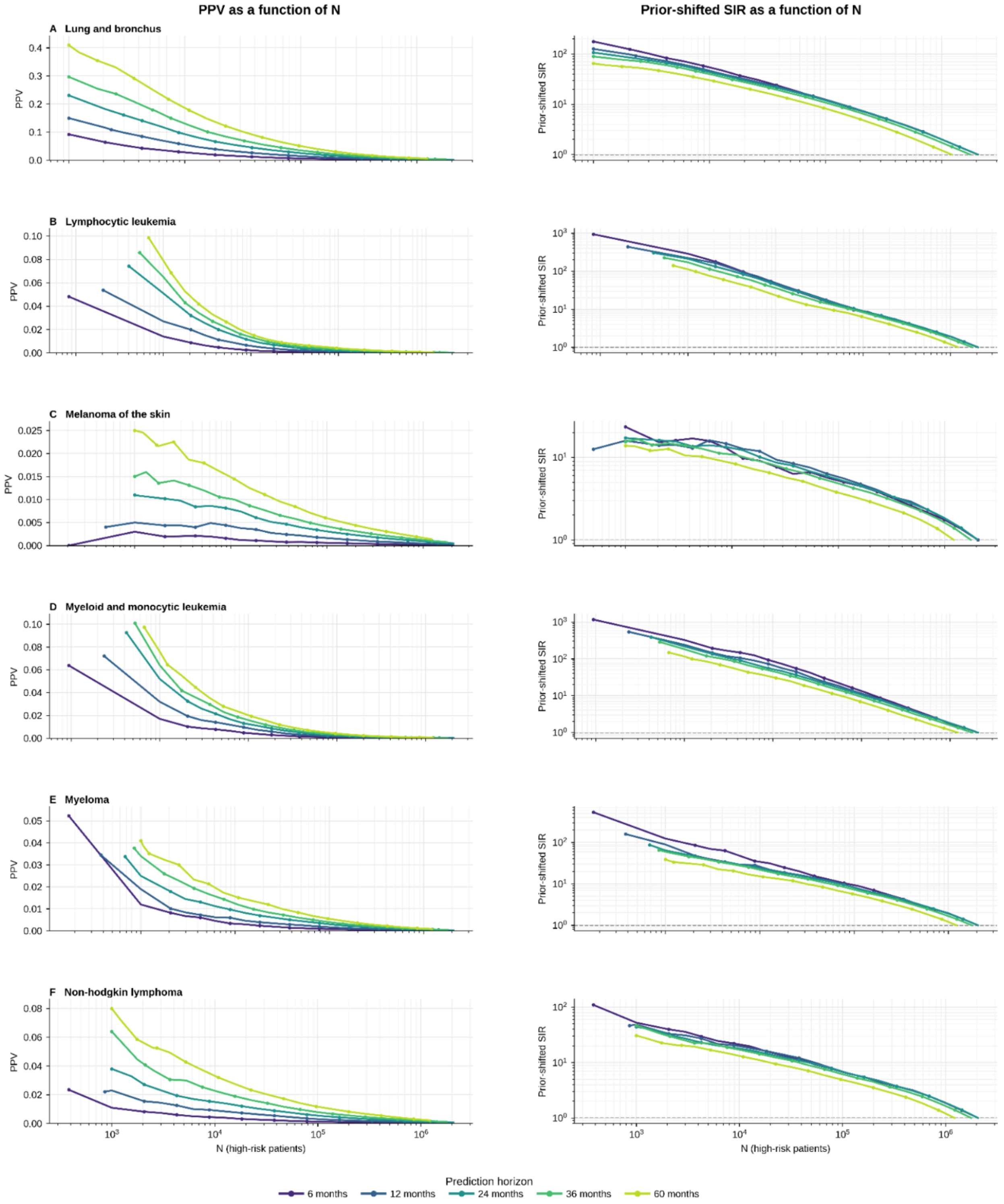

***Figure S22b: (PROV Cohort) Clinical utility of GenEHR fine-tuned model for nominating patients for screening by choosing an operating threshold***. *PPV (left) and SIR (right) plotted as a function of operating threshold N shows a net benefit of detecting one cancer at the cost of increasing false positives. (A-F) brain, breast, colon/rectum, corpus/uterus, esophagus, and kidney/renal-pelvis cancer.*

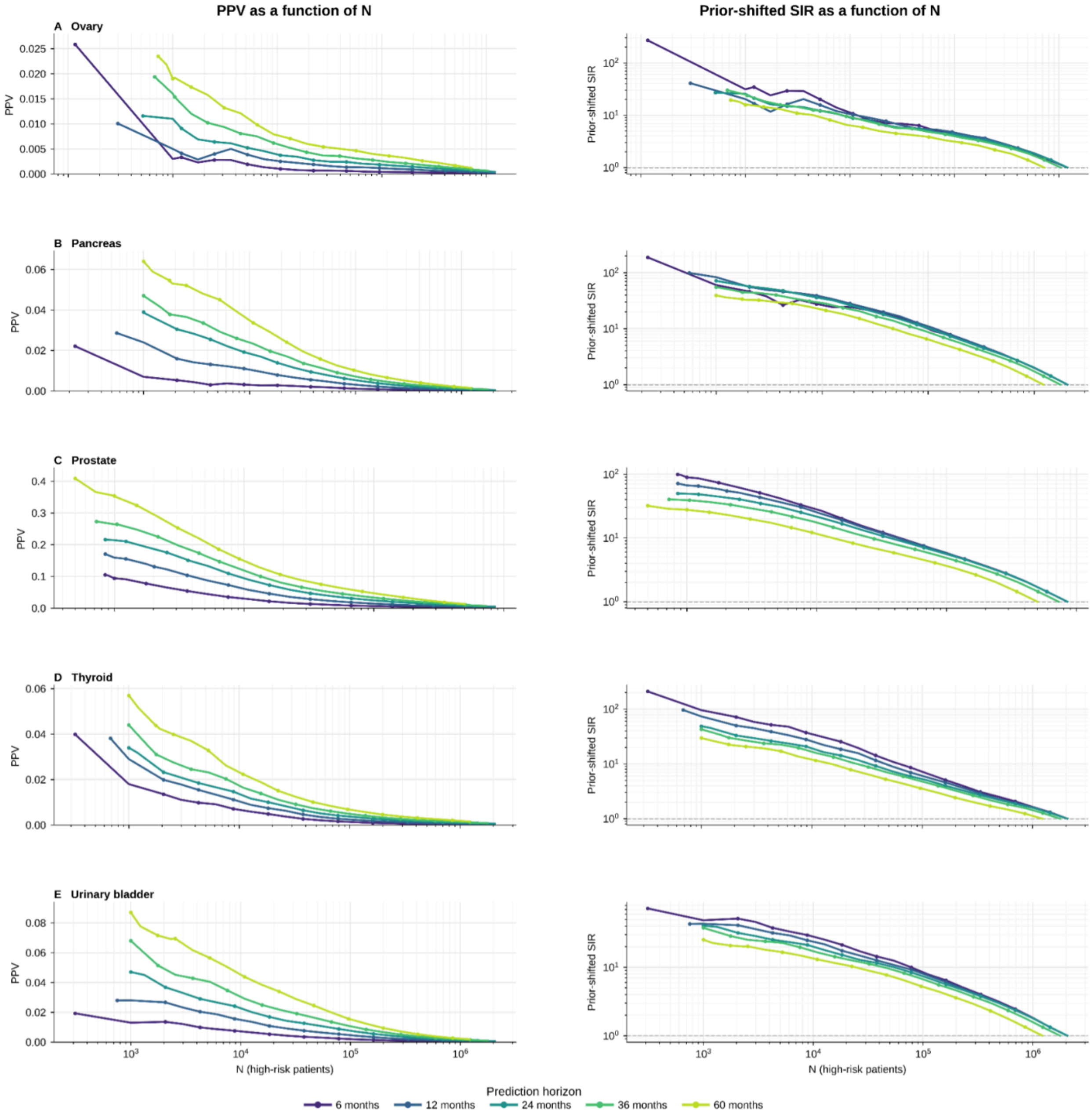


***Figure S22c: (PROV Cohort) Clinical utility of GenEHR fine-tuned model for nominating patients for screening by choosing an operating threshold***. *PPV (left) and SIR (right) plotted as a function of operating threshold N shows a net benefit of detecting one cancer at the*

*cost of increasing false positives. (A-F) brain, breast, colon/rectum, corpus/uterus, esophagus, and kidney/renal-pelvis cancer.*

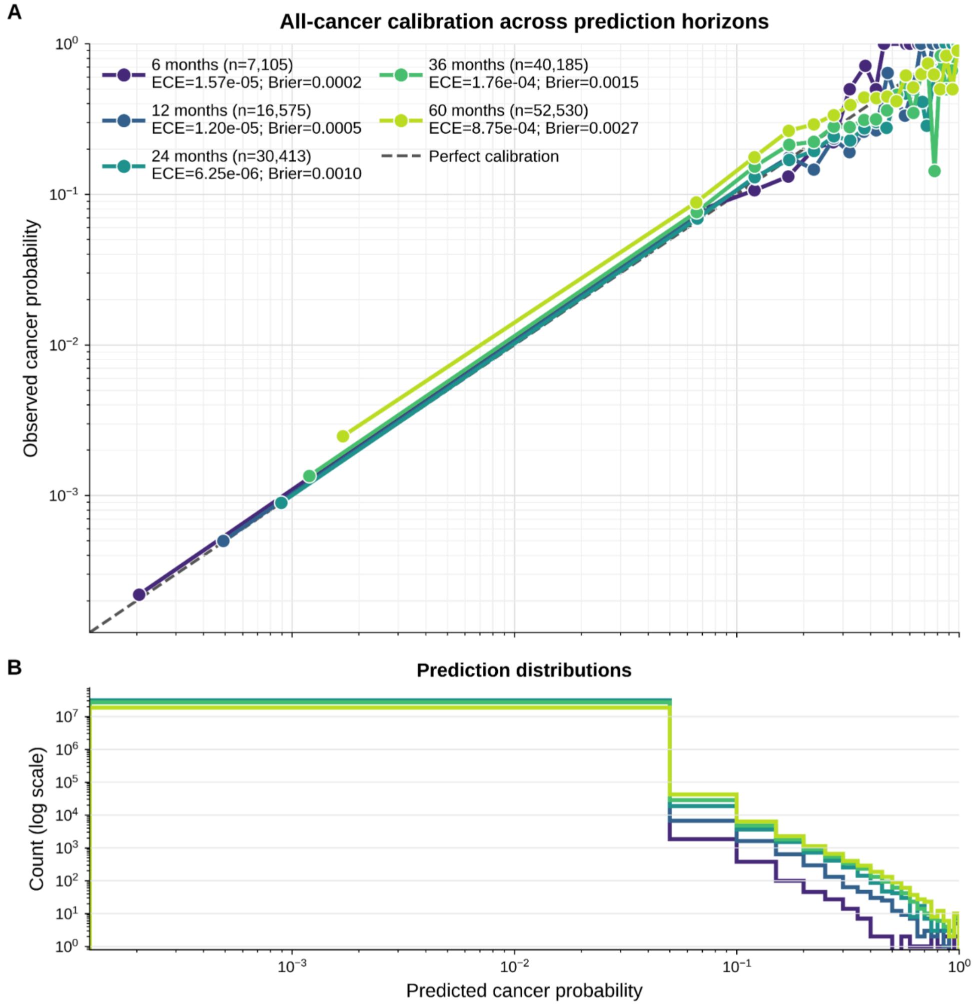


***Figure S23: (PROV Cohort) Pan-cancer risk model is well calibrated across horizons and cancer types.*** *Observed cancer frequency against predicted probability across calibration bins for different horizons, where diagonal line indicates perfect calibration. Both ECE and Brier score describe fine-tuned cancer-risk probabilities (not next event calibration in Fig 3).*

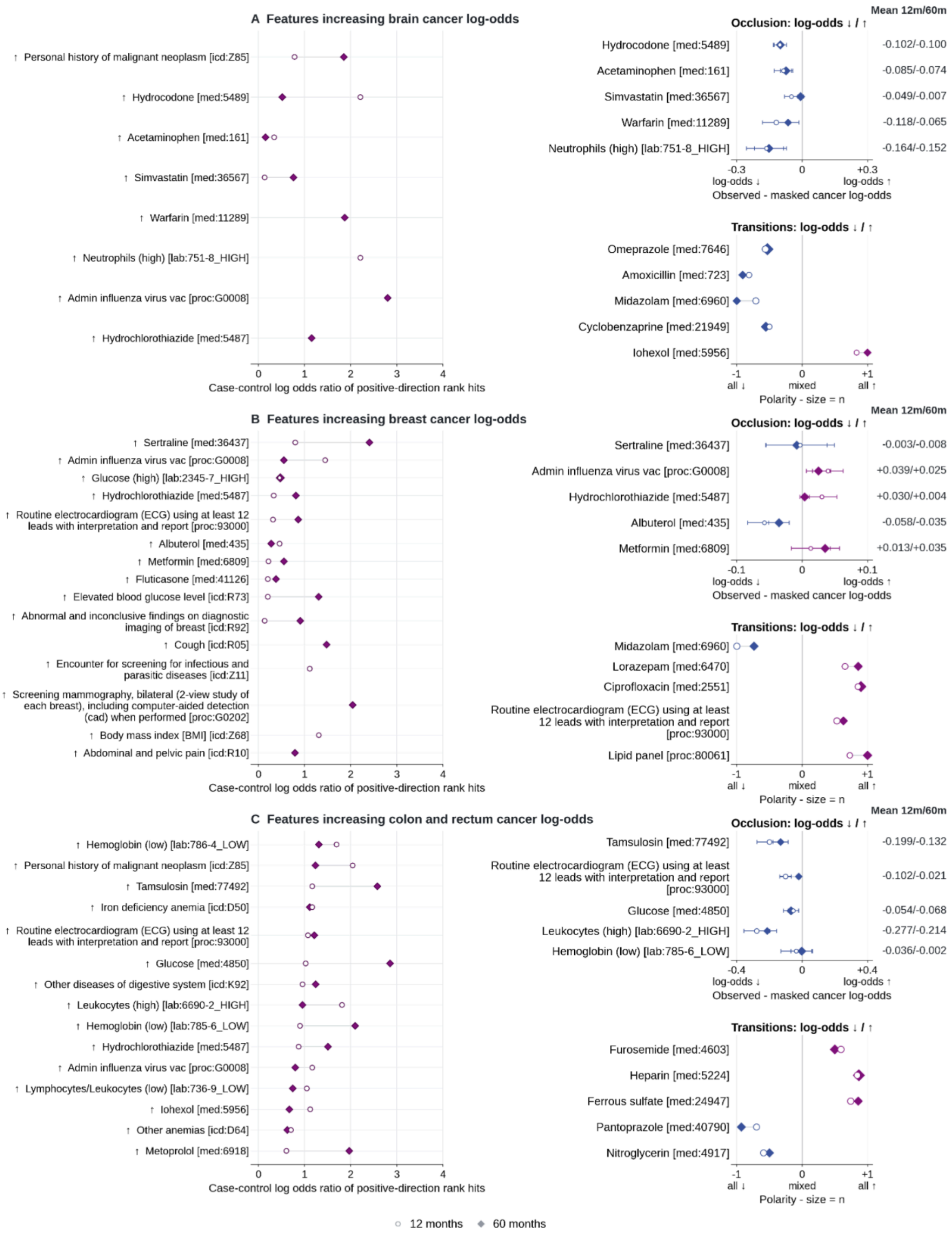


**Figure S24a:** ***Interpretation of features for selected cancer types (brain; breast; colon and rectum) in PROV cohort.*** *(Left) Case-control enrichment among top-ranked positive IG scores, x-axes values are log odds ratio of feature rank hits. (Upper right) Occlusion shows mean predicted log odds with the event observed minus masked, with whiskers of +/-1.96*

*occurrence level errors, positive means observed feature score is higher. (Lower right) Score-change directions around medication and procedure transitions (-1 all retained changes decrease the score, +1 all increase it).*

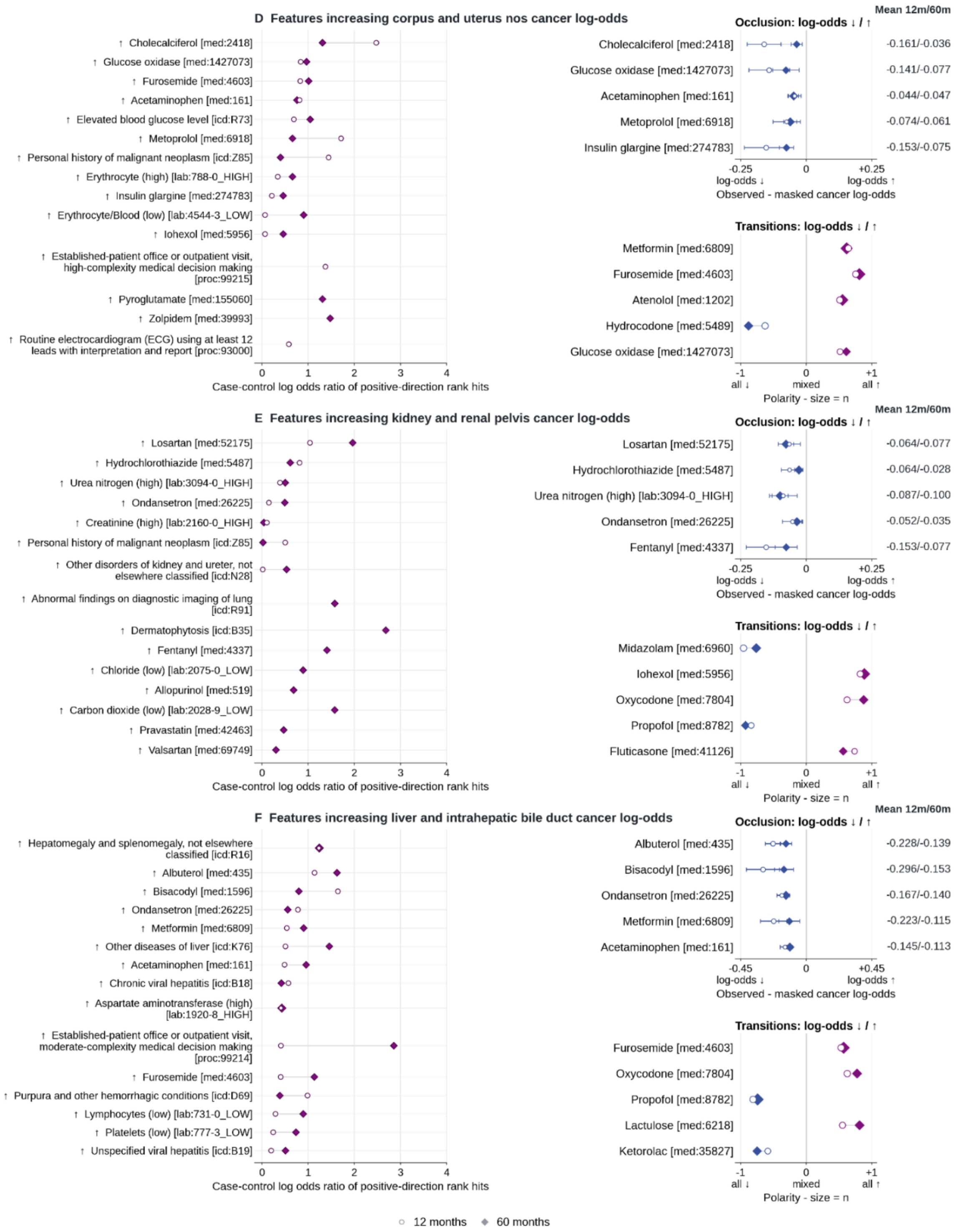


***Figure S24b: Interpretation of features for selected cancer types (corpus and uterus; kidney and renal pelvis; liver and intrahepatic bile duct) in PROV cohort.*** *(Left)*

*Case-control enrichment among top-ranked positive IG scores, x-axes values are log odds ratio of feature rank hits. (Upper right) Occlusion shows mean predicted log odds with the event observed minus masked, with whiskers of +/-1.96 occurrence level errors, positive means observed feature score is higher. (Lower right) Score-change directions around medication and procedure transitions (-1 all retained changes decrease the score, +1 all increase it).*

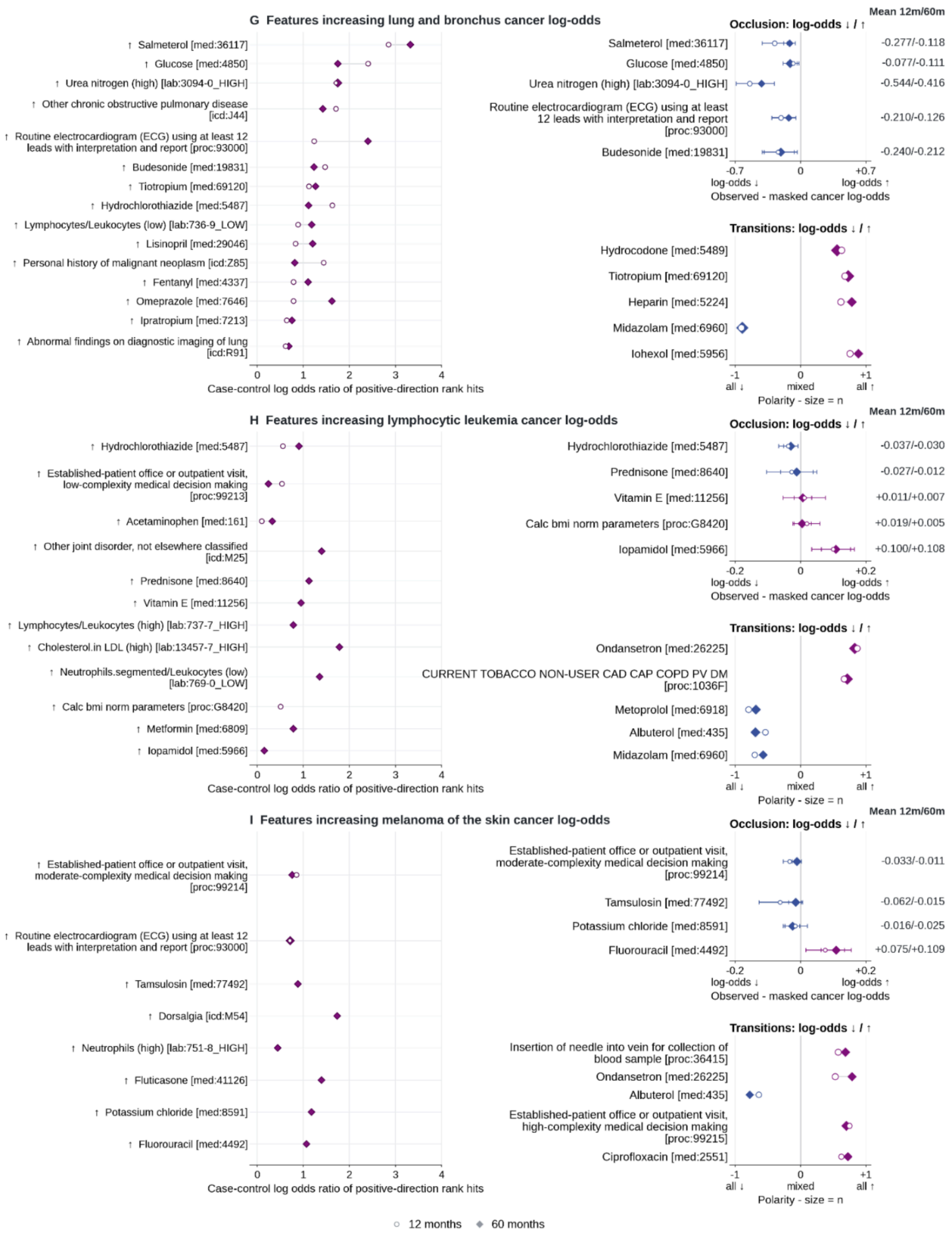

***Figure S24c: Interpretation of features for selected cancer types (lung and bronchus; melanoma of the skin;myeloid and monocytic leukemia) in PROV cohort.*** *(Left) Case-control enrichment among top-ranked positive IG scores, x-axes values are log odds ratio of feature rank hits. (Upper right) Occlusion shows mean predicted log odds with the event observed minus masked, with whiskers of +/-1.96 occurrence level errors, positive means observed feature score is higher. (Lower right) Score-change directions around medication and procedure transitions (-1 all retained changes decrease the score, +1 all increase it).*

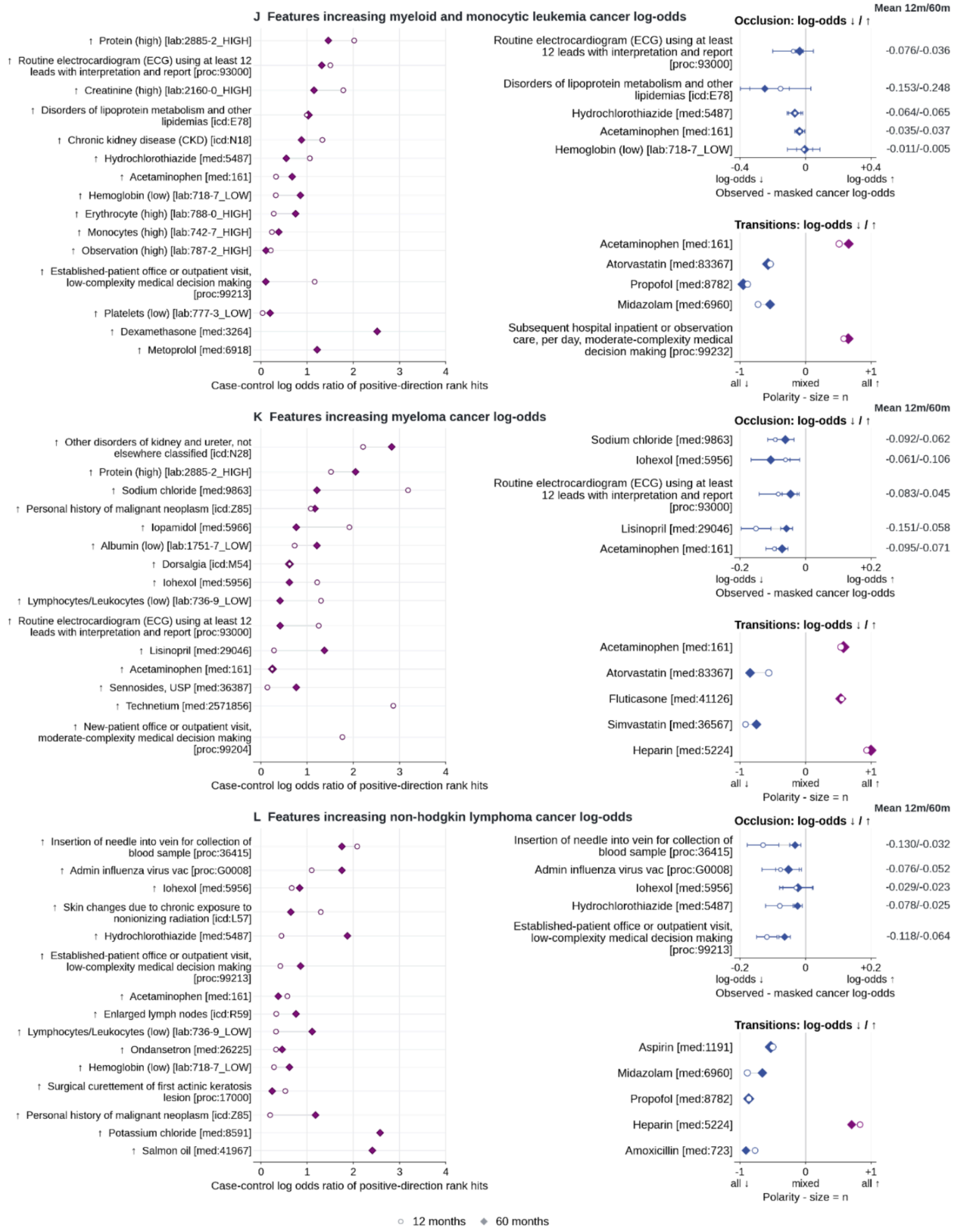

**Figure S24d:** ***Interpretation of features for selected cancer types in PROV cohort (myeloid and monocytic lymphoma; myeloma; non-Hodgkin lymphoma).*** *(Left) Case-control enrichment among top-ranked positive IG scores, x-axes values are log odds ratio of feature rank hits. (Upper right) Occlusion shows mean predicted log odds with the event observed minus masked, with whiskers of +/-1.96 occurrence level errors, positive means observed feature score is higher. (Lower right) Score-change directions around medication and procedure transitions (-1 all retained changes decrease the score, +1 all increase it).*

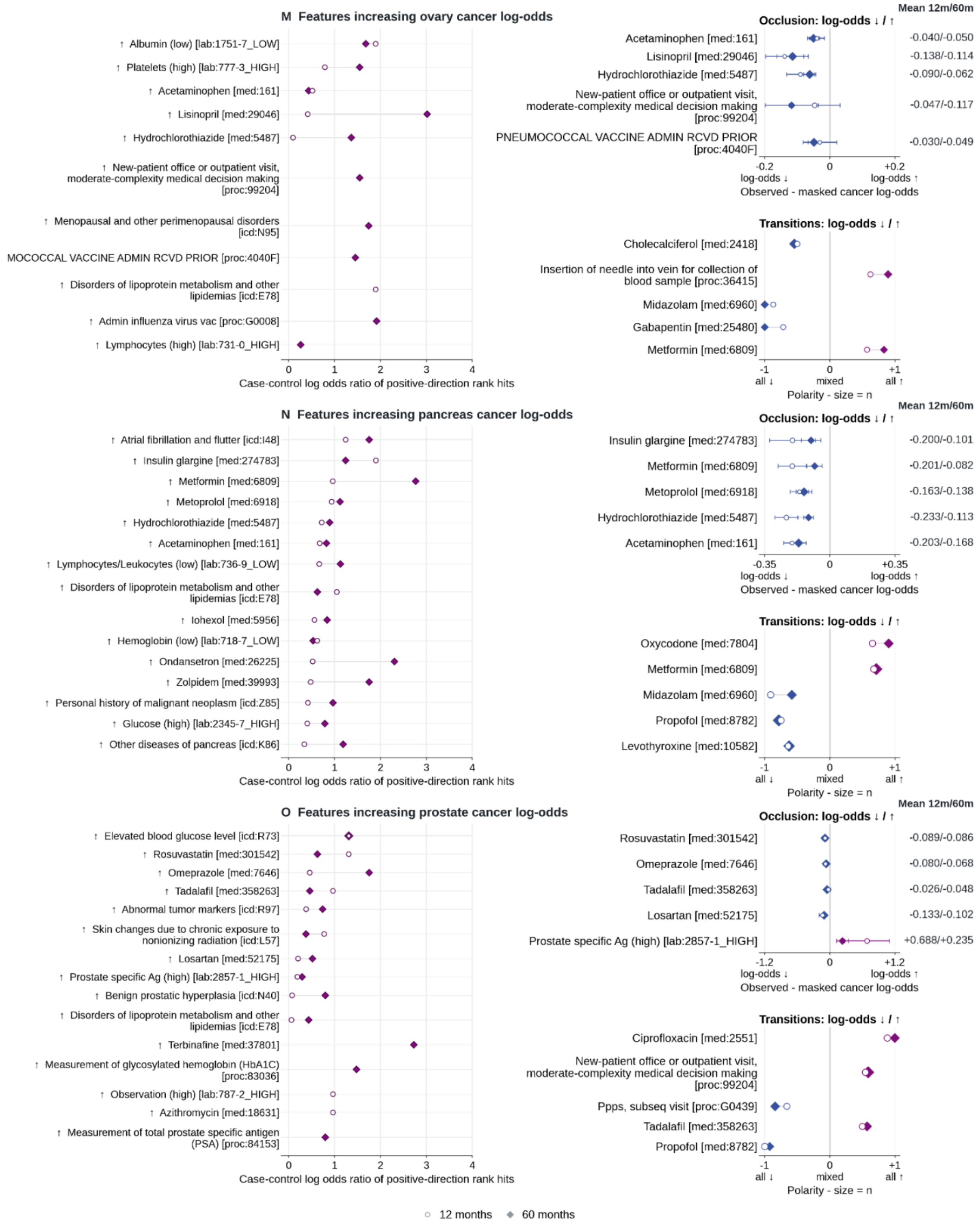

***Figure S24e: Interpretation of features for selected cancer types in PROV cohort (ovary; pancreas; prostate).*** *(Left) Case-control enrichment among top-ranked positive IG scores, x-axes values are log odds ratio of feature rank hits. (Upper right) Occlusion shows mean predicted log odds with the event observed minus masked, with whiskers of +/-1.96 occurrence level errors, positive means observed feature score is higher. (Lower right) Score-change directions around medication and procedure transitions (-1 all retained changes decrease the score, +1 all increase it).*

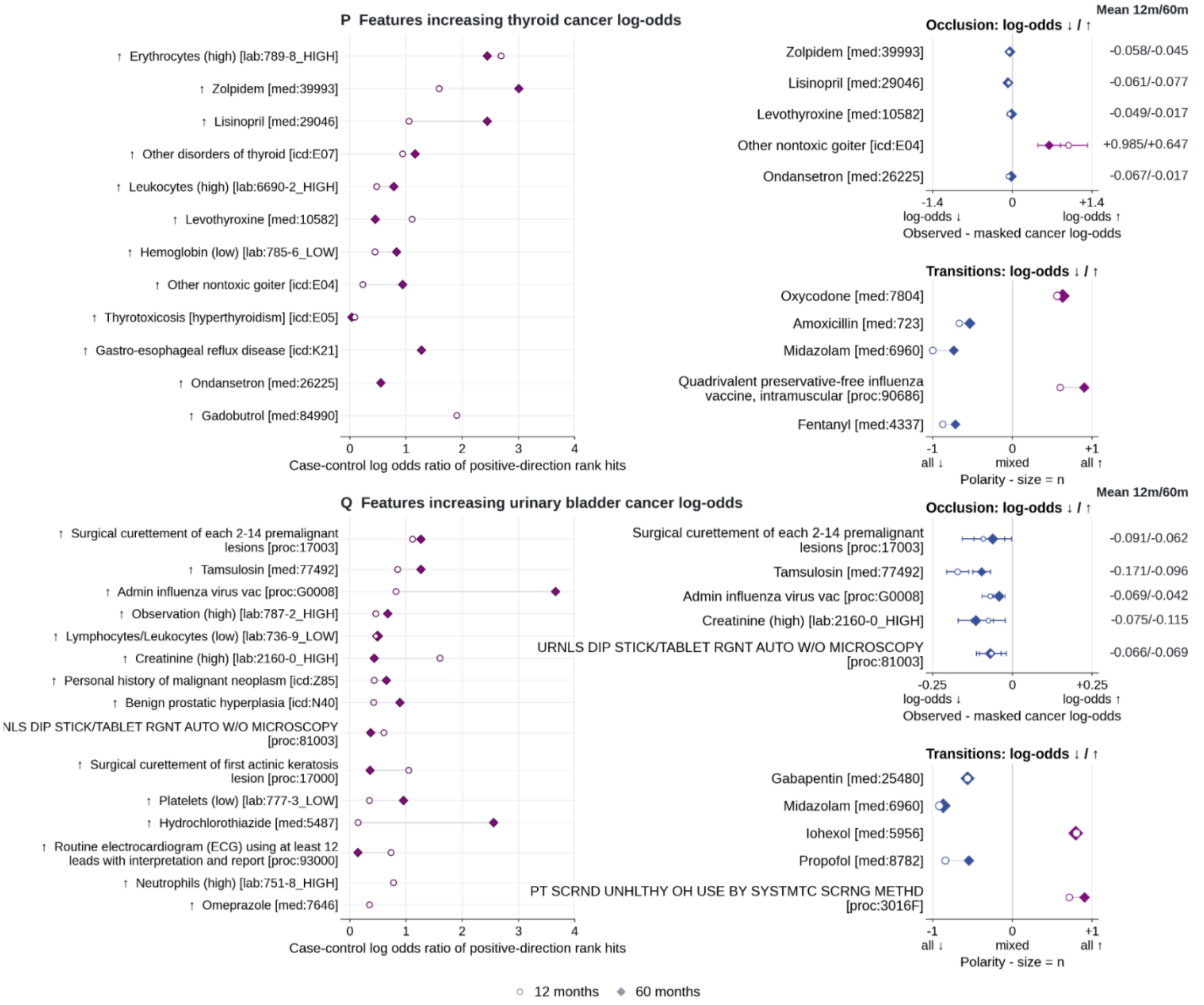


***Figure S24f: Interpretation of features for selected cancer types in PROV cohort (thyroid, urinary bladded).*** *(Left) Case-control enrichment among top-ranked positive IG scores, x-axes values are log odds ratio of feature rank hits. (Upper right) Occlusion shows mean predicted log odds with the event observed minus masked, with whiskers of +/-1.96 occurrence level errors, positive means observed feature score is higher. (Lower right) Score-change directions around medication and procedure transitions (-1 all retained changes decrease the score, +1 all increase it).*

### US-VA Results.

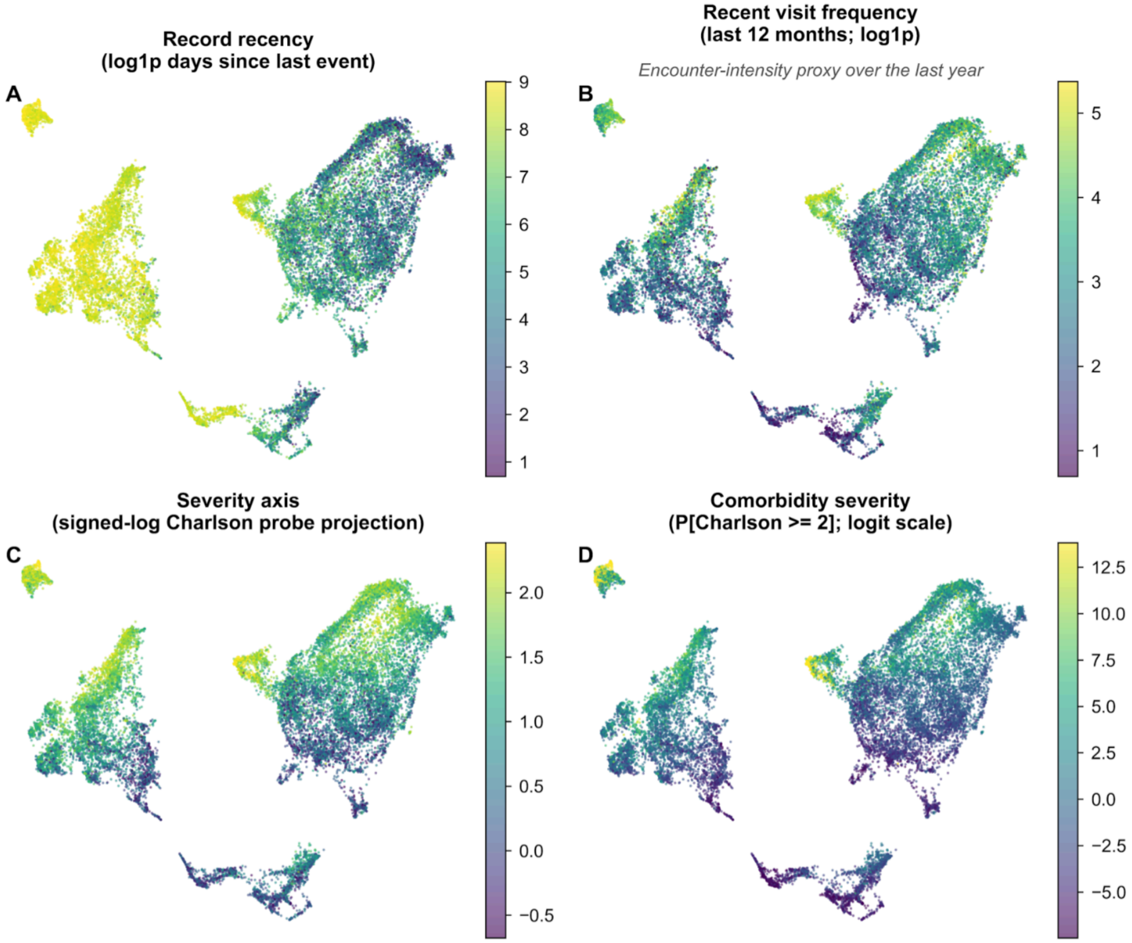


***Figure S25: Patient health state representation (VA Cohort).*** *All four panels are two dimensional projections of patient trajectory representation obtained from the penultimate layer of GenEHR. Each has a different overlay: (A) time gap since last visit (B) number of visits in preceding year as a proxy for acuity. (C) a linear probe was fitted on feature representation for predicting CCI overlay with probe prediction (D) only patients who have CCI above chosen threshold. Gradients in CCI imply that representations capture information about overall morbidity and healthcare use.*

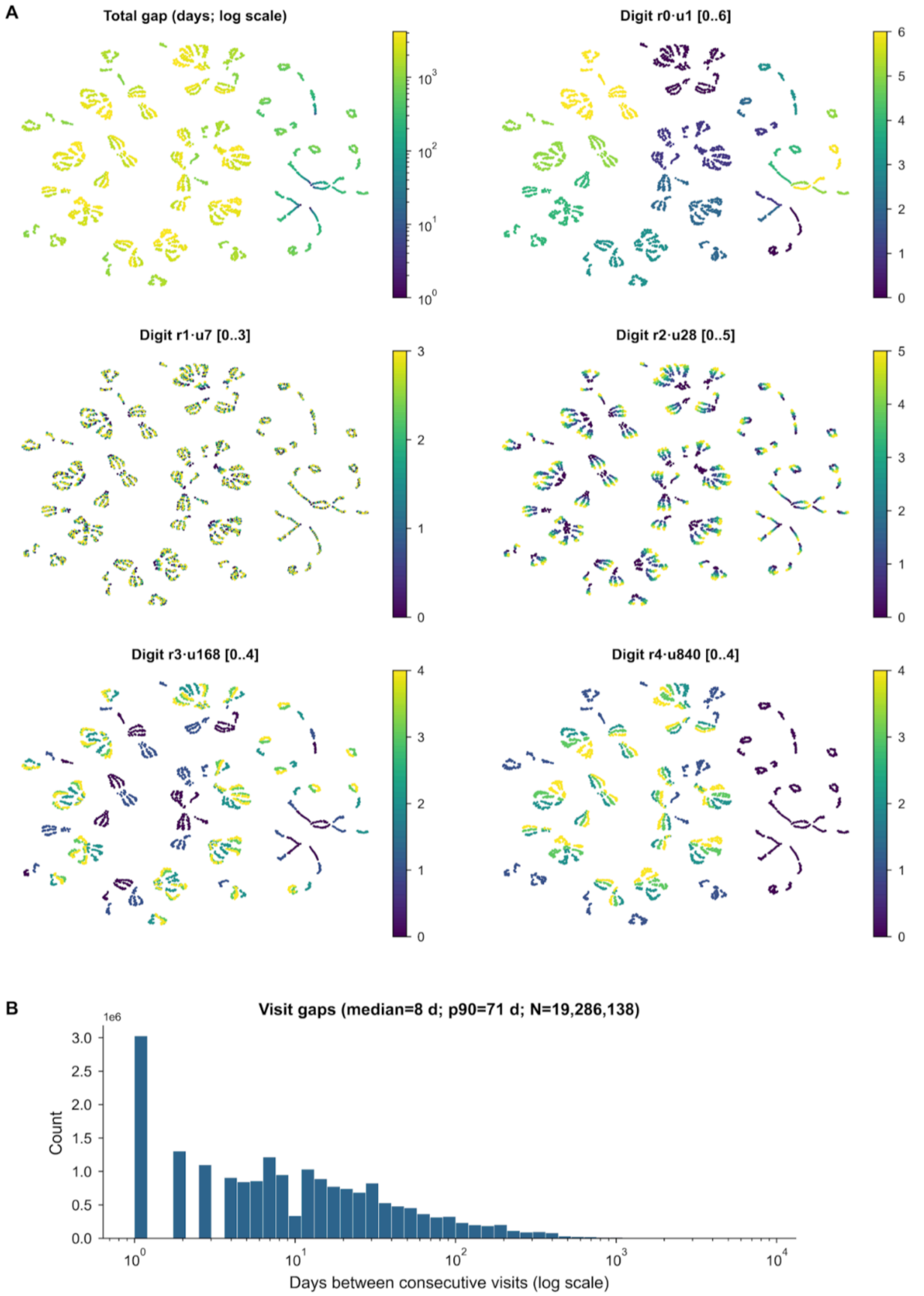


***Figure S26: Learned RATE representation and observed time gaps in VA.*** *UMAP projection of learned representation of 4200 days, colored by the value of each radix digit together with the observed distribution of inter-visit gaps. (A) Each point represents one encoded gap from 1 to 4200 days where color is the gap length. The other five show the digit with place values 1, 7, 28,*

*168, and 840 days with color indicating lower to higher values on each scale. (B) Inter-visit time interval distribution.*

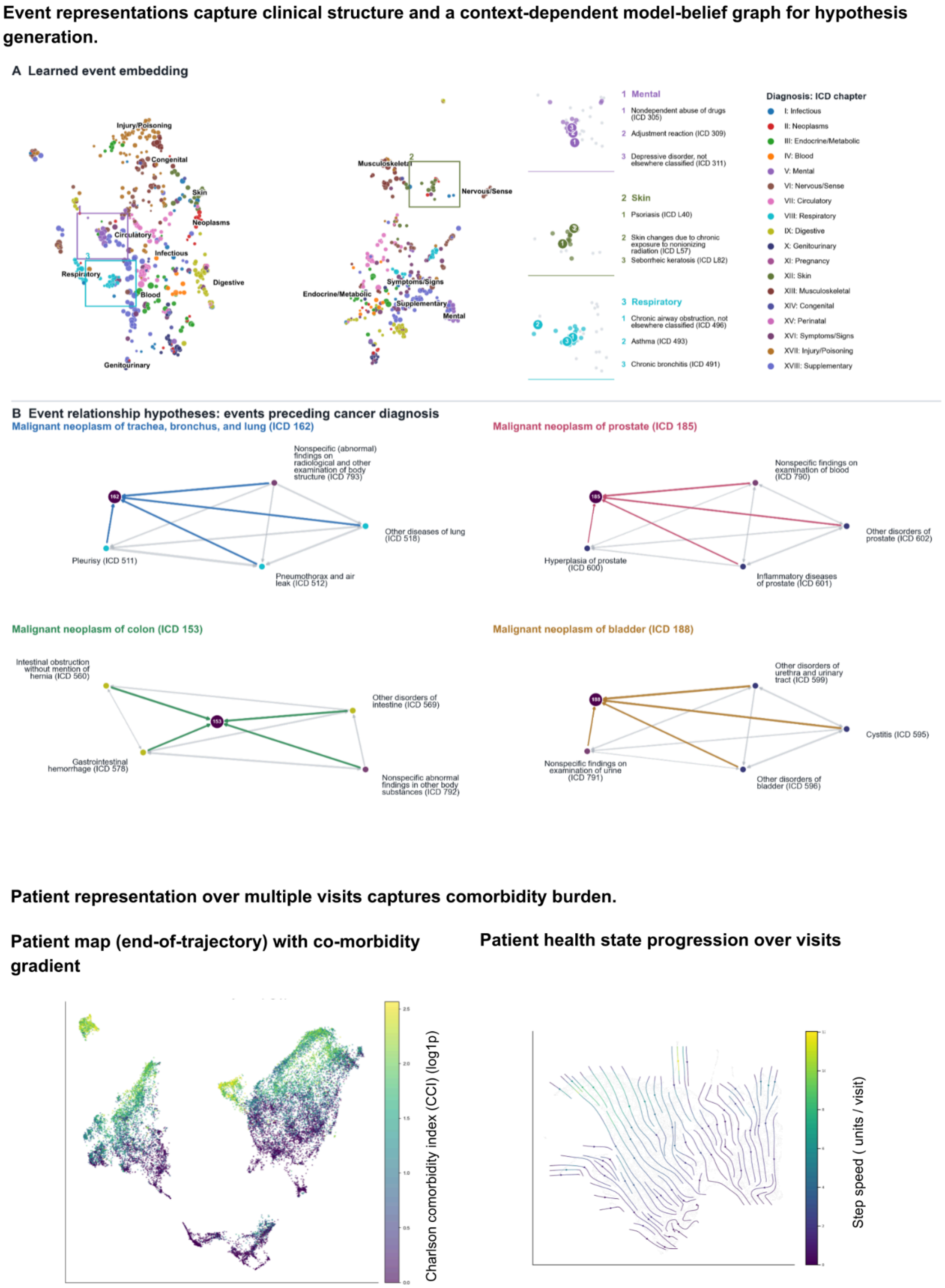


***Figure S27: For the VA cohort.*** *Left:* ***(A)*** *UMAP projection of learned event embeddings using cosine distance between embedding vectors. Right: closeup of three regions numbered 1 to 3 shows GenEHR learns hierarchy of clinical events without explicit prior knowledge and without*

using text. ***(B)*** *Event relationship preceding cancer diagnosis inferred using cPMI between pairs of events for selected cancers (ICD codes 162, 153, 185, 188). For held-out patients whose preceding visit contains an event $e_1$, cPMI compares the average model probability assigned to $e_2$ normalized against the model averaged probability of $e_2$ across all positions and patients. In the cPMI belief graph nodes are clinical events and edges are informational links. Diagnosis neighborhood of cancer with edges among preceding events. Dashed edges: events with weaker mutual information scores that are plausibly less specific to that cancer. (****Bottom)*** *UMAP neighborhoods of patients using their end-of-trajectory <eos> representation reflect gradient of comorbidity burden (left), and transitions along trajectories are informative about how the learned state of patients evolves over time using their <visit> representation over multiple visits. Flow (arrows) indicates how patient health evolves over visits (right) averaged over groups of held-out patients.*

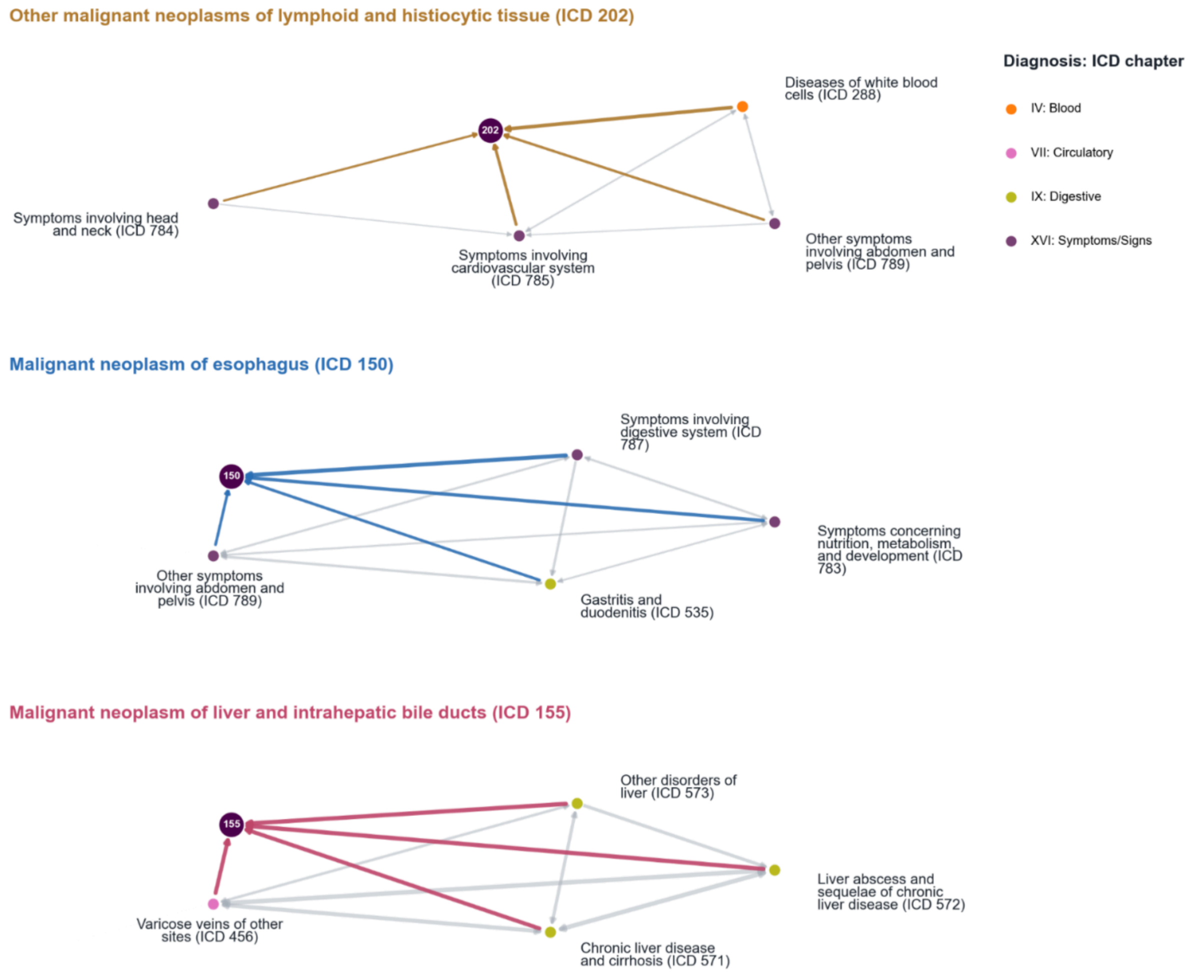


***Figure S28a: Breakdown of within modality belief graphs across cancer types (US-VA).*** *Event relationship preceding cancer diagnosis inferred using cPMI between pairs of events for selected cancers (ICD codes 202, 150, 155). For held-out patients whose preceding visit contains an event $e_1$, cPMI compares the average model probability assigned to $e_2$ normalized against the model averaged probability of $e_2$ across all positions and patients. In the cPMI belief*

*graph nodes are clinical events and edges are informational links. Diagnosis neighborhood of cancer with edges among preceding events. Dashed edges: events with weaker mutual information scores that are plausibly less specific to that cancer.*

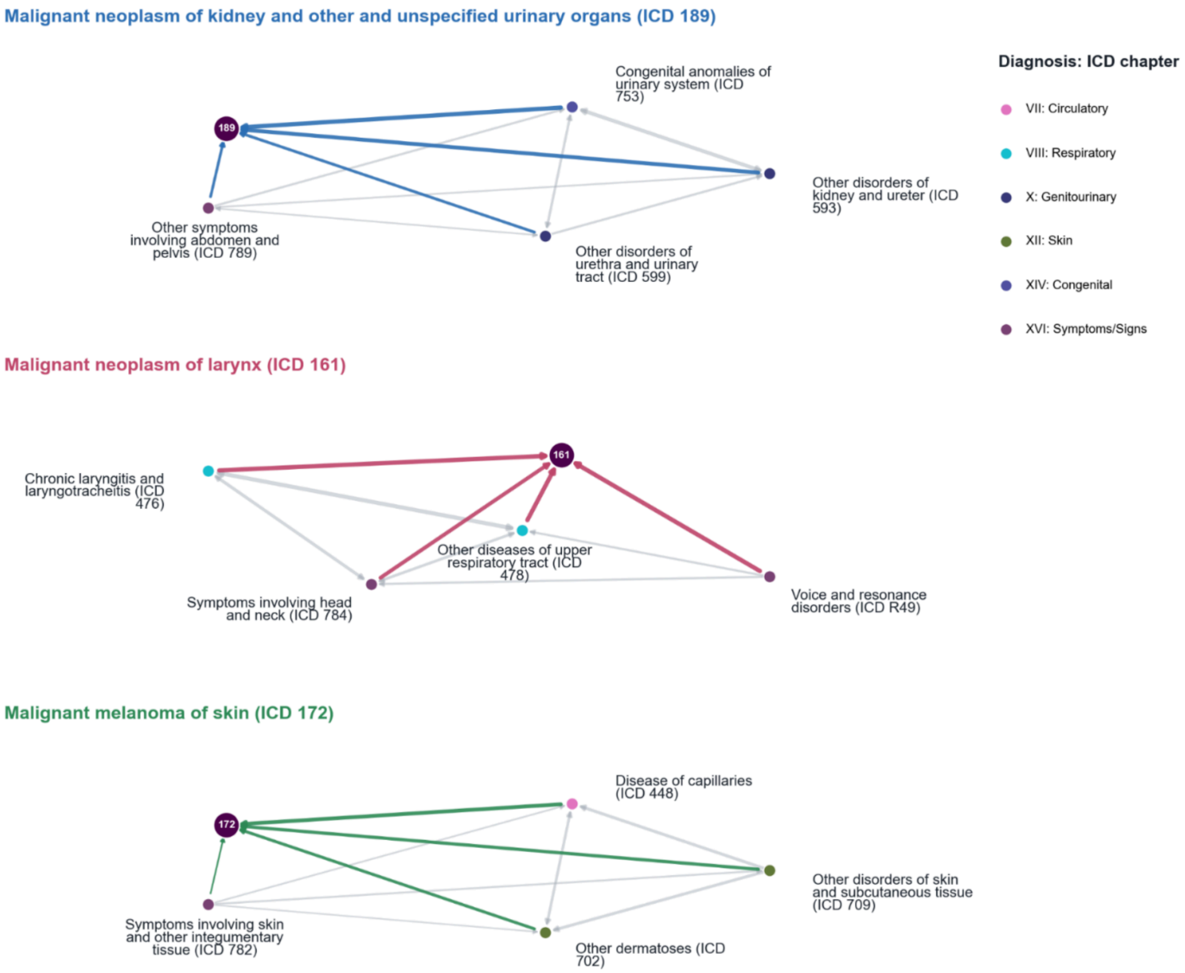


***Figure S28b: Breakdown of within modality belief graphs across cancer types (US-VA).*** *Event relationship preceding cancer diagnosis inferred using cPMI between pairs of events for selected cancers (ICD codes 189, 161, 172). For held-out patients whose preceding visit contains an event $e_1$, cPMI compares the average model probability assigned to $e_2$ normalized against the model averaged probability of $e_2$ across all positions and patients. In the cPMI belief graph nodes are clinical events and edges are informational links. Diagnosis neighborhood of cancer with edges among preceding events. Dashed edges: events with weaker mutual information scores that are plausibly less specific to that cancer.*

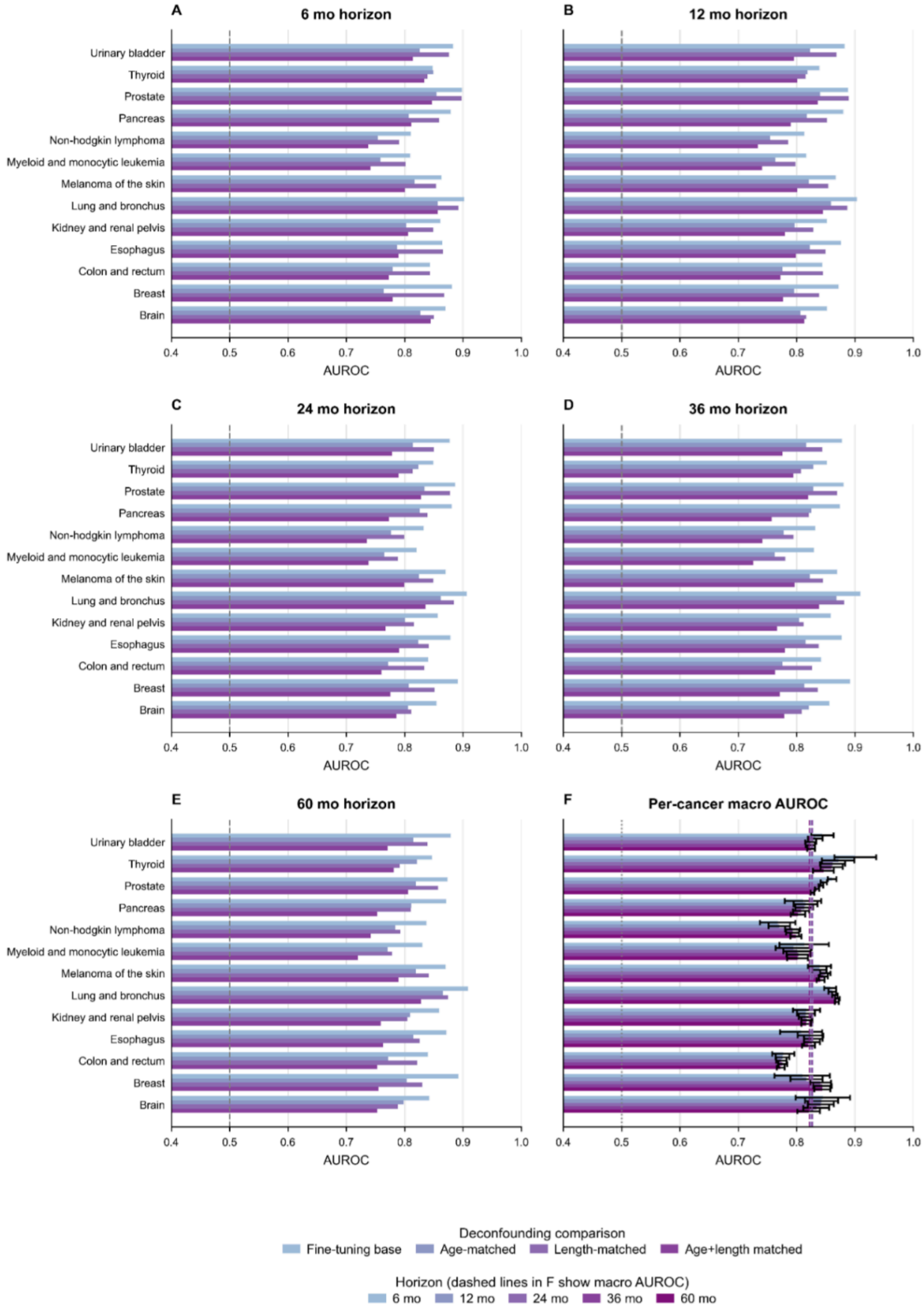


***Figure S29a: Sensitivity of US-VA cohort cancer discrimination to age and trajectory length on cancer risk.*** *Cases and controls were matched by age bracket at the endpoint of trajectory as well as with an equal number of events. Age matched results in a drop of performance, but overall performance shows the model just doesn't rely on age and learns from the two combined. Length matched shows the results are not sensitive to the number of codes in patient trajectory. (A-E) are results for 6, 12, 24, 36, and 60 month horizons, with each row comparing original evaluation with age matching, event count matching, and both combined.*

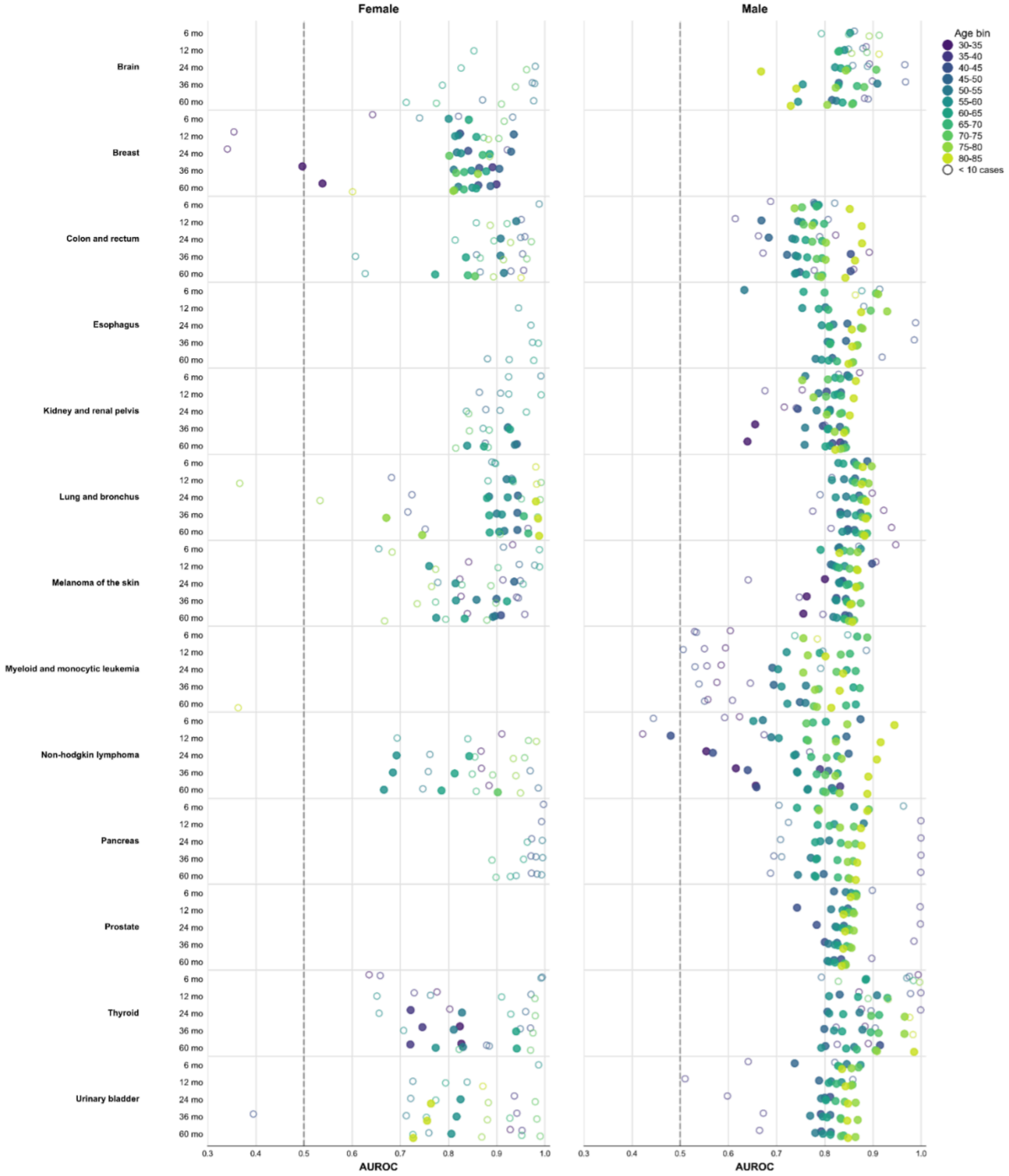


***Figure S29b: (US-VA Cohort) Breakdown of predictive performance by age-brackets and sex.*** *Performance by age and sex bracket for different groups assessing heterogeneity and data sparsity. (Left) Performance on female population for each cancer (Right) Performance on male population for each cancer. Both are reported for all six (6, 12, 24, 36, and 60 months)*

*prediction horizons. Color encodes five year age groups, and hollow circle means fewer than 10 positive outcomes in each bin.*

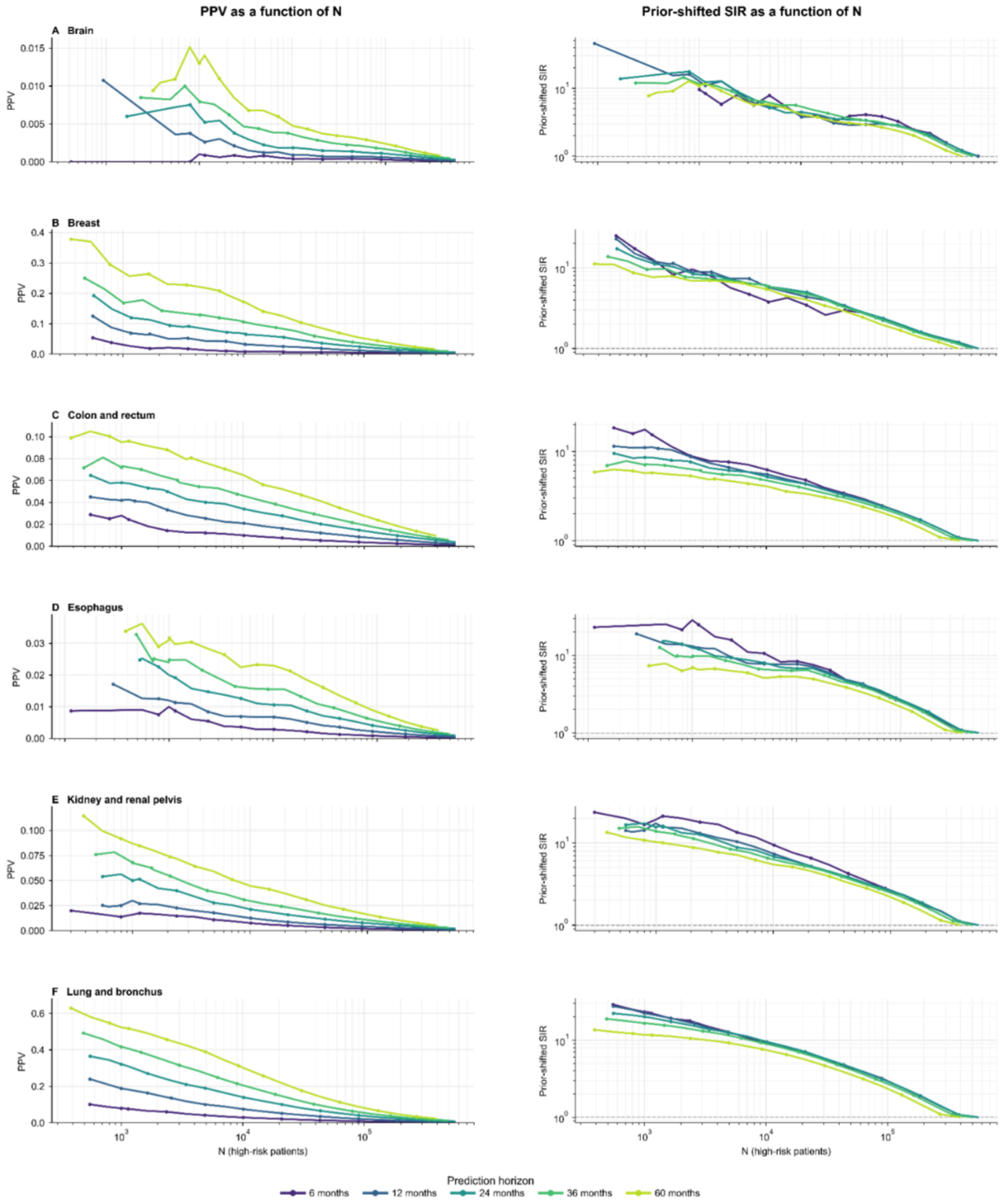


***Figure S30a: (VA Cohort) Clinical utility of GenEHR fine-tuned model for nominating patients for screening by choosing an operating threshold****. PPV (left) and SIR (right) plotted as a function of operating threshold N shows a net benefit of detecting one cancer at the cost of increasing false positives. (A-F) brain, breast, colon/rectum, corpus/uterus, esophagus, kidney/renal-pelvis, and lung and bronchus cancer.*

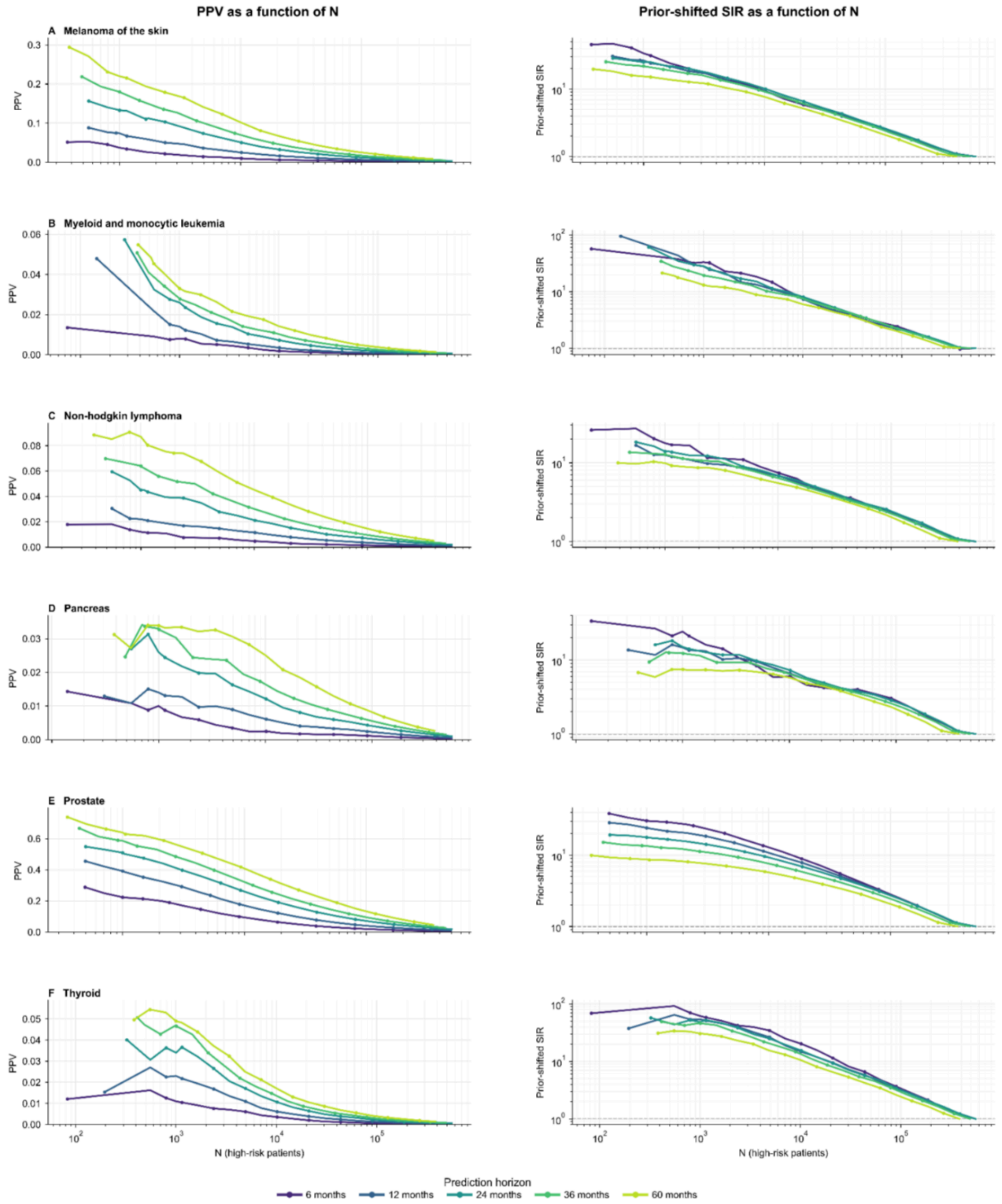


***Figure S30b: (VA Cohort) Clinical utility of GenEHR fine-tuned model for nominating patients for screening by choosing an operating threshold****. PPV (left) and SIR (right) plotted as a function of operating threshold N shows a net benefit of detecting one cancer at the cost of increasing false positives. (A-F) Melanoma of the skin, myeloid and monocytic leukemia, non-hodgkin lymphoma, pancreas, prostate, and thyroid cancer.*

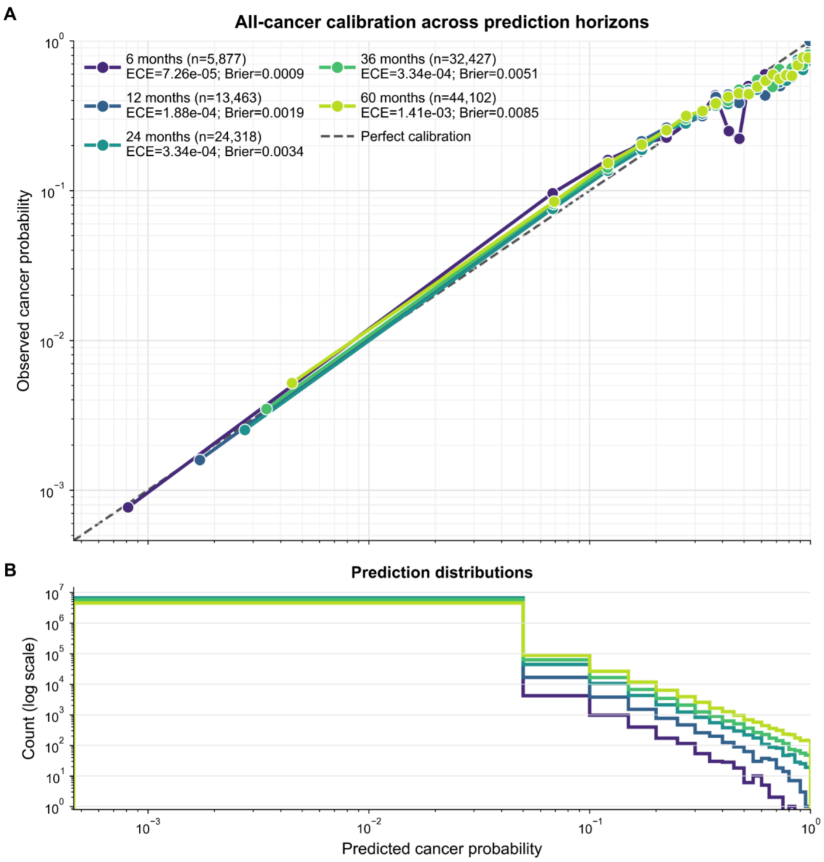


***Figure S31: (VA Cohort) Pan-cancer risk model is well calibrated across horizons and cancer types.*** *Observed cancer frequency against predicted probability across calibration bins for different horizons, where diagonal line indicates perfect calibration. Both ECE and Brier score describe fine-tuned cancer-risk probabilities (not next event calibration in Fig 3).*

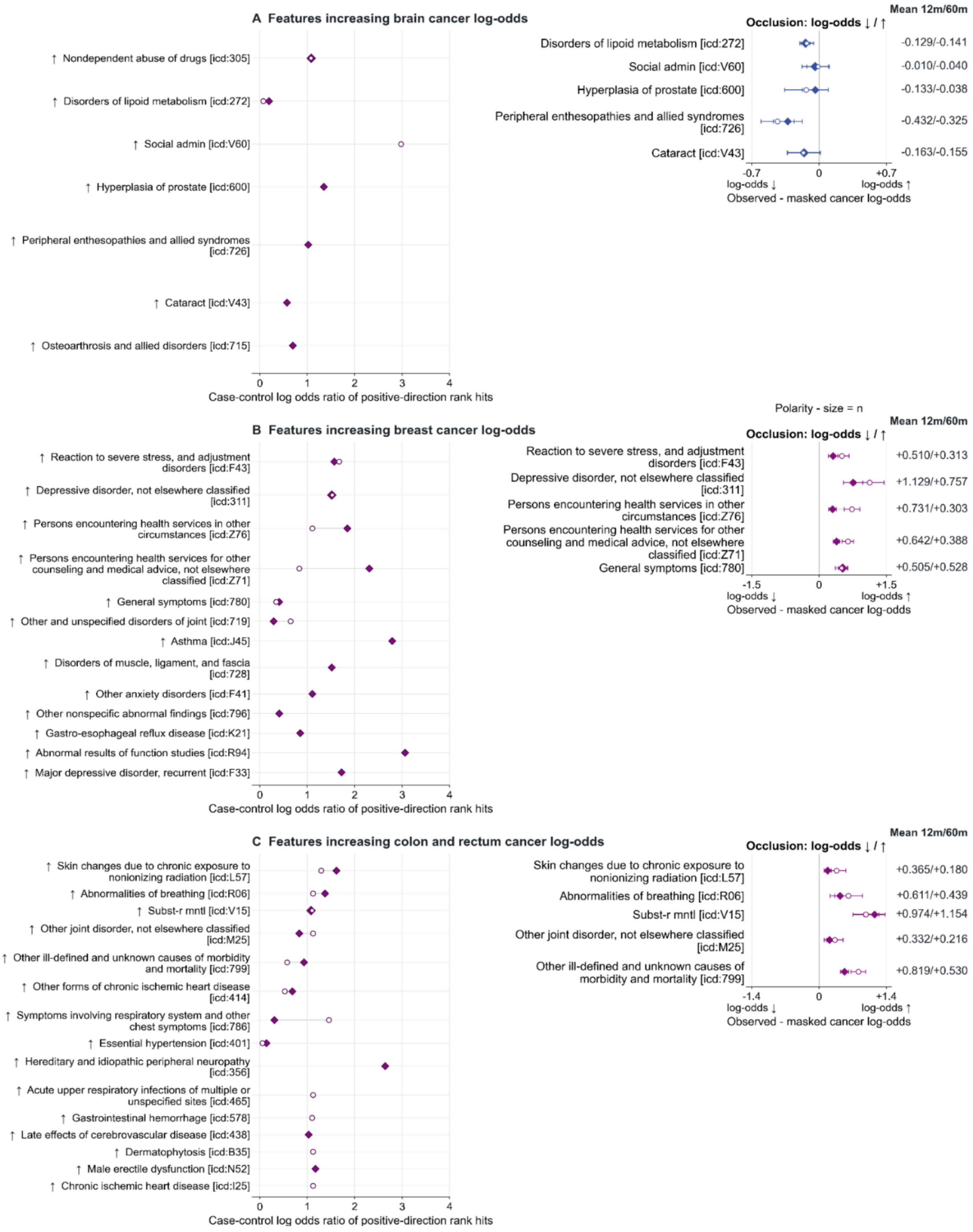


***Figure S32a: Interpretation of features for selected cancer types (brain; breast; colon and rectum) in VA cohort.*** *(Left) Case-control enrichment among top-ranked positive IG scores, x-axes values are log odds ratio of feature rank hits. (Right) Occlusion shows mean predicted*

*log odds with the event observed minus masked, with whiskers of +/-1.96 occurrence level errors, positive means observed feature score is higher.*

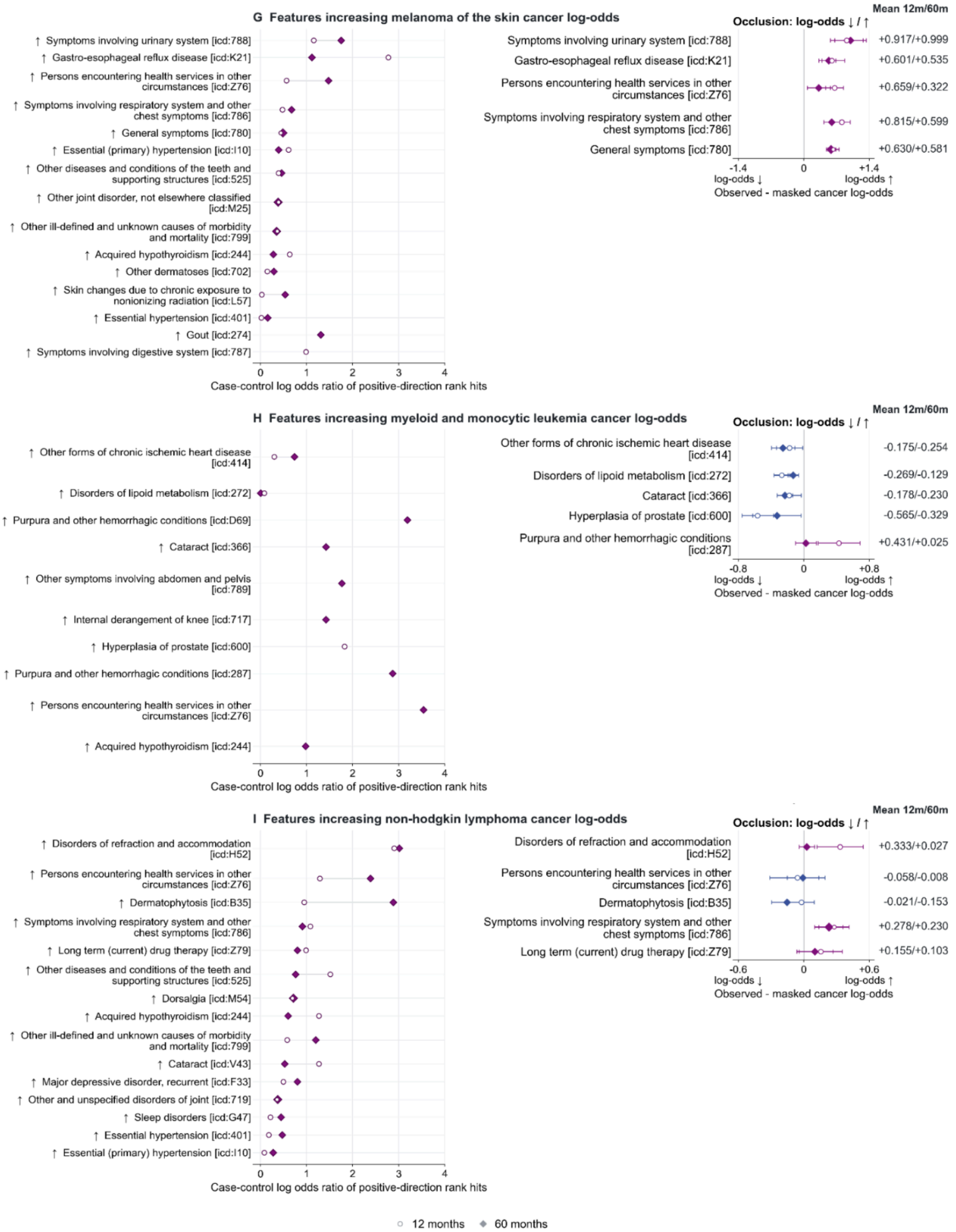


**Figure S32b:** ***Interpretation of features for selected cancer types (melanoma of the skin; myeloid and monocytic leukemia; non-hodgkin lymphoma) in VA cohort.*** *(Left) Case-control enrichment among top-ranked positive IG scores, x-axes values are log odds ratio*

*of feature rank hits. (Right) Occlusion shows mean predicted log odds with the event observed minus masked, with whiskers of +/-1.96 occurrence level errors, positive means observed feature score is higher.*

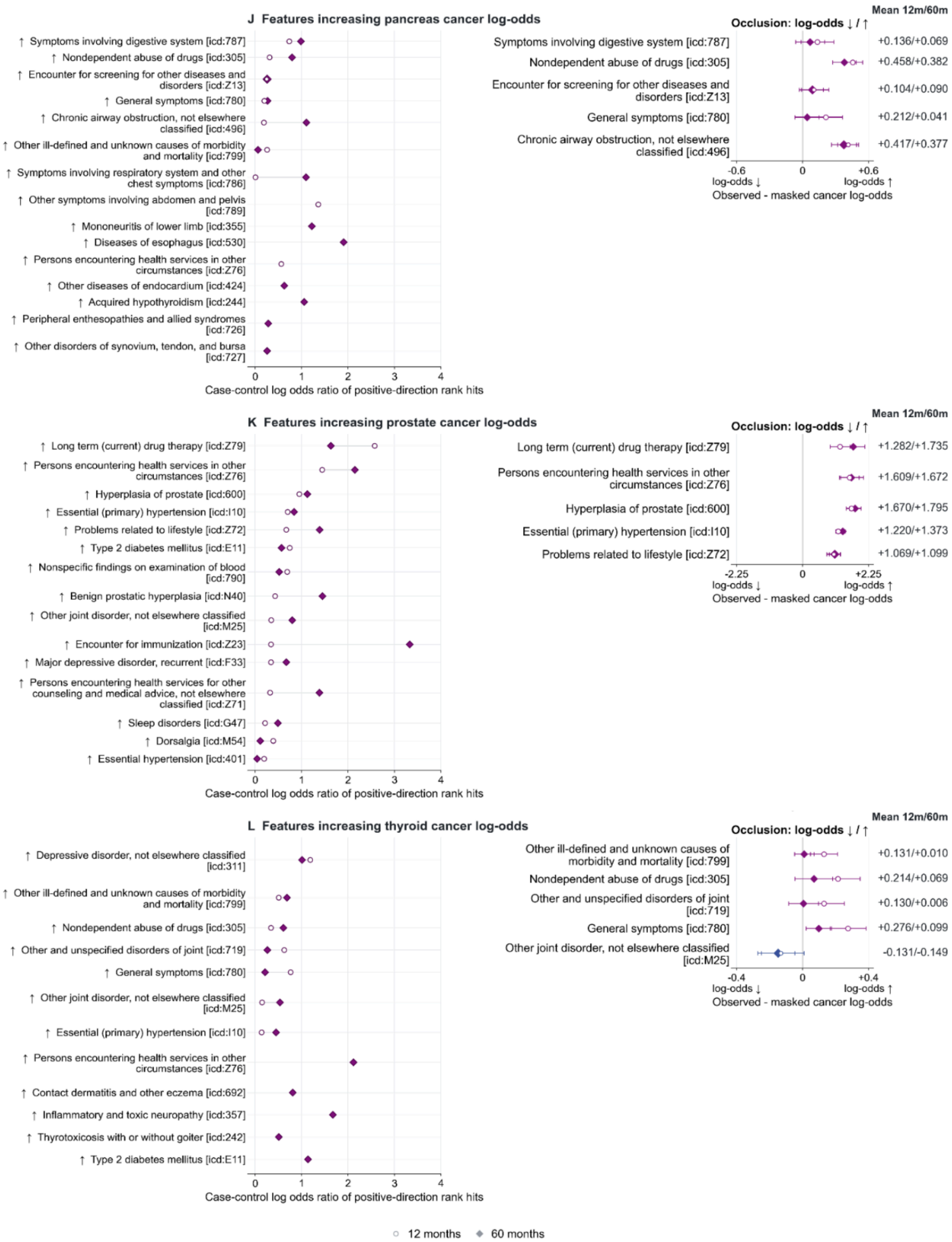


***Figure S32c: Interpretation of features for selected cancer types (pancreas; prostate; thyroid) in VA cohort.** (Left) Case-control enrichment among top-ranked positive IG scores, x-axes values are log odds ratio of feature rank hits. (Right) Occlusion shows mean predicted*

*log odds with the event observed minus masked, with whiskers of +/-1.96 occurrence level errors, positive means observed feature score is higher.*

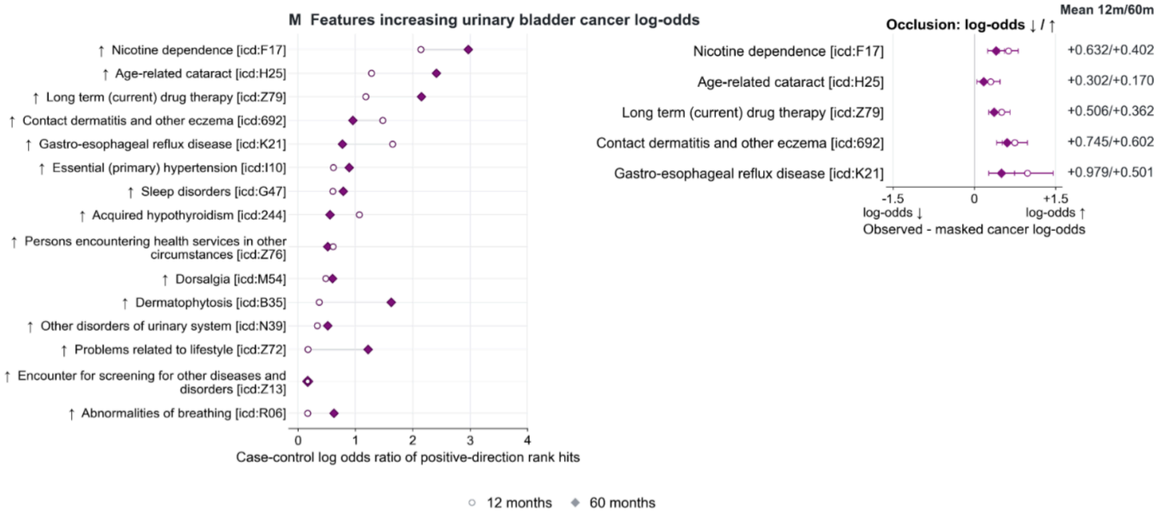


***Figure S32d: Interpretation of features for selected cancer types (urinary bladder) in VA cohort.*** *(Left) Case-control enrichment among top-ranked positive IG scores, x-axes values are log odds ratio of feature rank hits. (Right) Occlusion shows mean predicted log odds with the event observed minus masked, with whiskers of +/-1.96 occurrence level errors, positive means observed feature score is higher.*

### UK Biobank Results.

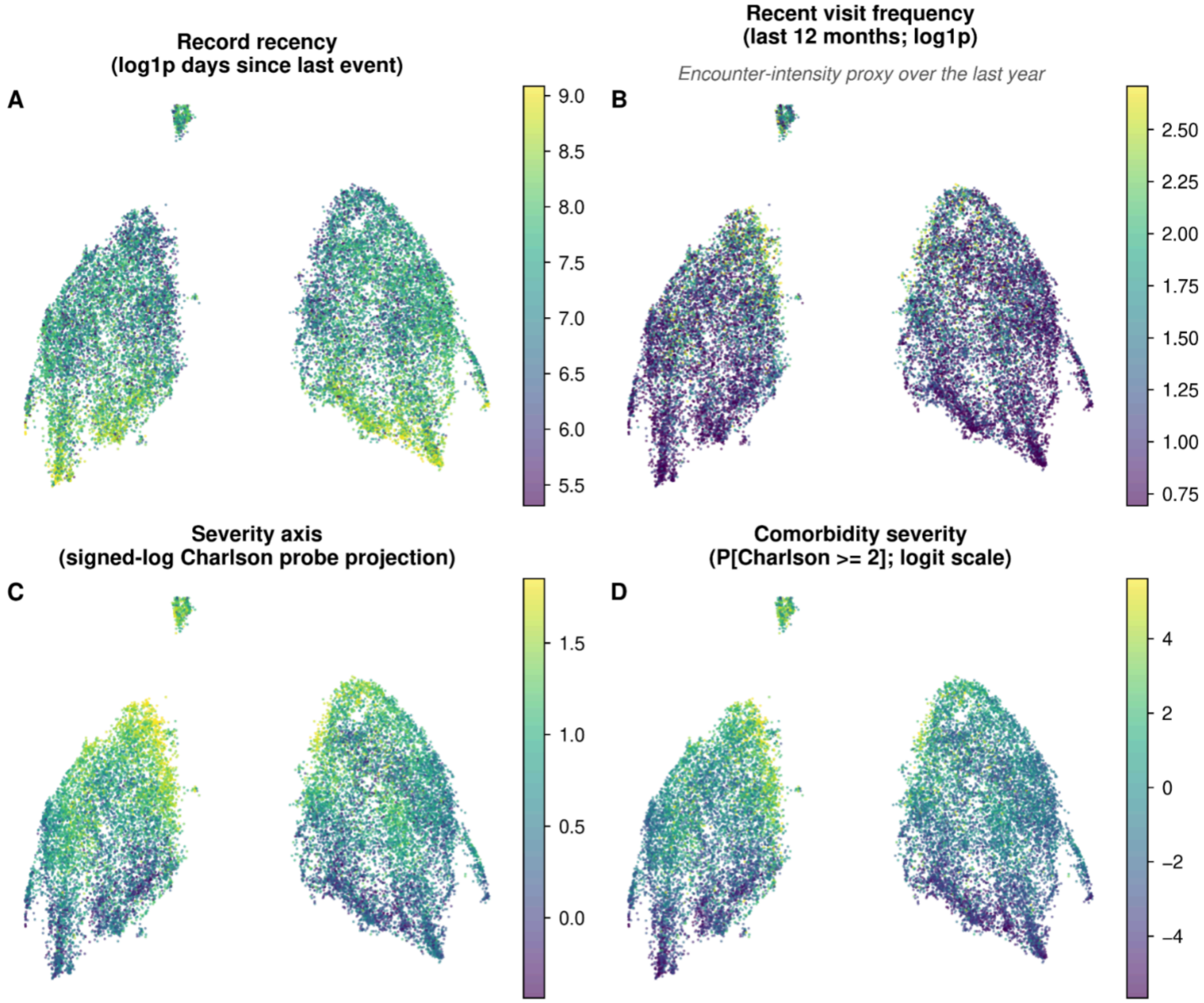


***Figure S33: Patient health state representation given by pooled encoding of all events in patient's trajectory (UKBB cohort).*** *All four panels are two dimensional projections of patient trajectory representation obtained from the penultimate layer of GenEHR. Each has a different overlay: (A) time gap since last visit (B) number of visits in preceding year as a proxy for acuity. (C) a linear probe was fitted on feature representation for predicting CCI overlay with probe prediction (D) only patients who have CCI above chosen threshold. Gradients in CCI imply that representations capture information about overall morbidity and healthcare use.*

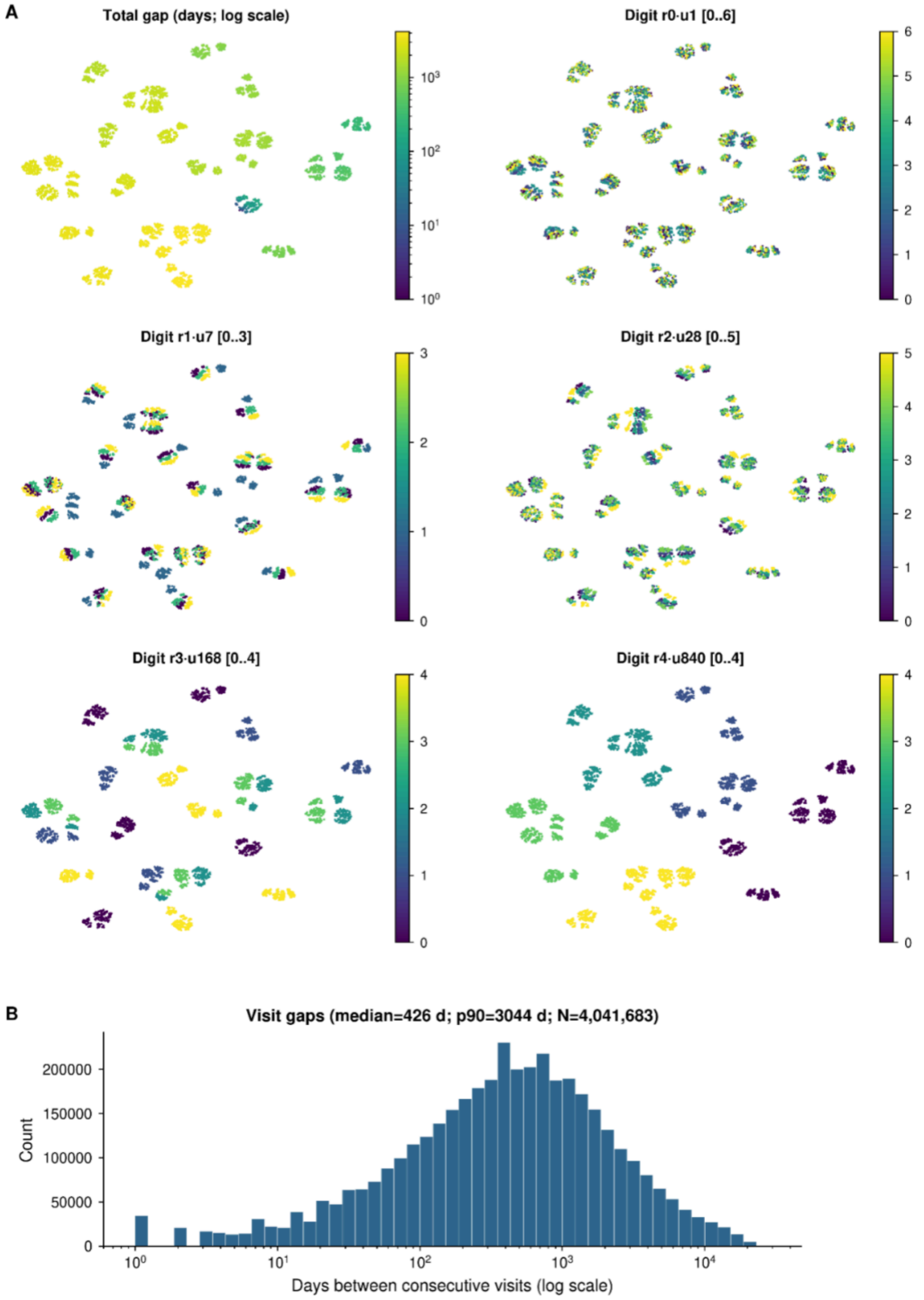


***Figure S34: Learned RATE representation and observed time gaps in UKBB cohort.*** *UMAP projection of learned representation of 4200 days, colored by the value of each radix digit together with the observed distribution of inter-visit gaps. (A) Each point represents one encoded gap from 1 to 4200 days where color is the gap length. The other five show the digit with place values 1, 7, 28, 168, and 840 days with color indicating lower to higher values on each scale. (B) Inter-visit time interval distribution.*

**Event representations capture clinical structure and a context-dependent model-belief graph for hypothesis generation.**

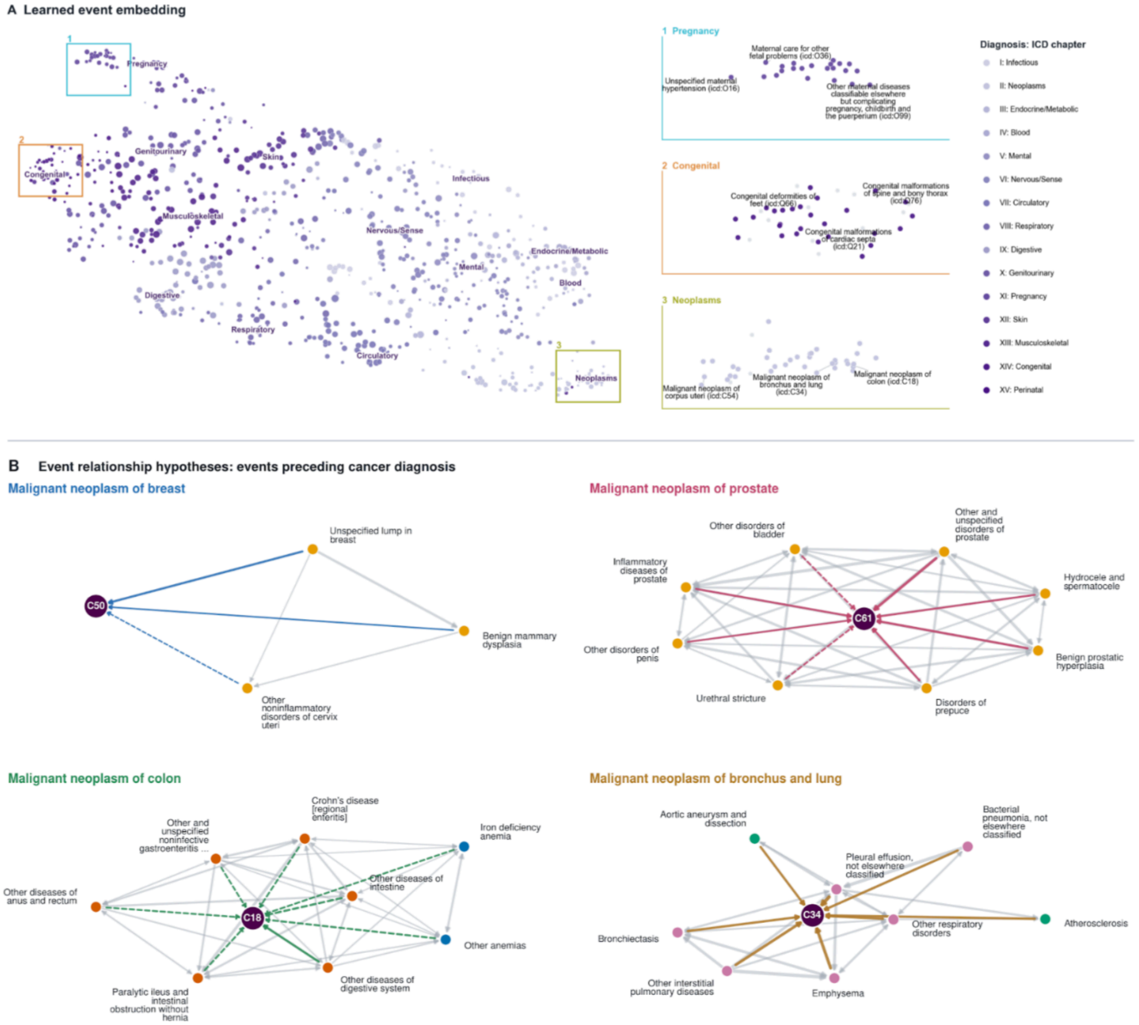


**Patient representation over multiple visits captures comorbidity burden.**

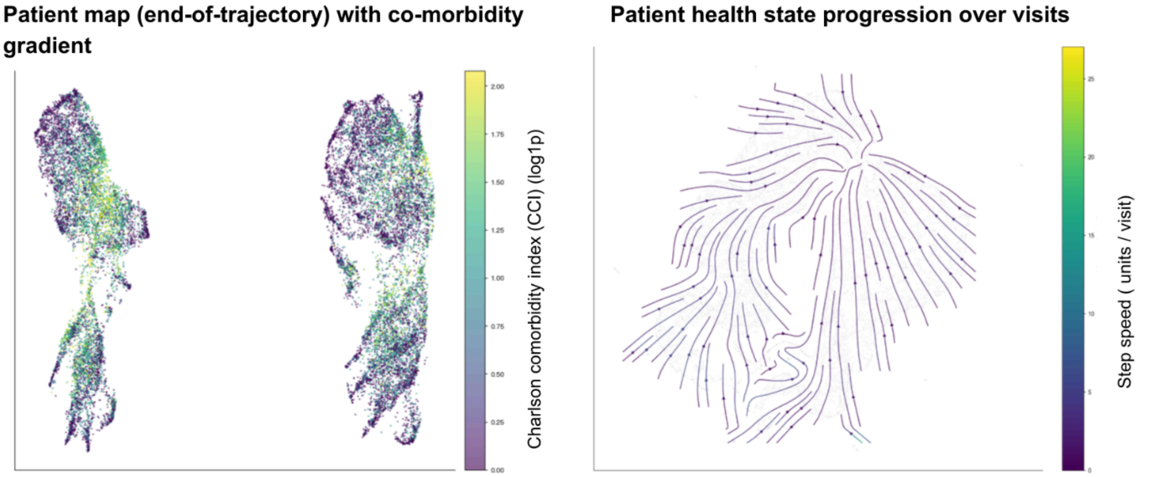


***Figure S35: For the UKBB cohort.*** *Left:* ***(A)*** *UMAP projection of learned event embeddings*

*using cosine distance between embedding vectors. Right: closeup of three regions numbered 1 to 3 shows GenEHR learns hierarchy of clinical events without explicit prior knowledge and without using text.* ***(B)*** *Event relationship preceding cancer diagnosis inferred using cPMI between pairs of events for selected cancers (ICD codes C50, C61, C18, C34). For held-out patients whose preceding visit contains an event $e_1$, cPMI compares the average model probability assigned to $e_2$ normalized against the model averaged probability of $e_2$ across all positions and patients. In the cPMI belief graph nodes are clinical events and edges are informational links. Diagnosis neighborhood of cancer with edges among preceding events. Dashed edges: events with weaker mutual information scores that are plausibly less specific to that cancer. (**Bottom)*** *UMAP neighborhoods of patients using their end-of-trajectory <eos> representation reflect gradient of comorbidity burden (left), and transitions along trajectories are informative about how the learned state of patients evolves over time using their <visit> representation over multiple visits. Flow (arrows) indicates how patient health evolves over visits (right) averaged over groups of held-out patients.*

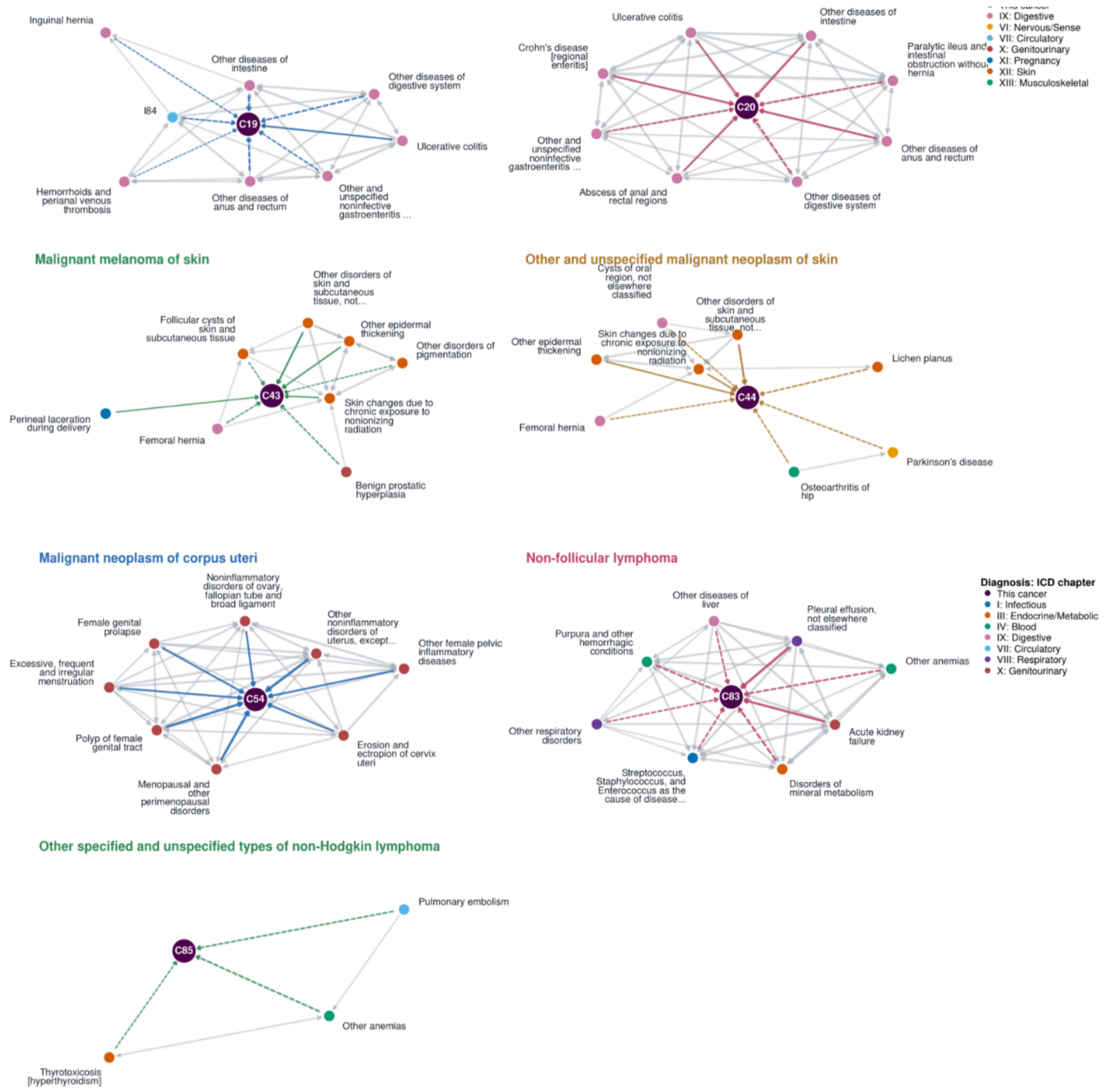


***Figure S36: Breakdown of within modality belief graphs across cancer types (UKBB).*** *UKBB first occurrence only has the diagnosis modality available. Event relationship preceding cancer diagnosis inferred using cPMI between pairs of events for selected cancers (ICD codes C19, C43, C54, C85). For held-out patients whose preceding visit contains an event* $e_1$*, cPMI compares the average model probability assigned to* $e_2$ *normalized against the model averaged probability of* $e_2$ *across all positions and patients. In the cPMI belief graph nodes are clinical events and edges are informational links. Diagnosis neighborhood of cancer with edges among preceding events, Dashed edges: events with weaker mutual information scores that are plausibly less specific to that cancer.*

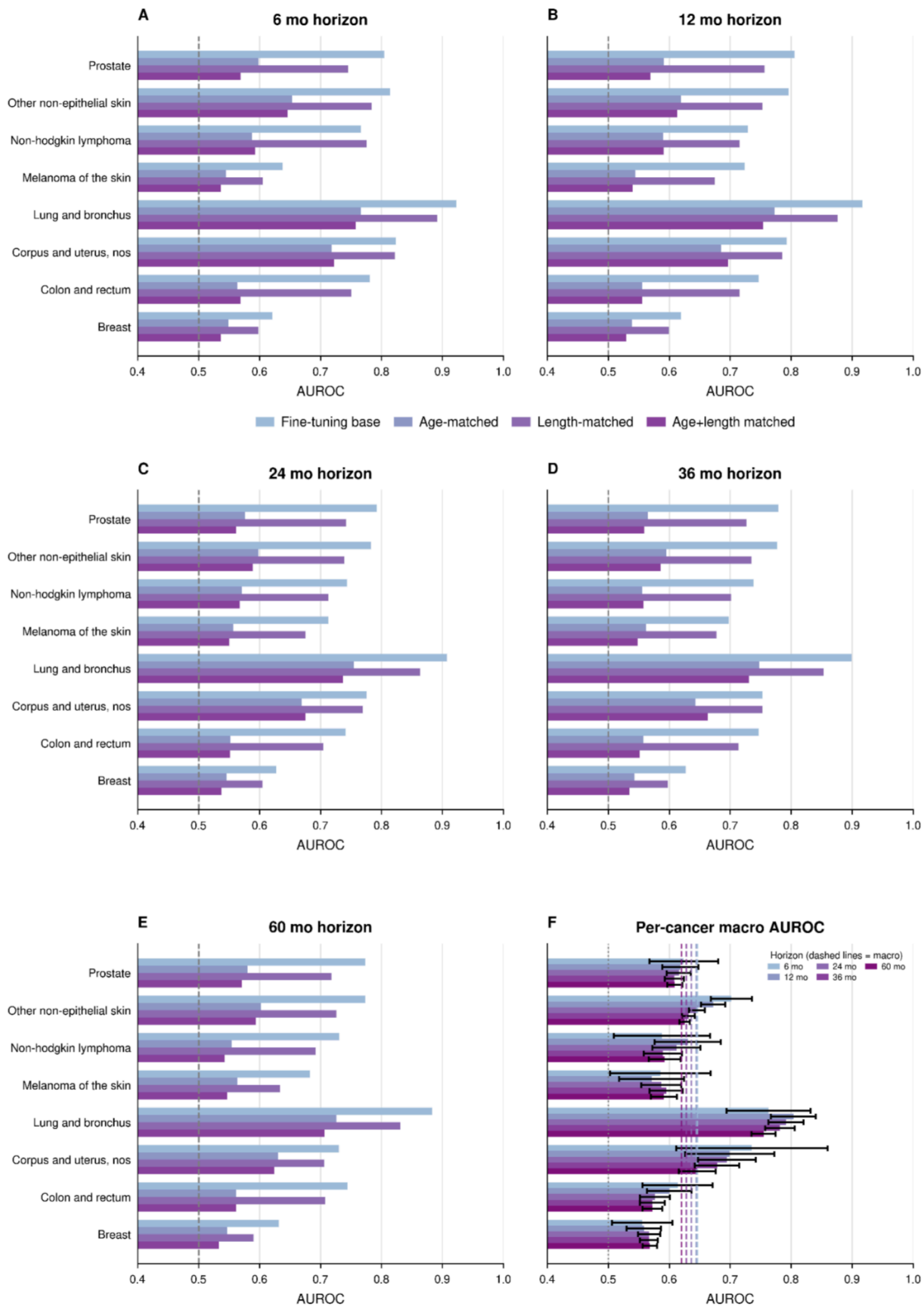


***Figure S37a: Sensitivity of UKBB cancer discrimination to age and trajectory length on cancer risk.*** *Cases and controls were matched by age bracket at the endpoint of trajectory as well as with an equal number of events. Age matched results in a drop of performance, but overall performance shows the model just doesn't rely on age and learns from the two combined. Length matched shows the results are not sensitive to the number of codes in patient*

trajectory. (A-E) are results for 6, 12, 24, 36, and 60 month horizons, with each row comparing original evaluation with age matching, event count matching, and both combined.

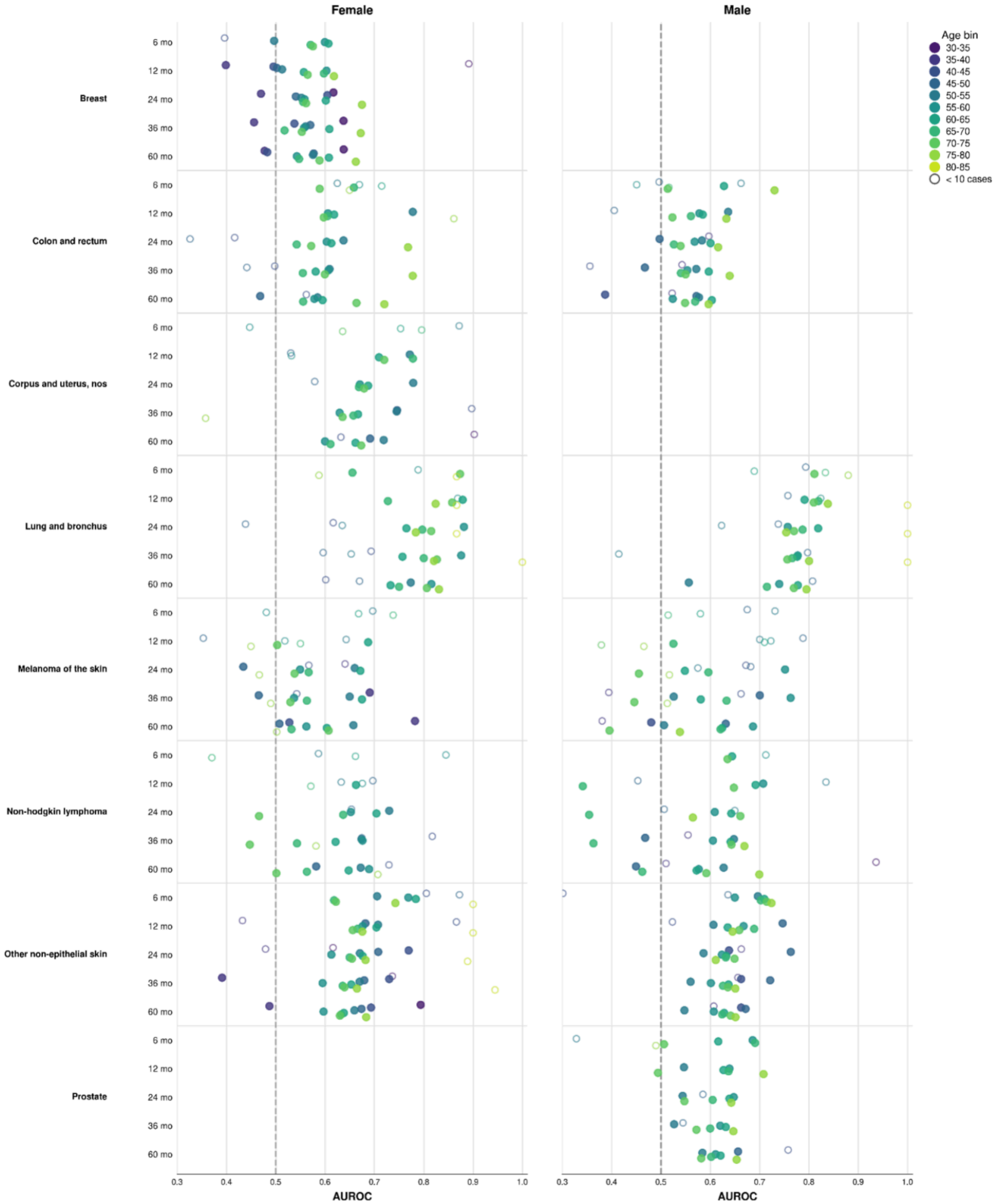


***Figure S37b: (UKBB Cohort) Breakdown of predictive performance by age-brackets and sex.*** *Performance by age and sex bracket for different groups assessing heterogeneity and data*

*sparsity. (Left) Performance on female population for each cancer (Right) Performance on male population for each cancer. Both are reported for all six (6, 12, 24, 36, and 60 months) prediction horizons. Color encodes five year age groups, and hollow circle means fewer than 10 positive outcomes in each bin.*

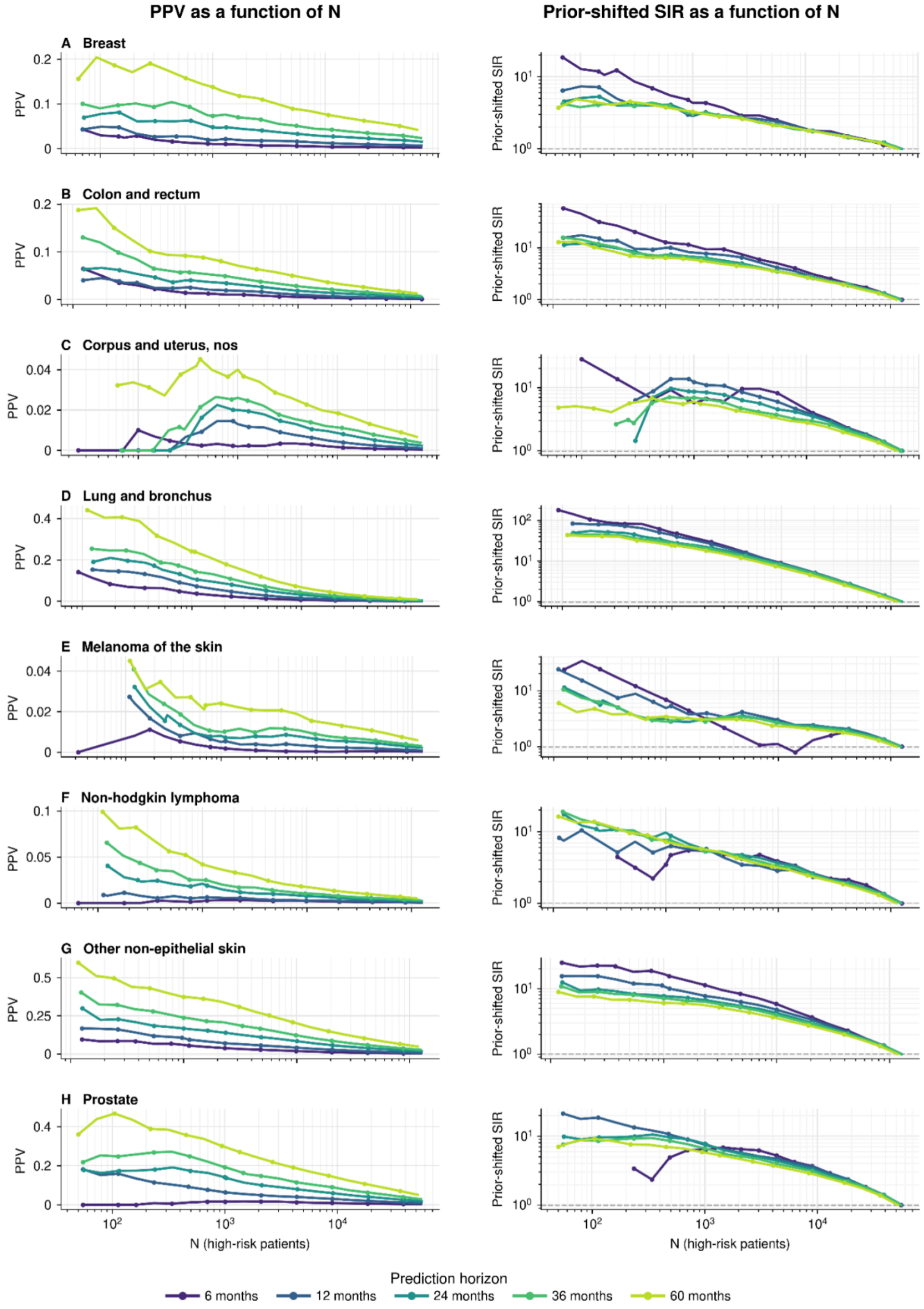

***Figure S38: (UKBB Cohort) Clinical utility of GenEHR fine-tuned model for nominating patients for screening by choosing an operating threshold****. PPV (left) and SIR (right) plotted as a function of operating threshold N shows a net benefit of detecting one cancer at the cost of increasing false positives. (A-F) breast, colon/rectum, corpus/uterus, lung and bronchus, melanoma of the skin, non-hodgkin lymphoma, other non-epithelial skin, and prostate cancer.*

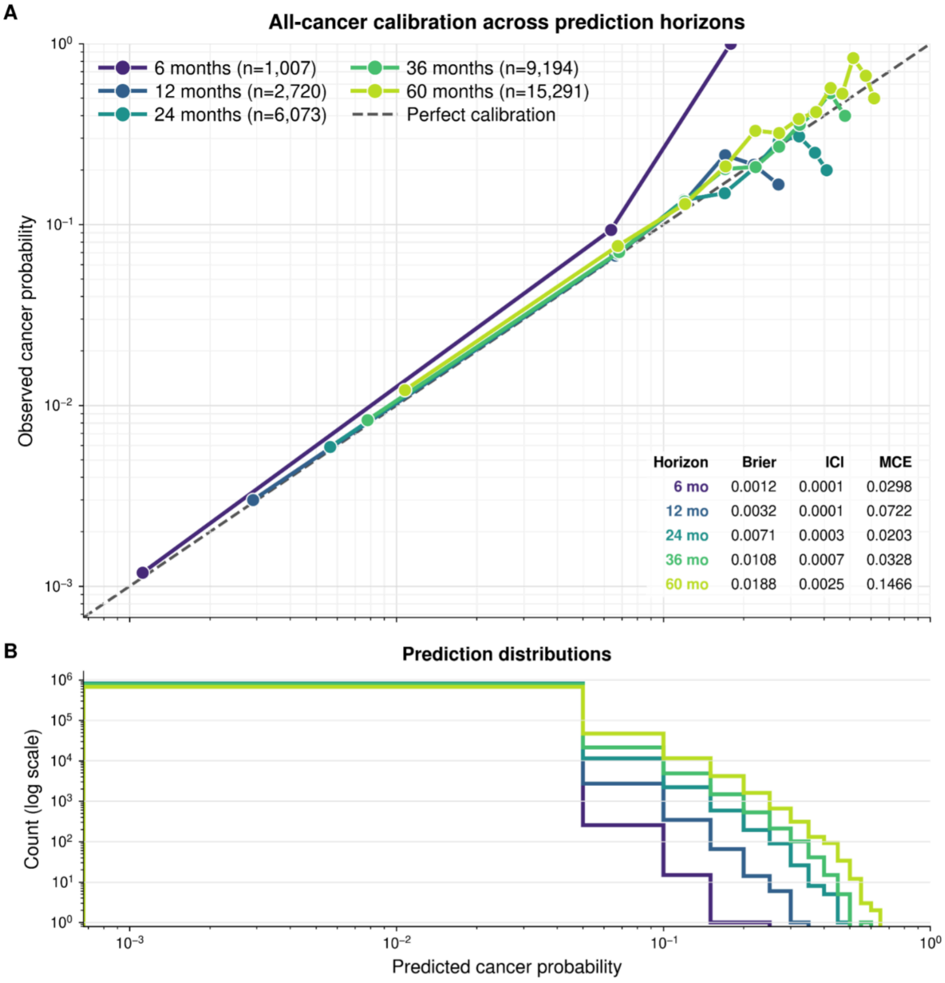


***Figure S39: (UKBB Cohort) Pan-cancer risk model is well calibrated across horizons and cancer types.*** *Observed cancer frequency against predicted probability across calibration bins for different horizons, where diagonal line indicates perfect calibration. Both ECE and Brier score describe fine-tuned cancer-risk probabilities (not next event calibration in Fig 3).*

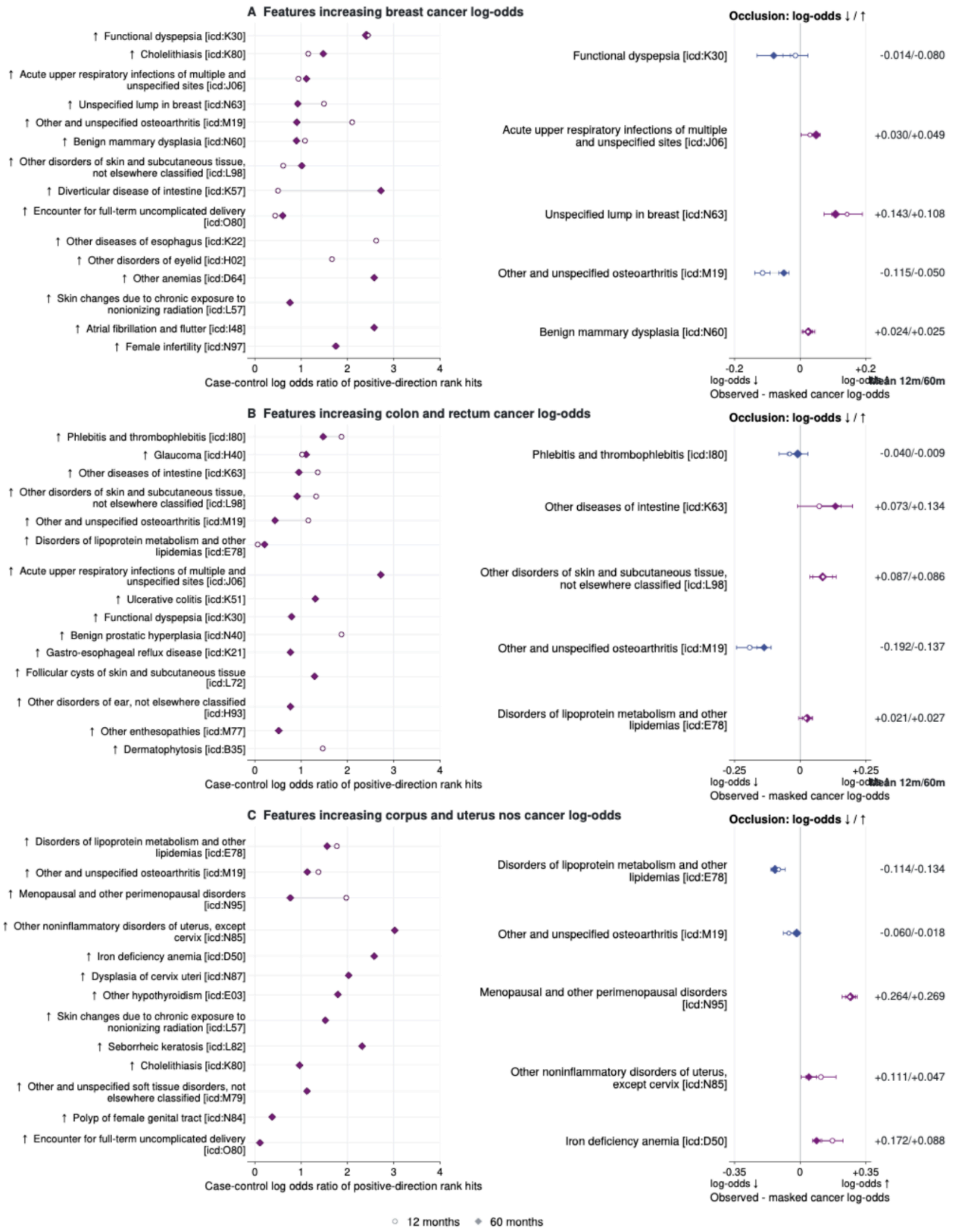


**Figure S40a:** ***Interpretation of features for selected cancer types (breast; colon and rectum; corpus and uterus nos) in UKBB cohort.*** *(Left) Case-control enrichment among top-ranked positive IG scores, x-axes values are log odds ratio of feature rank hits. (Right) Occlusion shows mean predicted log odds with the event observed minus masked, with whiskers of +/-1.96 occurrence level errors, positive means observed feature score is higher.*

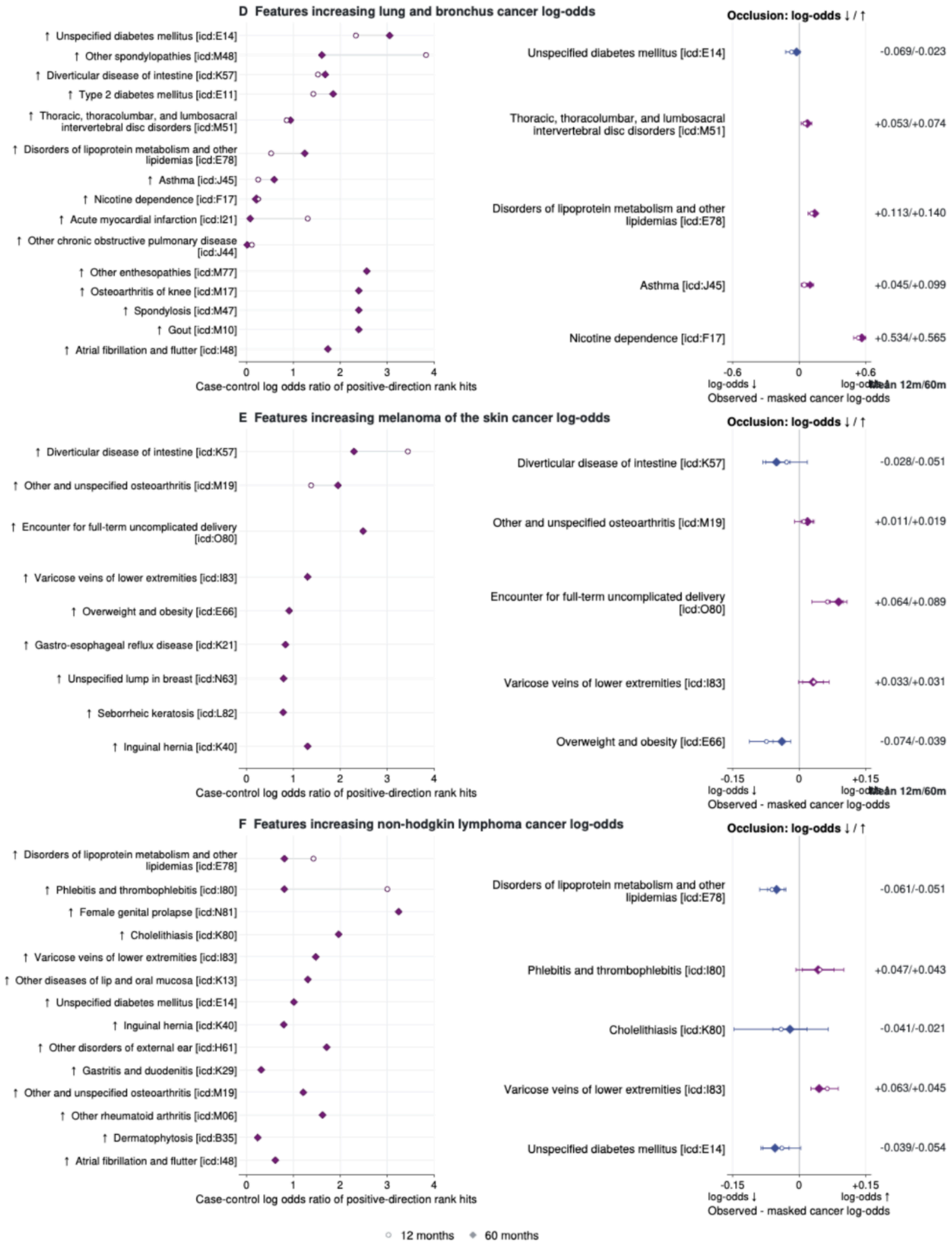


**Figure S40b:** ***Interpretation of features for selected cancer types (lung and bronchus; melanoma of the skin; non-hodgkin lymphoma) in the UKBB cohort.*** *(Left) Case-control enrichment among top-ranked positive IG scores, x-axes values are log odds ratio of feature rank hits. (Right) Occlusion shows mean predicted log odds with the event observed minus masked, with whiskers of +/-1.96 occurrence level errors, positive means observed feature score is higher.*

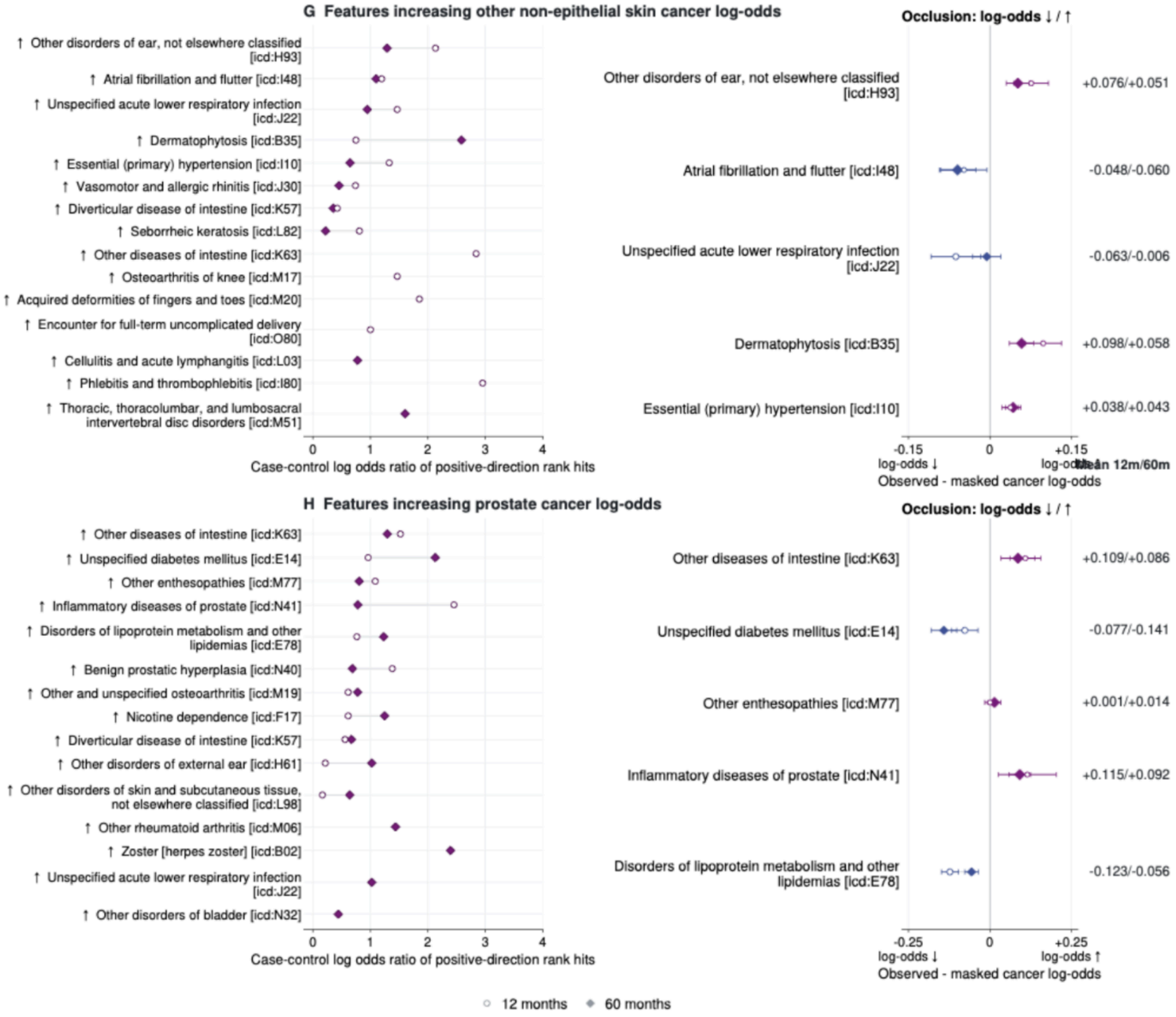


**Figure S40c:** ***Interpretation of features for selected cancer types (other non-epithelial skin; prostate cancer) in UKBB cohort.*** *(Left) Case-control enrichment among top-ranked positive IG scores, x-axes values are log odds ratio of feature rank hits. (Right) Occlusion shows mean predicted log odds with the event observed minus masked, with whiskers of +/-1.96 occurrence level errors, positive means observed feature score is higher.*

### All Of Us Results.

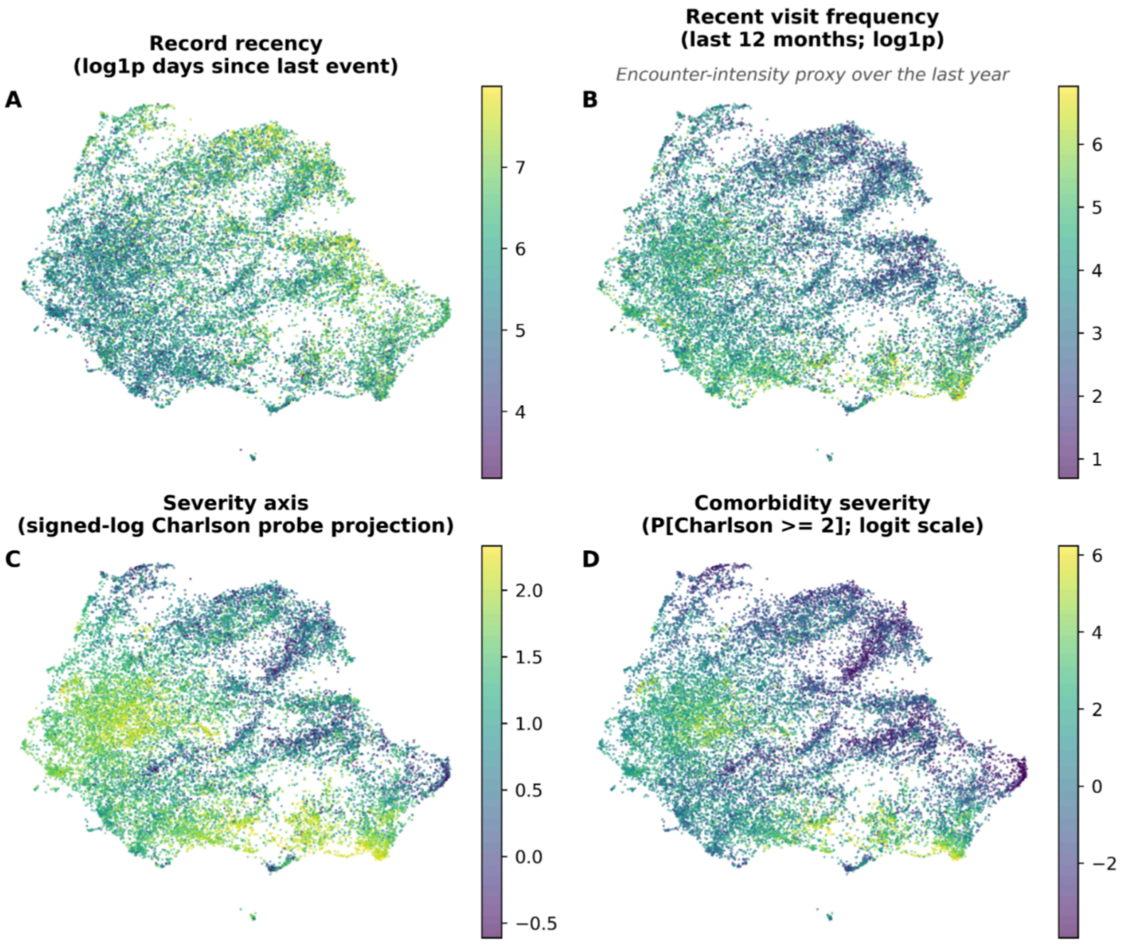


***Figure S41: Patient health state representation given by pooled encoding of all events in patient's trajectory (AoU cohort).*** *All four panels are two dimensional projections of patient trajectory representation obtained from the penultimate layer of GenEHR. Each has a different overlay: (A) time gap since last visit (B) number of visits in preceding year as a proxy for acuity. (C) a linear probe was fitted on feature representation for predicting CCI overlay with probe prediction (D) only patients who have CCI above chosen threshold. Gradients in CCI imply that representations capture information about overall morbidity and healthcare use.*

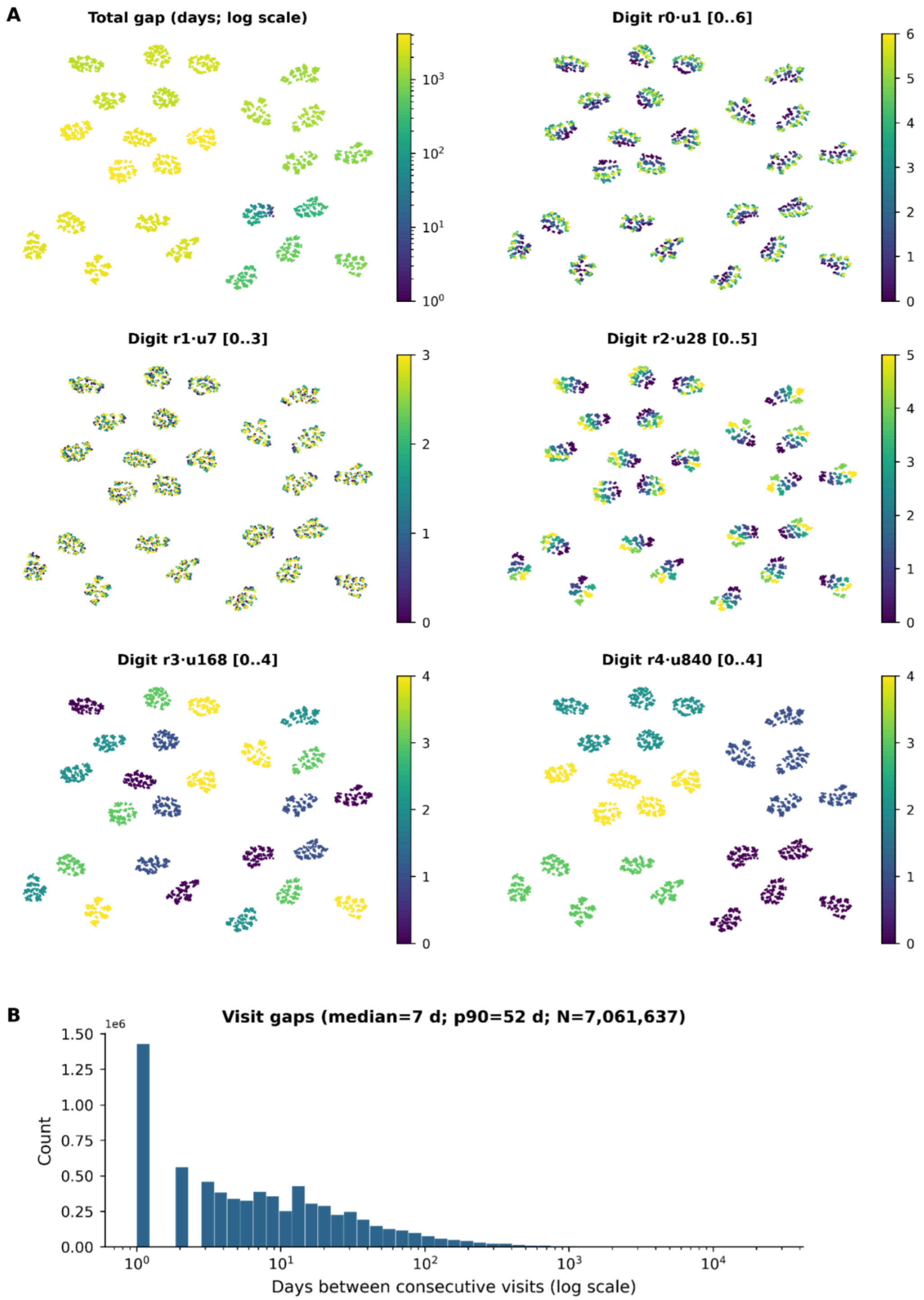


***Figure S42: Learned RATE representation and observed time gaps in AoU cohort.*** *UMAP projection of learned representation of 4200 days, colored by the value of each radix digit together with the observed distribution of inter-visit gaps. (A) Each point represents one encoded gap from 1 to 4200 days where color is the gap length. The other five show the digit with place values 1, 7, 28, 168, and 840 days with color indicating lower to higher values on each scale. (B) Inter-visit time interval distribution.*

**Event representations capture clinical structure and a context-dependent model-belief graph for hypothesis generation.**

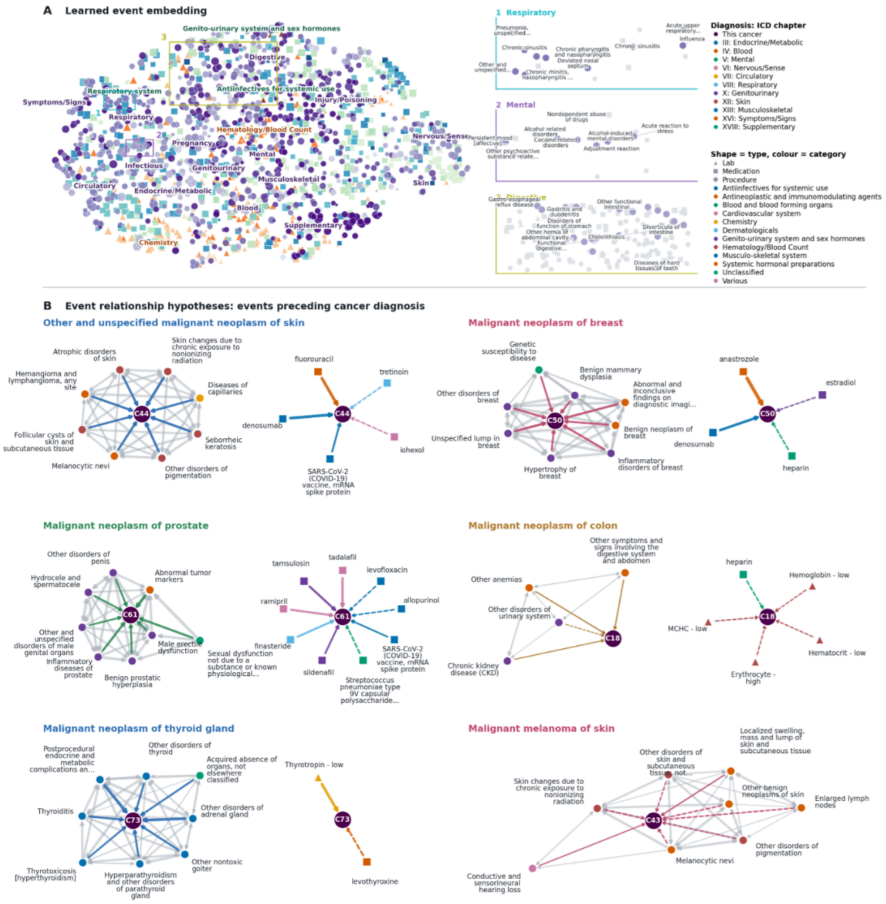


**Patient representation over multiple visits captures comorbidity burden.**

**Patient map (end-of-trajectory) with co-morbidity gradient**

**Patient health state progression over visits**

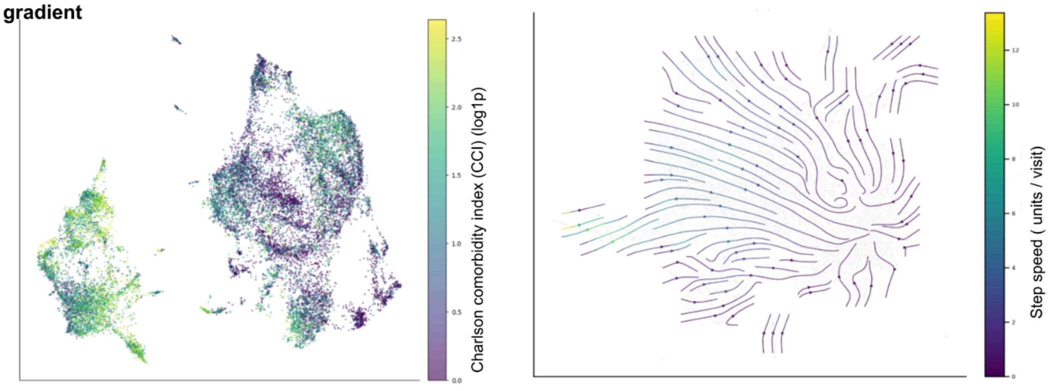


***Figure S43: For the AoU cohort.*** *Left:* ***(A)*** *UMAP projection of learned event embeddings using cosine distance between embedding vectors. Right: closeup of three regions numbered 1 to 3 shows GenEHR learns hierarchy of clinical events without explicit prior knowledge and without using text.* ***(B)*** *Event relationship preceding cancer diagnosis inferred using cPMI between pairs of events for selected cancers (ICD codes C50, C34, C61, C73). For held-out patients whose preceding visit contains an event* $e_1$*, cPMI compares the average model*

*probability assigned to $e_2$ normalized against the model averaged probability of $e_2$ across all positions and patients. In the cPMI belief graph nodes are clinical events and edges are informational links. Left: Diagnosis neighborhood of cancer with edges among preceding events, Right: Cross-modality hub - medications, procedures, and labs preceding cancer diagnosis. Dashed edges: events with weaker mutual information scores that are plausibly less specific to that cancer. (**Bottom**) UMAP neighborhoods of patients using their end-of-trajectory <eos> representation reflect gradient of comorbidity burden (left), and transitions along trajectories are informative about how the learned state of patients evolves over time using their <visit> representation over multiple visits. Flow (arrows) indicates how patient health evolves over visits (right) averaged over groups of held-out patients.*

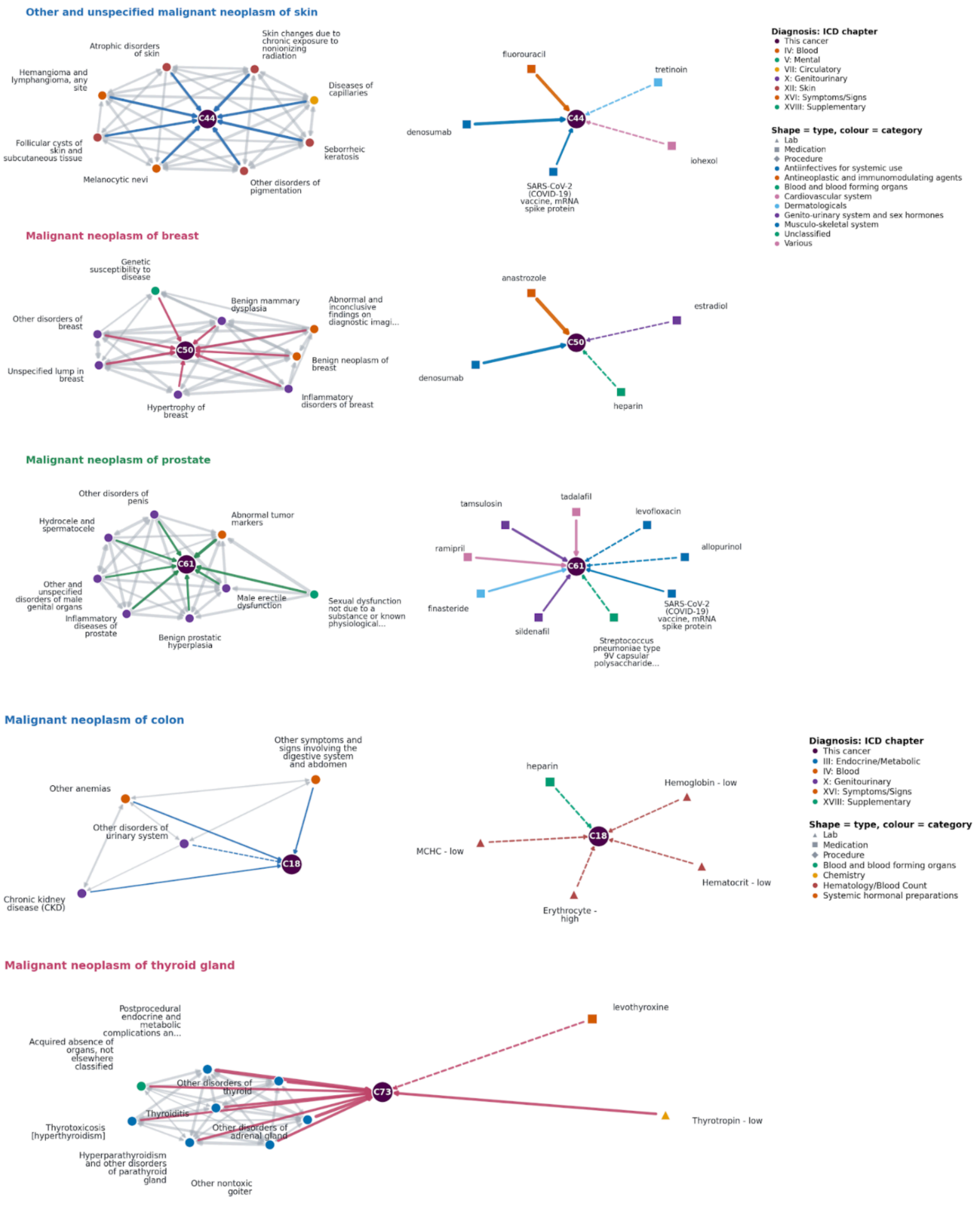

***Figure S44a: Breakdown of within and across modality belief graphs across cancer types (AoU Cohort).*** *Event relationship preceding cancer diagnosis inferred using cPMI between pairs of events for selected cancers (ICD codes C44, C50, C61, C18, C73). For held-out patients whose preceding visit contains an event* $e_1$*, cPMI compares the average model probability assigned to* $e_2$ *normalized against the model averaged probability of* $e_2$ *across all positions and patients. In the cPMI belief graph nodes are clinical events and edges are informational links. Left: Diagnosis neighborhood of cancer with edges among preceding events, Right: Cross-modality hub - medications, procedures, and labs preceding cancer diagnosis. Dashed edges: events with weaker mutual information scores that are plausibly less specific to that cancer.*

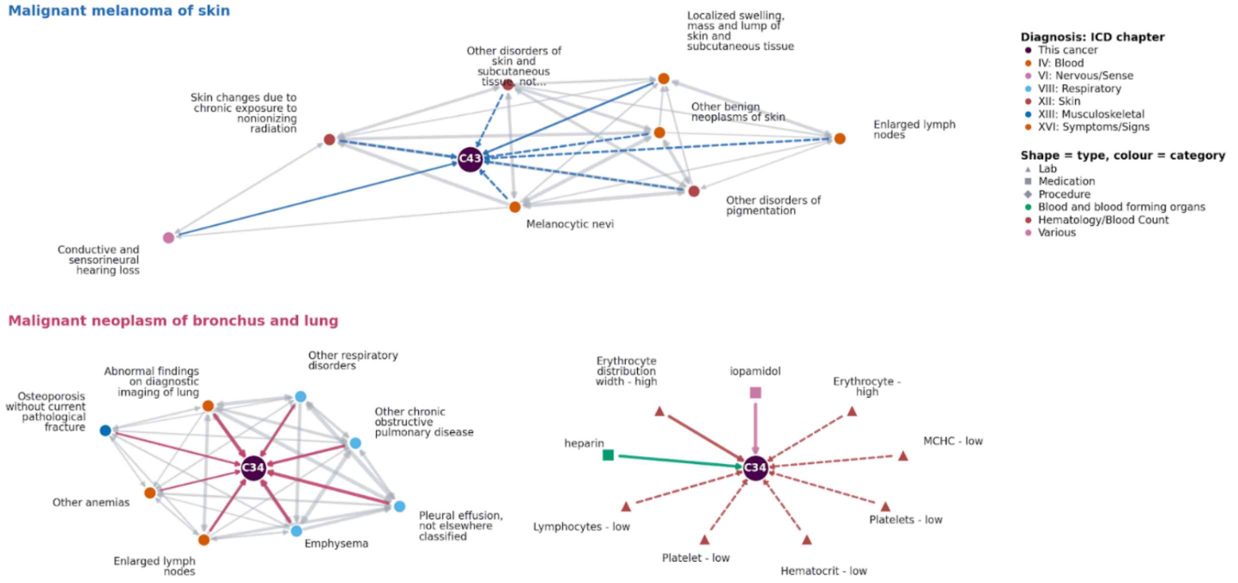


***Figure S44b: Breakdown of within and across modality belief graphs across cancer types (PROV Cohort).*** *Event relationship preceding cancer diagnosis inferred using cPMI between pairs of events for selected cancers (ICD codes C43, C34). For held-out patients whose preceding visit contains an event* $e_1$*, cPMI compares the average model probability assigned to* $e_2$ *normalized against the model averaged probability of* $e_2$ *across all positions and patients. In the cPMI belief graph nodes are clinical events and edges are informational links. Left: Diagnosis neighborhood of cancer with edges among preceding events, Right: Cross-modality hub - medications, procedures, and labs preceding cancer diagnosis. Dashed edges: events with weaker mutual information scores that are plausibly less specific to that cancer.*

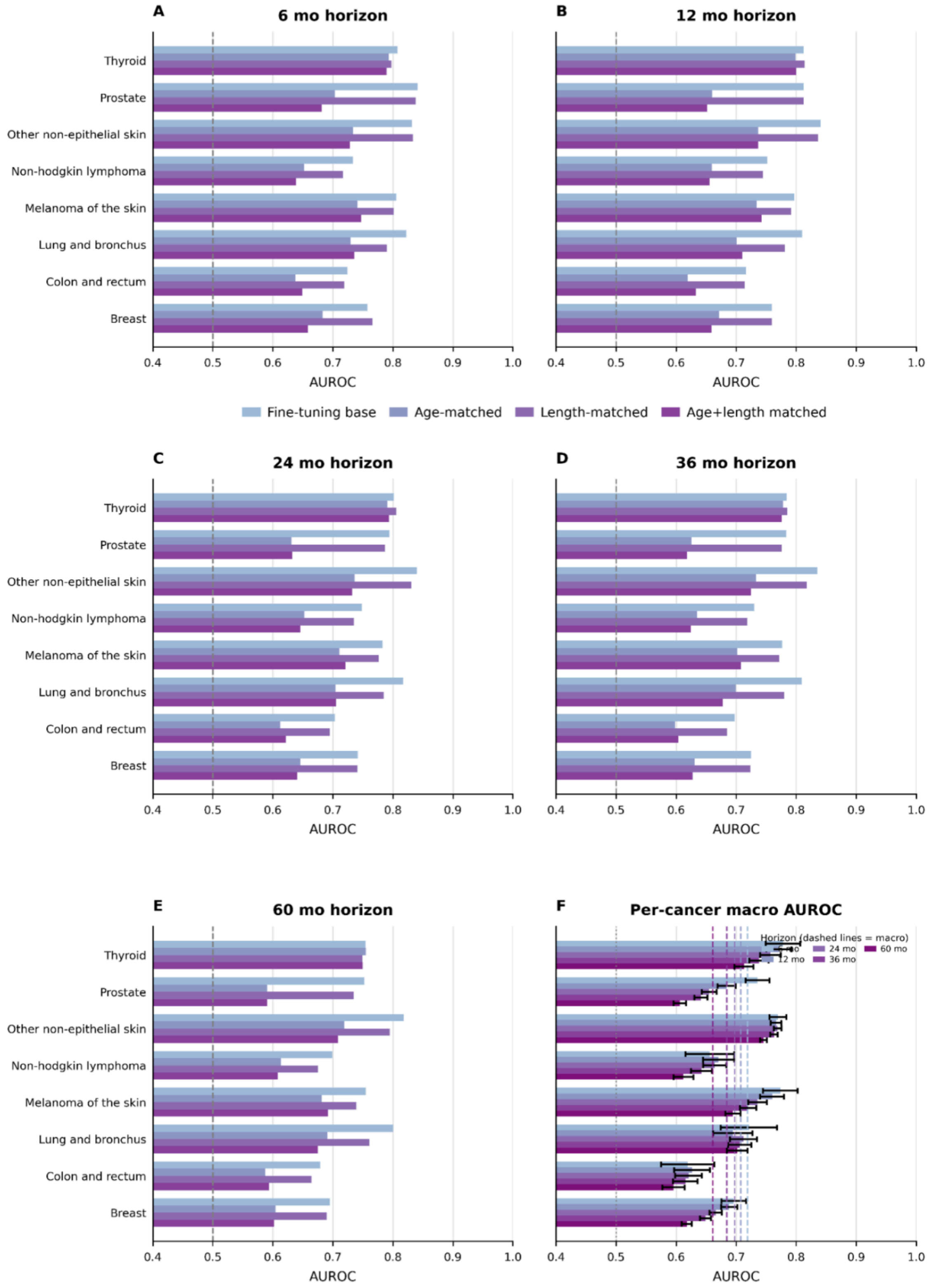


***Figure S45a: Sensitivity of AoU cohort cancer discrimination to age and trajectory length on cancer risk.*** *Cases and controls were matched by age bracket at the endpoint of trajectory as well as with an equal number of events. Age matched results in a drop of performance, but*

*overall performance shows the model just doesn't rely on age and learns from the two combined. Length matched shows the results are not sensitive to the number of codes in patient trajectory. (A-E) are results for 6, 12, 24, 36, and 60 month horizons, with each row comparing original evaluation with age matching, event count matching, and both combined.*

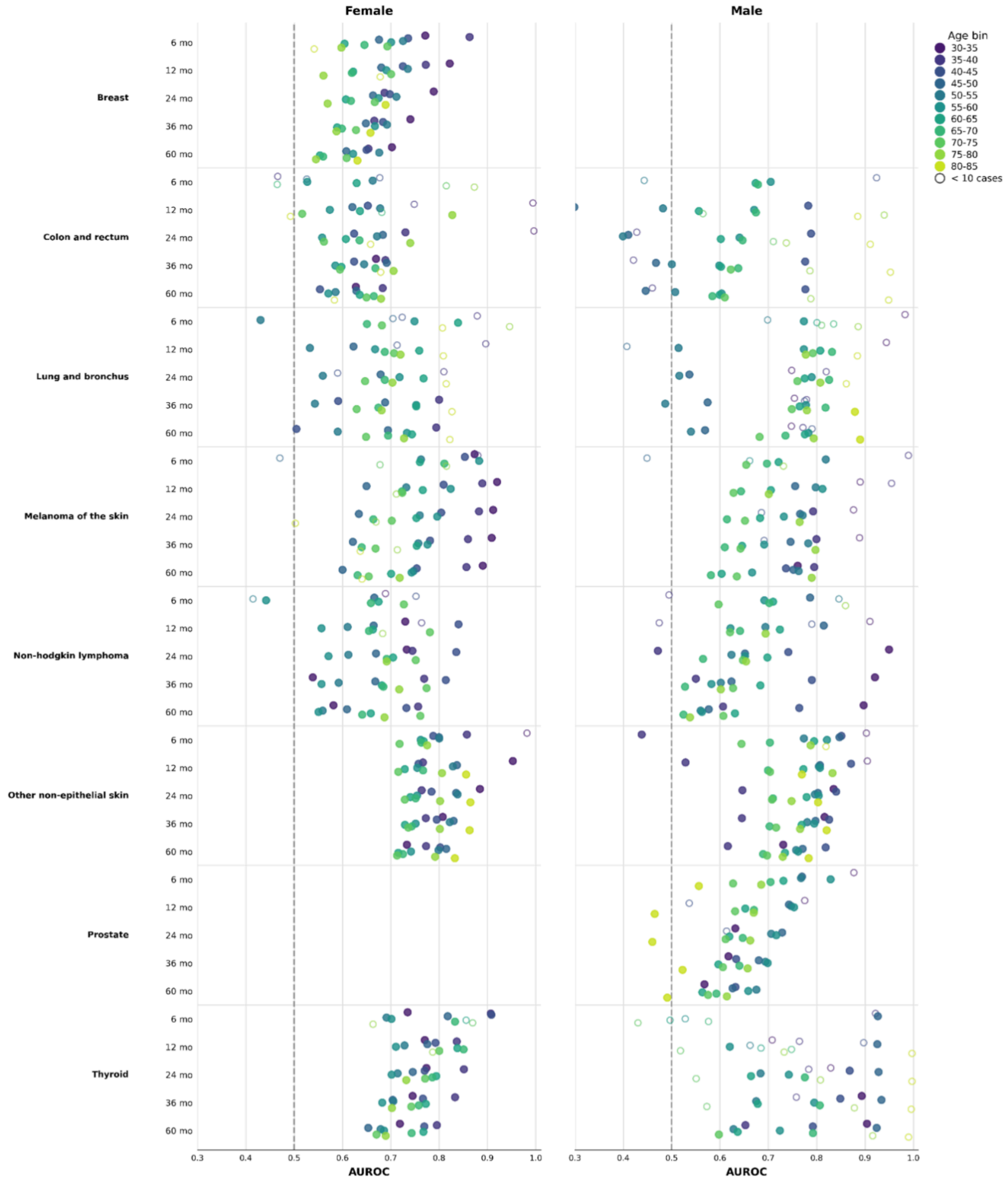


***Figure S45b: (AoU Cohort) Breakdown of predictive performance by age-brackets and sex.*** *Performance by age and sex bracket for different groups assessing heterogeneity and data sparsity. (Left) Performance on female population for each cancer (Right) Performance on male population for each cancer. Both are reported for all six (6, 12, 24, 36, and 60 months)*

*prediction horizons. Color encodes five year age groups, and hollow circle means fewer than 10 positive outcomes in each bin.*

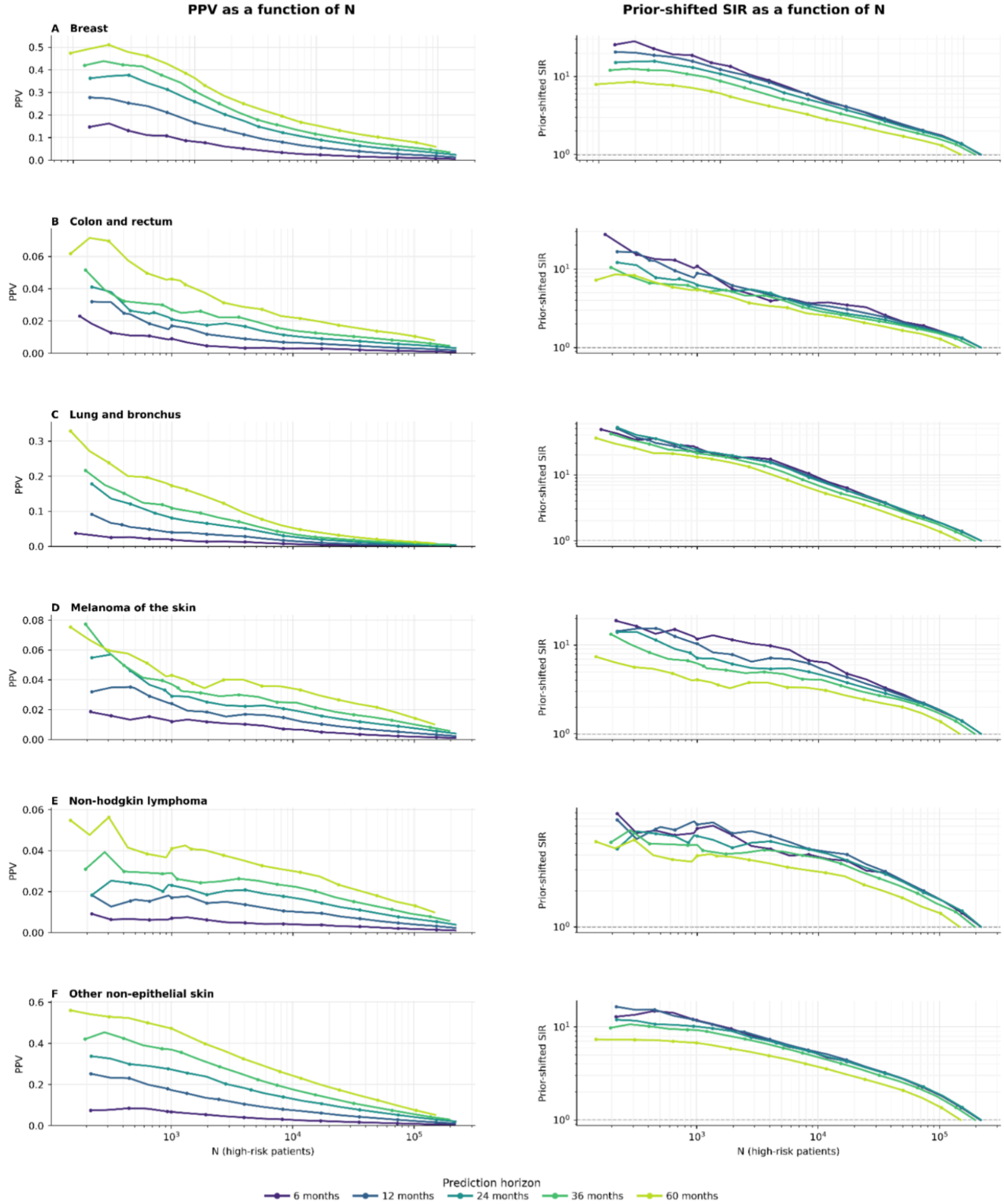

***Figure S46a: (AoU Cohort) Clinical utility of GenEHR fine-tuned model for nominating patients for screening by choosing an operating threshold**. PPV (left) and SIR (right) plotted as a function of operating threshold N shows a net benefit of detecting one cancer at the cost of increasing false positives. (A-F) brain, breast, colon/rectum, corpus/uterus, esophagus, and kidney/renal-pelvis cancer.*

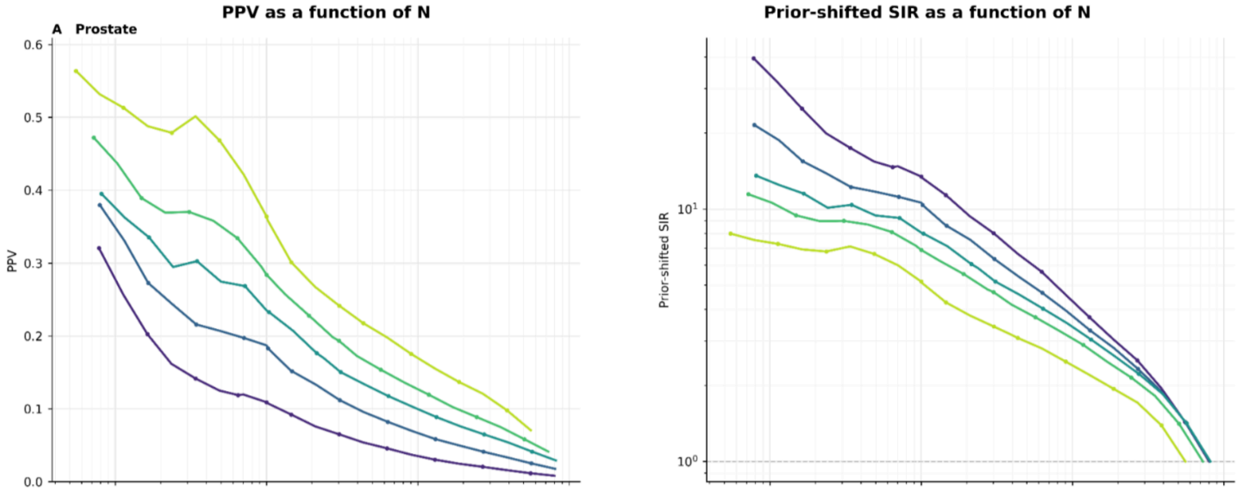


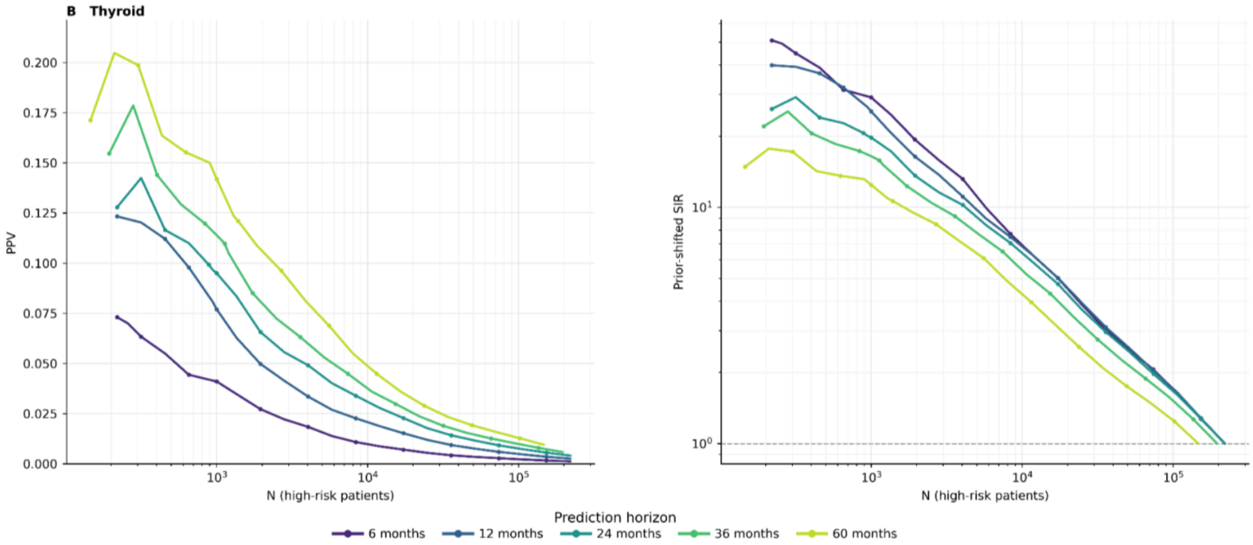


***Figure S46b: (AoU Cohort) Clinical utility of GenEHR fine-tuned model for nominating patients for screening by choosing an operating threshold**. PPV (left) and SIR (right) plotted as a function of operating threshold N shows a net benefit of detecting one cancer at the cost of increasing false positives. (A-F) brain, breast, colon/rectum, corpus/uterus, esophagus, and kidney/renal-pelvis cancer.*

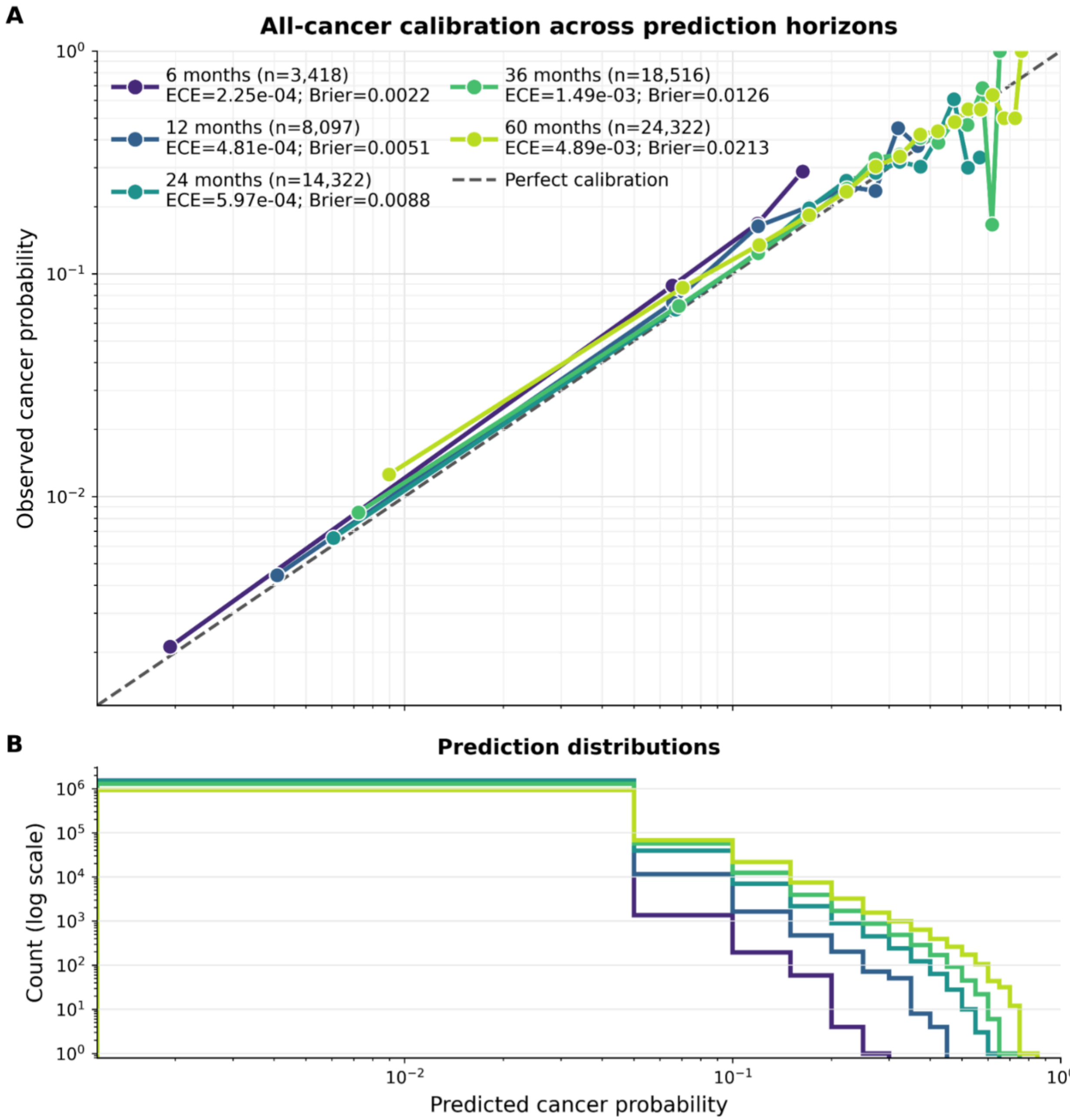


***Figure S47: (AoU Cohort) Pan-cancer risk model is well calibrated across horizons and cancer types.*** *Observed cancer frequency against predicted probability across calibration bins for different horizons, where diagonal line indicates perfect calibration. Both ECE and Brier score describe fine-tuned cancer-risk probabilities (not next event calibration in Fig 3).*

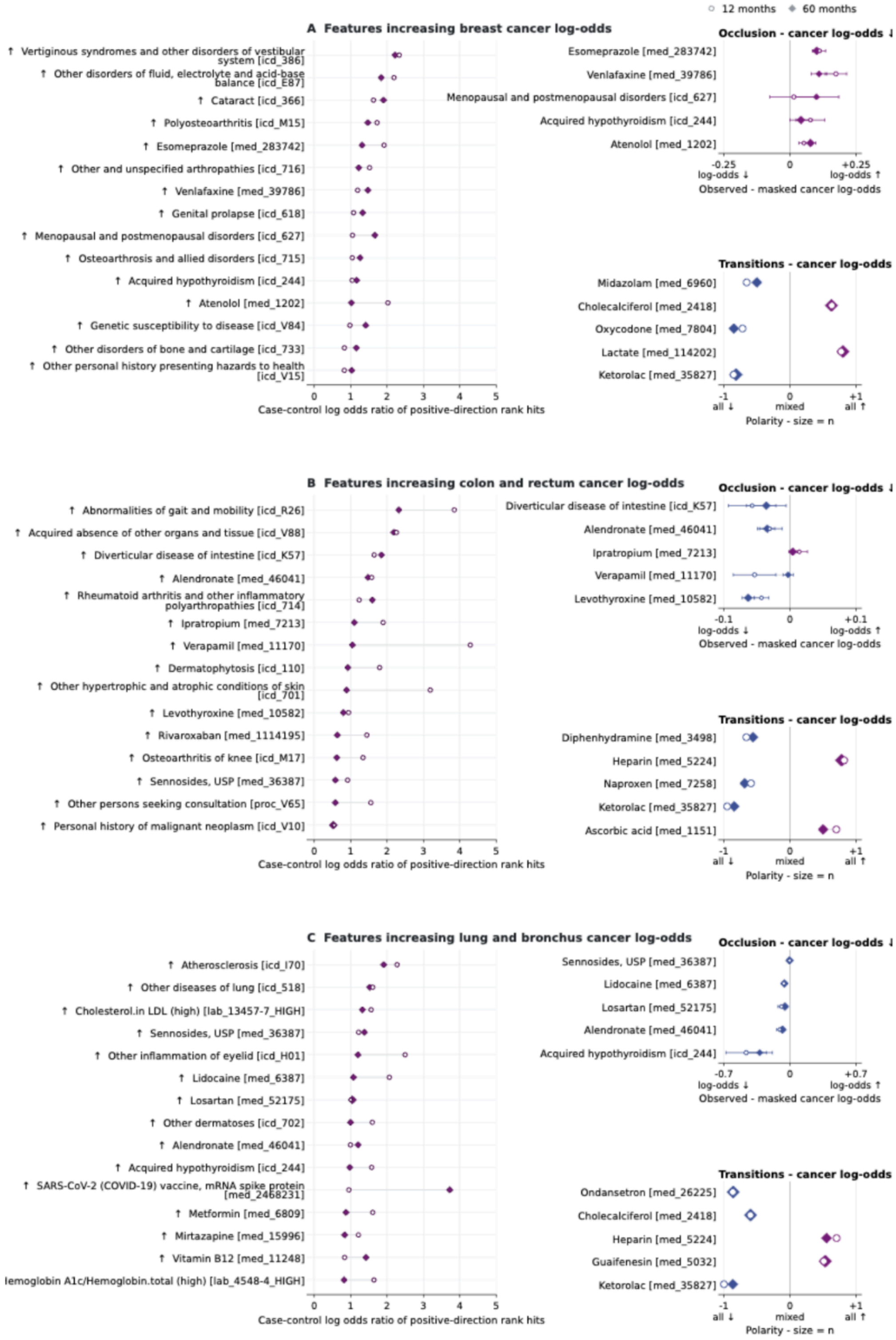


***Figure S48a: Interpretation of features for selected cancer types (breast; colon and rectum; lung and bronchus) in AoU cohort.*** *(Left) Case-control enrichment among top-ranked positive IG scores, x-axes values are log odds ratio of feature rank hits. (Upper right)*

*Occlusion shows mean predicted log odds with the event observed minus masked, with whiskers of +/-1.96 occurrence level errors, positive means observed feature score is higher. (Lower right) Score-change directions around medication and procedure transitions (-1 all retained changes decrease the score, +1 all increase it).*

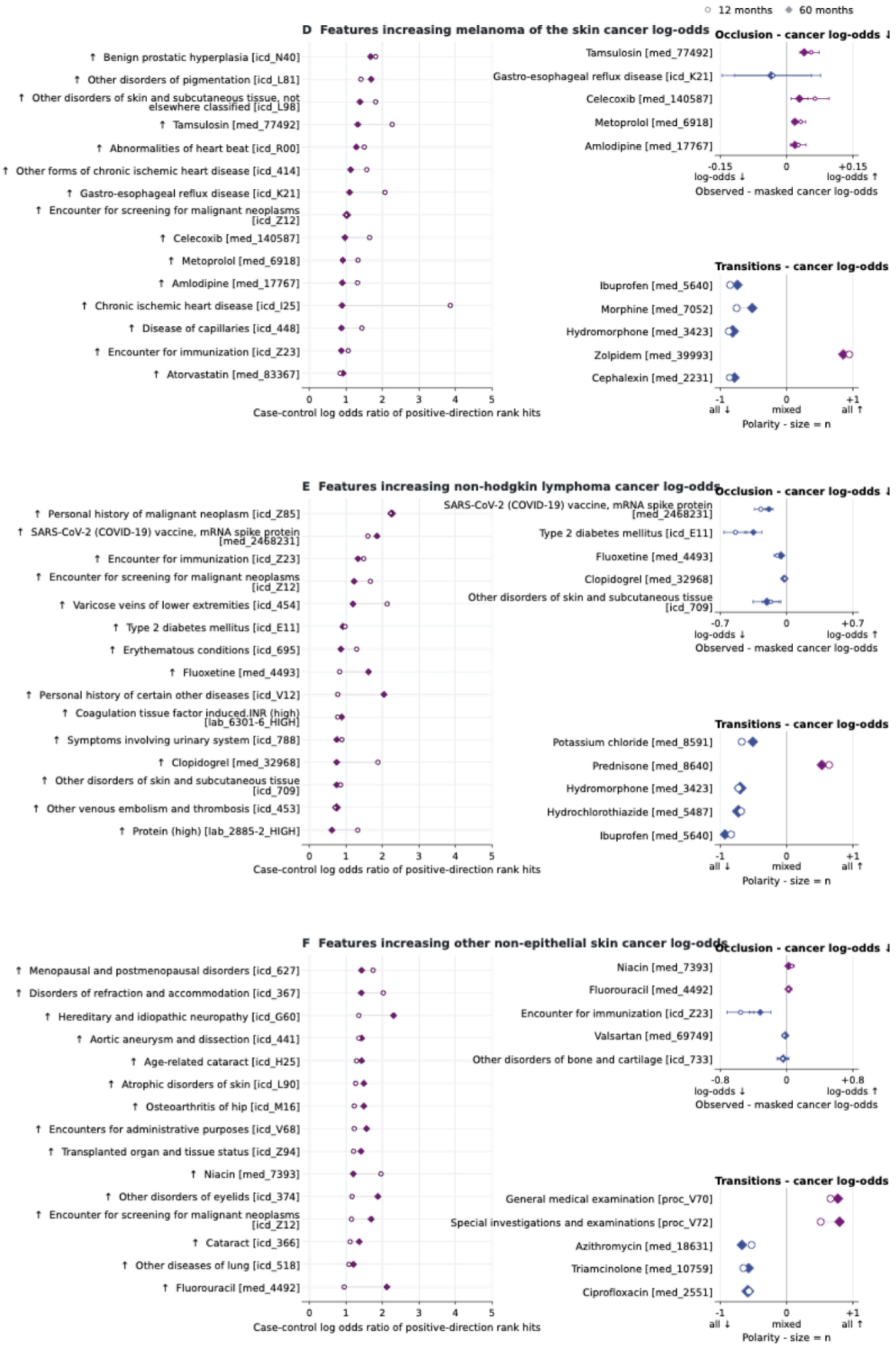


***Figure S48b: Interpretation of features for selected cancer types (melanoma of the skin; non-hodgkin lymphoma; other nopn-epithelial skin) in AoU cohort.*** *(Left) Case-control*

*enrichment among top-ranked positive IG scores, x-axes values are log odds ratio of feature rank hits. (Upper right) Occlusion shows mean predicted log odds with the event observed minus masked, with whiskers of +/-1.96 occurrence level errors, positive means observed feature score is higher. (Lower right) Score-change directions around medication and procedure transitions (-1 all retained changes decrease the score, +1 all increase it).*

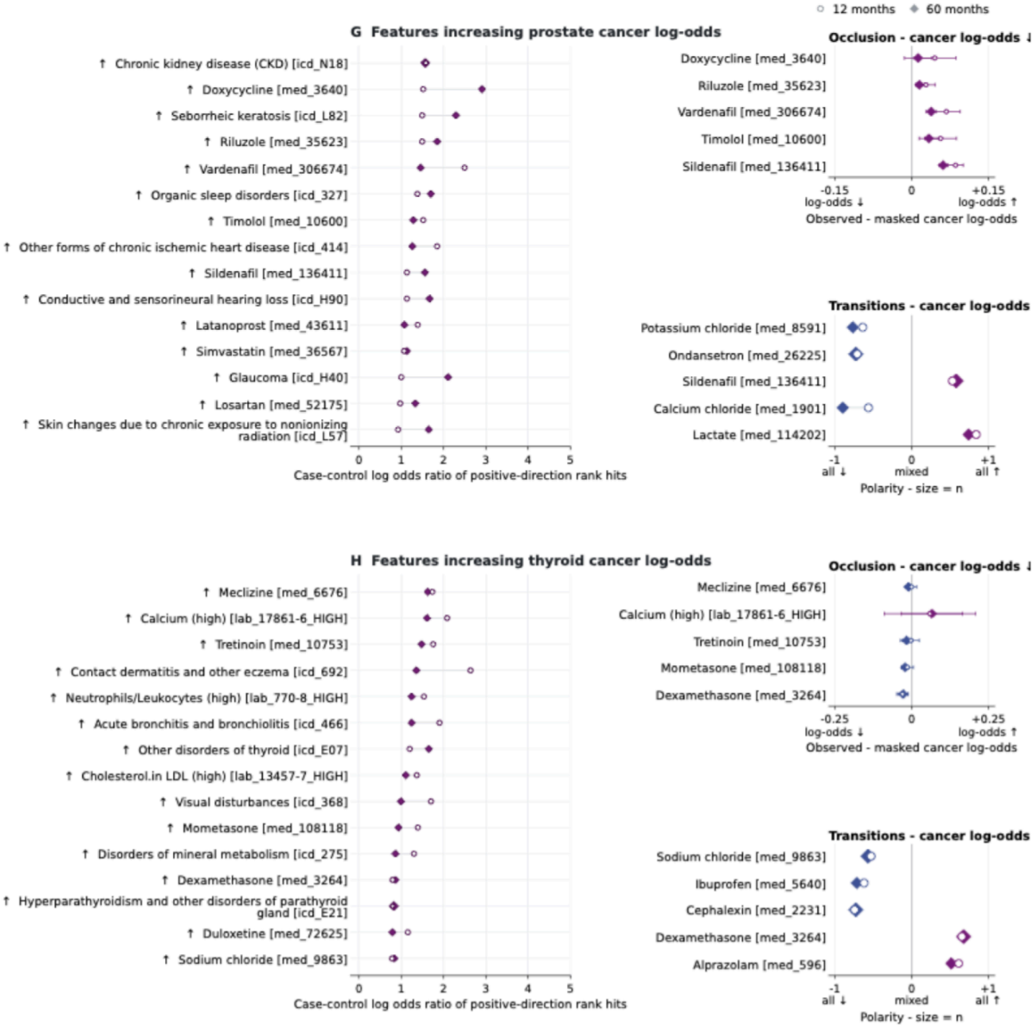


***Figure S48c: Interpretation of features for selected cancer types (prostate; thyroid) in PROV cohort.*** *(Left) Case-control enrichment among top-ranked positive IG scores, x-axes values are log odds ratio of feature rank hits. (Upper right) Occlusion shows mean predicted log odds with the event observed minus masked, with whiskers of +/-1.96 occurrence level errors, positive means observed feature score is higher. (Lower right) Score-change directions around medication and procedure transitions (-1 all retained changes decrease the score, +1 all increase it).*
